# Seeded Probes Do Not Identify a Reviewer’s Miss Rate: One-Sided Misclassification in Distribution-Free Risk Control

**DOI:** 10.64898/2026.10.01.26364465

**Authors:** Krishna Chaitanya Kotabhattara, Abhishek Siroha, Pijush Pal, Surya K, Venkata Siddharth Dhara, Priya Prabhakar, Nagaraj Honkote, Mounika Marreddy, Namita, Athmica Kishore K, Pramit Saha, Jacob M Agris

## Abstract

Distribution-free risk control certifies that the rate of defects that would change clinical management, among the answers an AI system releases, stays below a stated level, calibrated on cases a human reviewer has checked. The reviewer is an imperfect reference standard whose errors run one way: a missed defect enters calibration as a safe label. This one-sided misclassification lowers the risk on record at every release threshold, so the certificate is anti-conservative by a factor set by the reviewer’s miss rate on the system’s own defects. We show that seeded probes, which plant known defects and count the catches, do not identify this miss rate. A reviewer’s chance of missing a defect depends on how hard that defect is to spot, its detectability. A probe samples defects at detectabilities its designer chose. The system’s own defects fall at detectabilities nothing chose, and converting the probe’s miss rate into theirs requires the very rate being sought. This holds whenever detectability is not fully captured by an observed score, since a score that captured it would let probes covering its range identify the rate. Cytology field validation builds probe sets whose miss rate is a floor on the true one. Even then the probe bounds the rate from below only, so it can reject a certificate and cannot grant one. The finite-sample label-noise corrections assume noise independent of the case given its label, but a reviewer misses the defects that read as plausible, and the condition they need fails in all four simulated reviewer profiles we examine. Testing that condition on an independent sample takes 185 adjudicated cases for a reviewer missing 85% of defects and 147,938 for one missing 3%. A second reader who adjudicates the cases the careful reviewer passed finds a miss with 95% confidence within 498, so looking for the noise directly costs far less than testing a model of it. In a public corpus of model answers graded by physicians, a physician passed 26.3% of the defects the other graders agreed were there (95% interval 21.6% to 31.7%, resampling whole conversations), on criteria about seeking context and stating uncertainty, and individual rates ran from 8.0% to 54.5%.

## 1. Introduction

Distribution-free risk control gives a deployment a threshold 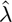 chosen on a calibration set. The risk of the released output then exceeds a stated level *α* with probability at most *δ*, under exchangeability and with no assumption on the model that produced it (Angelopoulos et al., 2025). For a clinical assistant the appeal is direct. Score each candidate answer with a verifier, release the answers scoring under 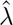, escalate the rest to a clinician, and the certificate bounds the rate at which a released answer carries a defect that would change care.

The calibration set is where that argument meets the ward. Someone has to say, case by case, whether the answer was safe, in every deployment we have examined that someone is a clinician, and clinicians miss defects. A missed defect does not enter calibration as a disagreement or as a missing label. It enters as a clean label on a case that was in fact an error, so the contamination is one-directional and points toward measured safety. Widening the certificate to absorb the reviewer’s miss rate means tightening *α*, the level calibration runs at, and doing that right means estimating the rate.

Seeded probes are the instrument a mature safety programme reaches for here, and two regulated industries have run them for decades: aviation screeners work against Threat Image Projection, which inserts a synthetic threat into a live X-ray stream, and clinical laboratories work against proficiency testing, which sends known samples for analysis. Both literatures report probe material unrepresentative of the real thing (Riz à Porta et al., 2022; Parsons et al., 2001). It is tempting to read that as a settled direction and to price it, but Section 4 shows the record will not carry a signed law, let alone a magnitude.

Our contribution starts one step below the direction, because a deployment that accepts the direction still has to choose a number. The natural move is to bound the probe miss rate with an exact binomial interval (Clopper & Pearson, 1934), multiply it by a realism penalty greater than one, and shrink *α* by the result. We argue in Section 4 that no such penalty is defensible, and the argument turns on identification, with conservatism beside the point. Miss rate is a function of how detectable a defect is, a probe is planted at a detectability the seeder chose, a model’s own defects fall at a detectability nothing chose, and the model’s own can be measured only by finding out which real defects the reviewer missed. That is the quantity the exercise set out to estimate. The instrument’s design leaves the parameter unidentified, and more data does not help.

Corrupted calibration feedback has a literature close to this setting, and we borrow from it rather than argue with it, so Section 7 says which part of the claim below is not ours (Wang et al., 2026). The finite-sample corrections in that line assume class-conditional noise, noise independent of the case once its true label is fixed (Sesia et al., 2025; Clarkson et al., 2024; Penso et al., 2025), and a reviewer miss breaks that assumption, because the flip happens precisely when the case was an error and read well. The results that assume nothing about the noise bound the gap in both directions and leave its sign open (Barber et al., 2023; Clarkson et al., 2024), and under-coverage from one-directional noise is already reported (Einbinder et al., 2024; Bashari et al., 2025). One-sidedness fixes the sign. A miss can only move a case from defect to safe, so a miss rate of *m* scales the risk on record to at most (1 *− m*) of its true value at every threshold, and calibration reports success at a threshold that has not earned it. The direction is a known bound (Stoye, 2022). Dependence on the score adds a second failure, because the miss rate is not one number. When misses concentrate at low scores, the miss rate among released defects at a low threshold exceeds the overall rate, so a correction that divides by the overall rate under-corrects exactly at the thresholds a deployment uses. Three things follow, and Fig. 1 is the shape they make.

**Figure 1.**
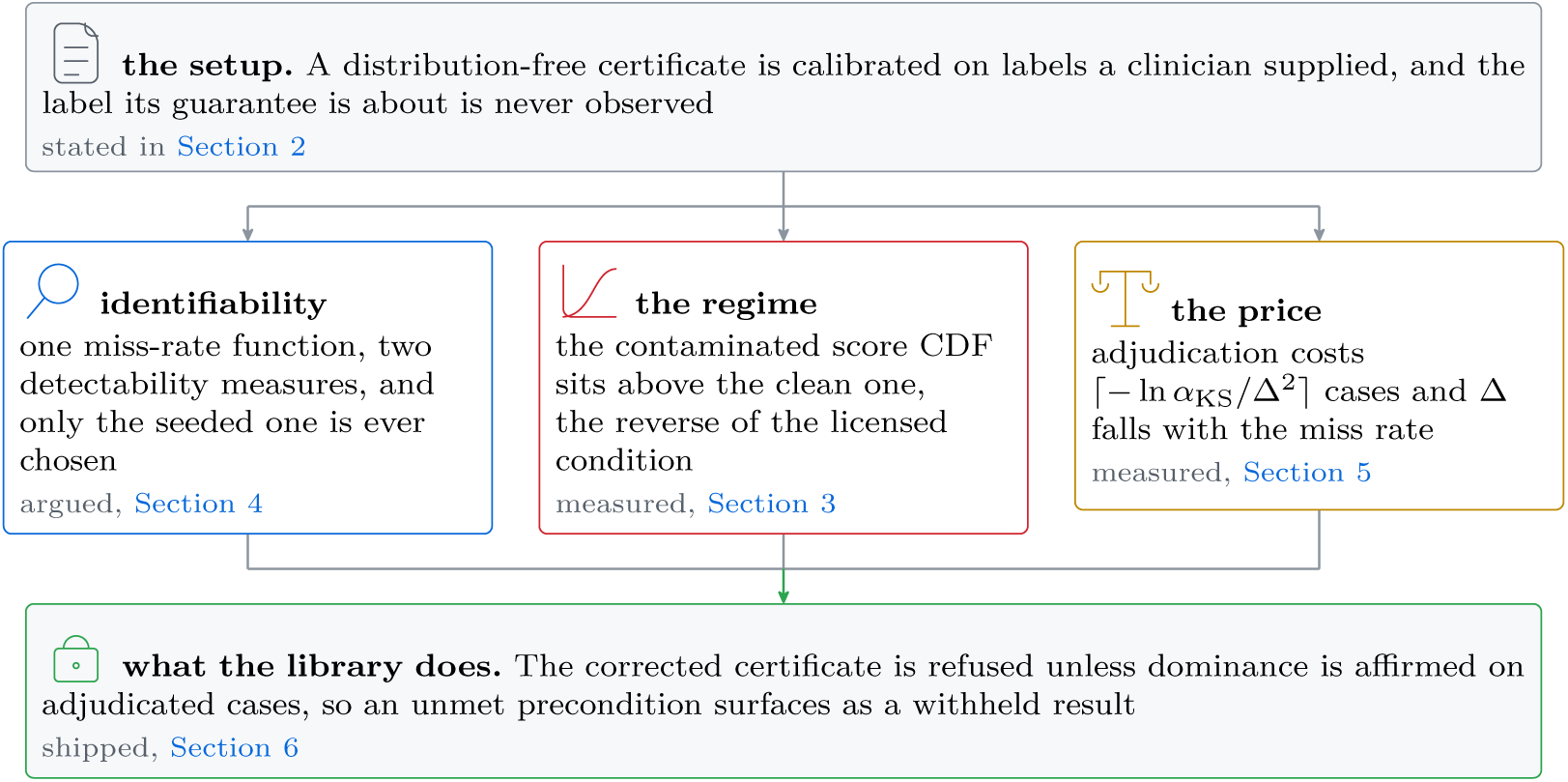
The three results, and the kind of evidence each one rests on. One premise feeds all three, all three feed one consequence, and no one of the three needs either of the others. The marker on a box is its kind of evidence, and they are not interchangeable: a deduction 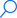, two measurements over four simulated reviewers 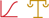, and a code path with a test.

## 2. Setup

### The certificate

A model produces an answer *y* for a case *x*. A verifier assigns a nonconformity score *s*(*x, y*) *∈* [0, 1], where a larger score means a stronger indication that the answer carries a defect. Let *Y ∈ {*0, 1*}* indicate that the answer contains a defect that would change care. Given a calibration sample and a candidate threshold *λ*, define the risk

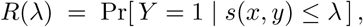

the rate at which a released answer carries a defect. Learn-then-Test selects the largest *λ* for which the hypothesis *R*(*λ*) *> α* is rejected at level *δ*, and the resulting 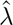 satisfies Pr[*R*(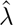) *≤ α*] *≥* 1 *− δ* over the draw of the calibration set (Angelopoulos et al., 2025). In words, *α* is the risk the deployment agrees to run and *δ* is the chance it accepts that the calibration sample was an unlucky draw. The guarantee needs no assumption about the model, the verifier, or the score. Figure 2 is the whole arrangement, and the argument of this paper concerns one of its six edges. Section A restates every symbol and verdict name without symbols.

**Figure 2.**
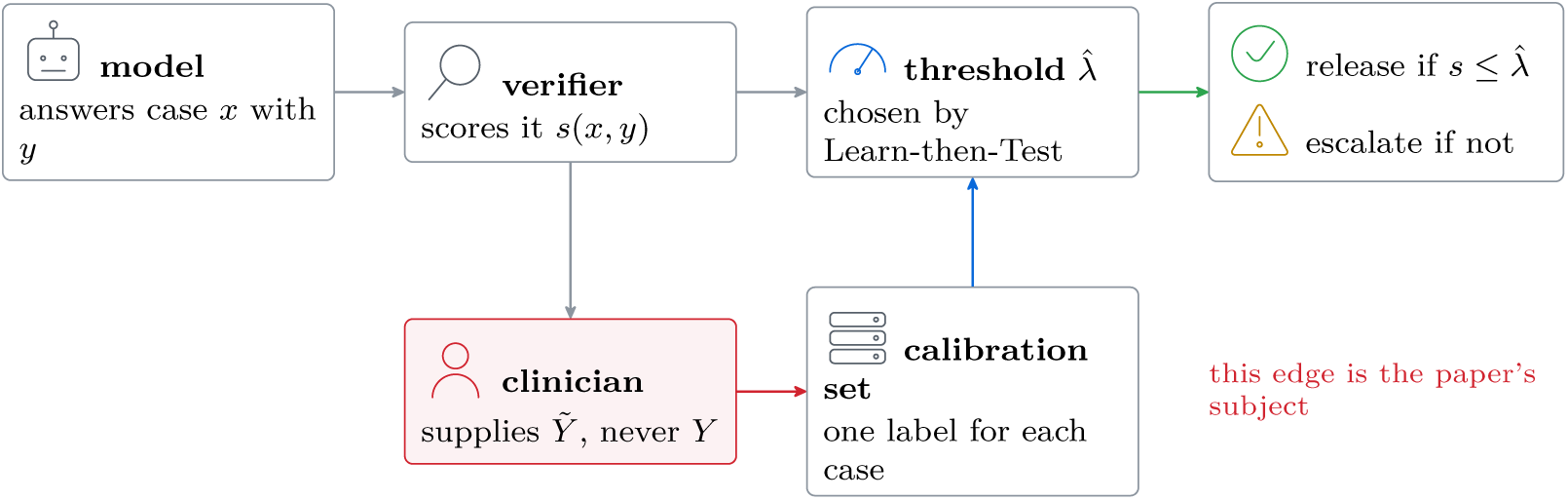
The arrangement, and the one edge this paper is about. The top row is the serving path in the order the code runs, from the model 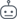 through the verifier 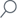 and the threshold 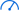 to release 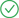 or escalation 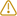. The row underneath is where 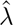 comes from, and it is a loop. Everything the certificate knows about safety arrives along the red edge, where the clinician 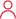 supplies a label, never the truth.

### The label the certificate never sees

The guarantee is stated in terms of *Y*, and *Y* is not observed. The observed variable is *Ŷ*, a clinician’s judgement, whose miss rate is *m* = Pr[*Ŷ* = 0 *| Y* = 1] and whose false-alarm rate is *ϕ* = Pr[*Ŷ* = 1 *| Y* = 0]. A false alarm costs clinician time and makes the certificate conservative. A miss does the opposite: it enters calibration as a clean label, the estimated risk at every threshold falls, 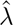 rises, and more is released. Throughout this paper we set *ϕ* aside and study *m* alone. That is the adversarial choice, because *ϕ >* 0 can only tighten the certificate we are arguing is loose.

Follow one missed defect around Fig. 3 and the asymmetry is visible without any algebra. The clean label enters calibration carrying the low nonconformity score the verifier gave it, because a defect a clinician approved is by selection one the verifier also found unremarkable. Every other error in this pipeline is corrected toward caution by the reviewer. This one is corrected toward permissiveness by the same reviewer, and the loop has no restoring term.

**Figure 3.**
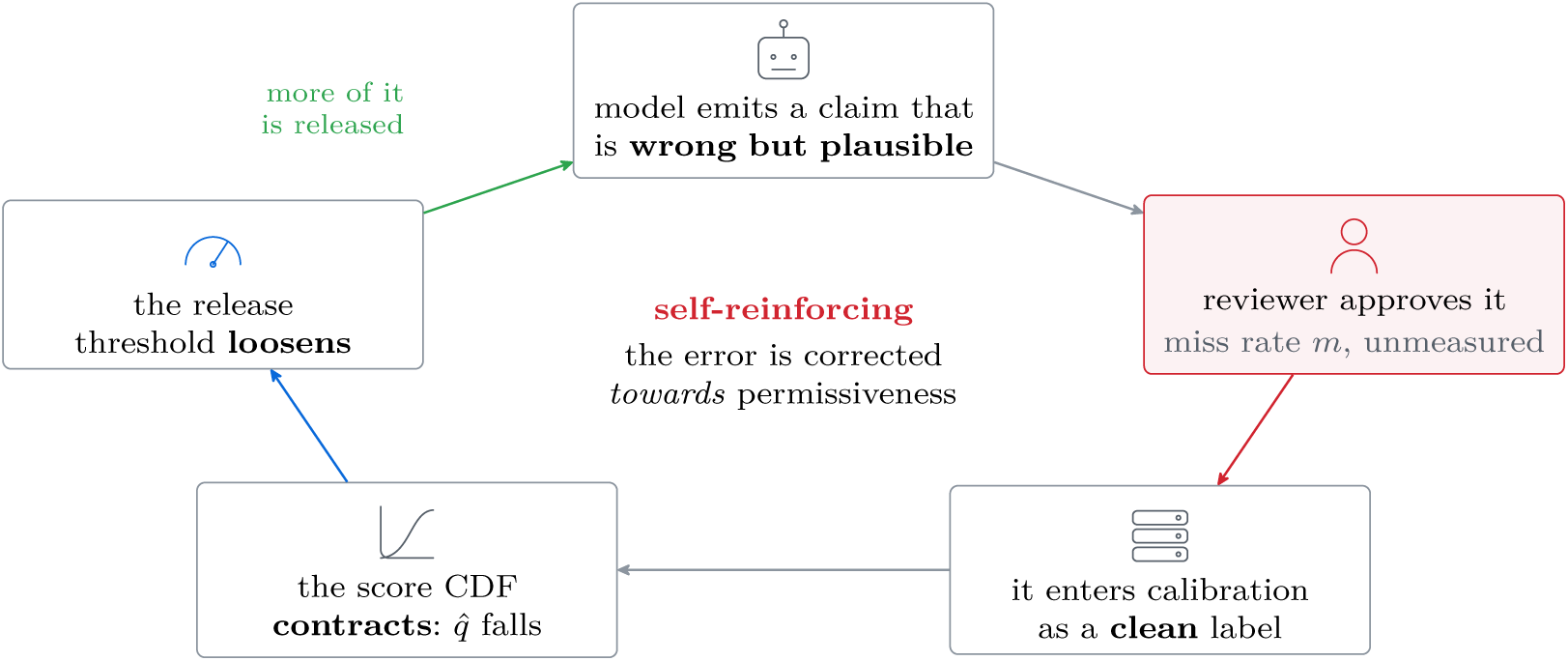
The reviewer miss is the one error in this pipeline whose correction points the wrong way. Read the cycle clockwise from the top. The node outlined in red is the hinge: a reviewer who misses a defect does not record *covered* incorrectly, they file a *clean* label for a case that was an error. That label carries a low score into calibration, the threshold loosens, and the green arrow releases more of exactly the answers that fed it.

Section O counts the same mechanism out in whole cases instead of a rate, at each of four reviewer profiles.

### The correction the deployment needs, and the instrument it can run

To restore the guarantee, a deployment must tighten *α* using an upper bound on *m*. The instrument available is the seeded probe. Insert *k* defects of known type into the review queue, count how many come back caught, and take a Clopper-Pearson upper bound on the share missed (Clopper & Pearson, 1934). Call the result *m̂* _probe_. The parameter the correction requires is *m*_natural_, the miss rate over the defects the model itself produces.

Our library implements the probe path in src/oversight/vigilance/probes.py, where converting the first quantity into the second requires the caller to declare a realism_penalty greater than one. Section 6 reports what we did about that parameter.

## 3. The contamination is one-directional and contractive

### What the published corrections require

Conformal coverage is robust to label noise under a condition on the score distributions rather than on the noise rate. Einbinder et al. show that coverage survives when the contaminated nonconformity-score distribution stochastically dominates the clean one, and that it degrades when the contamination pulls scores down (Einbinder et al., 2024). Sesia et al. give the coverage of a noisy-label conformal set as the nominal level plus an expected correction free to take either sign, so that noise is not a conservative perturbation (Sesia et al., 2025). Corrections of this family additionally assume the noisy label is conditionally independent of the input given the true label, *Ŷ ⊥ X | Y* (Penso et al., 2025). Both requirements fail for a clinical reviewer, and for one reason. Whether the reviewer misses a defect depends on how the case reads, so *Ŷ* depends on *X* given *Y*, and the defects that get missed are the ones that read well, and the same property gives them a low verifier score. The dependence is the mechanism itself.

#### What our reviewers produce

Figure 4 shows the two distributions and their difference for four reviewer profiles, computed by the simulation in examples/reviewer_certificate_demo.py. Contamination enters as follows: a fraction *p* = 20% of cases carry a real defect and a fraction *m* of those are missed, so missed defects make up a share *mp* of the sample, and each of them enters the calibration set carrying a score drawn from [0*, c*] with *c* = 0.25, the plausible range that made it approvable. A clean case carries a score drawn from [0, 1].

**Figure 4.**
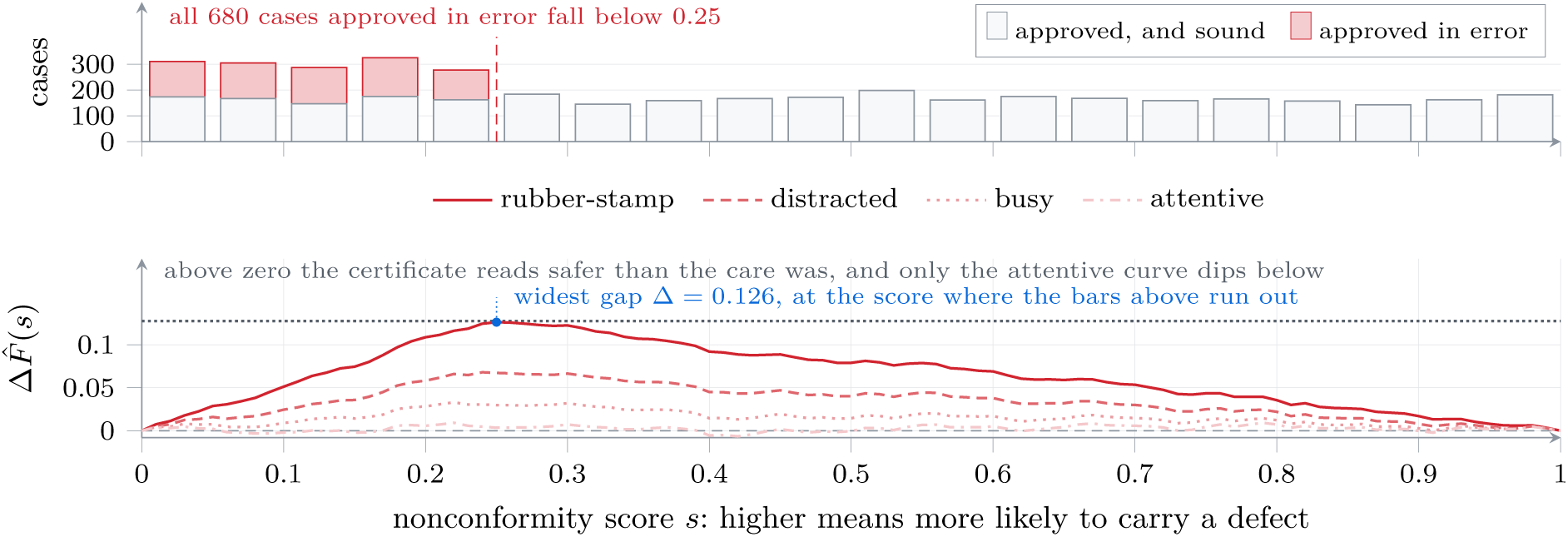
Where the contamination arrives, and why that is the regime published corrections exclude. 4,000 simulated scores at the worst profile. *Top:* approved cases in bins of 0.05, split into the ones the reviewer was right to approve and the ones they were not. *Bottom:* the resulting gap between the reviewer’s score distribution and the adjudicated one, for all four profiles. At every profile a test can resolve it never falls below zero, where a licensed correction requires it negative (Einbinder et al., 2024), and on the worst profile it is widest at the score where the bars above it run out.

The gap this opens needs no machinery to see. Pick any score *t* and ask what share of each sample falls below it. Among clean cases the answer is *t*, because the scores are spread evenly over the whole unit interval. Among missed defects the answer is *t/c*, because the same cases are crowded into the bottom *c* of it, and *t/c* is the larger of the two. So the contaminated sample carries an excess below every threshold, and the excess is the share of the sample that is contaminated, *mp*, times the amount by which that share was displaced, *t/c − t*. That displacement grows with *t*, is largest at *t* = *c*, where every missed defect has been counted once, and falls away above it. Its value there is the widest vertical distance between the two curves. We call it Δ:

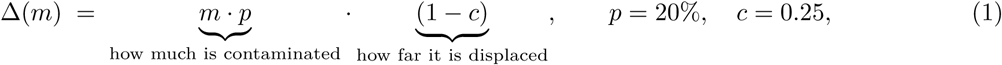

which is what contamination_gap computes and what Table 1 reports, under a floor of 10*^−^*^3^ that binds at no profile here, the smallest gap being 0.0045.

**Table 1.** No reviewer profile lands in the regime a correction requires. Contamination is the miss rate times the real defect rate, Δ follows Eq. (1), and inconclusive marks a gap a diagnostic of 4,000 cases cannot resolve. The last line is the control: shift the same contamination upward instead of downward and the same diagnostic does return conservative.

| Reviewer | Miss rate $m$ | Contamination | Planned $\Delta$ | Pairs needed | Diagnostic verdict |
| --- | --- | --- | --- | --- | --- |
| Rubber-stamp | 85% | 17.0% | 0.1275 | 185 | `anti_conservative` |
| Distracted | 45% | 9.0% | 0.0675 | 658 | `anti_conservative` |
| Busy | 20% | 4.0% | 0.0300 | 3,329 | `anti_conservative` |
| Attentive | 3% | 0.6% | 0.0045 | 147,938 | `inconclusive` |
| Dispersive control: contamination shifted *up* by 0.25 instead of down gives `conservative`. |  |  |  |  |  |

**Table 2.**
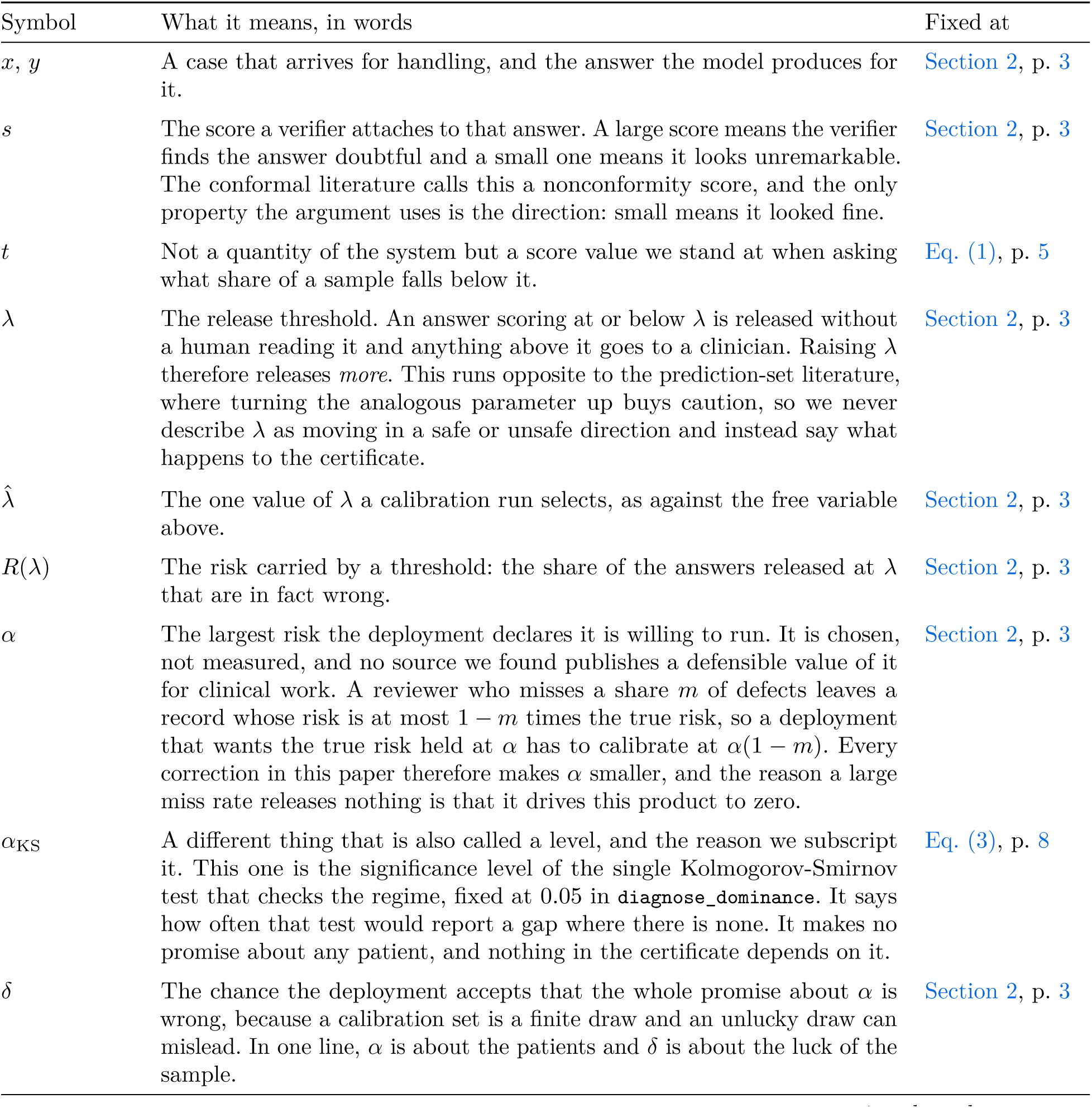
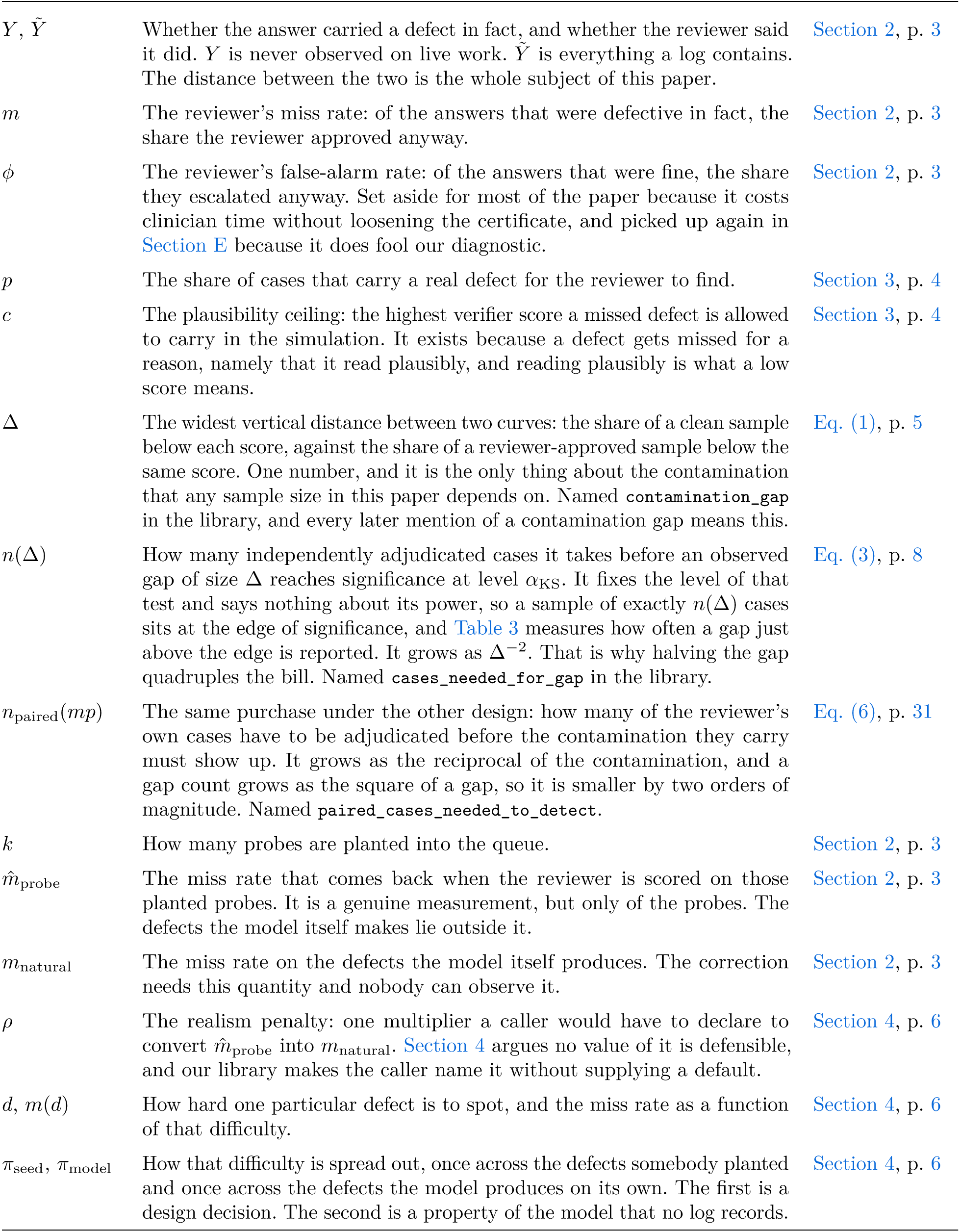
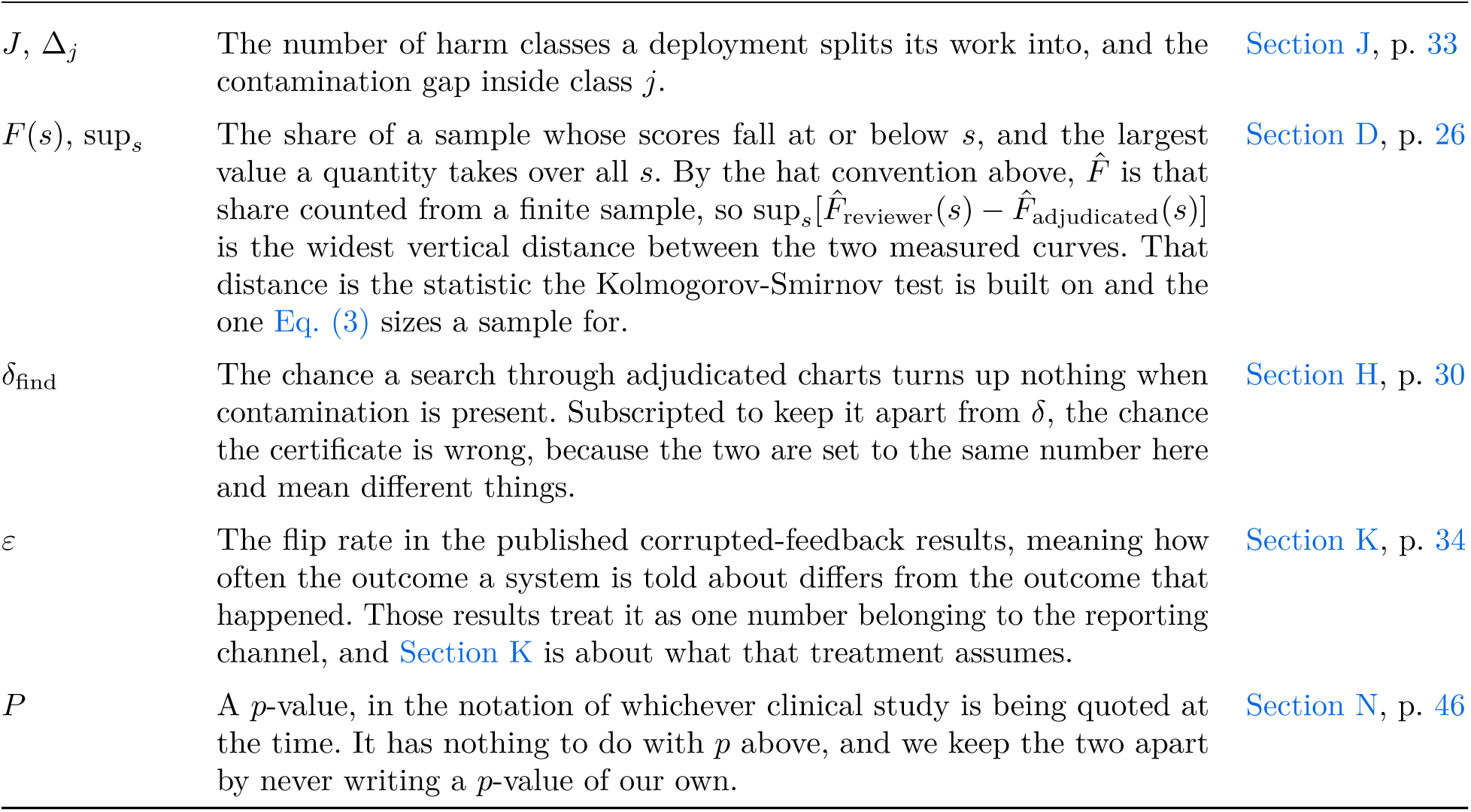
The symbols, in the order the argument introduces them. Two conventions hold throughout. A hat marks a number computed from a finite sample, never the quantity it estimates, so *m* is a property of a reviewer and *m̂* is a property of a measurement of that reviewer. A tilde marks what a log records as against what was true, so *Y* is the world and *Ŷ* is the record of it.

| Symbol | What it means, in words | Fixed at |
| --- | --- | --- |
| $x, y$ | A case that arrives for handling, and the answer the model produces for it. | Section 2, p. 3 |
| $s$ | The score a verifier attaches to that answer. A large score means the verifier finds the answer doubtful and a small one means it looks unremarkable. The conformal literature calls this a nonconformity score, and the only property the argument uses is the direction: small means it looked fine. | Section 2, p. 3 |
| $t$ | Not a quantity of the system but a score value we stand at when asking what share of a sample falls below it. | Eq. (1), p. 5 |
| $\lambda$ | The release threshold. An answer scoring at or below $\lambda$ is released without a human reading it and anything above it goes to a clinician. Raising $\lambda$ therefore releases *more*. This runs opposite to the prediction-set literature, where turning the analogous parameter up buys caution, so we never describe $\lambda$ as moving in a safe or unsafe direction and instead say what happens to the certificate. | Section 2, p. 3 |
| $\hat{\lambda}$ | The one value of $\lambda$ a calibration run selects, as against the free variable above. | Section 2, p. 3 |
| $R(\lambda)$ | The risk carried by a threshold: the share of the answers released at $\lambda$ that are in fact wrong. | Section 2, p. 3 |
| $\alpha$ | The largest risk the deployment declares it is willing to run. It is chosen, not measured, and no source we found publishes a defensible value of it for clinical work. A reviewer who misses a share $m$ of defects leaves a record whose risk is at most $1 - m$ times the true risk, so a deployment that wants the true risk held at $\alpha$ has to calibrate at $\alpha(1 - m)$. Every correction in this paper therefore makes $\alpha$ smaller, and the reason a large miss rate releases nothing is that it drives this product to zero. | Section 2, p. 3 |
| $\alpha_{\text{KS}}$ | A different thing that is also called a level, and the reason we subscript it. This one is the significance level of the single Kolmogorov-Smirnov test that checks the regime, fixed at 0.05 in `diagnose_dominance`. It says how often that test would report a gap where there is none. It makes no promise about any patient, and nothing in the certificate depends on it. | Eq. (3), p. 8 |
| $\delta$ | The chance the deployment accepts that the whole promise about $\alpha$ is wrong, because a calibration set is a finite draw and an unlucky draw can mislead. In one line, $\alpha$ is about the patients and $\delta$ is about the luck of the sample. | Section 2, p. 3 |
| $Y, \tilde{Y}$ | Whether the answer carried a defect in fact, and whether the reviewer said it did. $Y$ is never observed on live work. $\tilde{Y}$ is everything a log contains. The distance between the two is the whole subject of this paper. | Section 2, p. 3 |
| $m$ | The reviewer’s miss rate: of the answers that were defective in fact, the share the reviewer approved anyway. | Section 2, p. 3 |
| $\phi$ | The reviewer’s false-alarm rate: of the answers that were fine, the share they escalated anyway. Set aside for most of the paper because it costs clinician time without loosening the certificate, and picked up again in Section E because it does fool our diagnostic. | Section 2, p. 3 |
| $p$ | The share of cases that carry a real defect for the reviewer to find. | Section 3, p. 4 |
| $c$ | The plausibility ceiling: the highest verifier score a missed defect is allowed to carry in the simulation. It exists because a defect gets missed for a reason, namely that it read plausibly, and reading plausibly is what a low score means. | Section 3, p. 4 |
| $\Delta$ | The widest vertical distance between two curves: the share of a clean sample below each score, against the share of a reviewer-approved sample below the same score. One number, and it is the only thing about the contamination that any sample size in this paper depends on. Named `contamination_gap` in the library, and every later mention of a contamination gap means this. | Eq. (1), p. 5 |
| $n(\Delta)$ | How many independently adjudicated cases it takes before an observed gap of size $\Delta$ reaches significance at level $\alpha_{KS}$. It fixes the level of that test and says nothing about its power, so a sample of exactly $n(\Delta)$ cases sits at the edge of significance, and Table 3 measures how often a gap just above the edge is reported. It grows as $\Delta^{-2}$. That is why halving the gap quadruples the bill. Named `cases_needed_for_gap` in the library. | Eq. (3), p. 8 |
| $n_{\text{paired}}(mp)$ | The same purchase under the other design: how many of the reviewer’s own cases have to be adjudicated before the contamination they carry must show up. It grows as the reciprocal of the contamination, and a gap count grows as the square of a gap, so it is smaller by two orders of magnitude. Named `paired_cases_needed_to_detect`. | Eq. (6), p. 31 |
| $k$ | How many probes are planted into the queue. | Section 2, p. 3 |
| $\hat{m}_{\text{probe}}$ | The miss rate that comes back when the reviewer is scored on those planted probes. It is a genuine measurement, but only of the probes. The defects the model itself makes lie outside it. | Section 2, p. 3 |
| $m_{\text{natural}}$ | The miss rate on the defects the model itself produces. The correction needs this quantity and nobody can observe it. | Section 2, p. 3 |
| $\rho$ | The realism penalty: one multiplier a caller would have to declare to convert $\hat{m}_{\text{probe}}$ into $m_{\text{natural}}$. Section 4 argues no value of it is defensible, and our library makes the caller name it without supplying a default. | Section 4, p. 6 |
| $d, m(d)$ | How hard one particular defect is to spot, and the miss rate as a function of that difficulty. | Section 4, p. 6 |
| $\pi_{\text{seed}}, \pi_{\text{model}}$ | How that difficulty is spread out, once across the defects somebody planted and once across the defects the model produces on its own. The first is a design decision. The second is a property of the model that no log records. | Section 4, p. 6 |
| $J, \Delta_j$ | The number of harm classes a deployment splits its work into, and the contamination gap inside class $j$. | Section J, p. 33 |
| $F(s), \sup_s$ | The share of a sample whose scores fall at or below $s$, and the largest value a quantity takes over all $s$. By the hat convention above, $\hat{F}$ is that share counted from a finite sample, so $\sup_s[\hat{F}_{\text{reviewer}}(s) - \hat{F}_{\text{adjudicated}}(s)]$ is the widest vertical distance between the two measured curves. That distance is the statistic the Kolmogorov-Smirnov test is built on and the one Eq. (3) sizes a sample for. | Section D, p. 26 |
| $\delta_{\text{find}}$ | The chance a search through adjudicated charts turns up nothing when contamination is present. Subscripted to keep it apart from $\delta$, the chance the certificate is wrong, because the two are set to the same number here and mean different things. | Section H, p. 30 |
| $\varepsilon$ | The flip rate in the published corrupted-feedback results, meaning how often the outcome a system is told about differs from the outcome that happened. Those results treat it as one number belonging to the reporting channel, and Section K is about what that treatment assumes. | Section K, p. 34 |
| $P$ | A $p$-value, in the notation of whichever clinical study is being quoted at the time. It has nothing to do with $p$ above, and we keep the two apart by never writing a $p$-value of our own. | Section N, p. 46 |

One simplification runs against us. A calibration set holds only what the reviewer approved, and the defects they caught never reach it, so missed defects occupy a share of a real approved set larger than *mp* by the factor 1*/*(1 *− p* + *mp*). Every gap here uses the smaller share. Our gaps are therefore lower bounds, the sample sizes that follow from them upper bounds, and the price of the honest alternative is if anything overstated.

### What the verdicts do when only the seed changes

Every score in the diagnostic comes from a pseudo-random generator, so each verdict in Table 1 is one draw. We re-ran each profile at 400 seeds, changing nothing but the seed, and Section D reports every share with an exact Clopper-Pearson interval (Clopper & Pearson, 1934). Three profiles reproduce their verdict in every draw. The busy reviewer comes out anti_ conservative in 332/400 draws and inconclusive in the rest, because its planned gap of 0.0300 sits only just above the 0.0274 that 4,000 cases resolve, so a single draw can fall short of it. Falling short costs detection only, because inconclusive withholds the certificate. Across all 1,600 draws the number that returned conservative was 0.

### The certificate is a valid interval for the wrong quantity

Nothing above says the arithmetic is wrong. The Clopper-Pearson interval on the reviewer’s labels is exact and Learn-then-Test controls the risk it was given, but both bound Pr[*Ŷ* = 1 *| s ≤ λ*], the rate at which a released answer is one the reviewer would have flagged. A patient cares about Pr[*Y* = 1 *| s ≤ λ*], and the two differ by exactly the mass the reviewer misses.

## 4. The probe does not identify the parameter

### Miss rate is a function of detectability

Wolfe et al. had observers search for targets at three prevalences, and the same observers missed 11%, 25% and 52% of the common, rarer and very rare targets (Wolfe et al., 2005). The observers held a separate decision threshold per target type, so one aggregate rate cannot summarise them, and a correction that tightens *α* by one number is correcting for an average over a mixture whose weights it does not know.

Clinical work gives the axis a definition. Bednarczyk et al. score generated clinical content on a detectability scale running from “immediately and universally obvious” at one end to content that goes unnoticed until it has changed care, even with the source to hand (Bednarczyk et al., 2026). With that scale in hand the miss rate is a function *m*(*d*) of detectability *d*, and the two quantities in play are

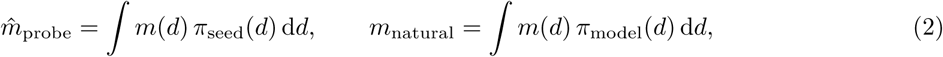

where *π*_seed_ is the detectability distribution of the planted defects and *π*_model_ that of the model’s own.

### The identifiability argument

A scalar realism penalty *ρ* asserts *m*_natural_ = *ρ · m̂* _probe_ for a *ρ* that can be fixed in advance. It holds when *π*_seed_ and *π*_model_ differ by a transformation whose effect on Eq. (2) is a constant factor. Two facts stand against fixing *ρ*, and Fig. 5 draws both on one axis. First, *π*_seed_ is chosen and cannot be otherwise, because to plant a defect a person must decide how hidden it is and that decision *is π*_seed_. Second, *π*_model_ is chosen by nobody: it is a property of the model, the prompt and the case mix, and it moves when any of the three moves. So a probe run reports the first integral in Eq. (2) and is silent about the second. That much is only a gap in coverage. The stronger statement follows, that collecting more of the same data does not close it.

**Figure 5.**
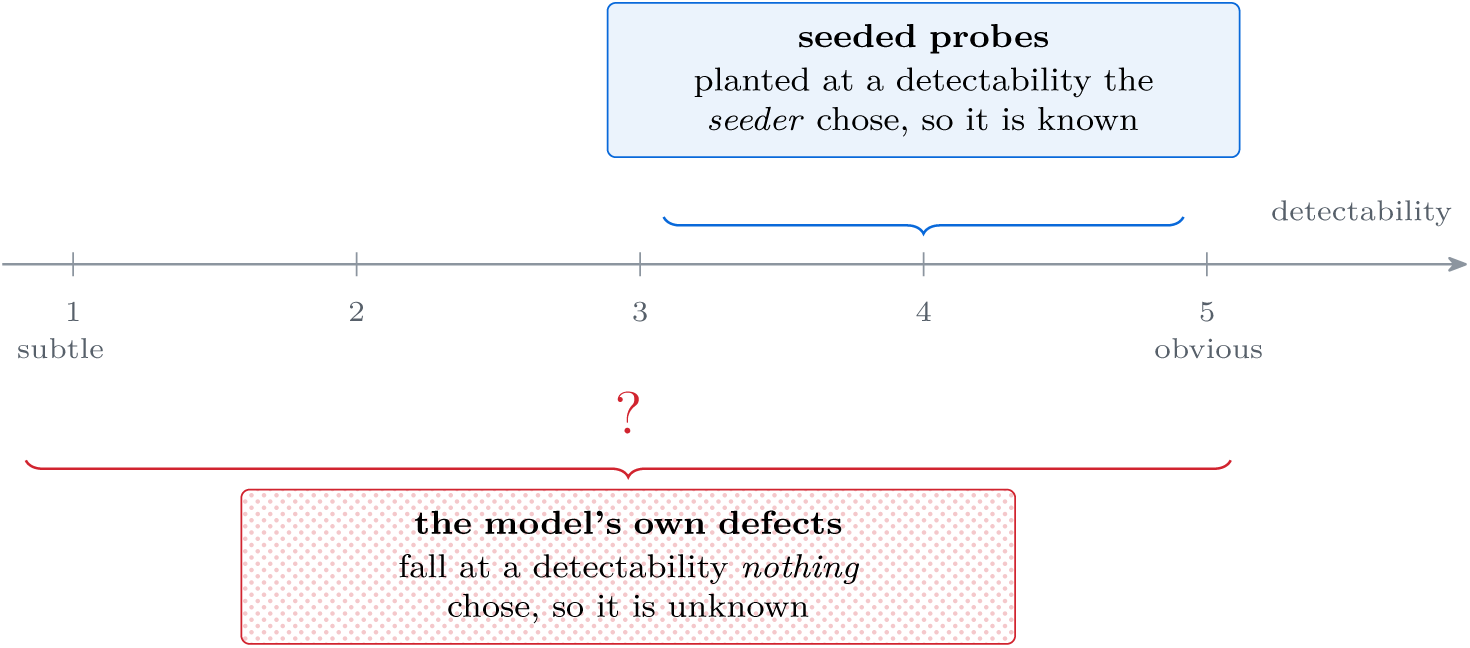
The probe and the target sit on the same axis and only one of them has a known position. Detectability runs from subtle to obvious, on the five-level clinical scale of Bednarczyk et al. (2026). The blue brace spans where seeded probes fall, a span known because the seeder chose it. The red brace spans the whole scale, because the model’s own defects can fall anywhere, and the question mark stands where a distribution would be drawn if we could draw it.

**Proposition 1.** With *ϕ* = 0, let a deployment see what a probe run sees: the share of seeded defects the reviewer missed, and the share of cases the reviewer flagged. Two deployments differing in both the real defect rate *p* and the miss rate *m*_natural_ can agree on both numbers while one of them releases (1 + *m*_natural_)*/m*_natural_ times as many defective answers as the other.

The proof is four lines of arithmetic and sits in Section B on page 24: the probe fixes one integral, the flag rate fixes one product, and doubling the defect rate while halving the share the reviewer catches leaves both alone. So *ρ* is unidentified, not unmeasured, and no amount of probe running tells the two deployments apart. Their score distributions do differ, and that difference has a stated limit. If the chance of a miss depended only on the verifier score, and probes covered every score at which the system’s own defects fall, the miss rate would be identified by weighting each flagged case by the inverse of its catch probability, as in positive-unlabeled learning with a known labelling probability (Bekker & Davis, 2018; Coudray et al., 2022). The result therefore needs the chance of a miss to vary among defects with the same score. A defect that reads as plausible to the verifier and to the reviewer alike is that case. Without the score route, the missing labels have to come from someone other than the reviewer. That is the expense Section 5 prices. Setting *ρ* conservatively does not repair the gap either. The certificate is then valid only if the unknown ratio falls under the chosen bound, an assumption about the model’s defects inside a procedure whose selling point is that it makes none.

### The seeded miss rate is a property of the seed set

A reader may grant all this and still expect the record to bound *ρ* from above, since probes are easier than real defects and only the size of the gap is open. The record does not support even that. Most of the studies that compare a seeded condition against a realistic one do find the seeded one easier, but one finds the reverse at *P* = .035 and one finds that denser seeding made detection worse. The sign is a tendency only, so a penalty set conservatively may be set conservatively in the wrong direction. Six published miss rates from one instrument and one disease span a range with no difference in reviewer attention behind it. Section L on page 37 is that record study by study, in Table 6 and Fig. 12. We quote no aggregate ratio from it, because averaging effects that disagree in sign would manufacture the constant we are arguing does not exist.

The same pattern is published for language models, and that brings the analogy home. On multiplication, Wu et al.’s synthetic errors covered 99.0% of gemma-3’s own error modes, and its recognition still fell from 83% on them to 20% on the errors it made itself. A second model on the same task fell from 100% to 78% (Wu et al., 2025). Matching the support of two distributions does not match the integrals taken against them. A seeder also cannot drift the seeded distribution toward the natural one, because the obstacle there is law and no matter of taste: in the one clinical domain where seeded probes are federally mandated, 42 CFR 493.945(b)(1) requires every slide preparation in a proficiency test set to carry unanimous agreement among at least three board-certified anatomic pathologists, and that agreement is needed only on one of four coarse categories, so the material that cannot clear even that bar is the equivocal material (Code of Federal Regulations, 2026). A certificate needs the level, and every source in Table 6 on page 39 supplies an ordering, a direction, or a level that belongs to a seed set someone chose.

### A probe can reject a certificate and never grant one

The record leaves a probe one use, and it runs in one direction. Suppose a deployment declares that the probe miss rate is a floor on the miss rate among the defects it releases, which cytology field validation arranges by discarding the slides readers miss most (Young et al., 2006). One-sidedness then bounds the true risk from below by the recorded risk divided by one minus that floor, and a reviewer who misses enough probes pushes that bound past the level and rejects the certificate. Nothing bounds it from above. The miss rate among released defects can approach one without contradicting any probe result, so the largest true risk consistent with the record is one, whatever the probes show (Stoye, 2022). Capping it would take a probe whose items are as hard to spot as the defects the system releases, and those are selected for being hard to spot by verifier and reviewer together. A probe is therefore a test that can fail a certificate and cannot pass one. That squares the result with laboratories that found probes predictive of routine work (Jenny & Jackson, 1993), since catching gross failure is the side a probe reaches. The library computes this identified set and has no code path on which a probe certifies (risk_ identified_set).

## 5. The price of the honest alternative

### Stop modelling the noise, and buy adjudicated labels

Adjudicated means labelled by a second, independent reader. Draw a sample of cases, have each adjudicated independently, and compare the score distribution of the approved cases against the adjudicated ones. If the approved distribution stochastically dominates, the published correction applies. That is a two-sample test on distribution functions, and the sample size at which an observed gap of Δ reaches significance at level *α*_KS_ follows from the Kolmogorov-Smirnov tail bound,

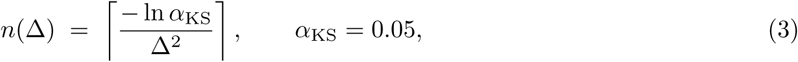

which cases_needed_for_gap computes in src/oversight/vigilance/dominance.py. The subscript separates this test’s level from the deployment’s risk level *α*. The gap enters squared, so halving it quadruples the charts a clinician must adjudicate.

### The asymmetry, and what it belongs to

Substituting Eq. (1) into Eq. (3) gives the cost of detecting each reviewer, plotted on logarithmic axes in Fig. 9 on page 33. Resolving the gap left by a reviewer who misses 85% of defects takes 185 adjudicated cases, and one who misses 3% takes 147,938, a ratio of about 800. Inverting Eq. (3) at a fixed budget states the same fact as a reach: our shipped diagnostic runs at 4,000 cases, resolves a gap of Δ *≥* 0.0274, and so reaches only a reviewer who misses at least 18.2%. That one number explains the inconclusive verdict in Table 1 without appeal to the simulation: the attentive reviewer’s contamination lies below the floor of the instrument aimed at it. The asymmetry belongs to the test. Section H on page 30 adjudicates the reviewer’s own cases instead of a separate sample, and the careful reviewer costs 498 cases instead, a factor of 297, because comparing two distributions has become counting one. We ship the separate sample because that is the design the published corrections are stated for, a limitation of what we built, and our measurement stands either way.

### One reviewer miss rate we could measure, and what it prices

A grid prices a range. One value of *m* can be measured instead. The HealthBench meta-evaluation publishes the grades 186 physicians gave to model answers on health questions, each answer judged against named criteria by at least two of them (Arora et al., 2025). Hold one physician out, let the majority of the others on that criterion stand as the reference, and a defect the majority recorded while the held-out physician did not is a miss by that physician, which peer_miss_rate computes. Physicians passed 257 of the 977 defects their peers agreed were present, a miss rate of 26.3%. Those grades cover 472 criteria on 352 answers from 134 conversations, so the interval resamples whole conversations and runs from 21.6% to 31.7%. Every one sits in HealthBench’s context-seeking and hedging themes, the only answers three or more physicians graded. Within them physicians passed 22 of 76 accuracy grades and 43 of 210 helpful-and-safe grades. They flagged 12.1% (11.0% to 13.3%) of the answers their peers had cleared. That rate falls inside the grid of Table 1 and prices the diagnostic at 1,925 cases in each arm, 1,564 to 2,398 at the ends of the Clopper-Pearson interval in Section M on page 43. A large service could buy that sample. So the obstacle is not the price. Among the 17 physicians who saw at least twenty such defects the rate ran from 8.0% to 54.5% on one task and one rubric. Part of that range is sampling noise, since physicians sharing one rate would still spread across 28.8% by chance, and each physician saw different answers, so how much of the rest belongs to the physician is not settled here. Across all disagreement in this corpus, who did the grading explains little (Borgohain & Mariathas, 2026). A deployment that borrows a value for *m* has borrowed one measured on other reviewers and other cases, this one included. Section M on page 43 gives the method, the digest of the release we read, and the three reasons this number is a floor.

### Where the money in an oversight pipeline goes

The usual way to make an answer look checked is to add reviewers: one model drafts, a second objects, a third settles it. That is the 3-model panel we ship. LangGraph makes each one a node in a graph and DSPy makes each one a declaration a compiler optimises (LangChain, Inc., 2026b; Khattab et al., 2024), and what a declaration costs to run is a property of the graph and never of the framework that declares it. One stage of our stack visits every claim against every document, so its count is a product: 15 claims by 19 documents is 285 calls from that stage alone. Re-pricing the same stack at a lean shape moves the bill by a factor of 60 and moving it to a self-hosted endpoint by 13, so shape dominates vendor. Table 11 on page 59 prices four shapes stage by stage, and Fig. 16 on page 48 separates what a runtime bounds from what it costs.

## 6. What the library does about it

A negative result about a shipped module raises a fair question: why keep the module. The measurement is useful and the correction is not, and Fig. 6 is how the code separates them.

**Figure 6.**
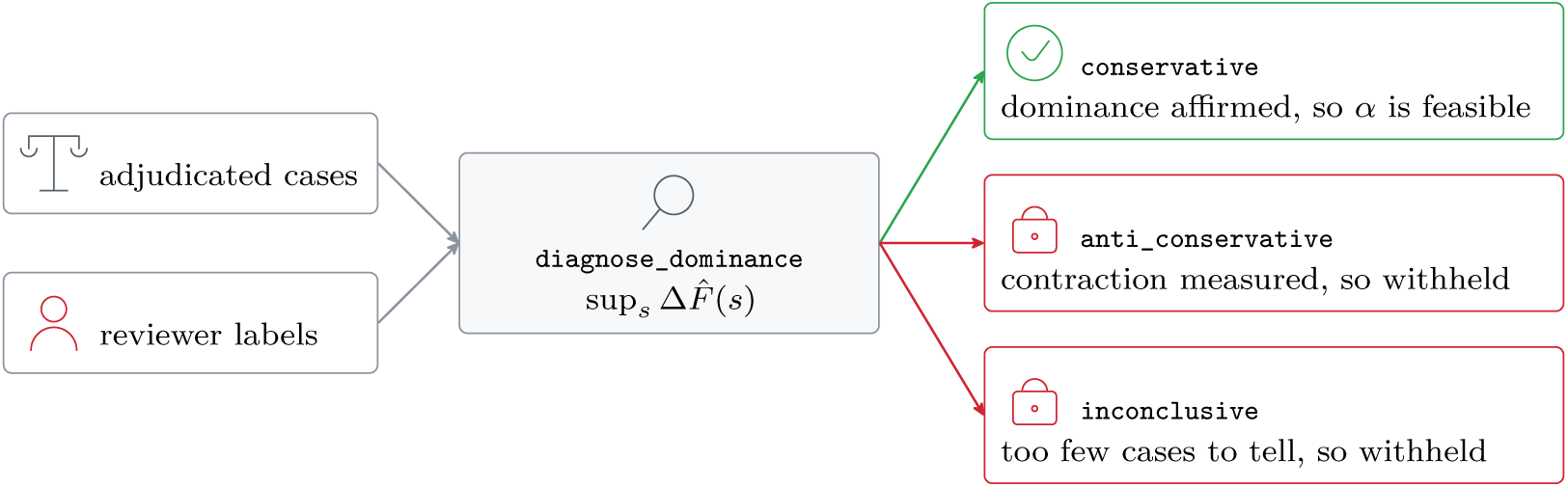
The correction is gated, and two of the three verdicts fail closed. `adjust_alpha` takes the diagnostic as a required argument, so the gate cannot be skipped by omission, and it refuses inconclusive on the same branch as anti_conservative 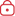, because absence of detected contraction is not evidence of dominance. Only conservative 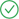 returns a feasible *α*, returned 0 times across 1,600 draws.

Failing closed is the discipline the library follows: an outage, a missing label, or an unmet precondition produces a result that says so, and never a clean pass. The probe path still reports a miss rate with an exact interval, per reviewer and per defect class, because a reviewer who misses a quarter of planted defects is worth a conversation whatever the realism gap. It no longer converts that measurement into a risk level. adjust_alpha survives as a function a caller reaches for by name, and no module calls it, so no certificate this library issues rests on a realism penalty. Where it still refuses a penalty at or below one, the refusal rests on failing closed alone, never on a direction Table 6 on page 39 leaves open, and a test over the library’s source, documentation and shipped examples holds it to that. The same principle governs this paper’s own numbers, which Section C and Table 20 on page 75 account for one at a time.

### What the library reads, and why one prompt is not enough

The reason a reviewer’s miss rate is not small is visible in the size of the thing being reviewed. The 20 synthetic charts this repository ships run 11,589 to 39,633 tokens each, at 3.89 characters to the token by the tokeniser, spread over 15 to 23 narrative documents apiece, 350 in all and a median of 17 per chart. Send one whole record in one prompt and any claim the answer makes can still be checked, because it is on the page. The claim it never made cannot be checked that way. Figure 18 on page 51 is the two readings side by side. OmissionDetector therefore spends one call per document instead of one per record, 17 for a chart of median size, and refuses to report an omission without a located span.

## 7. Related work

### Corrupted feedback, and the exogeneity the stochastic corrections need

Wang, Zecchin and Simeone bound miscoverage for online conformal prediction under corrupted coverage feedback at three levels of assumption, from one that assumes nothing about the corruption sequence to a stochastic one with an i.i.d. flip rate (Wang et al., 2026). This is the closest published work to ours and the relationship is layered: their general level covers a one-directional reviewer-miss sequence, and their probe-based resolution of the circularity and their online estimator for an unknown flip rate are theirs. We add the layer below. Their stochastic level requires the flip to be independent of the true miscoverage indicator, and that is neither a symmetry requirement nor an artefact of one proof technique. Xi et al. corrupt the label and leave the reported bit alone, so their induced flip is asymmetric and their correction survives it (Xi et al., 2025), and Section K locates the same independence step in four papers. Direction closes the published escape route as well, because under-coverage in Xi et al. needs the expected prediction set to reach *K*(1 *− α*) of the *K* labels, while a one-directional miss reports (1 *− m*)*α* with no dependence on *K* at all, and Feldman et al. exhibit the unsafe regime on real health expenditure data (Feldman et al., 2025).

### Label noise, wider certificates, probe realism

Three published corrections for label noise each need an input a deployment cannot obtain: stochastic dominance of the contaminated score distribution (Einbinder et al., 2024), a signed coverage correction (Sesia et al., 2025), and a conditional independence our setting violates (Penso et al., 2025). Our contribution against them is that the input cannot be had, not a new correction, and Section I covers two further lines that leave the origin of a calibration label alone. The empirical half is borrowed as openly: the vigilance literature supplies the mechanism behind a detectability-dependent miss rate (Wolfe et al., 2005; Evans et al., 2013), and Section L lists the seeded-defect measurements one at a time. We neither claim nor aggregate them, because they disagree in sign. We add the identifiability step that follows once a distribution-free certificate sits downstream of a reviewer, and that the fairness requirement written into federal proficiency-testing law is itself a truncation of the probe distribution (Code of Federal Regulations, 2026). The truncation is documented in the cytology record itself, which names the latent variable as the slides’ difficulty, keeps a slide only once readers agree on it, and measures that slides failing the rule were called negative 5.3% of the time against 1.2% for those that passed (Birdsong et al., 2006; Young et al., 2006). That record treats the truncation as fairness. Our addition is to read it as the reason the probe cannot be converted. The nearest formal precedent, that a false-negative rate estimated by rescreening negatives alone is a bounded function that does not recover the true rate, concerns a different design and a different mechanism (Renshaw, 1999). Commutability in clinical chemistry asks the same question of reference materials and answers it by measuring the same patient samples both ways, and that check is exactly what is unavailable here, because finding which real defects were missed is the target.

### Positive-unlabeled learning and spectrum bias

A missed defect recorded as safe makes the calibration labels positive-unlabeled data, and the reviewer’s catch rate is what that literature calls the label frequency (Elkan & Noto, 2008). It is identified when every defect is equally likely to be caught. When the chance depends on the case, the problem is called ill-defined, because an absent label can be explained by a low chance of a defect or a low chance of a catch (Bekker & Davis, 2018; Bekker et al., 2019; Bekker & Davis, 2020). Those papers state this in a sentence and move to assumptions that restore learning. Our proposition is the version of that remark for a probe set whose make-up a person chose. Its structure follows Link, who showed that unequal detection probabilities leave a population size unidentified in capture-recapture however large the sample (Link, 2003). That a test measured on one case mix misstates its accuracy on another is spectrum bias in diagnostic accuracy research (Ransohoff & Feinstein, 1978; Mulherin & Miller, 2002).

### Results that need no assumption on the noise

The bounds that assume nothing about the corruption hold in both directions and leave the sign open, in total variation for conformal prediction beyond exchangeability and as a two-sided Kolmogorov-Smirnov term under a contaminating mixture (Barber et al., 2023; Clarkson et al., 2024). The Huber form of the first inflates non-coverage by at most a factor of one over one minus the contaminating fraction, which bounds the risk on record from below. Our contraction bounds it from above, that a one-sided miss lowers the risk on record at every threshold. That is the part that fixes the sign, and its direction is the known lower bound on prevalence under a test with no false positives (Stoye, 2022). One-directional contamination of a calibration set is already shown to inflate error when the contaminating scores sit low (Bashari et al., 2025), which is the closest structural match to a reviewer who misses the defects that read well.

### Four standing routes to *m*, and where each stops

A reader who accepts that the planted defect cannot supply *m* will reach for one of four alternatives. *Reweight the calibration set.* Conformal prediction under covariate shift reweights each point by a likelihood ratio between the two distributions (Tibshirani et al., 2019), with bounds on what leaving exchangeability costs (Barber et al., 2023) and an online version that tracks a shift as it happens (Gibbs & Candès, 2021). All three need that ratio. Here it is the ratio of detectability densities over seeded and over natural defects, and the natural one lives only on the defects somebody noticed, so writing it down is writing *m* down. *Use several raters and no truth.* Latent class analysis recovers each rater’s error rate if their errors are conditionally independent (Dawid & Skene, 1979; Hui & Walter, 1980). That is the condition a clinical reviewer breaks hardest: a defect that reads well reads well to the second reader too, and such a model cannot tell correlated misses from a low prevalence. *Spend a few real labels well.* Prediction-powered inference draws valid conclusions from a small labelled sample beside a large unlabelled one (Angelopoulos et al., 2023), and risk control has been put on that footing (Einbinder et al., 2025). This is the strongest of the four, and the one we concede most to: its labelled sample comes from the same cases as the unlabelled one, so Section H prices it and Section 5 does not, and that price is low. *Defer instead of certifying.* Learning to defer optimises the joint human and machine system (Mozannar & Sontag, 2020), and a conformal set can aid without gating (Straitouri et al., 2023). Both are scored against labels, so deferring to the reviewer whose miss rate is unknown moves the problem without removing it.

### The name for what is left

A parameter that data constrains to a set but not to a point is partially identified, and the contaminated-sampling model in Eq. (1) is the classical case of it (Horowitz & Manski, 1995; Manski, 2003). We follow that literature’s discipline: report the bound the evidence supports and refuse the point estimate it does not. That is what adjust_alpha refusing to return a number amounts to. None of the distribution-free risk control we build on is in question here (Bates et al., 2021; Angelopoulos et al., 2024; 2025). Each holds conditional on the calibration labels being those named in the theorem, and this paper is about what happens when a human supplies them.

### Where the shipping tools sit

The guardrail and evaluation tools a team reaches for pass or fail an output against a threshold an author picks, and one of them documents picking it as the 75th percentile of observed scores (Confident AI, 2026). That is the same object as 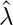 carrying a different warrant, a description of the scores already seen and no statement at level *δ* about an unseen one. Section R on page 50 searches six such trees at named commits and reports the difference as one of claim and never of defect (promptfoo, Inc., 2026; NVIDIA Corporation, 2026; Exploding Gradients, 2026; Guardrails AI, Inc., 2026; UK AI Security Institute, 2026).

## 8. Responsible use: what this does not show, and how it should be used

### Every reviewer number here is simulated except one

The four profiles, their contamination levels and the score distributions in Fig. 4 on page 5 come from examples/reviewer_certificate_demo.py under a stated contamination model. The grid those profiles sit on brackets 9 published human miss rates, 8.3% to 83%. That fixes its range without estimating any rate inside it, and Table 7 prices the diagnostic at every one of them. The one rate we measured ourselves is narrower than it looks. Physicians grading model answers passed 26.3% (21.6% to 31.7%) of the defects their peers agreed were there, but they were handed the criterion to check, easier than judging unaided whether an answer is safe to release, and none of them was in a clinic or answerable for the answer. Section M on page 43 gives the method and the three reasons that number is a floor under *m* for a task adjacent to ours rather than a value of *m* for ours. Every other number here prices a diagnostic under the model without measuring a clinical workflow, and every case count is an order of magnitude.

### The load-bearing gap in our own argument

The claim is about a level, and the ordering is already published. Fang et al. found that reviewers who caught more injected errors caught more natural ones (Fang et al., 2026), a correlation across seventeen reviewers that ranks them without fixing the rate at which any one of them misses a model-produced defect, and a certificate needs the rate. Publish the joint detectability distribution and *ρ* becomes derivable, weakening the claim from unidentified to expensively identified. It is the result here most likely to be overturned.

### Most of our probe-realism sources are analogies

Threat Image Projection, blood lead proficiency testing and seeded cytology slides all sit outside our domain, and none is a language model omitting a contraindication from a discharge summary, so we name the domain at every use and take direction, never magnitude. Wu et al. are the exception, measuring synthetic against on-policy errors in language models directly (Wu et al., 2025), and Section F gives three more.

### What the result is for, and what it must not be used to support

This negative result exists to stop one move: pricing a clinical distribution-free certificate with a seeded-probe miss rate scaled by a chosen realism factor. It is not licence to stop running probes. They remain a usable signal of reviewer attention. Nothing here establishes that any oversight arrangement is safe, nothing here is a claim about a device, and no regulator has reviewed this work.

### Clinical impact, and what we can mitigate

The failure is silent: a certificate that is internally valid and anti-conservative releases more unreviewed clinical text than its level implies. Our mitigation is the gate of Section 6, and Section E measures the two ways it can confirm wrongly. We have no cheap substitute for the independent adjudication Section 5 prices, and Section Q sets that price beside what published pipelines paid for the loops and panels a reader would reach for first.

## Data Availability

All data produced in the present study are contained in the manuscript or in the replication deposit at doi:10.5061/dryad.g79cnp66x, which holds the code, the generated score sets and the record of every run. The deposit is private for peer review, and the code version behind this revision, including the conversation-level interval and the criterion breakdown of the physician miss rate, will be added before it is released. The measured physician miss rates come from two third-party corpora, the HealthBench meta-evaluation and the CODE-test reader annotations, that are openly available from their original sources and are not redistributed here, so the deposit carries the loaders and the pinned checksums instead. No patient record was accessed at any point and no new human data were collected. Every reviewer figure reported from simulation is reproducible from the deposit under the seeds recorded there.

https://openaipublic.blob.core.windows.net/simple-evals/healthbench/2025-05-07-06-14-12_oss_meta_eval.jsonl

## A. Notation and borrowed terms

Each symbol below is defined where the argument first needs it, and this section collects the definitions in one place so that no reader has to hold nine pages in their head or open the source to recover one. The right column says where the symbol is fixed. Nothing here is new, and nothing here requires reading the library. Reading the library is for checking that we did what we say. That is a separate errand from following the argument.

The library also returns three verdicts and one refusal by name, and those names appear in the tables of this paper exactly as the code emits them, so that a reader who does go to the source finds the same word. The meanings are below, and they are about the *direction* the label noise pushes the certificate, never about whether the reviewer is any good.

- `conservative`. The reviewer-approved scores sit *higher* than the clean ones. That is what the literature means by saying one distribution stochastically dominates another. This is the one case in which the published label-noise corrections apply, because noise of this shape makes the certificate stricter than advertised. The library sets `certifiable` only here.
- `anti_conservative.` The reviewer-approved scores sit *lower* than the clean ones. That is what missing a plausible-looking defect does. The certificate is then looser than advertised by an amount nothing in the published corrections bounds. Every profile our simulation can see lands here, and that is the finding of Section 3.
- `inconclusive.` Neither direction reached significance, so the regime was not told apart from no gap at all. It never means that the two tests disagreed. The harmful direction is tested first and reported first, so if both were significant the answer would be `anti_conservative`. That is the fail-closed order. Read this as *not measured*, never as *safe*.
- `underpowered.` The tables print this as no test ran. It is not a fourth verdict but a flag carried alongside inconclusive. The sample on hand is smaller than *n*(Δ) demands for the gap the caller named, so the library declines to run the comparison at all, because any result it reported there would be unsupported, and it fills the gap it reports with the worst case, never with a measurement. This is the refusal Section 6 describes, and an empty cell in an attentive row of Table 3 records that refusal.

**Table 3.** Three verdicts survive a change of seed and the fourth says why it does not. Each profile was simulated at 400 seeds numbered from zero, with the reviewer model, the diagnostic size of 4,000 cases and *α*_KS_ = 0.05 held fixed. Column two is the count of draws whose verdict matched the one in Table 1 on page 6 over the count of draws taken, and column three is the two-sided Clopper-Pearson interval on that share. That interval is exact and therefore asymmetric and bounded. Columns four and five describe the statistic underneath the verdict, sup [*F̂* reviewer(*s*) *− F̂* adjudicated(*s*)], over the draws in which a test ran, and the fifth column is a standard deviation across those draws. It is not a standard error of their mean. The attentive rows read no test ran because the refusal in diagnose_dominance precedes the statistic: at that profile’s gap the diagnostic is underpowered in every draw, so there is no test result to average. The table reports variability under reseeding of one simulation and says nothing about variability across reviewers, case mixes or models, none of which this simulation varies.

| Reviewer | Draws agreeing | 95% interval on the share | Mean widest gap | Its spread |
| --- | --- | --- | --- | --- |
| Rubber-stamp | 400/400 | 0.991 to 1.000 | 0.1300 | 0.0094 |
| Distracted | 400/400 | 0.991 to 1.000 | 0.0713 | 0.0093 |
| Busy | 332/400 | 0.790 to 0.866 | 0.0362 | 0.0090 |
| Attentive | 400/400 | 0.991 to 1.000 | no test ran | no test ran |

Four more phrases are used throughout. *Exchangeability* is the sampling assumption the certificate keeps, and it is the only borrowed word in this paper that the argument cannot do without. It says the calibration cases and the cases still to come may be treated as a single pool that arrived in no particular order, so that shuffling the whole collection would leave the joint distribution of their scores as it was. Cases that are independent and identically distributed are exchangeable. The reverse does not follow, so exchangeability is the weaker of the two requirements, and the results we build on are published for the stronger one, independent draws from a single distribution. Either is enough for everything argued here. The assumption fails when the case mix drifts, when the model is replaced part-way through, or when today’s cases are correlated with yesterday’s, and the published relaxations of it are weighed in the main text. Nothing in this paper attacks exchangeability. The label is what we attack. The *approved arm* is the sample of cases the reviewer waved through. A real calibration set is made of exactly those cases, so it contains their misses. The *adjudicated arm* is the sample whose true labels were established independently of that reviewer, and that is the sample Section 5 prices. A *seed set* is one particular collection of planted defects, chosen by one particular person, and Section 4 is about why that choice does not cancel out.

## B. The two deployments a probe run cannot tell apart

This is the proof of Proposition 1, written out at length because the whole argument of the paper turns on it. Two deployments hand in the same probe numbers, and one of them is far more dangerous than the other. Measurement precision is not what separates them.

Write the two observables down. A probe run plants defects with detectability distribution *π*_seed_ and counts how many came back caught, so what it measures is *∫m*(*d*) *π*_seed_(*d*) d*d*, an average of the miss-rate function over the seeded detectabilities and nothing outside them. Counting escalations gives the share of cases the reviewer flagged, and with no false alarms a case is flagged only when it carries a defect the reviewer caught, so that share is *p* (1 *− m*_natural_). Two numbers, and among the unknowns are the defect rate *p*, the miss-rate function *m*, and the model’s own detectability distribution *π*_model_.

Now build the second deployment. Double the defect rate to 2*p* and let the reviewer’s miss rate rise from *m*_natural_ to (1 + *m*_natural_)*/*2, so the share caught, 1 *− m*_natural_, is halved. The flag rate is 2*p* times half of what it was, which is what it was. The probe count is untouched provided the rise in *m* is placed at detectabilities the seeder never used, and Fig. 5 is the picture of why that room exists: the blue brace is part of the scale and the red one is all of it. Two side conditions make the construction legal. The defect rate must satisfy 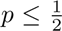, since 2*p* has to remain a probability, and that one is mild. The second needs care. Write *q* for the share of *π*_model_ lying outside the seeded span, and write *B* for the share of all cases that lie outside the span and are already missed, so that *B ≤ m*_natural_. The miss rate inside the span is held fixed by construction, so all the extra missing must happen outside it, and outside the span a miss rate cannot exceed one. That caps the extra at *q* and gives the requirement

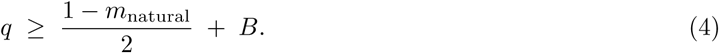

The first term is the extra miss mass the twin has to find and the second is the room already spent. Dropping *B* would state a condition that is necessary without being sufficient, and it is not a rounding detail: at *m*_natural_ = 0.06, *q* = 0.48 and an outside miss rate of 0.10 the shortened condition holds and no twin exists. A reviewer checking this construction should check Eq. (4) itself, never the shorter form, and the condition is easiest to satisfy where the argument matters most, because a reviewer who misses little leaves *B* small.

Count what each deployment releases. Defective answers escaping per case are the defect rate times the share missed: *p m*_natural_ in the first deployment and 2*p·* (1 + *m*_natural_)*/*2, which is *p* (1 + *m*_natural_), in the second. The ratio is (1 + *m*_natural_)*/m*_natural_, which is large exactly when the first deployment looks good. A reviewer who misses one defect in twenty has a twin who releases twenty-one times as much and files the same paperwork.

Two remarks keep the result in its place. First, adding false alarms back does not rescue the argument for the probe, because *ϕ >* 0 adds a third unknown to the same two equations. Second, the two deployments are not identical in every respect: their nonconformity score distributions differ, since a missed defect enters with a low score. Reading a miss rate off that difference is exactly the measurement Section 5 prices, and it needs labels the reviewer did not supply. The probe measures a different quantity, so it is no cheaper route to the same one.

### The share the gap is computed at

The same flag-rate accounting fixes a factor the main text states without deriving. A reviewer working through a stream whose defect rate is *p* catches and escalates *p* (1 *− m*) of the cases, so the calibration set that reaches the certificate is the remaining 1 *− p* (1 *− m*), or 1 *− p* + *mp*. The missed defects are *mp* of the original stream and all of them land in that set, so their share of it is

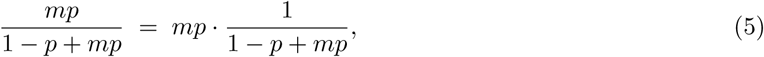

larger than *mp* by that second factor whenever *p > mp*, and that is whenever the reviewer catches anything at all. At the values used throughout, *p* = 20% and *m* at the busy profile’s 20%, the factor comes to 1.19, so the true contamination occupies about a fifth more of the approved set than the share we compute the gap at. Every gap in this paper uses the plain *mp*. That understates the separation between the two curves, so the gaps are lower bounds and the case counts they imply are upper bounds. The direction is safe for a deployment. A deployment budgets from our numbers and so buys more adjudication than it strictly needs, never less. It is not safe for the argument of this paper, and that has to be said in the same breath. A larger true contamination opens a wider gap, a wider gap is cheaper to detect, and the price of the honest alternative is the number our conclusion leans on. Correcting the share would therefore lower every count we quote. The factor enters the count squared, so at the busy profile the correction divides the requirement by about 1.19^2^, and it shrinks toward one as the miss rate rises or the defect rate falls. We leave the plain *mp* in place, because a reader who wants the corrected figure can divide by the factor above, and because no verdict in Table 1 on page 6 turns over under the correction. At the attentive profile the factor is 1*/*(1 *− p* + *mp*) = 1.24, and it lifts the gap from 0.0045 to 0.0056, still far under the 0.0274 that 4,000 cases resolve. The other three profiles were already resolved, and a wider gap only resolves them sooner.

## C. Reproducibility

Every number in this paper that describes our own system is emitted by a command, and there are five of them: four in the library, and one in this manuscript’s own tree that is named at the end of this list. The statistical argument comes from `.venv/bin/python examples/vigilance_report.py`, which imports examples/reviewer_ certificate_demo.py, runs the contamination model defined there, and writes the entire measurement to standard output as one JSON document: the four regime rows, the reseeding sweep behind each verdict, the score histogram, the cohort counts, the cost curve and the reach of a fixed diagnostic. The context sizes and every price come from `.venv/bin/python examples/context_report.py`. That command reads the chart corpus on disk and the priced configurations in examples/cost_model.py. The test count comes from pytest --collect-only, and the counts of files and lines from a walk of the source tree. Table 20 names, for every quantity in the paper, the command that produced it and the function inside that command. The fifth command is tools/census.sh. It produces Section R on page 50 and lives in this manuscript’s own tree, outside the library, because most of what it measures is other people’s repositories. No quantity here was typed in and then described as measured, and the manuscript will not typeset one that no generator emitted, nor a bare numeral in the text unless an exemption for it is written down.

The report is itself under test, and that matters more than the report existing. tests/test_vigilance_ report.py asserts the properties the argument rests on, never the values it happens to produce: that no draw of any profile returns conservative, that each verdict stated here is the modal verdict over reseedings, that the control which shifts contamination upward inverts the verdict, that no case approved in error scores above the plausibility ceiling, and that a smaller detectable gap never costs fewer adjudicated cases. Each of those was confirmed by breaking the code it covers and watching the assertion fail.

The simulated reviewer draws its scores from a generator seeded at 11, and every verdict the paper states from that simulation is also re-run at 400 seeds so that its variability under reseeding is reported alongside it, in Table 3. A reader who changes the seed reproduces a draw from the same distribution, and the same numbers do not come back. That is why the share of agreeing draws, never a single verdict, is the claim.

Measurements were taken on Python 3.12.9 on 2026-10-18. The library is 70 modules and 7,959 lines of source exposing 42 public symbols, with 5 required dependencies and 7 optional ones. The suite is 1,113 tests over 57 files and 11,074 lines, and the shipped evaluation corpus carries 20 chart fixtures.

### Limits of this reproduction

The library itself is not part of this submission, and no link to it is given, because its release date is not ours alone to set. The deposit the manuscript names carries the simulation code and the score sets every reported quantity is computed from, so every analysis in the paper can be repeated from that deposit alone. The deposit does not carry the library, so a reader cannot run the commands named above against our tree or read the code that does the accounting for the census and the cost model. A reader can check that the paper accounts for every number it states about our own system, and which command accounts for it. Table 20 names the generator of each one, the single exception being the census control, whose generator is named beside it.

### Use of language models in this work

No quantity in this paper was produced by a language model. Every number describing our own system comes from the seeded simulation, the library functions in Table 20 or the shell script behind Table 9, none of which calls a model, and every number from another study is quoted with its citation. A language model was used to draft and revise prose and the authors are responsible for all of it, and it acted as no reviewer, judge or label source in any measurement.

## D. Verdict stability under reseeding

Section 3 reports that three of the four reviewer profiles reproduce their verdict at every seed and that the busy profile does not. Table 3 is the measurement behind that statement, and its caption states the sweep and the estimator. The caption cannot say where the published draw sits inside the sweep: seed 11, the one Table 1 on page 6 is drawn at, is inside the swept range, so the verdict this paper prints is one of the draws the interval covers, never a draw held out from them.

The verdict is a threshold applied to a continuous statistic, so the statistic carries variability the verdict discards. The last two columns of Table 3 report the mean widest gap over the draws and its sample standard deviation across those draws. It is the standard deviation, because the question a deployment faces is how far its own single diagnostic can sit from the mean, not how well 400 draws pin the mean down. The three tested profiles agree on that spread to the third decimal while their means differ by a factor of nearly four. That is the behaviour of a two-sample Kolmogorov-Smirnov statistic whose dispersion is set by the sample size, and the size of the shift it is detecting does not enter. The attentive profile has no entry in those columns because no draw of it ran a test. Its refusal is raised before the statistic is computed, so its verdict is fixed by construction and the seed cannot move it. The interval on its share records that the construction held every time.

Two properties of the sweep bear on how the four rows may be read together. The simulation draws one adjudicated arm per seed and compares all four profiles against it, so at a given seed the reference arm is the same sample in all four rows, and the arm is identical across profiles draw for draw. That is deliberate, because it puts the four profiles on one shared reference, and it means the 400 draws behind each row are independent of each other while the four rows at one seed are not independent of one another. The consequence for the intervals in column three is that each is a valid interval on its own row’s share and the four are not simultaneous. A reader who wants a statement about all four at once should widen them, by Bonferroni or otherwise, and we do not make such a statement anywhere in the paper. 1,600 is therefore the number of profile draws, four per seed. The number of independent simulations is 400.

Figure 7 draws those four rows on one axis, and it adds a check the table cannot carry. diagnose_dominance decides on a two-sample Kolmogorov-Smirnov *p*-value that is the exact one at the equal arm sizes reported here, a property a test asserts while the call demands nothing, since forcing it would make an unequal-arm call on large samples arbitrarily slow. So the closed-form critical statistic behind Eq. (3) is an approximation to a boundary the sweep can locate directly. Among the busy profile’s draws the largest widest-distance value that came out inconclusive is 0.02725 and the smallest that came out anti_conservative is 0.02750. Those two differ by one step of the statistic. One step is all a widest distance over 4,000 scores per arm can move by, so the bracket is as tight as this sweep can make it. Equation (3) at the same size puts the critical gap at 0.0274, inside that bracket. The formula the cost curve is built on is therefore the number this simulation’s verdict changes at. That is a check on the two together, and it says nothing about reviewers. No other profile has such a bracket, because no other profile produced draws on both sides of it. That is also why the figure draws the full observed range: a range is what tells a reader whether a distribution reaches a threshold, and a mean with a standard deviation around it does not.

**Figure 7.**
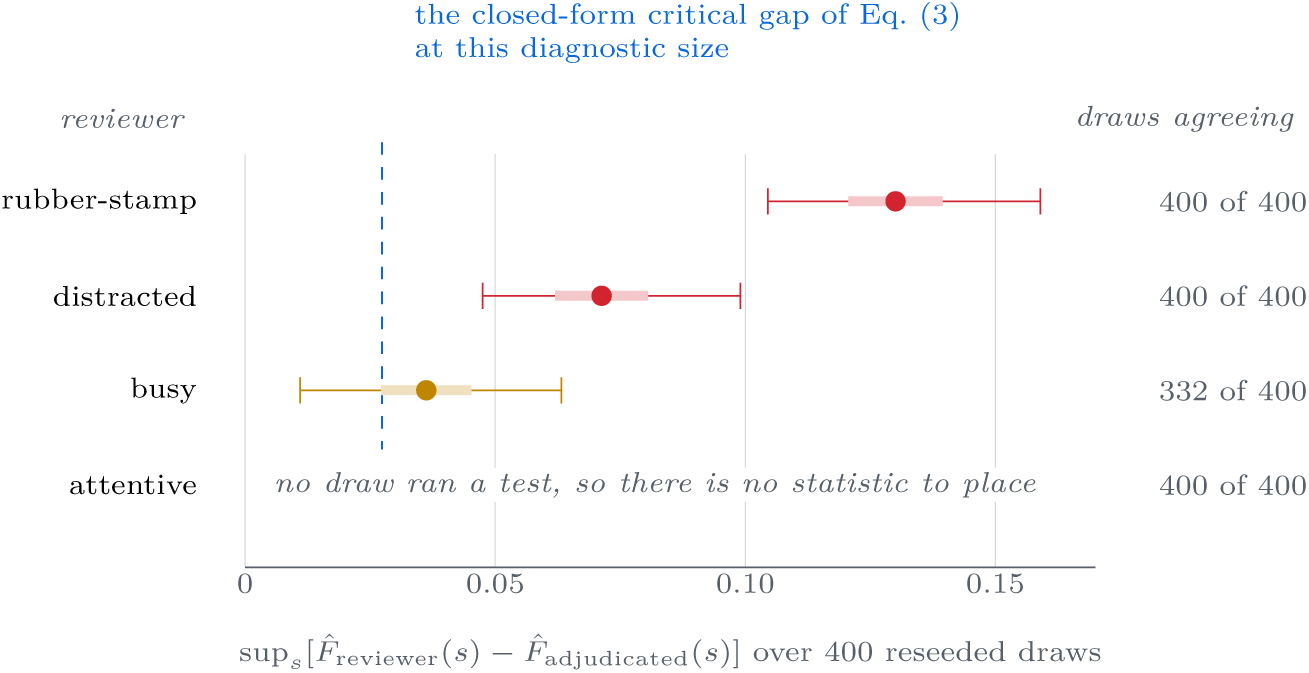
Mean widest gap by profile against the critical gap at 4,000 cases. Each row is a reviewer profile from Table 3 over the same 400 draws. The bar is one standard deviation either side of the mean widest gap, the capped line is the full observed range, and the dot is the mean. The dashed blue line is Eq. (3)’s critical gap at 4,000 cases. The busy row is amber and the only one whose range crosses the line. That crossing is the whole reason its verdict is not reproduced at every seed. The attentive profile has no mark at all: no draw of it ran a test, so there is no statistic to place.

The empty attentive row is a measurement. It is not a hole in the drawing. Every gap the report writes for that profile is empty, because no draw of it ran a test, and a zero on that axis would be a number no code path produced. So the figure cannot show where the attentive statistic would fall. That is the same absence Table 3 records as no test ran and the same one Section 5 prices at 147,938 cases.

The attentive line in Fig. 4 goes below zero. The four lines there are differences, each profile’s approved-arm curve minus its adjudicated-arm curve, and the other three stay at or above zero everywhere while the attentive line does not. Over the 101 scores the curves are compared at, the attentive difference is negative at 22 of them, and negative is the direction a licensed correction would need.

The sizes settle it. At its deepest the attentive difference falls 0.00650 below zero, and elsewhere on the same grid it rises 0.00925 above. The yardstick for a single score is the sampling error of the difference between the two curves at that one score. Write *F* for the height of the approved cu<u>rve there, *G* for the heigh</u>t of the adjudicated curve, and *n* for the 4,000 scores in each arm. That error is 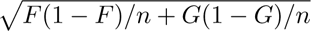, and it changes from one score to the next, so each excursion is measured against the error where it happens. The dip comes to 0.59 of its own error and the rise to 1.03. Over the whole grid the largest excursion of either sign is 1.90 of its own error, and that one lies above zero, away from the direction a correction would need.

The diagnostic also never reads that curve at this profile. Before comparing the two arms it asks how many cases the separation named for the profile would take, and Eq. (1) puts that separation for the attentive reviewer at 0.0045. That separation needs 147,938 cases in each arm. 4,000 is fewer, so the diagnostic refuses and reports no gap, and that is the empty entry named above. The claim in the main text is about a separation a test can resolve, and that separation is positive at every profile. The figure draws the raw height, and at one profile the raw height crosses.

## E. Two ways our own diagnostic affirms what does not hold

Section 6 treats diagnose_dominance as the mitigation, since it withholds the corrected certificate until the precondition is affirmed. A mitigation that can affirm wrongly is worth measuring, and this section measures it twice. Both measurements come from the same generator as the rest of the paper, at 100 draws per configuration.

### The per-test level and the uncorrected multiplicity

The call runs two one-sided Kolmogorov-Smirnov tests on the same pair of samples, one for each direction, each at *α*_KS_ = 0.05, and reports conservative only when the second is significant and the first is not. Under exact equality of the two arms, the null in which no label noise is present at all, the affirming verdict came out in 3/100 draws, a share whose exact interval is 0.006 to 0.085. That share is consistent with the level the test is built to have and is not a defect. Everything above the single call is a defect. Nothing corrects for the two tests sharing a sample, nothing corrects across the four profiles in Table 1 on page 6, and nothing corrects across repeated diagnostics on the same deployment over time. A team that reruns the diagnostic every month runs it twelve times a year, each run at *α*_KS_ = 0.05 on its own, and the library gives them no way to spend *α*_KS_ across those runs.

### False alarms lift the approved arm, and that is what dominance looks like

Section 2 sets the reviewer’s false-alarm rate *ϕ* aside on the grounds that *ϕ >* 0 can only tighten the certificate. That is true of the certificate and false of this diagnostic, and the distinction matters here. A false alarm escalates a clean case, so that case never reaches the approved arm. An independent catch by a careful reader looks like a low verifier score. If the falsely escalated cases are the ones the verifier scored low, then removing them lifts the approved arm’s score distribution, and a lifted approved arm is exactly the signature the test reads as benign noise. Table 4 is how much of that it takes. The contamination is held fixed in every row, so nothing about the harm changes across a row, only how much clean low-scoring mass the reviewer removed.

**Table 4.**
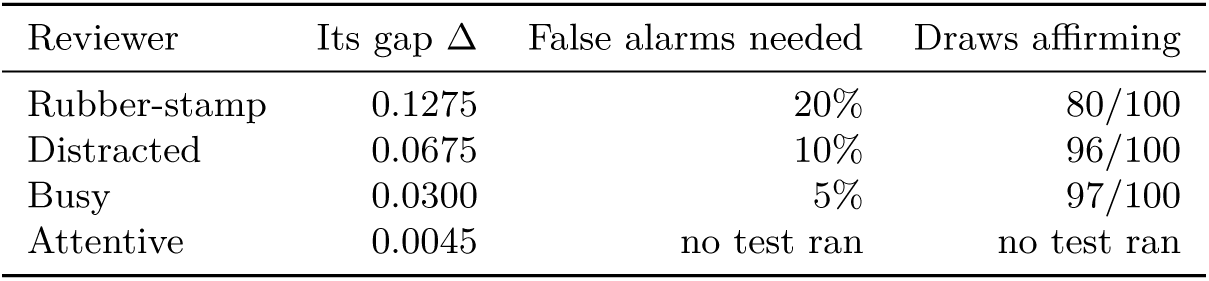
False alarms alone flip every tested profile’s verdict to conservative. Each row holds one profile’s contamination fixed and removes a share of the lowest-scoring clean cases from the approved arm, the way the reviewer’s own false alarms do. Column three is the smallest share in the swept grid 0%, 5%, 10%, 20% and 30% at which more than half the draws returned conservative, and column four is how many of 100 draws did. At a share of zero no tested row returns conservative in any draw, so the removal alone is what causes the flip. Reading down column three, the reviewer whose contamination is hardest to see is also the cheapest to disguise. The attentive row has no entry because no draw of it ran a test at any level, the same refusal Table 3 on page 26 records.

| Reviewer | Its gap $\Delta$ | False alarms needed | Draws affirming |
| --- | --- | --- | --- |
| Rubber-stamp | 0.1275 | 20% | 80/100 |
| Distracted | 0.0675 | 10% | 96/100 |
| Busy | 0.0300 | 5% | 97/100 |
| Attentive | 0.0045 | no test ran | no test ran |

The refusal inside `diagnose_dominance` catches some of this. It does not catch the part that matters. For the busy profile the largest shares strip so many cases from the approved arm that it falls below the sample its own gap requires, and the call returns inconclusive. The two loosest profiles are not caught that way. Their gaps need only 185 and 658 cases in each arm against the busy profile’s 3,329, so no share in the swept grid strips the approved arm below what they require, and at the largest share both still affirm in every one of 100 draws. The guard that stops the busy row is a sample-size check written for a different purpose, and it leaves untouched the rows where the contamination is worst. That is luck, and we do not present it as a safeguard.

This section narrows our own claim in Section 6. The diagnostic is necessary and it is not sufficient. A deployment that wants to rely on it has to bound *ϕ* as well as *m*, on the same independently adjudicated sample Section 5 prices, and nothing in this library bounds *ϕ* at all.

## F. Three mechanical limits and their directions

The main text defers three limits to this section, because each is a bounded technical qualification and no threat to the argument, and in each case the direction is the point.

### Equation (3) is asymptotic

It comes from a tail bound and holds in the limit. A parametrised test in our suite constructs samples at the predicted size, confirms that a two-sample test rejects there, and confirms that it fails to reject at a quarter of that size. That checks that the constant is not badly wrong in the regime we quote, without making the expression exact at small *n*. Were the constant wrong by a factor, every case count in Table 1 on page 6 would move by that factor together and the ratio between them would not move at all. The ratio is the claim. The expression also fixes only the level of the test, so *n*(Δ) is the size at which a gap of Δ arrives at the edge of significance. It is not the size at which that gap is sure to clear the edge. The busy profile measures the difference. Its planned gap of 0.0300 sits just above the 0.0274 that 4,000 cases resolve, and across 400 reseedings the diagnostic reported the contamination in 332/400 draws, short of all of them, as Table 3 records. A deployment that needs the gap found nearly every time should read *n*(Δ) as a floor and buy more than it.

### One reviewer and one defect class

We treat *m* as a scalar per reviewer for the cost calculation while arguing that it is a function of detectability. The simplification runs in the direction that favours the correction, since a per-class miss rate has more parameters to estimate and therefore a higher adjudication cost than the single number we price. Section J expands this into the classes a clinical deployment partitions on, ordered by the harm of the action being recommended, and finds that the cost lands hardest on the class a deployment would most want certified.

### One setting

Our certificate is a batch Learn-then-Test threshold held fixed, while the corrupted-feedback results we build on are long-run averages under a threshold that moves every round. Our negative result should not be read as applying to those procedures unchanged, nor their positive results as covering ours. The assumption audited in Section K carries across, because it constrains the corruption model and leaves the update rule alone.

## G. Sensitivity of the price to the two asserted numbers

Every case count in this paper comes out of one product. Equation (1) multiplies three numbers to get the gap Δ the diagnostic must be able to see, and Eq. (3) turns that gap into a sample size that goes as 1*/*Δ^2^. One of the three is the reviewer’s miss rate *m*, and the paper varies it across four profiles because varying it is the paper’s subject. The other two are not varied anywhere in the main text. They are the real defect rate *p*, the share of cases in which the model produces something that would change care if released, and the plausibility ceiling *c*, the top of the range a missed defect’s verifier score is drawn from. Both are ours by assertion. No study we have found measures either on generated clinical text, and Section L is the closest thing to evidence about the second. That is why the main text states them as inputs to a model and claims no finding for either.

A result proportional to an asserted number needs a sweep over that number, and squaring it makes the sweep compulsory. Figure 8 is that sweep. It holds the miss rate at the rubber-stamp profile, walks *p* across 11 values from 1% to 30%, walks *c* across 4 values up to 0.75, and asks the library’s own `cases_needed`_for_ gap for the sample size in every one of the 11 by 4 cells. The grid is required to contain the pair the paper uses, and the sweep refuses to run if it does not, because an interpolated marked point is the one way a figure like this misleads.

**Figure 8.**
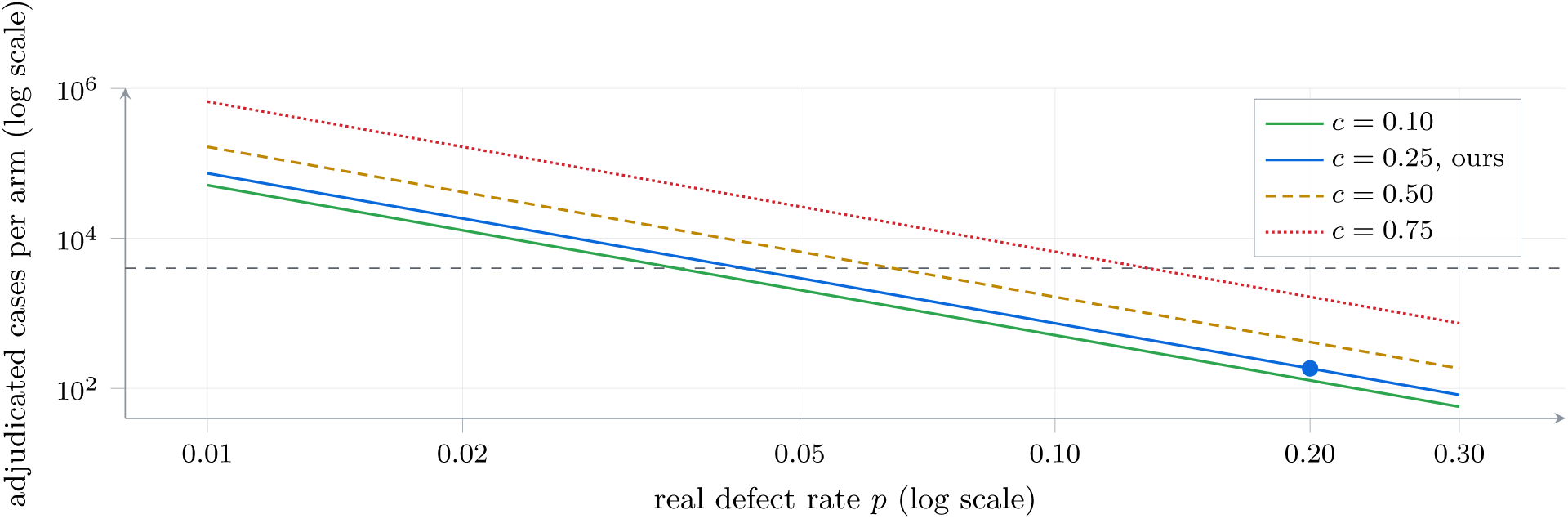
The price of the diagnostic against the two numbers the model asserts. Both axes are logarithmic, so each curve is Eq. (3) at one plausibility ceiling *c* and each is a straight line of slope *−*2. The miss rate is held at the rubber-stamp profile, which is the cheapest of the four, so every height on the plot is a floor. The filled dot is the pair this paper works at, a defect rate of 20% and a ceiling of 0.25, where the requirement is 185 cases in each arm. The dashed grey line is the 4,000 cases the shipped diagnostic is given, and a curve above it is a setting that diagnostic cannot resolve.

Read the figure by its slope first. Both parameters enter the gap linearly and the sample size as the inverse square, so halving either one costs a factor of four. Each parameter moves the price in the direction a deployment would rather it did not. A lower defect rate is the mark of a better model, and it makes the contamination rarer and therefore harder to see. A higher ceiling means the missed defects looked more ordinary to the verifier, and looking ordinary is what makes them dangerous. It crowds them less far below the threshold and so opens a narrower gap. Over the plotted grid the requirement runs from 57 cases to 663,415, a span of 11,639 at one profile, and the dot the paper works at sits near the cheap end of it.

The other three profiles are the same surface moved up. Only the product *mp* (1 *− c*) enters Eq. (3), so changing the miss rate multiplies every height by the square of the ratio and changes nothing about the shape. Going from the rubber-stamp profile to the attentive one multiplies the whole family by 803. The corner that follows is worth writing down. At the attentive profile, a defect rate of 1% and a ceiling of 0.75, confirming the condition takes 532,574,627 adjudicated cases in each arm. That number is what the model implies. Nobody would run that test. The simulation puts a floor under a planned gap so that its diagnostic is never asked to resolve nothing at all, and the corner sits below that floor, so we removed the floor to size it and did not quote the floor’s own answer.

None of this tells a deployment which *p* and which *c* it has. That is why we state the sweep and state no tolerance. The two numbers are properties of a model and a reviewer working together, they are measurable in principle by adjudicating a sample, and adjudicating a sample is the very purchase whose price they set. The argument of the paper does not turn on the cell. Contamination runs one way and contracts the recorded risk whatever *p* and *c* are, the score condition fails in the same direction at all four profiles, and the two deployments of Section B remain indistinguishable. The cell fixes only how much adjudication the honest alternative costs, and Fig. 8 is there so that a reader who disputes our two numbers can price their own without reading our code.

## H. The paired design and its cost

This section is the largest concession in the paper, and it is against our own headline number. The cost in Section 5 is the cost of one particular design, and a different design of the same purchase costs two orders of magnitude less.

### What the shipped diagnostic draws

`diagnose_dominance` compares two samples of scores and asks whether one stochastically dominates the other. Our example supplies them the way the test expects, and `simulate_review` in `examples/reviewer_certificate_demo.py` draws the adjudicated arm as its own set of cases, unconnected to the cases the reviewer approved. That is a faithful model of the published corrections. Those corrections are conditions on two score distributions and say nothing about which cases the scores came from. It is not a faithful model of how a hospital would buy adjudication. We expect a hospital to hand the adjudicator the charts the reviewer had already signed off.

### Adjudicating the reviewer’s own cases turns a comparison into a count

When the two labels sit on the same case, each adjudicated chart reports both what the reviewer said and what was true, so there is nothing to compare across distributions. The contaminated share of the approved stream is *mp*, the reviewer’s miss rate times the share of cases that carry a real defect, and it is now something to count. Write *δ*_find_ for the chance we accept of looking and finding nothing when something is there. The subscript is there because *δ* on its own already means the chance the certificate itself is wrong, and these are two different things that happen to be set to the same number. Then the smallest number of adjudicated charts that leaves at most that chance is

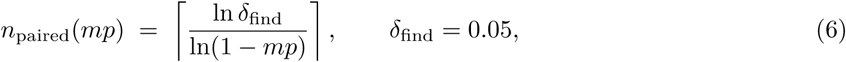

which `paired_cases_needed_to_detect` computes in the same module as Eq. (3). It takes that number as a required argument and supplies no default from the one beside it, for the same reason the subscript is written. The contamination enters once here and squared there, and that single difference in exponent is the whole saving. Table 5 is the comparison profile by profile.

**Table 5.** The same purchase priced two ways. The middle column is Eq. (3), one arm of a two-sample test against a separately drawn sample. The next is Eq. (6), the same contamination found by adjudicating the reviewer’s own cases. The last column is the number that pins the contamination down to a width of 0.01 at 95% confidence, a stronger purchase than finding it at all, and its ordering is the reverse of every other column in this paper.

| Reviewer | Contamination $mp$ | Separate arm | Paired, to find | Paired, to pin down |
| --- | --- | --- | --- | --- |
| Rubber-stamp | 17.0% | 185 | 17 | 21,881 |
| Distracted | 9.0% | 658 | 32 | 12,787 |
| Busy | 4.0% | 3,329 | 74 | 6,114 |
| Attentive | 0.6% | 147,938 | 498 | 1,156 |

### What the table says, including one result against us

First, the careful reviewer stays the expensive one to find, 498 charts against 17, so the direction of the asymmetry survives. Second, its size does not: the ratio falls from 800 to 29, and the attentive reviewer’s own bill falls by a factor of 297. 498 adjudicated charts is a small purchase, well short of a research programme, so the sentence in our abstract about affordability is a sentence about a two-sample test and we have scoped it to say so. Third, and this is the one that runs hardest against us, the last column reverses the order. Pinning the contamination down to a fixed width costs 1,156 charts for the careful reviewer and 21,881 for the rubber stamp, because a rare event has a narrow interval around it in absolute terms, and a fixed absolute width is what `probes_needed_for_width` is asked to buy, with the contamination *mp* passed to it as the rate to plan against. Under this design the careful reviewer is the cheap case. Fig. 9 on page 33 is a statement about a test on distribution functions, and any reading of it as a statement about the problem is a reading we withdraw.

**Figure 9.**
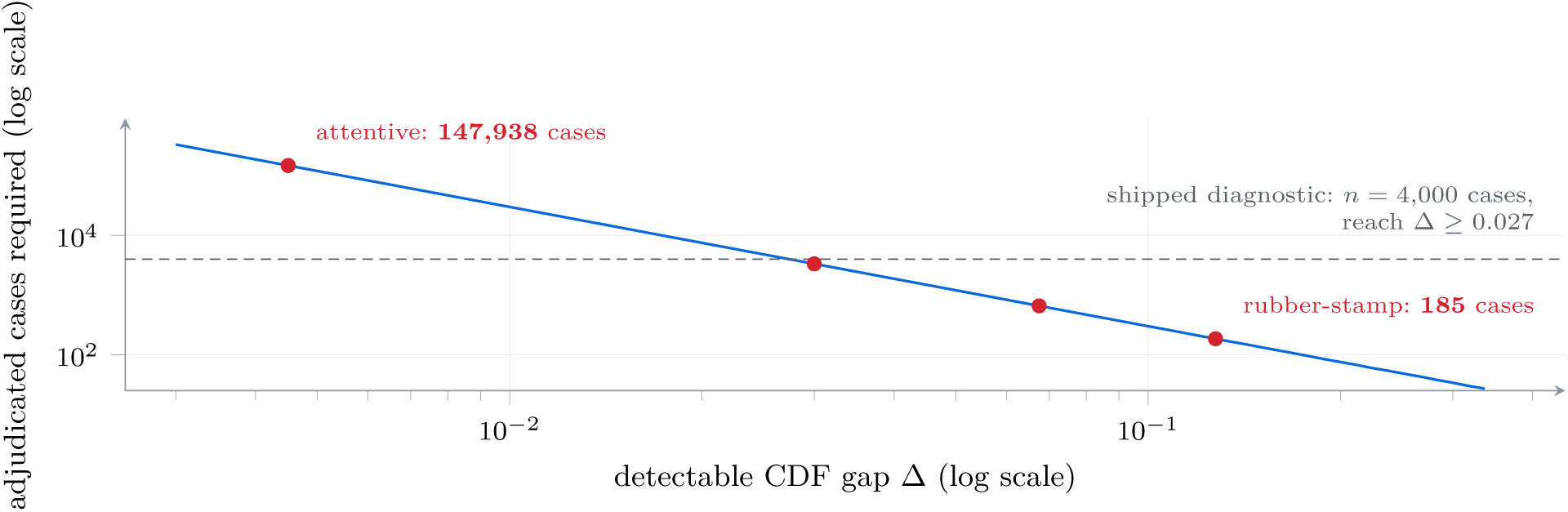
The diagnostic is affordable only where the gap is already large. The horizontal axis is the gap Δ the test must be able to see and the vertical axis is the number of adjudicated cases that takes, both logarithmic, so Eq. (3) is the solid line of slope *−*2. The four marks are the profiles of Table 1, and the dashed line is the sample size the diagnostic we ship is given: everything below it is affordable.

### Why we still ship the separate arm

The separate arm is not the better design. The published corrections we gate on are stated as conditions on two score distributions (Einbinder et al., 2024; Sesia et al., 2025), and `adjust_alpha` exists to refuse them when the condition is unmet. The claim in Section 6 on page 9 that no certificate rests on a realism penalty is enforced by a test that reads the library’s own source for callers of that function and fails if it finds one, and the only caller anywhere in the repository is the script that produced the numbers in this paper. A paired count measures the contamination itself and never tests that condition. That makes the correction unnecessary, and it licenses nothing, because a deployment holding adjudicated labels for its own cases can calibrate on those labels and skip the whole question. `zero_error_ floor` in `src/oversight/calibration/ltt.py` says a flawless run needs 14 such cases before it could certify at *α* = 20% with *δ* = 0.05 at all, so the floor is not the obstacle. Clinician hours are. A library that shipped only the paired route would be a better library and a smaller claim, and we say which of the two we built.

### What the concession does not touch

It does not touch Section 4. That result is about the planted probe, and an adjudicated chart is not a planted probe: it is the real distribution, read by a second person. Section 5 already calls buying adjudicated labels the defensible move, and this appendix says the defensible move is cheaper than we priced it, so it strengthens the recommendation while weakening the number beside it. Pairing also leaves the adjudicator with the same problem. The second reader has a miss rate too, unmeasured by the same argument, so the unknown moves from the reviewer to the adjudicator and does not go away. Section N is the section that refuses to treat the adjudicated arm as truth.

## I. Adjacent lines of work

Two lines of work sit next to ours without meeting it. The first widens what a certificate covers. Nguyen et al. trade off multiple risks under one budget (Nguyen et al., 2024), Rahman et al. stratify risk control by role for tool-calling agents (Rahman et al., 2026), and Gurram et al. price selective risk control per field of a structured output (Gurram, 2026). Each takes the calibration label as given, so each inherits whatever the reviewer supplied, and the wider the coverage the more labels the reviewer had to supply. The second line scrutinises the surrounding evaluation. Traub et al. audit how selective classification is benchmarked (Traub et al., 2024), Hu et al. examine conformity scores themselves (Hu & Su, 2026), Han et al. test validity by a martingale argument (Han & Qu, 2026), and Parris et al. set out an inspection scheme for model-written code, which is the software analogue of the review step we study (Parris, 2026). These change what a certificate is checked against and leave where its labels came from alone.

A clinical reporting guideline gets nearer than either line and still stops short. TRIPOD+AI asks an author whose outcome assessment “requires subjective interpretation” to describe the qualifications of the outcome assessors, and to report “any actions to blind assessment of the outcome to be predicted” (Collins et al., 2024). A clinician judging whether a generated summary was safe is that case exactly. Naming who they were satisfies the item, and a certificate computed from their labels can still be anti-conservative by an amount the item does not bound. Blinding an assessor to the model’s output controls something different from the effect at issue here, and we do not expect a subtle omission to be easier to see for being read blind.

One recent study shows what the borrowing looks like when nobody objects to it, and we name it here because the practice is otherwise easy to describe as a straw man. Brown et al. study whether suppressing a model’s poor predictions improves what a clinician and a model achieve together. They had no observed clinician responses to the two prediction tasks, so in their own words they “simulated clinician acceptance of ML predictions based on previous empirical work on acceptance of clinical decision support alerts” (Brown et al., 2026). Acceptance of an interruptive alert about a medication order is a different act, on different material, under a different workload, from judging whether a mortality or readmission prediction should be believed. Their conclusions are stated with care and we take no issue with them. The point is narrower and it is the one this paper is about: the borrowed rate enters the results with no interval and no limitation paragraph on whether it transports, and we know of no accepted way to supply either.

Aviation baggage screening and laboratory proficiency testing established that seeded material is unrepresentative of native error (Riz à Porta et al., 2022; Parsons et al., 2001; Buser et al., 2025), and Section L sets out the seeded-defect measurements in cytology, radiology and pharmacy review one study at a time, in the sign each one reports. Some of those studies set the seeded material against a native rate. Others hold the seed set fixed and vary one thing about the review, or hold the review fixed and vary the seed set, and those are in the table too, because a seeded rate that moves when nothing about the reviewer changes is the same objection stated a different way.

## J. The cost of stratifying by harm

Section F concedes in one sentence that we price a scalar miss rate where Section 4 implies a per-class one. This section expands that concession, because the question it answers is the one a deployment asks first: not whether a certificate holds on average, but which recommended actions may be released without a clinician and which may not.

### We follow a published stratification

Conditioning a distribution-free guarantee on something narrower than the whole output stream is established work. Rahman et al. stratify conformal risk control by the role a tool call plays (Rahman et al., 2026), Gurram et al. do it per field of an extracted document and build a ladder of guarantee tiers over the result (Gurram, 2026), and Nguyen et al. trade several risks against a single budget (Nguyen et al., 2024). Gurram et al. also report the verdict a stratified certificate tends to reach, that the tier matching the deployment unit is the weakest one on the ladder, and we claim neither the construction nor that finding. What this section adds is what happens to the diagnostic of Section 5 when the strata are ordered by harm and the labels still come from the reviewer under study.

### The stratification a clinical deployment wants is the harm of the recommended action

A recommendation to repeat a laboratory measurement and a recommendation to image a chest are not one object under review even when a verifier scores them alike, because the second commits the patient to a dose of ionising radiation and the first commits them to a second blood draw. Our library carries an ordinal severity on the claim in src/oversight/attest/claims.py and one named constraint set in src/oversight/attest/profiles.py, and `certify_claims` requires every constraint to hold at one threshold while correcting across the grid it searches. That severity grades how bad a claim is when it is wrong, and it is supplied by the caller. It is not the harm of the action the claim recommends, and nothing in the library derives the second from the first, so the strata it partitions the release into are wrong claims above a severity. They are not recommendations above a harm. We claim none of that construction here, and this paper contributes no method.

### The cost concentrates in one stratum

Let the release be partitioned into *J* harm classes and write Δ*_j_* for the contamination gap in class *j*. A reviewer’s miss rate is a function of detectability, the premise of Section 4, and the detectability distribution of the defects a model produces inside one harm class is not the distribution it produces inside another. A dominance diagnostic run on the pooled stream therefore licenses nothing about any single class, and the honest cost is *n*(Δ*_j_*) rather than *n*(Δ). That much is arithmetic. The order the Δ*_j_* fall in is not.

### The stratum that most needs the certificate is the least affordable

The vigilance literature this paper already leans on reports that detection improves when a target is attended to and degrades when it is rare (Wolfe et al., 2005; Evans et al., 2013). The size of the second effect matters here, because it is the mechanism the main text asserts and does not quantify. Wolfe and colleagues had the same observers search for the same target at three prevalences and miss 7% of it at 50% prevalence, 16% at 10% and 30% at 1% (Wolfe et al., 2005). Nothing was seeded and nothing about the target changed. Rarity alone moved the miss rate by a factor of four. In these studies the observer moves the threshold and keeps the same ability to tell the two classes apart, since false alarms fall as prevalence falls (Evans et al., 2013), and a threshold is exactly what a calibration set records. We expect a reviewer handed a recommendation that will irradiate a patient to read it more carefully than one handed a repeat electrolyte panel, because that is the behaviour a safety programme is built to produce. We have no measurement of it. Its consequence for measurement is unwelcome. Careful review means a smaller contamination gap, *n* grows as Δ*^−^*^2^ by Eq. (3), and where the high-harm class is also the rarer one it supplies adjudicated cases at the lowest rate while requiring the most of them. Both terms move the same way, and neither is a modelling choice a deployment could reverse.

### What that says about autonomy

Section 5 concludes that the affordable diagnostic exists only where the answer was never in doubt. Stratified by harm, the same conclusion applies to whole harm classes: the harm classes in which a reviewer’s residual miss rate can be measured on a realistic adjudication budget are the low-harm, high-volume ones. In those classes the case for releasing without a clinician never rested on a certificate in the first place. That is not an argument against granting autonomy over low-harm work, and the arithmetic here agrees that is where it belongs. It is an argument against the inverse move, of reading a certificate fitted on pooled review as licence over a high-harm class inside that pool, because a pooled diagnostic has its power where the pooled volume is and the volume is not in that class.

### Direction of this limit

Every step above makes the result of this paper harder to escape. That is why it sits in this section and Section 8 does not carry it. A deployment that hoped to get out from under Section 4 by conditioning on harm finds the conditioning legitimate, published, and expressible in our own library on a caller-supplied severity, with no implementation over the harm of an action, and finds that it multiplies the adjudication bill while concentrating the shortfall in the class it most wanted certified. We know of no measurement of Δ*_j_* across clinical harm classes for a language model under human review, and Section L does not supply one, so the ordering argued here rests on the vigilance mechanism and on no measurement of it.

## K. Exogeneity in four published results

Section 7 states that every rate-dependent correction for corrupted feedback needs the corruption event to be independent of the case that produced it, unconditionally or given observed covariates. We locate the requirement here, because it is easy to mistake for a symmetry requirement and it is not one. It enters in four separate places. Wang, Zecchin and Simeone reach their stochastic level by drawing the flip as an independent Bernoulli variable, so the flip rate is a constant of the channel and does not depend on the case (Wang et al., 2026). Xi et al. substitute an unconditional flip probability for a conditional one inside the proof of their noise-robustness lemma. That substitution is the step that lets a label-noise rate be carried through to a coverage statement (Xi et al., 2025). Wang et al. pull an observation indicator out of an expectation under intermittent feedback, and that is valid when whether a case is observed does not depend on what the case was (Wang et al., 2025). Feldman et al. assume a conditional independence between the corruption mask and the data when they re-weight (Feldman et al., 2025).

A reviewer miss violates all four for a single reason: the miss happens *because* the defect read plausibly, so the event and the case are the same event described twice. Two apparent escapes do not work. Conditioning on observed covariates does not restore independence, because the covariate that drives the miss is the defect’s detectability. Nothing in a deployment records it, since a deployment could record it only by knowing which defects were missed. And modelling the unreviewed case as an erasure moves the problem without solving it, since the erasure probability is then the miss rate itself, and the miss rate is the quantity under estimation (Wang et al., 2025).

Figure 10 puts the two channels side by side, and because both panels hold the same four elements at the same four places, anything that differs between them is a claim, not a drafting choice. One thing differs, an arrow. In the upper panel nothing arrives at the die, and the space to its left carries the words *nothing arrives here from the case* and is not left blank, since a blank region reads as a gap in the drawing instead of as the content of the assumption. That absence is what makes the flip rate, written *ε*, a constant of the channel and no function of the case. A constant is estimable from any sample of it, and that is the step all four results above take. In the lower panel the arrow is present, drawn red and routed out of the top of the case, so its tail is the detectability *d* of Eq. (2) and its head is the miss rate *m*(*d*). The word **unrecorded** sits beside *m*(*d*) because that is why the repair named above is unavailable, and hard is not the word for it.

**Figure 10.**
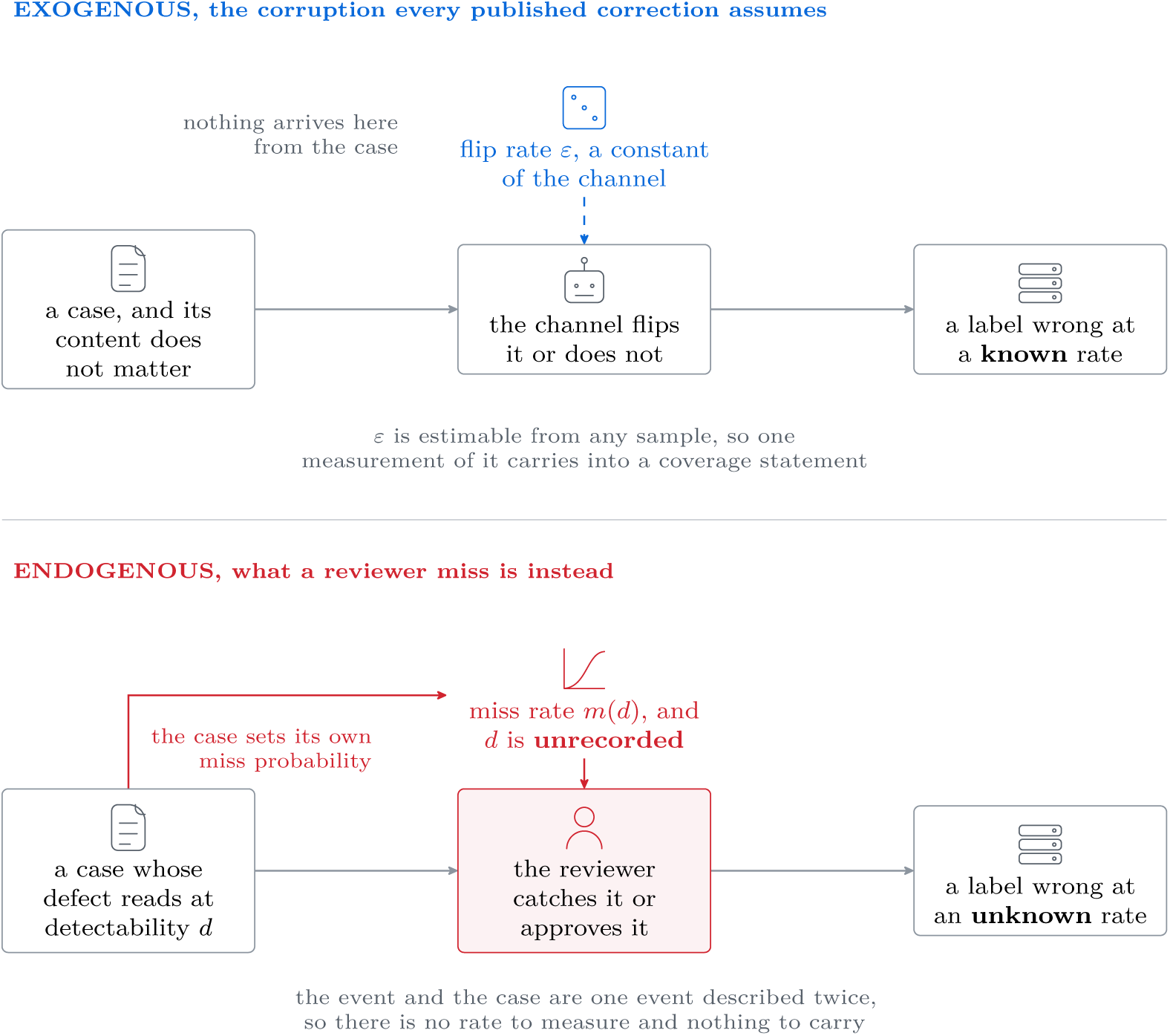
The difference between an assumption and a violation of it is one arrow. Both panels carry the same four elements in the same places, so the comparison is positional. In the upper panel the flip rate *ε* is a constant of the channel and nothing reaches it from the case. That is why one measurement of *ε* can be carried into a coverage statement. In the lower panel the red arrow out of the case is present: the defect’s detectability *d* decides the miss rate *m*(*d*), so the corruption event and the case are one event described twice. The figure is drawn by hand and carries no data. It states the premise behind Eq. (2) and the reason the four results collected here do not cover ours.

Figure 11 then puts the four steps beside each other with the one reason under them, because four proofs audited in four sentences is a paragraph a reader finishes without holding the shape of it.

**Figure 11.**
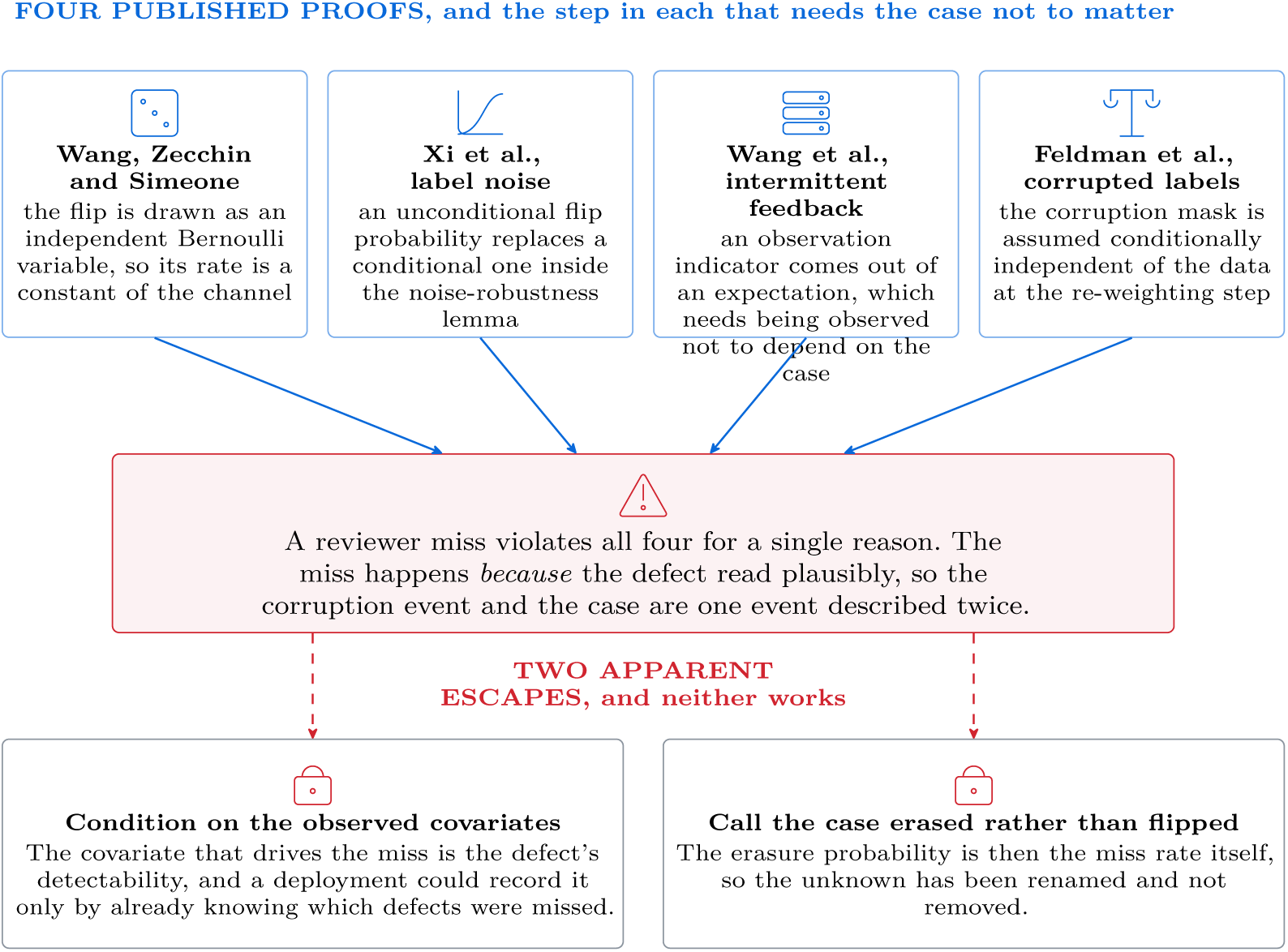
Four proofs, four different steps, one shared premise. The top row is the four places the paragraph above locates, cited left to right as (Wang et al., 2026; Xi et al., 2025; Wang et al., 2025; Feldman et al., 2025). All four boxes are one colour and the arrows converge on one point, because the claim is not that there are four inputs but that they reduce to the box beneath them. The lower row is the two escapes a reader reaches for first. Nothing here is measured, and that is why the figure carries no axis.

Read that figure as a funnel, top row to hinge to bottom row, because the vertical order is the order this section argues in. The icon on each box of the top row names the kind of step it stands for: an independent draw, a claim about a distribution, a rule for which cases reach the calibration set, and a re-weighting. Four steps that look nothing like each other reduce to the single sentence beneath them, and that sentence is one sentence because there is one reason. The two boxes in the bottom row carry the withheld glyph and no cross, since neither repair is wrong in general and both are unavailable here.

## L. Seeded and native detectability, study by study

Section 4 argues that a probe-derived miss rate is a measurement of the probes, and shows the spread inside one instrument in Fig. 12. Table 6 is the whole set the argument rests on, one row per study, so that a reader can check the direction column against the sources themselves. Every number in it is quoted from the study cited on its row and none of it is our measurement. The final column is the direction relative to the assumption a realism penalty encodes, that probes are easier than the real thing, and its values fall into four families. Where a study compared a probe against the real thing and the probe was easier, it reads *as assumed*. Where the probe was harder or indistinguishable it reads *against*, of which *equivalence assumed, not found* is the special case: the two were matched by construction on the properties that decide detection, so agreement there was built in before any measurement. Where what moves the measured rate is something other than the reviewer, the column names that something, because such a row reports no direction at all. And where a study publishes no rate against a real defect at all, the column names the arm it did compare against, which is where both mature programmes end up: *ordering only* for one of them and *probe against probe* for the other. Figure 13 sorts the studies into those families, and the column does not add up to a signed law, which is the point of the table.

**Table 6.**
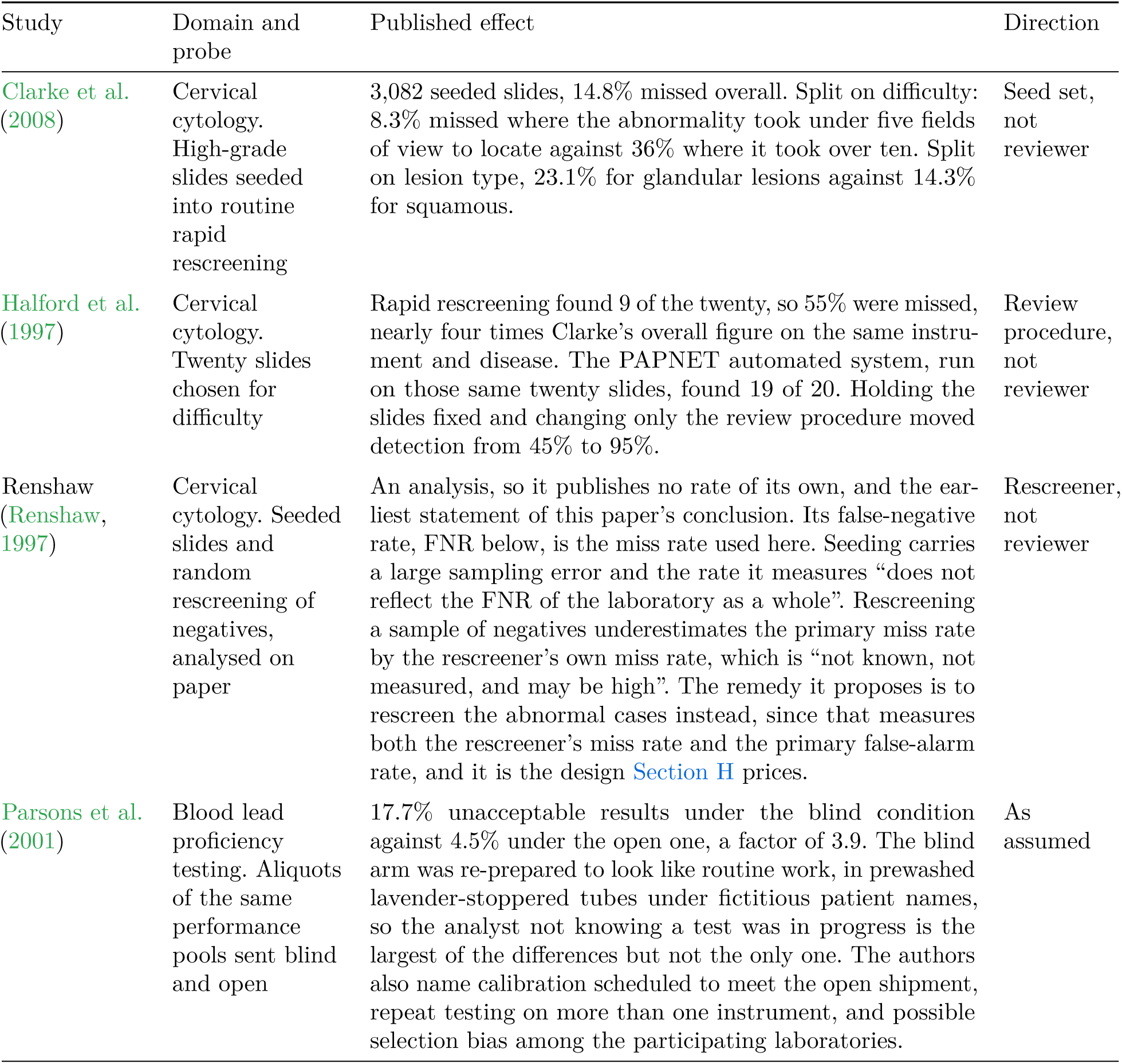
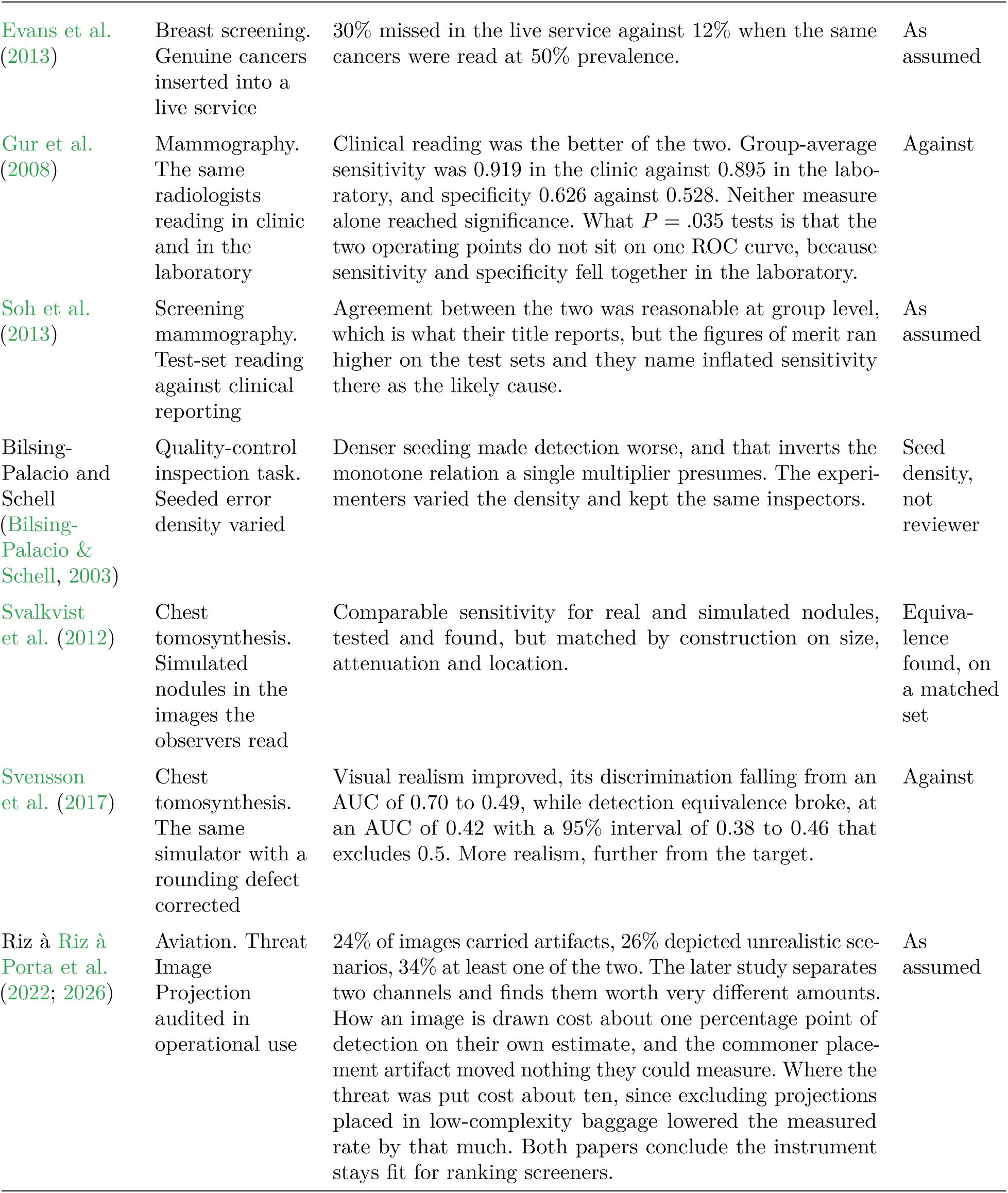
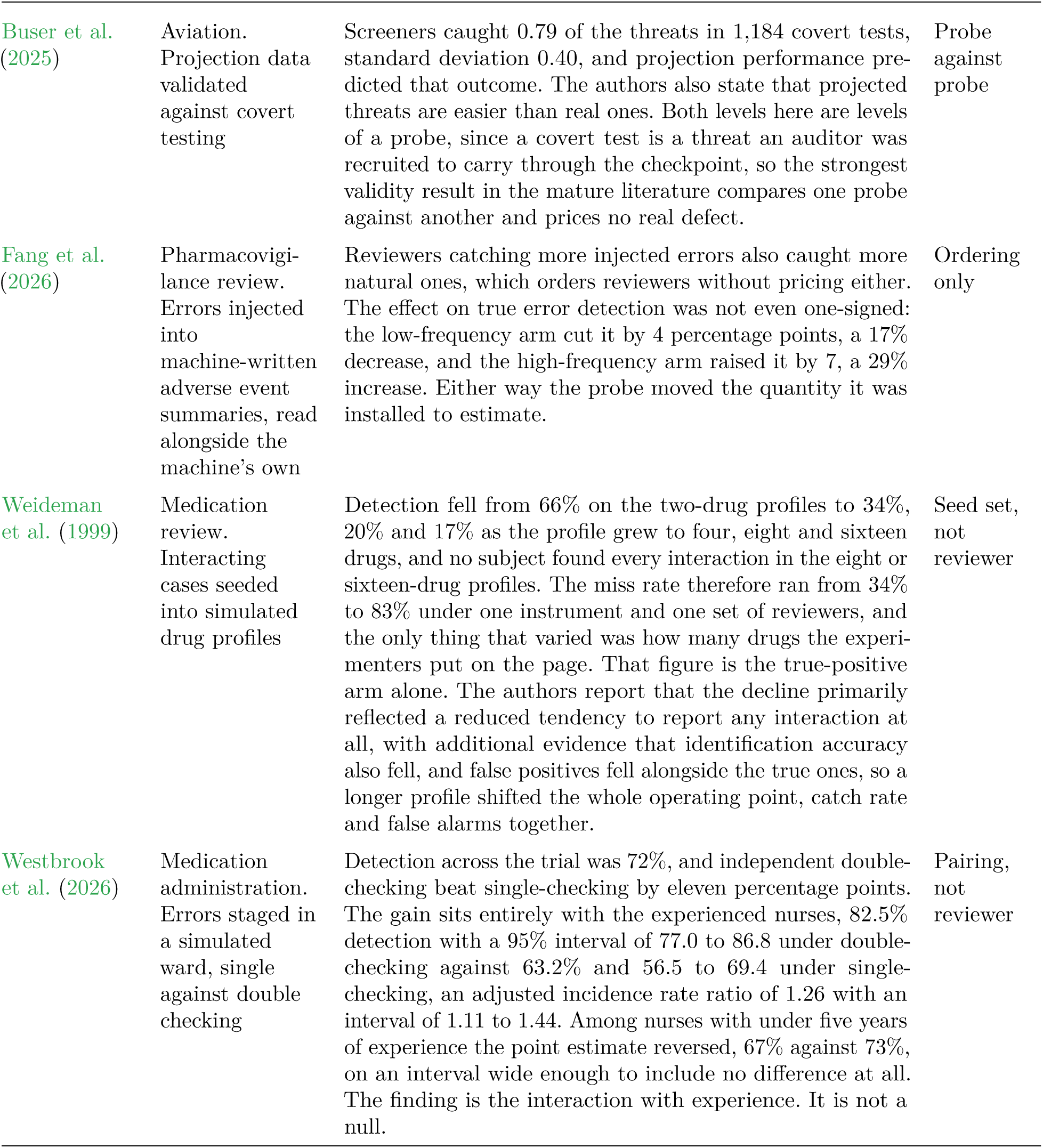
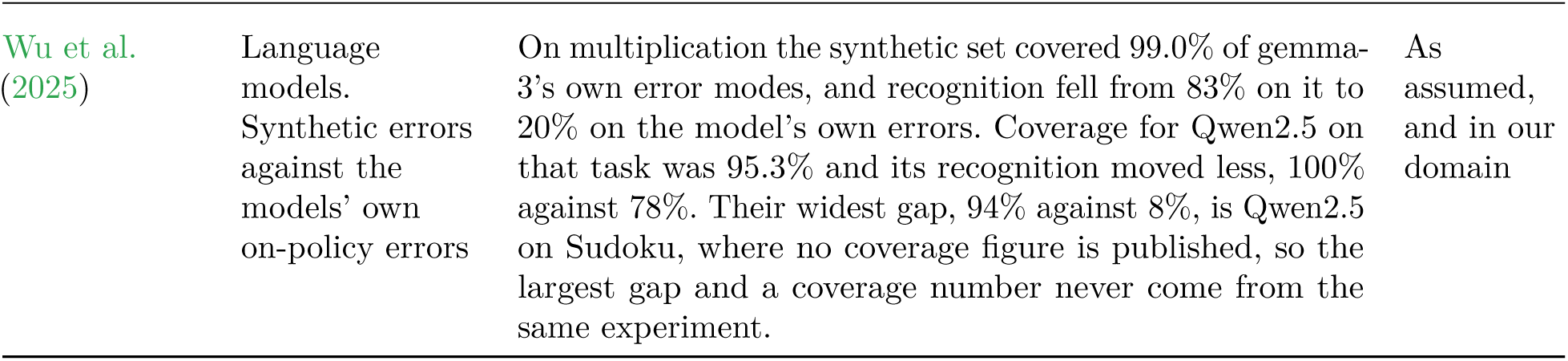
The seeded-against-native record supplies no signed law. One row per study, with the seeded material, the published effect and its direction relative to the assumption that probes are easier than real defects. Rows disagree in sign, so no average of them is reported anywhere in this paper.

**Figure 12.**
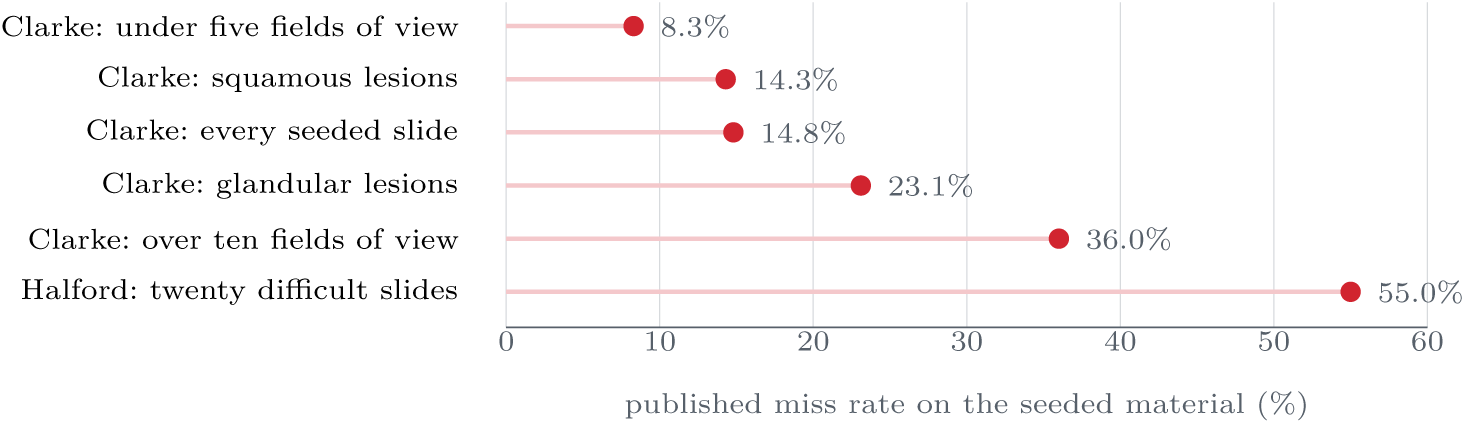
Six published miss rates from one instrument and one disease, and no difference in reviewer attention behind any of them. All six are cervical cytology rapid rescreening: five are Clarke et al.’s 3,082 seeded high-grade slides split on their own difficulty and lesion-type covariates, and the sixth is Halford et al.’s twenty slides chosen for difficulty (Clarke et al., 2008; Halford et al., 1997). The first five differ in which slides were seeded. Halford et al. also show the other way a rate on this axis can move, since running an automated system over their same twenty slides took detection from 45% to 95% with the slides held fixed. Neither route runs through the reviewer, and a realism penalty is one number applied to whichever of these a deployment happened to run.

**Figure 13.**
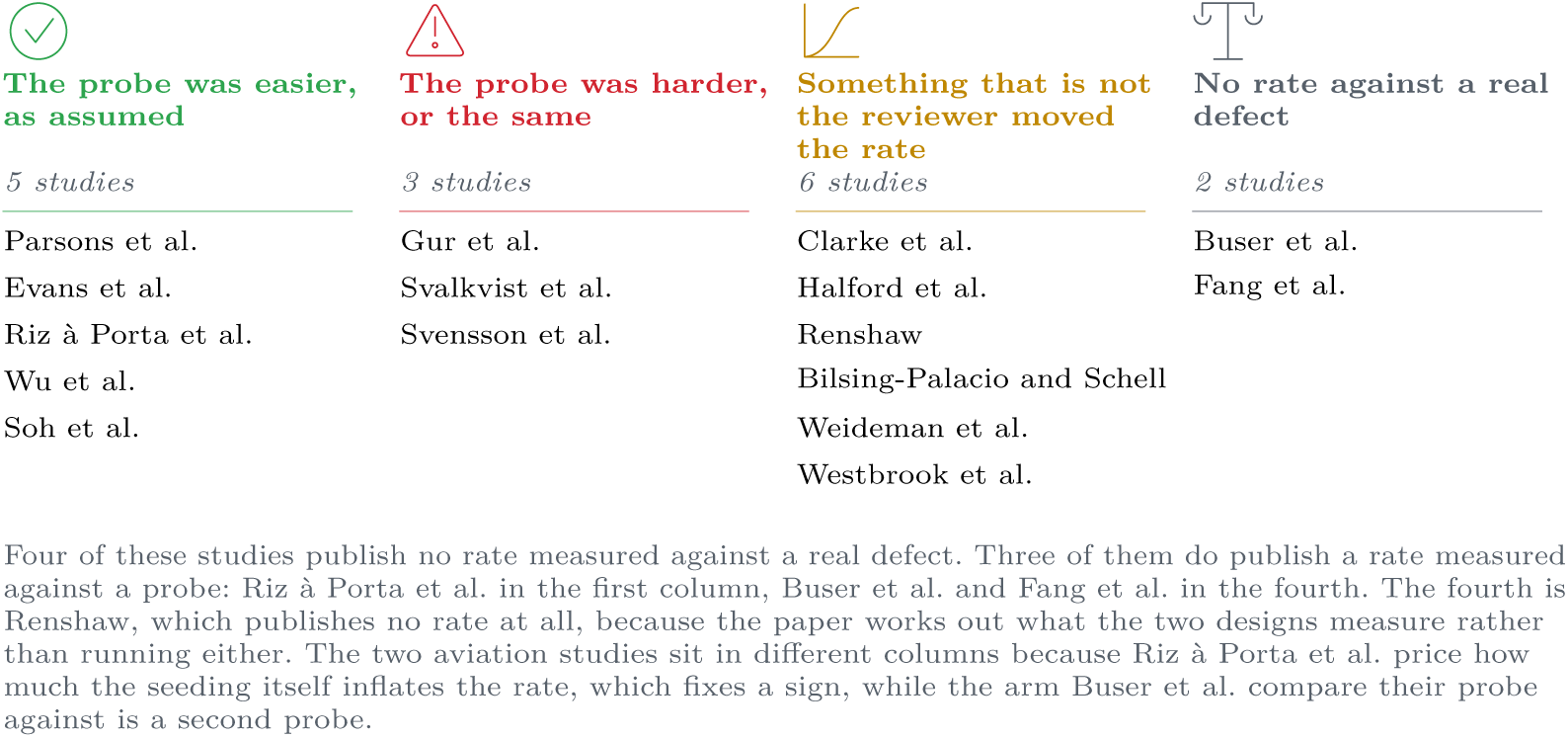
The Direction column of. Table 6**, sorted.** Every study in the table placed by what its own result says about the assumption a realism penalty encodes. All four families are populated, the family that contradicts the assumption is the smaller of the two signed ones, and no family is larger than the one whose studies report no direction at all, because what moved the measured rate in them was the seeding, the interval or the case mix, and none of it was the reviewer. No rate is plotted and none is averaged: the sixteen studies report unacceptable proficiency results, cancers missed in service, detection AUC, which is one number for how well a reader separates the diseased cases from the healthy ones, and error modes recognised, which are four different quantities.

The spread Fig. 12 draws is inside a single instrument and a single disease, where the confounds between studies are smallest. Across methods the sign is not stable either. Of the five studies cited on this line, three find the seeded material easier to work against and one finds it harder, so the tendency is the assumed one without being a law, and a fifth finds that seeding errors more densely made detection worse, and that inverts the monotone relation a single multiplier presumes (Parsons et al., 2001; Evans et al., 2013; Gur et al., 2008; Soh et al., 2013; Bilsing-Palacio & Schell, 2003). Three further rows set a simulated or synthetic probe against the real thing, and the paragraph after Fig. 13 reads them out one by one and keeps them out of that count. One of the three found the probe harder than the target, one had its agreement built in by construction, and the third is the only study in the table run on language models.

The column that agrees with the assumption is no more reassuring than the column that contradicts it. A study finding its probe easier than the real thing fixes a sign, and Section 4 argues that a sign is not a factor, so none of the four outcomes licenses carrying a multiplier from one deployment to the next.

Two of the comparisons need reading before the table, because they are the two a reader is most likely to expect to break the argument. The first is the attempt to fix the problem by building a better probe. A lung-nodule simulator was found to round its attenuation values, the defect was corrected, and the corrected nodules were then rated more realistic by observers than the originals. Detection equivalence went the other way: against the corrected nodules the detection AUC was 0.42, an interval that excludes chance in the direction opposite to the one realism was supposed to buy (Svalkvist et al., 2012; Svensson et al., 2017). Making the probe look more like the target moved it further from the target on the axis a certificate would have to price. The second is the language-model case, which removes the need to argue by analogy from imaging at all. Wu et al. sampled a model’s own errors on a task, built a synthetic error set of the same kinds, and then measured two separate things: how much of the model’s own distribution of error modes the synthetic set covered, and how often the model recognised an error of each kind. On multiplication the synthetic set covered 99.0% of gemma-3’s own error modes, and recognition was 83% on the synthetic errors against 20% on the errors the model made itself. Qwen2.5 on the same task gives coverage of 95.3% with recognition of 100% against 78%, so the gap is there in one model and small in the other. The widest gap they report is Qwen2.5 on Sudoku, 94% against 8%, and for Sudoku they publish no coverage figure, so that gap and a coverage number cannot be read off the same experiment (Wu et al., 2025). Near-complete coverage of the error modes with a four-fold gap in detection, on the one task where both quantities are published for the same model, is the identifiability claim of Section 4 restated as an experiment.

One result runs the other way in the same setting. Stenzl et al. took facts extracted from oncology literature summaries, modified them at three graded levels named subtle, moderate and contradictory, and reported that detection scores fell as the modification grew more severe (Stenzl et al., 2026). That is the monotone relation between severity and detectability which a single realism penalty presumes. Here it is a finding, and we do not discount it. Two things keep it from supplying the penalty. The detectors were panels of language models, and where the same study measured people it found them agreeing with each other at a kappa of 0.43, so the monotone curve is a property of the automated panel alone. And the study plants errors at graded severities without ever comparing them against errors the pipeline made on its own, so it fixes a shape and never a level. It is not a row in Table 6 for the first of those two reasons only. Six rows of that table have no native arm either, so a missing arm is not what keeps a study out. The detector keeps this one out. Every row of the table records what a person missed, and this study records what a panel of models missed.

The two regulated industries that have run this instrument for decades belong in the last of those four families, and aviation is the clearer of the two about why. It states outright that its projected threats are easier than real ones, so a screener scoring ninety per cent against projections is not to be expected to find ninety per cent of actual threats, and it puts a figure on how much the seeder’s hand is worth, because excluding projections placed in low-complexity baggage lowered the measured rate by about ten percentage points (Riz à Porta et al., 2022; 2026; Buser et al., 2025). The literature does not supply a rate against a real defect, and no published level is missing either. Its strongest validity result compares projected performance against covert testing, and the covert level is printed, an average hit rate of 0.79 with a standard deviation of 0.40 across 1,184 tests, which projection performance predicted at *p* < .001 in a model clustered within screener (Buser et al., 2025). A covert test is a threat an inspector recruited an auditor to carry through the checkpoint, so 0.79 is the level of a second probe and not the level of a threat somebody brought meaning to use it. Both aviation papers add that the instrument remains fit for the purpose it was built for, which is ranking and training screeners, and on their own estimate the drawing of the image costs about one percentage point of detection against the ten that placement costs. Ranking is the use it survives, and ranking is what Section 4 shows a certificate cannot be priced from. The closest clinical analogue withholds more, ordering pharmacist reviewers by their catch rate on errors injected into machine-written summaries of adverse event reports without pricing what the same reviewers miss on the errors the machine made by itself (Fang et al., 2026). A programme that has run the instrument at scale for longer than this field has existed, and that still cannot price what a screener misses on a threat nobody planted, is evidence about the instrument and not about the diligence of the programme.

### How the federal mandate truncates its own probe set

Section 4 states that 42 CFR 493.945 requires unanimous agreement among at least three board-certified anatomic pathologists, and that the requirement removes the hard material. The requirement sits in paragraph (b)(1). That paragraph also demands tissue biopsy confirmation for the premalignant and malignant slides. The three pathologists must agree on one of the four coarse response categories tabulated at (b)(3)(ii)(A). Those four run from unsatisfactory through normal, low grade, and high grade with carcinoma. Paragraph (a)(1) then requires, in its own words, that “test sets should be comparable so that equitable testing is achieved within and between proficiency testing providers”. Two features of that structure matter here. Agreement is demanded at a resolution four categories wide, so a slide two pathologists would place either side of one of those three boundaries cannot enter the set at all, and the paragraph binds a testing program that HHS has approved, not a laboratory, so the truncation is upstream of any laboratory’s discretion and is written into the fairness requirement instead. The regulation does not say what removes the hard material in practice, and the distinction matters because a referee will look the paragraph up. Section 493.945 prescribes no validation step of any kind. The procedure that does the removing is a recommendation from the field, and Birdsong et al. are the ones making it, writing that field validation of the slides before their use in graded test sets is “mandatory for the test to be considered acceptable” to the consortium that reviews such sets. That validation measures every candidate slide under live testing conditions and discards the ones that behave inconsistently between readers, and the rejection rates they publish were measured in the College of American Pathologists interlaboratory comparison program, and no federal requirement demanded them. It rejected 31.8% of conventional low-grade slides against 15.8% of their liquid-based equivalents. On the high-grade slides the ordering reverses, 9% conventional against 17.6% liquid-based, so the loss is not even one-signed across preparation methods. Their plainest figure is that fewer than half the slides admitted to that program under the three-pathologist rule went on to pass field validation at all (Birdsong et al., 2006). Those rejection rates carry two consequences for the argument in the main text. The rejection is the point of the procedure and not a defect in it, because a slide that two competent readers grade differently cannot be scored fairly against either of them. And a slide that competent readers grade differently is, by that same fact, one of the harder ones, so the fraction discarded is a lower bound on how much of the difficult mass a compliant probe set has to leave out. A programme that set out to make its probes more realistic would have to fail this validation to succeed.

### What range of reviewers the four profiles cover

The miss rates in Table 1 are a grid. They are not estimates. No published study set out to measure how often a clinician misses a defect in a model’s answer, so no rate in that grid is fitted to a study. Section M recomputes two such rates ourselves from marks other groups released. That is a different thing from a published estimate, and it is read there with that in mind. One demand can still be made of a grid. It has to reach the range that published human measurements occupy, because a case count computed at a miss rate nobody has observed prices nobody. Table 7 collects the 9 rates from the studies above that report the share of present defects a qualified reader passed, and puts beside each one the cases per arm Eq. (3) would ask for at that rate. The published rates run from 8.3% to 83%. The grid runs from 3% to 85%, so it brackets them on both sides, and its careful end is more careful than any reviewer in this literature.

**Table 7.**
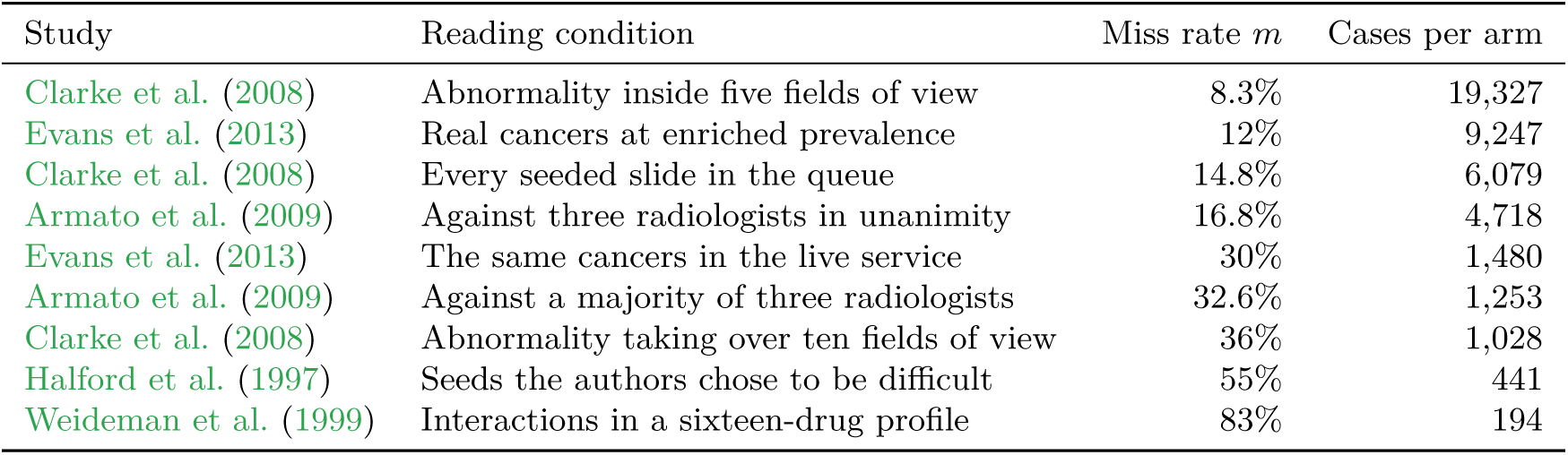
Published human miss rates, and what the diagnostic would cost at each. One row per published reading condition, ordered by rate. The middle column is the share of defects known to be present that a qualified reader passed. The right column is Eq. (3) evaluated at that rate, in cases for one arm of two. No row estimates the miss rate of a clinician reading a model, and the table is here to fix what range the four profiles cover.

Read the right-hand column as a price. It is not an estimate. Every rate in the table was measured on a slide, a mammogram, a chest scan or a drug profile, and none of them transports to a clinician reading a model. Transporting one would repeat the error Section 4 objects to, which is carrying a rate measured in one setting into a second setting that shares none of its parameters. The column carries the shape of the cost. At the most careless rate anybody has published the diagnostic asks for 194 cases in each arm, and at the most careful it asks for 19,327. Both arms need independent adjudication, so the first figure is a study somebody could run and the second is not.

Two rows have to carry their sample size with them. Armato et al. measured each thoracic radiologist against a reference three other radiologists defined, and report a mean sensitivity of 83.2% when the three agreed unanimously and 67.4% when a majority sufficed, over 25 scans from one site (Armato et al., 2009). Both standard deviations exceed 25 percentage points, so those two rows are the closest in kind to the quantity a certificate needs and the least settled in value. Their own reading of the spread is the sentence this paper would have written, that experienced thoracic radiologists “may not perform well when measured against the ‘truth’ established by other experienced thoracic radiologists”.

Section M leaves the grid behind and measures two such rates directly, and Section N turns the same question on the other arm, and asks what fixes the rate of the adjudication this paper prices as the honest route.

## M. Reviewer miss rates measured from published grades

Every other reviewer number in this paper is a point on a grid. These two we computed from marks clinicians gave, and this appendix says how, so that a reader can judge how far they travel. The first is measured on grades physicians gave to written model answers, and its reference standard is the other physicians. The second is measured on electrocardiogram readings, and its reference standard was settled by different people before the readers we score saw the tracings.

### The corpus

The HealthBench meta-evaluation releases the grades 186 physicians gave to answers that language models wrote to health questions (Arora et al., 2025). Each answer arrives with named criteria, and a physician marks each criterion met or not met on that answer. Every criterion on every answer carries marks from at least two physicians, and many carry more. The release we read holds 60,896 marks over 14,592 model answers. Its digest begins 6a343ed3fcb6, and `examples/physician_miss_rate.py` carries the full digest, the byte count, the row count and the address to fetch it from. The corpus is somebody else’s. Its licence is permissive, but it also carries a canary string and a request that its examples not be reposted, so our loader prints no question, no answer and no criterion. It prints counts.

### The estimator

Take one criterion on one answer and call that a case. A physician who marks the criterion not met has flagged a defect in the answer. Now hold one physician out and let the others who read the same case vote. If a majority of them flagged a defect, the case had one, and if the held-out physician did not flag it, that is one miss by that physician. If a majority cleared the case and the held-out physician flagged it, that is one over-call. The held-out physician’s own mark never enters the vote that judges them. That is the reason for holding them out at all. The function `peer_miss_rate` does this and returns both counts with exact intervals.

Holding a reader out is not our own idea. Warby and colleagues did the same thing on sleep spindle scoring, where 24 technologists read each epoch a mean of 5.3 times over and the reference standard was the group’s own consensus (Warby et al., 2014). Their reason was ours. A reader who helped settle a standard is being graded in part on their own answer. They also measured what the correction is worth. The average expert scored an F1 of 0.75 against the consensus the experts had built together and 0.67 against a consensus that same expert had not contributed to. Eight points of F1 was what a reader gained from having helped write the answer key, and it is why the second arm below leaves out the two readers who helped settle its standard.

Two kinds of case cannot be scored, and the function reports each of them instead of dropping them quietly. A case that fewer than three physicians read leaves the held-out physician with a single peer, and a single peer cannot outvote anybody, so every disagreement would become a miss for one of the pair and an over-call for the other. That is a disagreement rate wearing a miss rate’s name, and the function refuses to compute on it. 55,546 marks fall out for this reason. A case whose remaining peers split evenly has no majority at all, and breaking that tie would need the held-out physician’s own mark. 1,185 marks fall out for this reason. 4,165 scored marks survive.

With exactly three readers the window this leaves is narrow. A mark can be scored only when the other two readers agree with each other. So when two readers go one way and one the other, only the lone dissenter is scored, and each of the two in the majority faces a tied vote among their own peers.

### What it comes to

Physicians passed 257 of the 977 defects their peers agreed were present. That is a miss rate of 26.3%. The two-sided Clopper-Pearson interval 23.6% to 29.2% treats the grades as independent. They are not: they cover 472 criteria on 352 answers from 134 conversations, and resampling whole conversations gives 21.6% to 31.7%, the interval we report. Every scored defect sits in HealthBench’s context-seeking and hedging themes, the only answers graded by three or more physicians. Physicians passed 22 of 76 accuracy grades (28.9%), 43 of 210 helpful-and-safe grades (20.5%) and 27.8% of the 691 grades on seeking context or stating uncertainty. They flagged 12.1% (11.0% to 13.3%) of the cases their peers had cleared. Among the 17 physicians who were shown at least twenty peer-confirmed defects the rate ran from 8.0% to 54.5%, with a median of 26.3%. That raw range overstates the real spread, because with about twenty defects each, physicians sharing one rate would still differ by 28.8% by chance. A DerSimonian-Laird random-effects fit on the proportion scale gives Cochran’s Q of 43.5 on 16 degrees of freedom and an I squared of 63%, a between-physician standard deviation of 10.3% in the true miss rate, and a middle band of true rates of 7.5% to 47.9%. That fit assumes the physicians graded comparable answers, and they did not, so it cannot separate the physician from the case mix. An analysis of all disagreement in this corpus finds that physician identity explains little of it (Borgohain & Mariathas, 2026). Put that rate through Eq. (3) and the diagnostic costs 1,925 cases in each arm, or 1,564 to 2,398 at the two ends of the Clopper-Pearson interval.

### Why the estimator does not flatter itself

The scored set is chosen by what the peers agreed on, and a case everybody missed drops out of it. An estimator built that way owes the reader a demonstration that it does not read low by construction. Plant a known miss rate, let three readers miss independently at that rate, and the estimator returns the planted rate, because the two ways a case can be scored carry the same factor and it cancels. `tests/test_raters.py` checks that recovery at several planted rates. Correlated misses are where it reads low, and the same file plants those and asserts that the estimate lands under the rate the readers were given.

### Three reasons to read the number as a floor

First, the physician was handed the criterion to check. Judging a criterion somebody else wrote is easier than reading an answer cold and deciding whether anything in it is unsafe to release, and the second is what a certificate’s reviewer does. Second, the reference is the peers, so a defect all three readers passed is recorded as a miss by nobody, since nobody was left to record it. Both of those push the measured rate below the rate a release reviewer would have. Third, none of these physicians was in a clinic, holding the case, or answerable for the answer, and that one could push either way. We cannot sign it. So the honest reading is that 1,925 is an upper bound on the case count. It is not an estimate of it.

### A second arm, where the standard is not the readers

The second of those three reasons is the one that can be tested, so we tested it on a corpus built the other way round. The CODE study released a set of 827 electrocardiogram tracings, one per patient, each marked for the presence or absence of 6 named abnormalities (Ribeiro et al., 2020). Two certified cardiologists read every tracing on their own. Where the two agreed, their common reading became the standard. Where they disagreed, a third senior specialist who was shown both readings decided. The same tracings were then read again by three other groups: two fourth-year cardiology residents, two third-year emergency residents and two fifth-year medical students, each person taking half the set. Every group’s sheet is published beside the standard. So the reference here was fixed before the readers we score ever saw the material, and a defect a whole group passes is still on the record as a defect.

### What the second arm comes to

Take one abnormality on one tracing as a case. That gives 4,962 cases. The standard records a defect in 158 of them and calls the other 4,804 clean. The 3 groups who had no hand in the standard passed 68 of the 474 recorded defects put in front of them, a rate of 14.3% with an interval of 11.3% to 17.8%, and flagged 0.5% of the clean cases. Put that rate through Eq. (3) and the diagnostic costs 6,470 cases in each arm. The function that computes it is `reference_miss_rate`. It takes the standard as an argument and does not build one from the readers. `examples/ecg_reader_miss.py` pins a digest for each of the six files. The standard’s digest begins 4a0ed7059333.

The rate this arm reports is close to the rate the physicians reached on written answers. That is worth saying because nothing forced it. Two different tasks, two different kinds of reference and two sets of clinicians who never met, and the two numbers land within a few points of each other.

### Grade does not order the rate

The cardiology residents passed 15.2% (10.0% to 21.8%) of the recorded defects and the emergency residents 16.5% (11.0% to 23.2%). The medical students passed 11.4% (6.9% to 17.4%), a figure below either resident group. Those intervals overlap, so we do not claim the students read better. We claim the reverse of a claim. Nothing here lets a deployment pick a miss rate off a reviewer’s grade, and 11.4% to 16.5% across three groups doing the same reading is the same lesson the spread across physicians taught above, reached on another task against another kind of standard.

### What the second arm gives up

Four things, and none of them is small. It is electrocardiogram reading, so the task is not the one a certificate’s reviewer does. Each group’s sheet is two people who split the tracings between them, so the rate is a group average and no per-person spread can be read off it. The two cardiologists’ own sheets are in the release, at 12.7% (7.9% to 18.9%) and 7.6% (4.0% to 12.9%), but they are the readers whose agreement built the standard, so those two rates measure the standard partly against itself and no number above is computed from them. Warby et al. measured what that costs, and it was eight points of F1 on their own task (Warby et al., 2014). Pooling all 5 sheets gives 100 of 790 defects passed, a rate of 12.7% (10.4% to 15.2%) with 0.4% of the clean cases flagged and 8,310 cases per arm. We report that here so the choice is visible, but 14.3% is the number to read. Last, the released standard and the source paper’s first table do not quite agree: the file carries 158 recorded defects and the table prints 157, a difference of 1 case in ST. We compute from the file, because the file is what the readers’ sheets are aligned to, and we report the difference.

### What the numbers do not settle

They are two rates on two tasks, one rubric and one set of model answers, and one reading task with one set of tracings. Neither identifies *m* for any deployment. Both depend on the readers, the task and the cases, and a deployment that borrows a single value has borrowed one measured on other readers and other cases, these two included. Section 4 stands unchanged. The argument there is that a seeded probe cannot supply *m*, and neither a rate measured on somebody else’s corpus from three independent readers per case nor a rate measured against somebody else’s adjudicated standard is a seeded probe.

## N. The adjudicated arm is a chosen distribution too

Section 5 treats independent adjudication as the honest alternative, and a clinician objects before the sentence is finished: adjudication is a design, and a design has a rate of its own. The objection is right, and we put it at full strength, because the numbers behind it were measured by radiology on radiologists.

### The same cancers, four review designs, four answers

Hofvind et al. took 231 interval breast cancers from one population screening programme and had six radiologists review them four different ways, varying whether the reviewer was told the case was an interval cancer and whether the prior screening mammograms were mixed in among normal ones. The share classified as missed came out at 19.9%, 23.4%, 35.9% and 33.8% across the four designs (Hofvind et al., 2005). Nothing about the cancers changed between those four numbers. A systematic review of 46 studies of the same task reports classifications ranging from 4% to 40% (Mullooly et al., 2021), and it does not attribute that range to design, so neither do we. Hofvind et al. is quoted here because it is the study that varied the design and held the material fixed.

### Context changes how a case reads

Egglin and Feinstein showed the same 24 angiograms to six radiologists on two occasions and changed only the proportion of abnormal cases in the set the 24 were embedded in. Sensitivity moved from 60% to 75% (*P* = .04), and the area under the receiver-operating-characteristic curve, which plots sensitivity against the false-alarm rate as the reader’s threshold is swept, moved from 0.82 to 0.88 (*P* = .02) (Egglin & Feinstein, 1996). That area is the AUC of Fig. 13, and we write ROC for that curve from here on. The laboratory effect Gur et al. report, quoted earlier in Section L, is the same finding reached by a different manipulation.

### Satisfaction of search

Berbaum et al. added a simulated nodule to images that already carried a native lesion, leaving the native abnormality physically unchanged, and accuracy on that native lesion fell. Two accuracy measures are quoted. One is the area under the reader’s ROC curve, which that literature writes *A_z_*. The other is *d^′^*, and it says how far apart the diseased and the healthy cases sit on the reader’s decision axis. Averaged over the individual readers’ curves the drop reached significance on both, *t* = 2.364 at *P* = .025 for *A_z_* and *t* = 2.648 at *P* = .017 for *d^′^*, where *t* is the paired statistic Berbaum et al. report. On the pooled curve the two measures split, *t* = 1.573 at *P* = .080 for *A_z_* and *t* = 1.934 at *P* = .047 for *d^′^* (Berbaum et al., 1990). All four are reported here, including the pooled result that clears 0.05 and the pooled result that does not, because keeping only the convenient ones is the move this paper is against.

### The same radiologist, later in the day

Krupinski et al. had forty attending radiologists and residents read for fractures at the beginning and at the end of a working day. The detection area under the curve fell from 0.885 to 0.852 (*P < .*05), alongside measured increases in symptoms of visual fatigue (Krupinski et al., 2010). The reader did not change. The hour did.

### The programme that reviews readers does not score the reviewer

RADPEER is a peer review programme running at scale in United States radiology, and its design repays close reading because it is what a real oversight arrangement looks like at scale. A radiologist who encounters a prior study in the course of normal work scores the agreement of the original interpretation on a published scale (Jackson et al., 2009). The multifacility study reports 20,286 cases from fourteen facilities, 2.9% scored at 2 or above and 0.8% of the total validated by a committee (Borgstede et al., 2004), and a workstation-integrated programme reports 29,044 reviews by sixty-six radiologists, 0.69% scored 2 to 4 and 0.42% scored 3 to 4, with 75.2% of the disagreements being a missed diagnosis (O’Keeffe et al., 2016). Every one of those rates is a property of the reader under review. None of them is a miss rate for the reviewer, and the reason is structural. It is no oversight in the programme’s design: the reviewer’s own reading is the standard the other reading is scored against, so nothing is left over to score the reviewer with. Bender et al. examined that standard directly, showing twenty-five known-discrepant cases to twenty-one attending radiologists and finding a kappa of 0.11 on the score assigned, with the readers disagreeing about whether a discrepancy had occurred at all in twenty of the twenty-five cases (Bender et al., 2012).

### Model-as-judge is not a fifth route

None of this is repaired by replacing the clinician with a language model as the adjudicator. The strongest published case for that method reports a judge agreeing with human preference at about the level at which two humans agree with each other (Zheng et al., 2023). That is an agreement rate, not a miss rate. Judges have also been shown to favour their own generations by an amount that tracks their ability to recognise them (Panickssery et al., 2024). Substituting a judge for the reviewer replaces one unknown *m* with another that is correlated with the system under test. That is the one direction Section K shows a correction of this family cannot absorb.

### Effect on the argument

It does not weaken Section 5. The adjudicated sample is still the only route to *m* we can price, and a review design that is fixed and reported is one a reporting guideline asks for (Collins et al., 2024). It does two other things. It puts published numbers behind the claim in Section 4 that a reviewer’s rate is a property of the reading conditions, since four of the manipulations above moved a rate without altering the material being read. And it adds a requirement to the honest alternative that Section 5 does not price: whoever buys the adjudication has to fix the review design before buying it, because Hofvind et al.’s four answers are all available from the same cases.

## O. The hundred-case cohort

Figure 14 is the contamination model of Section 2 with the rates cleared away. Two of its numbers are sums a reader can check on sight, twenty is three plus seventeen and ninety-seven is eighty plus seventeen, and both are computed from the profile and never typed in, so both hold at each of the four reviewers named under the figure. That list of four carries more than its length suggests, because three percent of a fifth of a hundred rounds up to one: even the reviewer this paper calls attentive puts a mislabelled case into the calibration set.

**Figure 14.**
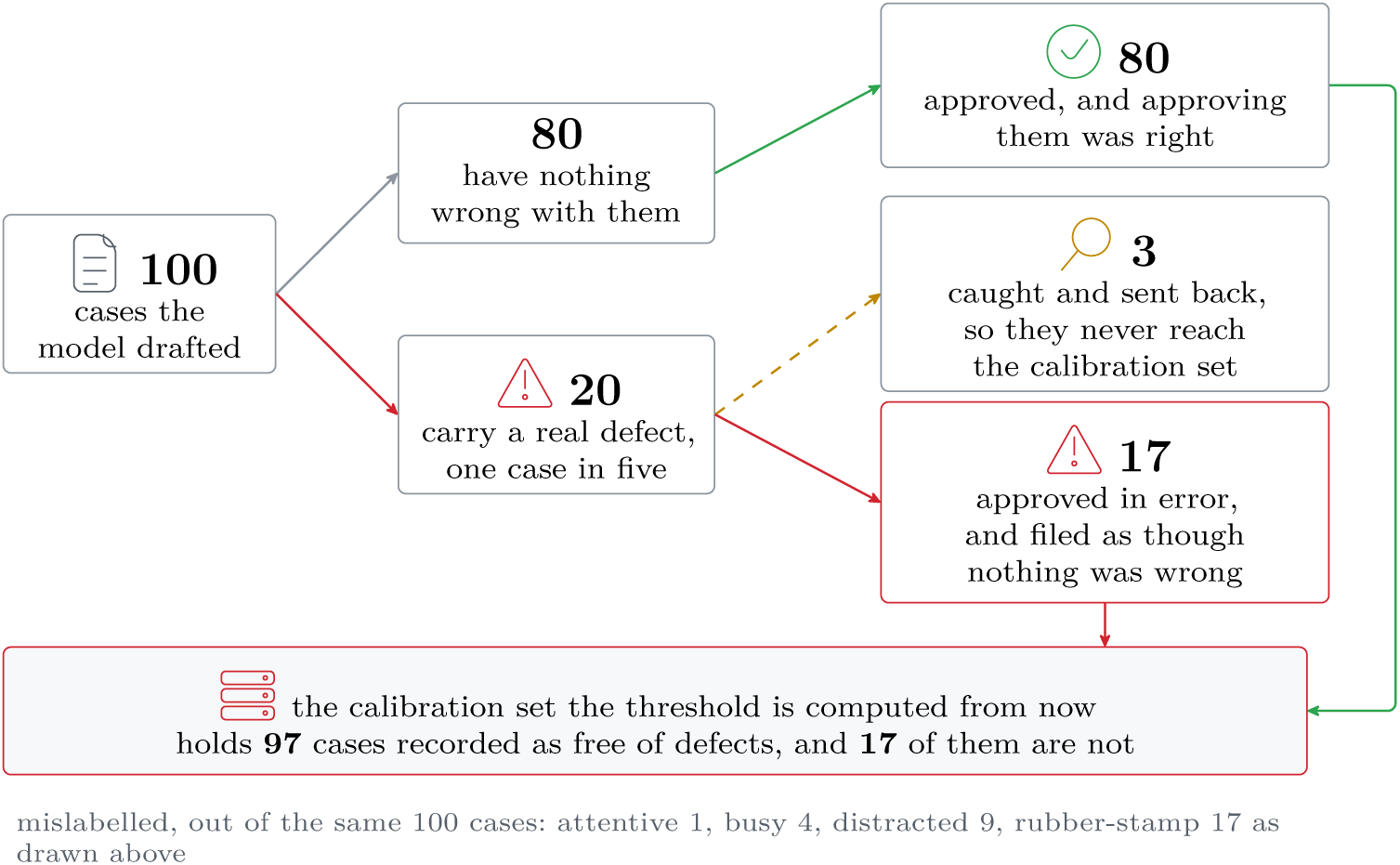
The same contamination as whole cases instead of a rate. One hundred drafted cases, split by what is wrong with them and then by what the reviewer did about it. The seventeen missed defects do not enter the record as disagreements or as gaps: they join the eighty sound cases, so the threshold is computed from ninety-seven cases marked clean, seventeen of them wrongly.

## P. Pipeline structure and published costs

### Two kinds of stage

Every stage in the stack we price is one of two kinds, and Fig. 15 is the difference. Adding a reviewer adds one call, so that bill is a sum and it grows one term at a time. The stage that changes the arithmetic is the one that visits every claim against every document, because its count is a product of two quantities the record decides. The split drawn under the figure comes from re-pricing the identical stack with one extra document and recording which stages moved. That is why 8 of the configured stack’s calls stay put and 304 follow the record.

**Figure 15.**
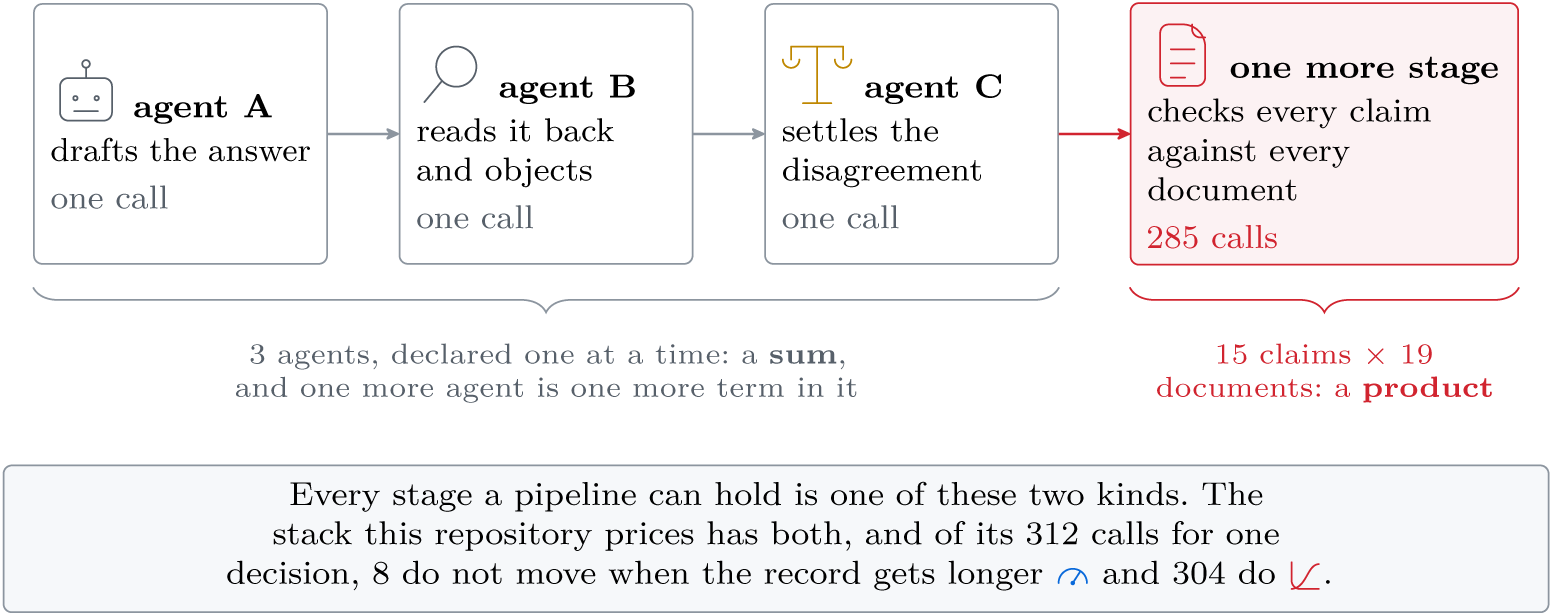
Adding an agent adds a term. Adding a fan-out adds a factor. The first three stages are the pattern our own stack starts from, one call each. The fourth 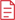 visits every claim against every document, and it is the one whose count the record controls.

**Figure 16.**
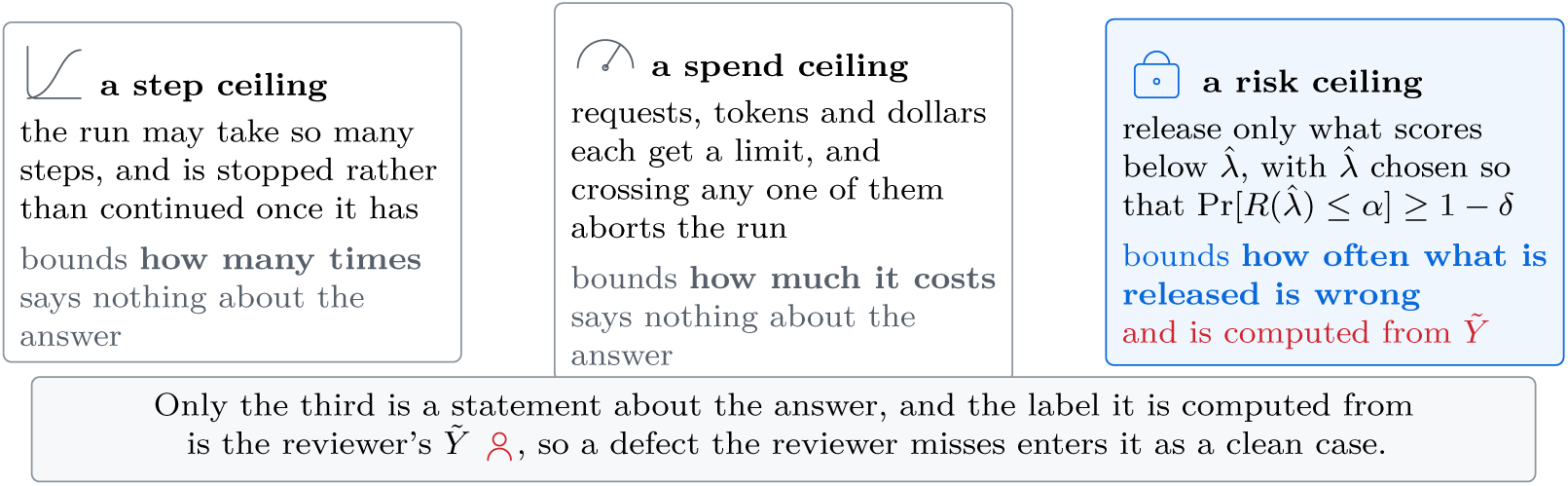
A step ceiling and a spend ceiling bound the run. Only a risk ceiling bounds the answer. The first two are read off each runtime’s own documentation (LangChain, Inc., 2026b; Pydantic Services Inc., 2026) and neither is a defect: a step limit stops a loop and a spend limit stops a bill. Section R on page 50 searches six shipped systems for the third.

#### What the frameworks execute

LangGraph runs a graph as a sequence of super-steps and documents a recursion limit that caps how many of them one run may take, which bounds iterations and says nothing about spend (LangChain, Inc., 2026b). LangChain wraps any step in a retry whose own field documents three attempts as the default, and it retries a step that raised an exception, so a well-formed answer with a fact missing from it is never a retry that library would take (LangChain, Inc., 2026a). DSPy treats each call site as a declaration and compiles the program around it, and its own account of the compiler is that it optimises a program “for quality or for cost” (Khattab et al., 2024), the composition of retrieval with a language model having been introduced under the earlier name in the same line of work (Khattab et al., 2022). AutoGen frames the same composition as a conversation among agents (Wu et al., 2024). Reading the documentation of the mainstream runtimes in September 2026, the one that publishes a monetary ceiling and a request count able to abort a run is Pydantic AI (Pydantic Services Inc., 2026). None of that is a defect in any of them and none of it is a comparison with anything we ship. It is one observation: a stage is cheap to declare in all of them, and the graph is what gets billed.

## Q. What published pipelines paid

The measurements exist, and they agree in direction. Kim et al. report a solo setting that used 6.0 model calls per case against 20.3 for a five-agent group, and say in the same sentence that this is “a high computational cost without a corresponding increase in accuracy” (Kim et al., 2024). That is the medical-domain form of the point. Liu et al. benchmark agent systems on clinical decision tasks and report more than 10 times the token usage and more than 2 times the latency of a single call for what they call “modest performance benefits” (Liu et al., 2026). Poser et al. run a three-provider consensus panel over structured extraction and state that “a consensus model multiplies the costs” (Poser et al., 2026). Outside medicine the accounting is the same. Kapoor et al. find that agent designs reaching much the same accuracy differ in cost by “almost two orders of magnitude” (Kapoor et al., 2024), Smit et al. price a debate strategy that matches a single agent’s benchmark score for roughly eight times the dollar total (Smit et al., 2024), and the first author of the medical study above later holds the compute budget constant while varying only the coordination structure, and identifies a “capability-saturation threshold” past which more agents are unlikely to help (Kim et al., 2026). Cemri et al. catalogue how these systems fail and find the gains over single-agent baselines often minimal (Cemri et al., 2025). Figure 17 puts the four of these that report a factor on one axis, Kim, Liu, Kapoor and Smit.

**Figure 17.**
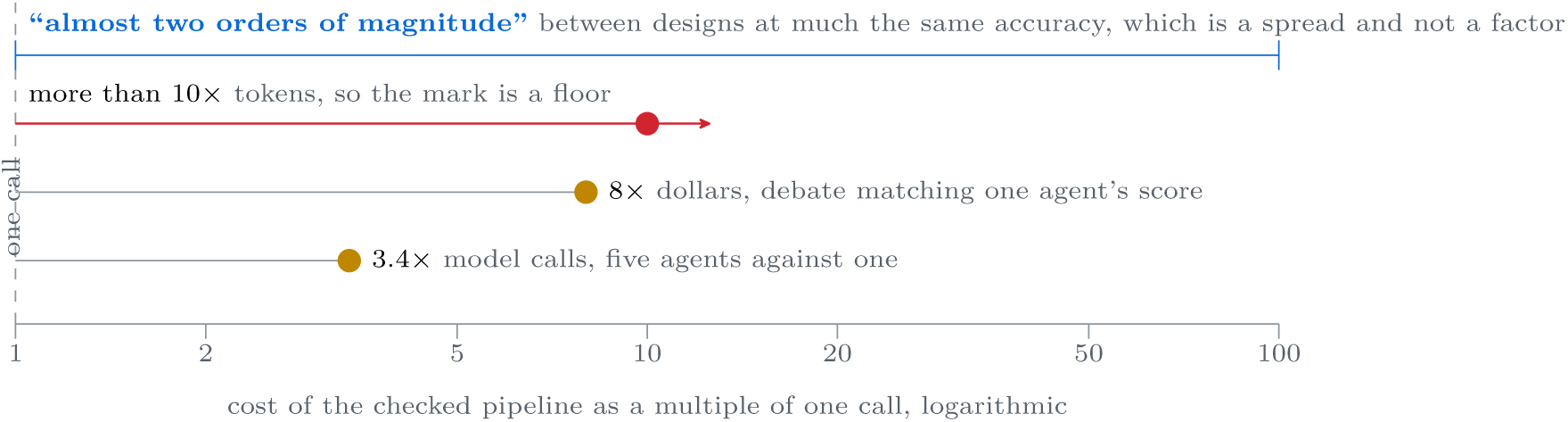
One ruler for four published measurements of what a checked pipeline costs. None of these numbers is ours. They were taken in three different units, so the axis is logarithmic and each mark carries the unit it was measured in, a factor on calls being a different claim from a factor on tokens or on dollars. Liu et al. report a floor, so their mark is an arrowhead. Kapoor et al. measured a spread between designs of similar accuracy, so their mark is drawn as a span.

**Figure 18.**
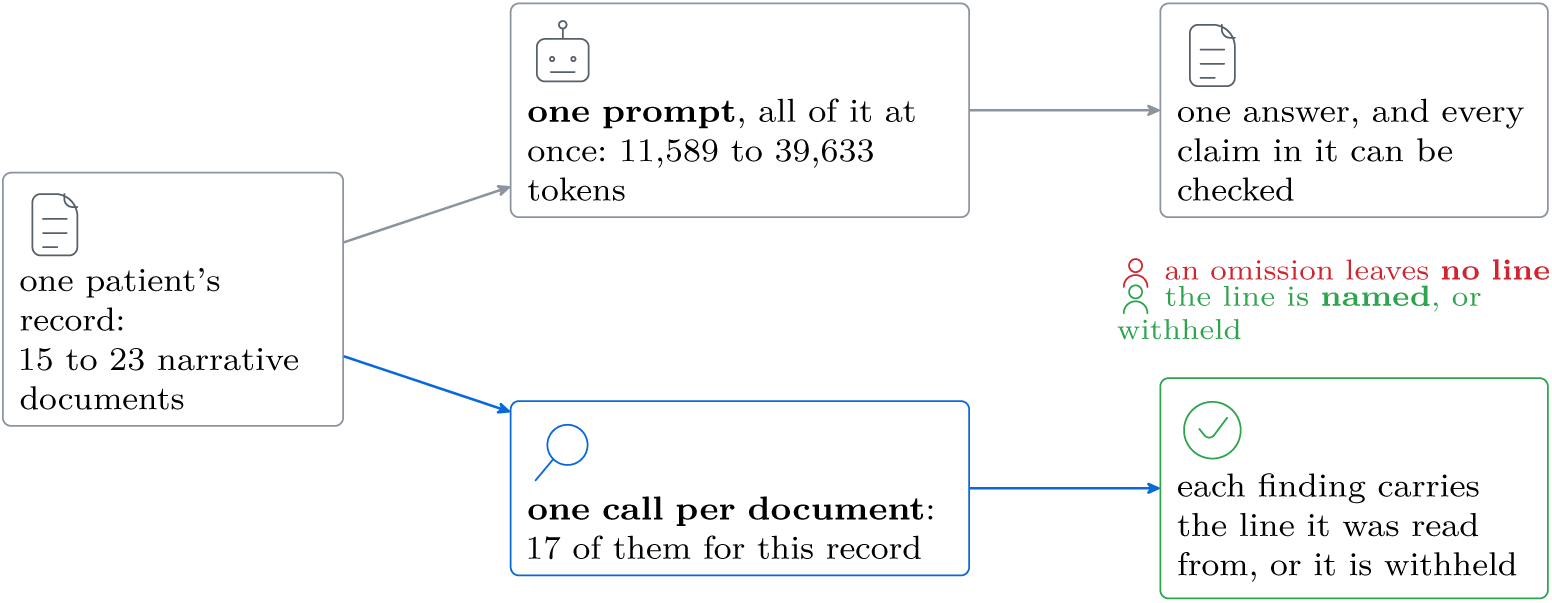
An omission is the one finding a reviewer cannot check against the answer. Both lanes read the same record. Above, one prompt returns one answer and any claim in it is on the page. Below, each document is read alone 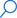, so a finding arrives with the line it came from 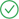 or not at all. This is the fan-out shape the paragraphs above price.

### The cost of a repair loop

A repair loop is a product as well, with the number of rounds as one of its factors. Chen et al. raised the conversation rounds of a multi-agent clinical system to 13 and then to 25 and report that the output token counts rose without a significant performance improvement, at a measured 0.12 dollars per primary consultation (Chen et al., 2025). Huang et al. hold the response budget fixed and find that multi-agent debate scores below plain self-consistency given the same number of responses, which places the apparent gain in the spend rather than in the debate (Huang et al., 2024). An analysis of compound inference systems gives the shape formally: the accuracy of a voting system is non-monotone in the number of calls, rising and then falling (Chen et al., 2024). Our own library carries the smallest version of the same thing. In `src/oversight/agents/litellm_agent.py`, LiteLLMAgent retries a malformed reply up to three times, and each retry appends the failed reply and a correction to the message list, so the third attempt is billed on a longer prompt than the first.

### What a loop returns when it never clears its bar

The sharpest form of the objection is that a developer can wrap an existing program in a retry loop and reach the same assurance for less money, so it is worth reading what such a loop hands back. DSPy ships the pattern as Refine, which takes a module, a number of attempts, a reward function and a threshold, and runs the module up to that many times. Its own source says that it “selects either the first prediction that exceeds the specified threshold or the one with the highest reward” and that “if no prediction meets the threshold, it automatically generates feedback to improve future predictions” (Stanford NLP, 2026). Read the second sentence again. When no attempt clears the bar the call still returns a prediction, the best of the failures, and the returned object carries no field that says the bar was never cleared. A caller who does not keep the reward beside the answer cannot tell a run that passed from a run that gave up. Our rule is the other one. A threshold that is not met yields inconclusive, and inconclusive is not a result.

The loop is also not billed the way a first reading suggests. Each round that fails costs one run of the module and then one further model call, and the prompt of that further call carries the source of the program, the input and output fields of every module in it, the whole trajectory of the round that failed and the source of the reward function (Stanford NLP, 2026). So a round that fails costs more than a round that succeeds, and the rounds that fail are the ones a loop is added to absorb. None of this is a defect in the library that ships it, which does what its documentation says. It is the same observation as before in a smaller frame: the loop bounds how many attempts are made, and nothing in it bounds what the last attempt is allowed to claim.

### The loop priced in the same units

The objection is about money, so it deserves a number. examples/cost_ model.py prices the loop shapes off the same rates and the same per-stage token budgets as the pipeline shapes of Table 11, one attempt billed like the decision call and one check billed like the verification pass. A retry of three attempts whose check is ordinary code spends 3 calls and 0.099 dollars a case at hosted prices, against 2 calls and 0.06 dollars for the leanest shape in that table. A check written in ordinary code cannot see an omission, because seeing one means reading the sources the answer was drawn from, and reading them is a model call. Pay for that and the retry spends 6 calls and 0.194 dollars. Refine costs more again, because its control flow adds a call of its own: it asks for advice after every failing attempt other than the last. That is one call fewer than there are attempts. Three attempts that all fail are therefore 5 calls and 0.163 dollars with a reward function written in ordinary code, and 8 calls and 0.257 dollars with one that reads the evidence. Those two figures understate Refine, because we charge its advice call the failing attempt’s own input and output while the prompt it sends carries the source of the program, the fields of every module in it and the source of the reward function as well.

None of this makes a loop a bad thing to write. The figures say something narrower than that. Every row above spends more than the two-call shape, and not one of them reports a rate at which the answer it returned still carries a defect. That rate comes from held-out cases somebody adjudicated and from a threshold fitted against them, and the two calls are where it comes from.

### Every option in one table

A developer weighing this is choosing between a handful of concrete things, so Table 8 puts them in one place with the price of each. The first row is the zero point, one model call with nothing reading it, because the honest comparison for any check is the bill without it. The rest are the loops above, and the last two columns are the part that does not appear in a price. A loop bounds how many attempts are made. It does not say what the attempt it returns is allowed to claim, so what comes back when nothing clears the bar is an answer either way.

**Table 8.**
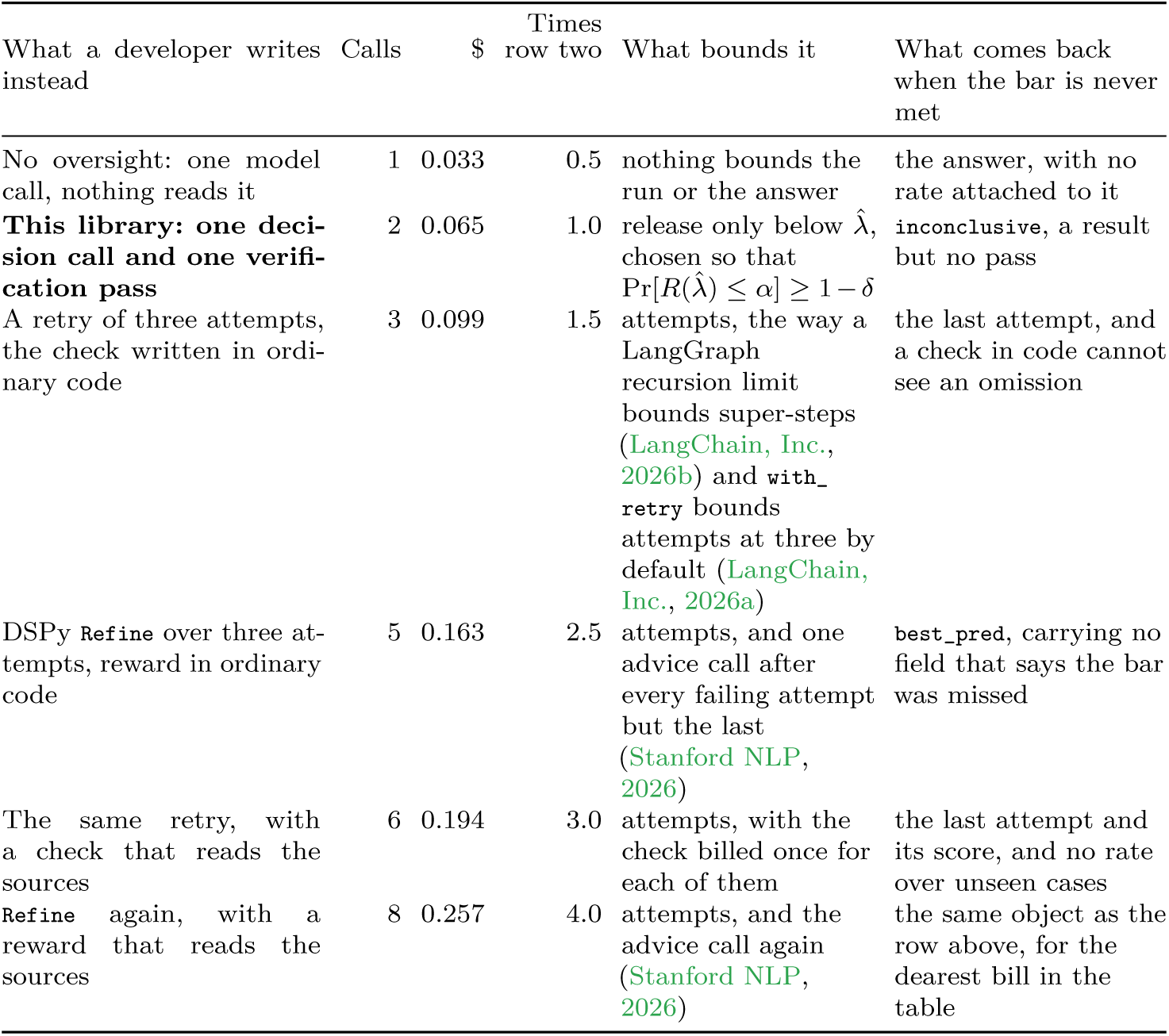
The two calls are the cheapest row here that can see an omission, and the only row that refuses. Each row is one decision at the hosted rate `examples/cost_model.py` prices from, an attempt billed like the decision call and a check billed like the verification pass, so the rows differ in shape and never in rates. A runtime named in a row is where that row’s bound was read from, and naming it is not a comparison with it: the identical loop written by hand bills the identical amount, because the call count follows the shape. A spend ceiling is a third kind of bound again, and the mainstream runtime documenting one able to abort a run is Pydantic AI (Pydantic Services Inc., 2026).

Two readings of that table are worth separating. Read down the money column and the question is what oversight costs. The answer is one further call and about twice the bill of no oversight at all. Read across the last two columns and the question is what any of it buys, and there the loops and the zero point answer alike: an answer, with nothing said about how often an answer like it is wrong. The two-call row is dearer than the zero point and cheaper than every loop, and it is the only row whose output has a third value in it.

### Relevance to identifiability

Section 5 prices the honest alternative in adjudicated cases, and the natural objection is that a pipeline could be made to adjudicate for itself if enough stages were added to it. The measurements above are why that objection does not close. The stages are cheap to declare and the calls are not, the returns are reported as minimal or absent at exactly the point where the count begins to multiply, and no arrangement of models supplies the one input the correction needs. That input is a judgement formed without reference to the reviewer under study.

## R. What shipped guardrail and evaluation systems compute

### One narrow question, asked of the source itself

A reader who has already built oversight out of existing tools will ask what this library computes that those do not, and the question earns an answer only once it is narrow enough to be measured. The narrow form is this: does any of them choose a release threshold whose warrant is a stated confidence? The alternative warrant is a score compared against a threshold an author picks. Learn-then-Test chooses 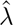 by rejecting a hypothesis at level *δ*, so its warrant is Pr[*R*(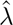) *≤ α*] *≥* 1 *– δ* over the draw of the calibration set (Angelopoulos et al., 2025). That machinery carries a small and particular vocabulary. Its presence in a source tree is therefore a fact about the tree.

### Method and control

Each repository was cloned at the commit named in Table 9, and every source, documentation and notebook file in it was searched for seven strings: conformal, learn-then-test, `learn_then_test`, clopper, hoeffding, bentkus and distribution-free. The extensions covered are Python, TypeScript, Markdown, Colang, Jupyter, reStructuredText, plain text and YAML, because a search that read only source and Markdown would be reading the wrong half of a Python machine-learning repository. A search that finds nothing is worth nothing unless the search is known to work, so each tree was also searched for one string certain to be in it. The controls returned between 81 and 990 files each. The seven strings returned no file in any of the six trees, across 11,433 files in total. The one near miss is a promptfoo document that names risk management as a governance activity, which is not an implementation of risk control.

**Table 9.** The vocabulary of distribution-free risk control is absent from all six trees, and the two control columns are why that absence can be read. Each tree was searched at the named commit on 2026-09-10. A control string certain to be present shows the search working inside each tree, and the last row runs the same seven strings against our own source, where five of them appear. The last column is what each system does compute. That part is not what this table is about: none of these projects advertises a distribution-free guarantee, so the zero is a difference in what is claimed and no defect at all.

| System | Commit | Control string, files carrying it | Of seven | What it computes |
| --- | --- | --- | --- | --- |
| promptfoo (2026) | b8ea544 | redteam, 990 of 4,352 | 0 | assertions and red-team probes over prompts and outputs |
| NeMo Guardrails (2026) | 39b9c5b | colang, 439 of 1,923 | 0 | programmable input and output rails in a rule language |
| DeepEval (2026) | f94d940 | faithfulness, 109 of 2,206 | 0 | metric scores in [0, 1] against a threshold an author sets |
| Ragas (2026) | 298b682 | faithfulness, 81 of 591 | 0 | metric scores for retrieval-augmented generation |
| Guardrails AI (2026) | 06d0ff2 | validator, 196 of 497 | 0 | validators that repair or reject a structured output |
| Inspect (2026) | 7772997 | solver, 403 of 1,864 | 0 | an evaluation harness of solvers and scorers |
| *this library* | *working tree* | `clopper`, 8 of 68 | 5 | the positive control: the instrument reading a tree that does implement it |

Six zeros are not yet evidence, because a search that can only return zero returns zero everywhere, and a reader is entitled to suspect that seven strings assembled by us are seven strings nobody says. So the last row of Table 9 runs the identical search against our own source. That is the one tree we know implements the thing being looked for. Five of the seven come back, clopper in 8 files of 68. The two that do not are bentkus, a bound this library does not implement, and `learn_then_test`, which our source spells as learn-then-test. Neither absence weakens the control, because a control needs the search to work, and no more than that. That row is the instrument reading a known positive, and tools/census.sh halts with an error if it ever reads zero, on the ground that the six rows above it would then be unreadable. The row is a control. It claims nothing about our library, because a null result is only worth the control that sits beside it.

### Scope of this result

An absent string is not an absent capability. A project could implement a Clopper-Pearson interval and name the function something else, and this search would miss it. The census supports something narrower: across six trees that a team would plausibly reach for when asked to put a safety check in front of a clinical model, the standard vocabulary of distribution-free risk control appears nowhere, and the shape of what each one returns is a score, a rule outcome or a validator verdict, and none of them is a bound at a confidence. It is not a ranking. Every one of these systems does something this library does not, and a red-team suite in particular has no counterpart here.

### The same object, a different warrant

The most useful comparison is the closest one. DeepEval documents that every metric takes a threshold parameter, defaulted to 0.5. That parameter decides whether the metric passes or fails, and its documentation gives the default without a procedure for choosing the number against a stated error rate (Confident AI, 2026). Our 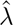 is the same kind of object, a single number a score is compared against, and it is arrived at differently. A threshold an author picks says where the line was drawn. That is the right instrument when the question is whether this release scores worse than the last one, and it says nothing about how often a released answer is wrong, because the number was never referred to an unseen draw. The distance between the two is one hypothesis test at level *δ*. That test is what this library adds, and Section 3 is the finding that adding it buys less than it appears to, because the labels the test consumes were produced by a reviewer who misses in one direction.

### The regulator already writes the gate this way, and that is not a point in our favour

The census above looks at tools a developer would reach for. A second place to look is the file a device maker sends to a regulator, and what is there came as a correction to us. A predetermined change control plan is the document in which a maker says in advance which future changes it may ship without a new submission, and what each change must clear before it ships. We had assumed no authorised plan of that kind fixed its bar on a confidence bound. Four do. Therapixel’s plan for MammoScreen 4 gives its acceptance criterion as “Positive lower bound of the 95% CI of the difference in endpoints between the version under evaluation (MammoScreen 4 on Envision) and the reference version (MammoScreen 2 on Dimension)” (Therapixel, 2025). The plan for Heartvue.Proton takes its bound on the worst case, “Lower bound of the 95% confidence interval of the lowest-performing segmentation output in the validation cohort” (Heartvue, 2026). So the shape of the object in this paper, a threshold that a release must clear at a stated confidence against a named earlier version, is a shape the regulator has already accepted. It is prior art for the shape, and we cite it as such.

### Both plans publish the criterion and withhold the result

Read those two plans again and notice which half is public. The criterion is public and it is numeric. The result of applying it to the release a hospital is running today is not. The user gets a version and a date. Therapixel notifies by “advisory notices sent by email at least 2 weeks before deployment” (Therapixel, 2025), and Heartvue’s notification is documentation that will “identify the modification implemented and the device version in use” (Heartvue, 2026). The plan that goes furthest promises labelling that accompanies each update with “a description of the modification, current Se/Sp and Time to Notification metrics, and a version history of all PCCP-authorized changes” (iSchemaView, 2026), which is a point estimate for each release, not a bound. One plan does print a verdict table, with the columns “Metric *|* Measure *|* Result [95% CI] *|* Pass/Fail”, for the submission alone, and one of its rows reads “Low-Density Non-Calcified Plaque Volume Correlation 0.414 [0.32, 0.45] Pass” (Cleerly, 2026). The margin that row was tested against is not printed anywhere in the summary. A reader therefore cannot tell what a pass on a correlation of that size is a pass against, which is the plainest way to say why a verdict is worth only as much as the threshold printed beside it. That remark is about what a public summary discloses. It is not about any device.

### The honest reading of this, including the part that cuts against us

A vendor operating an authorised plan of this kind computes something close to our number at every release already, because its own plan obliges it to. It does not publish the number. So the gap we are pointing at may be a gap in disclosure alone, and if that is the right reading then what this library contributes is the artefact and the discipline of issuing it, not the statistics underneath. We say it here. Two further limits belong here. All four plans are for imaging and triage devices, so none of them is a bound on what a language model leaves out, and nothing above should be read as saying otherwise. And the substantive documents of several vendors sit behind a login or an email form, so the sentence this appendix will support is that no such per-release statement is public and no vendor publicly claims to issue one. That is weaker than saying none exists.

## S. Three live runs of the shipped pipeline

Everything in the body of this paper about reviewer behaviour is simulated, and Section 8 says so. This section reports the one thing here that is not simulated. We ran the extraction and audit stages the library ships against a hosted model and recorded what came back. The section separates two questions a reader can merge: how often the machine misses something, and how often the person checking the machine misses something. The first is measurable, and is measured below. The second is *m*, and the whole paper is about why a seeded probe cannot supply it.

### What was run

The corpus is the 20 charts at data/charts. Each chart holds a set of clinical notes and a separate structured record of active medications and allergies. A *gold item* is one entry of that record, reduced to a single comparable string: an allergy contributes its full name and a medication contributes the first word of its name, which is the drug with no dose attached, and a medication is taken only when the record marks it active. Both are then lowercased and stripped of every character that is not a letter, so Lisinopril 10 mg becomes lisinopril. An entry is kept as gold only if that string then occurs somewhere in the same reduction of the notes. That last filter is the one that flatters us, and it is stated here and again below: the generated record is larger than the gold set we score against, and what the filter removed is exactly the entries a reader would have had to infer. 130 entries survive it across all 20 charts, and that is the denominator of every recall figure in this section. A named item *matches* a gold item when either of the two reduced strings contains the other, in either direction, which is deliberately generous: a stricter rule would score the formatting. The script `examples/accuracy_report.py` takes each chart through two stages. The first reads the whole set of notes at once and returns the medications and allergies it finds. The second reads each document on its own, is shown what the first stage already found, and is asked to name anything that document adds together with the sentence that supports it. The whole corpus went through both stages 3 times, and the pass reported here is the first of the three, chosen by position before any number was read. Each pass made 370 calls to Claude Sonnet 4.5, at 8 in flight at once, and every call answered, so no row was dropped for a provider failure. Every share below is reported three ways: the fraction it was computed from, that fraction as a percentage, and a two-sided exact Clopper-Pearson interval at *δ* = 0.05. Where a one-sided bound is also given it is marked as one, and it runs in the direction unfavourable to us.

None of these are sensitivity and specificity. The comparison here is against a gold set the generator wrote, not against a clinical reference standard, so the correct name for a share of gold items recovered is a percent agreement with that generated record and nothing stronger. We write it out in full without borrowing the diagnostic-accuracy vocabulary, because we expect the two to be read differently by anyone who assesses a device, and the second reading is one this corpus cannot support.

### What came back

The first stage named 122/130 gold items, a share of 93.8%, in 88.2% to 97.3% two-sided, with a one-sided lower bound of 89.2%. Of the items it named, 123/127 matched a gold item, a share of 96.9%, in 92.1% to 99.1%. The misses were not spread evenly: they fell on 5/20 charts. Reading each document on its own recovered 3/8 of the misses. That is 38% of them, on an interval of 8.5% to 75.5% so wide that it settles nothing on its own, and it brings the two stages together to 125/130, a share of 96.2%, in 91.3% to 98.7% two-sided, with a one-sided lower bound of 92.1%. The fraction of a chart’s documents the second stage flags is a nonconformity score, meaning a number the library can sort releases by without knowing the answer, and ranking charts by it separates the charts that carry a miss from the charts that do not with an area under the curve of 0.933. That one is the weakest number in the section and should be read with its interval attached. It ranks 5 against 15 charts, and resampling the charts within each class puts it in 0.787 to 1.000, an interval whose upper end is a perfect ranking. Nothing here establishes that the score ranks well. It establishes that it is not ranking at random, and no more than that. On timing, the artefact keeps one time for each whole-chart call and a median and a maximum for each chart’s document calls, so the figures we can state are over those per-chart times. They are not over all 370 calls one by one. Their median is 2.6 seconds and the largest of them is 7.7 seconds, at the concurrency above, and one whole chart took 60 seconds through both stages.

### The evidence check

Each of the 87 items the second stage proposed came with a quotation, and the script looks for that quotation in the document the proposal was drawn from, comparing letters only so that reformatting does not count as a mismatch. 1/87 could not be found, which bounds the rate at which this stage supplies evidence that is not in its own source at 5.3% or less. The check therefore fires, and a rule that fires is worth more than a rule that has only ever sat idle: over all 3 passes 1/252 quotations were absent from the document they were drawn from, and each of those proposals was refused. The failure is rare enough that it appears in only 1 of the 3 passes. That is worth stating because it is the reason a shorter run can report a clean zero and mean nothing by it. The gap in that bound should be said in the same breath. It is one narrow failure, a quotation with no source, and it is silent on the more common case of a proposal that quotes a real sentence and draws the wrong item out of it.

### How far the three passes moved

None of the figures above is a fixed property of the model. Across the 3 passes the first stage named between 122 and 123 of the 130 gold items, the two stages together named 125 to 125, the second stage made 80 to 87 proposals, and the area under the curve ran from 0.933 to 0.984. Recall moved little and the ranking moved a lot. This is why the reported pass is fixed by position before any outcome is seen, and the reported pass holds the lowest of those three areas. A reader shown one pass with no spread beside it has no way to tell a stable number from a lucky one, and the build refuses to typeset this section from an artefact that records fewer than two passes.

### Why these are a pilot

`data/README.md` records two properties of this corpus, and both push the numbers above upward. First, the notes and the structured record come out of the same generation pipeline, so the label is not independent of the input the model reads. That is circular evaluation, and it flatters any extractor. Second, gold was filtered to items that appear verbatim somewhere in the notes. That filter removes exactly the cases that need paraphrase or inference and keeps the easy ones. A real chart supplies neither courtesy. The corpus is also synthetic throughout. That is why it could be sent to a hosted model at all, and no real patient record was involved at any point. The fix is known. It is not yet done: have a second annotator label a sample of the charts without seeing the generator, and report agreement. Until that exists, treat every figure in this section as a working measurement of one pipeline against one model on one date, reproducible as a procedure, not as a set of digits.

### None of this measures *m*

A reader who wants a number for reviewer vigilance will be tempted by the recall figures above, and we name that temptation here and refuse it. Recall here is the machine’s, scored against a label the generator wrote. The quantity the certificate needs is the probability that a clinician looking at a released answer fails to see a defect that is there, on the defects that arise naturally. Nothing in this run observes a clinician. Running the pipeline harder, on more charts or a better model, moves the numbers in this section and leaves *m* exactly where Section 3 leaves it.

## T. Both arms in full on the external corpus

Every other measurement in this paper is on material we wrote, and this one is not. The section gives that run whole: what the corpus is and is not, both arms on every stratum, the half of the corpus where the published designs beat ours, the attribution the licence requires, and the two defects the run found in shipped code.

### What the corpus is

OmissionBench is 495 pairs of clinical notes over 112 consultation transcripts, released as ComposoAI/OmissionBench under CC BY 4.0. One note of each pair is a reference a clinician accepted and the other is that note with a single fact removed or altered, so a method is asked to rank the pair rather than to agree with a label. We scored the released evaluation split whole, at SHA-256 3e5ada2d6fdb, and the loader refuses any artefact scored against a different digest, because a figure quoted beside the published baselines has to be on the rows those baselines were measured on.

### What it is not

It is not real patient data, and no sentence in this paper says it is. Two of the four strata come from public corpora, and the ACI-Bench paper gives both of them the same data type in its own table, “role-played dialogue-note pairs”, with usage notes saying the data was “produced synthetically by medical annotators or patient actors in a single institution” (Yim et al., 2023). PriMock57 says the same in its title, “A Dataset Of Primary Care Mock Consultations” (Papadopoulos Korfiatis et al., 2022). The clinicians in both are real clinicians and the patients are actors. Those authors also give the reason no better corpus exists to test on: “collecting, storing, and sharing a large collection of real patient data is a highly legally and ethically challenging endeavor.” That is a limit the whole field works under. The corpus buys something narrower than realism and still worth having: the transcripts are human speech, somebody else wrote them, somebody else planted the defects, somebody else chose the measure, and the numbers we are compared against were published before we ran.

### The measure

Paired discrimination is the share of pairs in which the defective note scores lower, counting a pair scored equally as half. Perfect is 1 and guessing is 0.5. Intervals are percentile intervals from a paired bootstrap over the pairs, at 9,999 resamples, and the sign test excludes ties. One call over the whole transcript scored 0.661 (0.625 to 0.694), sign-test *p* of 1.8 *×* 10*^−^*^17^ on 258 wins, 138 ties and 99 losses. One call per transcript segment scored 0.705 (0.665 to 0.741), sign-test *p* of 5.6 *×* 10*^−^*^22^ on 329 wins, 40 ties and 126 losses.

A consultation transcript carries no document boundaries, so the segmented arm imposes them by cutting on speaker turns at about fifteen hundred characters, a median of 6 segments to a transcript, and the one-call arm is handed those same segments in a single prompt. The two arms therefore differ in how many calls read the same text and in nothing else. Both scored the same 990 notes, two to a pair.

### What the extra calls bought

Table 10 sets the two arms side by side. The segmented arm is 6.06 calls a note against 1, or 5,999 calls against 990 and 14,396,484 input tokens against 6,563,741, and $51.56 against $21.38 for the whole corpus. It took 42 minutes of wall clock against 9, at eight calls in flight in both. The two discrimination intervals overlap, so the extra calls bought a higher point estimate. They did not buy a demonstrated separation. Two things it did buy are not visible in that headline. It tied 40 pairs where one call tied 138, and on the mock primary-care stratum its interval clears one half where the cheaper arm’s does not. Against that, it discarded 1601 findings for an unlocatable quotation where one call discarded 212, and it returned 1 inconclusive note where one call returned 0. A refusal is a result here, and no gap in the table: the detector reports Inconclusive without scoring a note whose source it could not read whole.

**Table 10.**
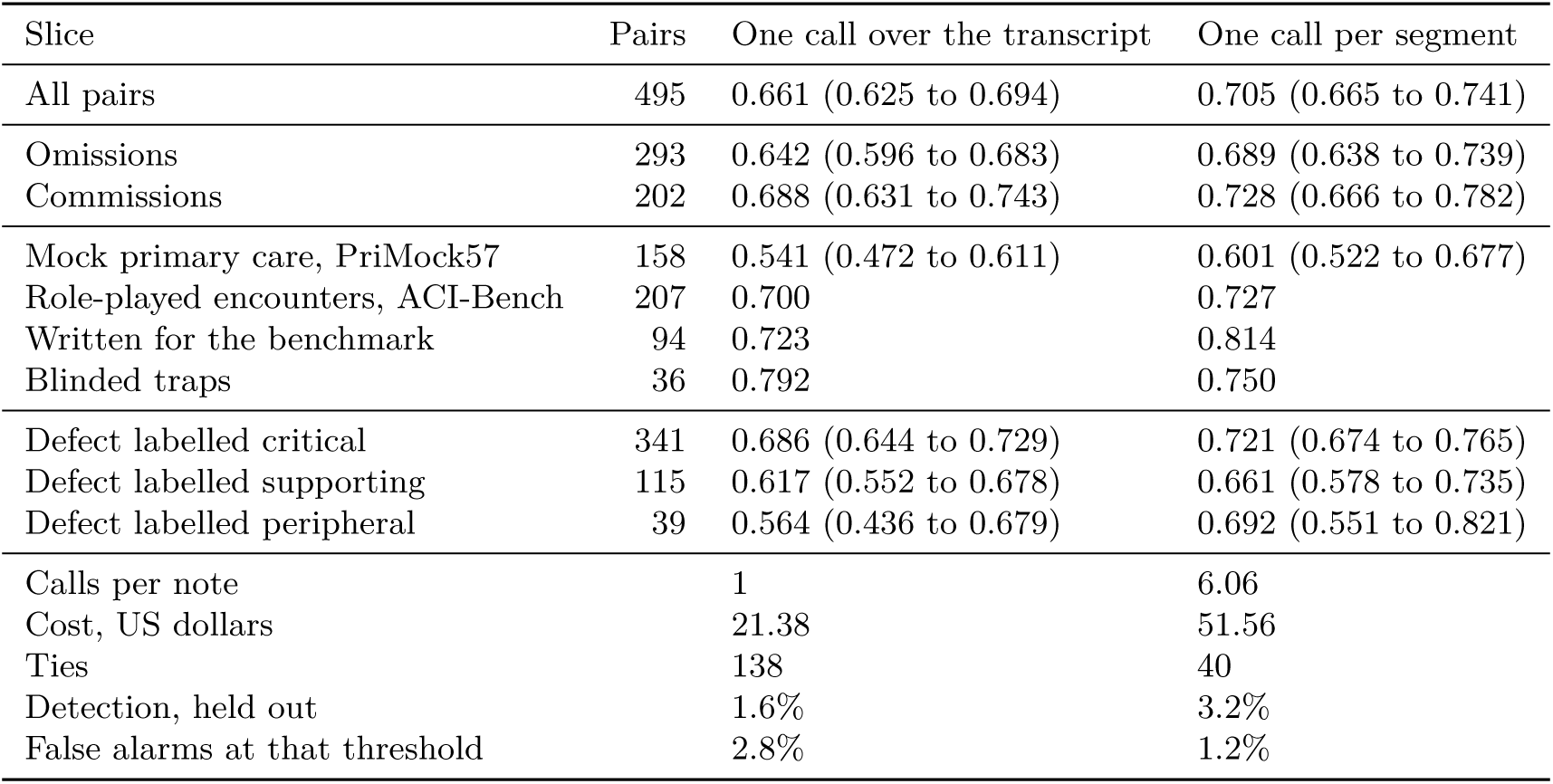
Both arms of the external run, on every stratum, severity and defect type the corpus labels. Chance is 0.5 and intervals are 95 per cent percentile intervals from a paired bootstrap. The two arms read the same text and differ only in how many calls read it. Strata are not one population and the totals should not be read as an average over them.

### Where we stand against the published designs

Every published figure here is read off Tables 4 and 8 of the benchmark’s own paper, and none off its dataset card. An earlier version of this supplement used the card. The card prints a shorter roster and does not say row by row which of its numbers are on the full evaluation set. Two of the sentences that followed from it were wrong in our favour and are corrected below. The judge behind every published figure is GPT-5.4 and ours is Claude Sonnet 4.5, so nothing here is a like-for-like contest. Their paper reports omissions and commissions in separate columns and never blends them, and this supplement follows that, because a design can lead on one class and trail on the other and most of them do. Their own caveat on the measure travels too: “0.500 is chance and it is a ceiling, since production has no clean twin.” Everything in the three paragraphs below is on the full 495-pair evaluation set. Their held-out subset is treated separately at the end.

On omissions their eight one-prompt designs run from 0.500, which is exactly a coin flip, up to 0.634. Both our arms are above all eight, but our one-call interval (0.596 to 0.683) contains the top of that range, so the honest reading is that the cheap arm matches the best of the eight rather than beating it. Three further designs in their table enumerate the transcript’s facts before scoring: a checklist of about twenty-five generated items at 0.647, which falls between our two arms, the three-stage pipeline they recommend deploying at 0.795, and a deliberately naive coverage recipe checking about sixty-two unaudited facts per consultation at 0.809, the highest omission figure in their table. The last two are above the top of both our intervals, and their other recommended design, a single call with an optimised prompt, scores 0.670 and falls between our two arms. **So the published work contains a design that beats our better arm on omissions on the same pairs, and this paper does not claim otherwise.** Their own reading of why is that enumerating facts, rather than decomposing the judgement, is what recovers omissions. That recipe’s two paired figures are computed over fewer pairs than every other row, because parse failures cost that arm some of its records: 289 omissions and 200 commissions in place of the full 293 and 202, 14 records short.

Enumeration is paid for on the other axis. On commissions the eight run from 0.792 to 0.944, and every one of them leads both our arms, which score 0.688 (0.631 to 0.743) on one call and 0.728 (0.666 to 0.782) on segments. That much this supplement said before and it stands. It also said that our arms sit below every design in their table on this class, and that is false. The checklist scores 0.744 and leads both our arms, but the coverage recipe scores 0.712 and their own recommended pipeline 0.661, and our segmented arm is above both. The pattern is not that they lead us and we trail: it is that whatever enumerates facts gains on omissions and loses on commissions. Their own two recommended designs bracket the range, 0.661 for the enumerating pipeline against 0.978 for the single call, a spread on this class wider than the distance between either of them and us.

At a threshold the comparison turns against us, and it does so against two rows in particular. Their two recommended deployments are read at their own rules on these same pairs: the pipeline catches 24.6% of defective notes at 2.7% false alarms for $0.45 a note, and the prompt-optimised single call catches 36.9% at 6.2% for $0.046. Ours catches 1.6% and 3.2%. That is the sharpest published result this run falls short of and no weaker row is quoted in its place. Against the eight the picture is less one-sided, because a detection figure only reads next to the false alarms beside it. The four of the eight that answer with a score run 1.0% at 1.5%, 2.4% at 2.1% and 4.7% at 5.7%, then 8.3% at 6.5%. Our segmented arm’s 3.2% at 1.2% is above the first two of those on both counts. The four that answer yes or no reach as high as 42.3% and flag 32.1% of clean notes doing it. That is outside the 10.0% their paper treats as the most a deployment could carry.

The strongest thing in their data is not a ranking and it is not ours. The coverage recipe carries the best omission score anyone has measured on this corpus and flags 98.7% of clean notes at its natural rule, so it is unusable as written. Swept to a calibrated threshold at their 10.0% budget it reaches 20.9%, against 8.3% for the best of the eight. Their paper draws the conclusion in one line: “A detection number with no false-alarm rate beside it is uninterpretable.” That is the same separation between ranking and thresholding this paper argues from its own tie counts, established here on somebody else’s corpus, by somebody else, against designs we did not choose.

Their held-out confirmation subset is a different set of pairs, 151 of them, and their Table 8 states outright that its rows are not directly comparable to the others. Two omission figures sit there, the extract-then-check pipeline at 0.786 and the same pipeline with a severity-grading stage at 0.762. Neither is compared with ours anywhere in this paper, and they are named only so that a reader who opens their table knows which rows we left out and why.

### The threshold is the weak result

Ranking a pair is a different task from setting the threshold a deployment has to set, and this is where the run is worst. Completeness in the library is one over one plus the flagged burden, which is coarse: many notes land on the same few values. Fixing the false-alarm budget at 2.1%, choosing a threshold on one half of the pairs and evaluating on the other, one call detects 1.6% of defective notes at 2.8% false alarms, having detected 3.2% on the half it was fitted to, and segments detect 3.2% at 1.2% against 4.5% fitted. Neither transfers intact and neither is near either of their two deployment rules. The thresholds themselves are 0.1064 and 0.0384 on the completeness score, and they are reported so that a reader can reproduce the two operating points. The mechanism is the ties: a tie costs a ranking measure half a win, and gives a threshold nothing at all. The remedy is a less lumpy score, not a different measure, and it is not done. On notes carrying no planted defect the held-out threshold nonetheless flags 2.7% on one call and 0.9% on segments.

### Do not pool the strata

Table 10 reports each of the four separately because they are not one population, and the card says so itself: its own generation pipeline failed on about 52 per cent of first-pass pairs drawn from the two public corpora against 18.9 per cent on the transcripts written for the benchmark. The stratum nearest the setting this paper describes is the mock primary-care one, and it is the weakest of the four in both arms. On one call it is 0.541 (0.472 to 0.611), an interval containing one half, which means that arm has shown nothing there. On segments it is 0.601 (0.522 to 0.677). Reading the paper as though the 0.705 headline held on primary-care consultations would be reading it wrongly.

### The severity ordering runs the right way, weakly

The corpus labels each planted defect critical, supporting or peripheral, and a detector worth deploying should do best on the critical ones. It does, in both arms: 0.721 (0.674 to 0.765) on segments against 0.661 (0.578 to 0.735) on the supporting ones. Read that ordering as reassurance only. The intervals overlap, the peripheral cell holds 39 pairs and its interval is correspondingly wide at 0.551 to 0.821, and we did not fix the ordering as a hypothesis before looking. A corpus with the severities balanced by design would settle it, and this one is not balanced.

### Two defects this corpus found in shipped code

Neither was found by a test we wrote. The first is in the evidence check. _locate in src/oversight/attest/verifier.py required a cited quotation to appear character for character in a supplied source, and a consultation transcript puts a space before every comma and wraps mid-sentence, so a model quoting one faithfully but tidying its spacing produced a quotation that no source contained and the finding was discarded as ungroundable. It now retries with whitespace deleted from both sides, and only for a quotation of at least twenty-four characters, because deleting whitespace joins words and a short quotation could then match a join the source never made. The floor is named SQUEEZE_ FLOOR and a test holds it in place.

The second is in the benchmark harness. `assert_scoring_blind` removed the note from the prompt before checking that the ground-truth defect did not appear in what was left, and it removed only the first copy, so a prompt carrying the note more than once, which is exactly what the segmented arm produces, could pass the blindness check with a copy still in the leftover. It now removes every copy. Both fixes ship with tests that fail against the previous code.

### Attribution

The two public strata carry the attribution their licences require, and it is reproduced verbatim from `examples/omissionbench_report.py`. That script writes it into the attribution field of every score file the run deposits. For the first: “PriMock57, Babylon Health, licensed under CC BY 4.0, github.com/babylonhealth/primock57. Changes made: transcripts normalised; the clinician reference notes were audited against their own transcripts and repaired; single-error variants were generated from the repaired notes.” For the second: “ACI-Bench, Yim et al. 2023, licensed under CC BY 4.0 on its figshare record, doi.org/10.6084/m9.figshare.22494601 (code at github.com/wyim/aci-bench, which carries no licence file of its own). Changes made: a 48-encounter subsample drawn at seed 20260728 from the 140 encounters remaining after 67 exclusions; transcripts normalised and rendered to synthetic speech; the dictation-derived reference notes were audited against their own transcripts and repaired; single-error variants were generated from the repaired notes.” We redistribute none of either corpus. The deposit carries per-pair scores, the loader, and the digest of the split we read.

### What this run does not measure

It does not measure *m*. It measures a detector against defects somebody planted. That is the machine’s recall. The reviewer’s miss rate is a different quantity, and Section 3 is about why the second cannot be read off the first. One further caveat belongs here because it bears on every figure above. The provider is not deterministic at temperature zero: a probe of one consultation’s thirteen segments, rerun on identical input, returned three ungroundable findings on the first trial and twenty-seven on the second. Every number in this section is therefore one run of one shape against one model on one date. It reproduces as a procedure, never as a set of digits, and that is the same caution Section S attaches to the chart-reading run.

## U. Four pipeline shapes priced stage by stage

Table 11 is what `examples/cost_model.py` prints. It prices a shape by summing its stages, and it says on its own first line of output that every per-stage token count is an assumption, so the money columns are projections. The call column is not an assumption. It follows from the shape: a stage that fans out over every claim and every source is a product, and a product grows where a sum does not. Between the first row and the last the calls fall by a factor of 156 and the input tokens fall from 960,000 to 14,000, which is the same fact told in tokens. At 100,000 decisions a year the gap between the two rows is the difference between a line item and a programme, and because the token counts are assumptions the ratio is the claim and the absolute figure is not. Section V counts four of those token budgets instead of assuming them, and reports what the ratio becomes.

**Table 11.** The shape of the graph moves the bill further than the choice of vendor. Calls and cost are both for one decision, and the two money columns are the same four shapes priced at a frontier hosted rate and at a self-hosted endpoint, so reading down a column shows what shape buys and reading across a row shows what vendor buys.

| Pipeline shape | Calls | \$ frontier | \$ self-hosted |
| --- | --- | --- | --- |
| Full stack, three-model panel, fan-out over every source | 312 | 3.90 | 0.303 |
| Retrieval-scoped fan-out, three-model panel | 39 | 0.56 | 0.043 |
| Single model, attestation only | 34 | 0.45 | 0.034 |
| Lean: one model, one verification pass | 2 | 0.06 | 0.005 |

The accuracy of the last row is not a projection, because we ran a shape with that call count. Section W reports what it recovered and what it gave up. Its token budget is another matter, and the difference runs against this row. The row prices a verification pass over retrieved evidence, and the arm we ran was shown every document of the record, or 58,190 characters at the median. So the money in this row is below what the arm we measured would be billed, and the call count is what the two have in common.

## V. The assumed token counts against measured ones

Every token budget in Table 11 was a number we wrote down. `examples/token_census.py` counts all 9 of them. It counts them without changing the library, which matters, because a measurement of a modified copy is a measurement of the copy. Two things in this library take a model from the outside. The map-reduce verifier takes the function that calls a model as an argument to its constructor, and every screening and verification stage takes an object with a decide method. Both are how an integrator supplies their own provider. The census passes a function and an object that call the model and, before handing the reply back, keep the two numbers the provider reports about that call: how many tokens it read and how many it wrote. The prompts, the fan-out and the response contracts are therefore the shipped ones. Nothing is counted on this machine, so no local counter can disagree with the provider that sends the bill.

The decision stage is counted separately, because its prompt belongs to whoever deploys the pipeline. The prompt used is the one the chart study of Section S already runs, over all 20 charts of the corpus, so the number is that study’s number. One run of the whole census is 368 calls, 183 seconds and 5.06 US dollars at the frontier rate this appendix prices from. It read 1,572,144 tokens and wrote 23,130, all of it against Claude Sonnet 4.5.

The first assumption to fall was not a token count. Table 11 assumes a decision breaks into 15 checkable claims. The shipped splitter produced 17 from the case, and each of them was scored against all 19 documents of the record, so the scoring stage made 323 calls where the table prices 285. That is the fan-out working exactly as described and costing more than described, because the multiplier was the thing assumed. Every other stage made the number of calls the table prices.

Table 12 has one pattern running down each half of it, and the two patterns point in opposite directions. On the input side, every stage that reads the whole record cost more than budgeted and every stage that reads one document or one short text cost less. On the output side, every stage that answers with a structured verdict wrote less than budgeted, and the two that write a list wrote more.

**Table 12.** Assumed token budget beside the token count the provider reported, for every stage the shape table prices. Rows are in the order the pipeline runs them, and a number in brackets after a stage is how many calls it made. The counted columns are the median call, and the ratio of counted to assumed is on the right, so a ratio near one means the budget was close and a ratio far from one is the size of the error.

| Stage | Input tokens |  | Output tokens |  | Counted over assumed |  |
| --- | --- | --- | --- | --- | --- | --- |
|  | assumed | counted | assumed | counted | in | out |
| Content safety screen | 2,000 | 15,288 | 100 | 178 | 7.64 | 1.78 |
| Prescreen | 4,000 | 15,085 | 100 | 135 | 3.77 | 1.35 |
| Decision over one chart [20] | 8,000 | 17,404 | 600 | 50 | 2.18 | 0.08 |
| Groundedness check | 8,000 | 15,535 | 400 | 404 | 1.94 | 1.01 |
| Backward verifier | 6,000 | 15,153 | 300 | 120 | 2.53 | 0.40 |
| Claim splitting | 4,000 | 634 | 800 | 1,514 | 0.16 | 1.89 |
| Claim scoring [323] | 3,000 | 2,758 | 200 | 32 | 0.92 | 0.16 |
| Omission scan [19] | 3,000 | 2,871 | 400 | 343 | 0.96 | 0.86 |
| Verification pass, retrieved evidence only | 6,000 | 4,149 | 900 | 200 | 0.69 | 0.22 |

The input side first, because it is the side that carries the money. Four stages are handed the case prompt itself, and they are content safety, the prescreen, the groundedness check and the backward verifier. Their input size is therefore fixed by the record, and the census reports here only what the pipeline already does. The largest error in the table is there: the content safety screen was priced at 2,000 tokens and read 15,288, 7.64 times the budget, because a screen for injected instructions and for leaked identifiers has to read every word that is about to be sent. The prescreen is 3.77 times its budget for the same reason, the backward verifier 2.53 times and the groundedness check 1.94 times. The decision stage over a whole chart read 17,404 tokens at the median chart and 45,164 at the largest one, against 8,000 budgeted. That is 2.18 times. That one matters most of all, because the panel pays for it once per model.

The stages that read less than budgeted read less because they read less. Scoring one claim against one document is 2,758 tokens against 3,000, the omission scan over one document 2,871 against 3,000, and the single verification pass over retrieved evidence 4,149 against 6,000. The splitter is the extreme case, at 634 tokens against 4,000, because all it reads is one short determination.

The output side is simpler and the error is one-sided. A score is a severity word, two numbers and one quoted line. That came to 32 tokens against 200 budgeted. A verdict from the backward verifier is 120 against 300, and a decision over a chart 50 against 600. The exceptions are the two stages that return a list where the others return a verdict. The splitter writes one claim for every checkable sentence, 1,514 tokens against 800, and the safety screen writes 178 against 100.

The honest reading of Table 13 is that the errors ran in both directions and largely cancelled. Two shapes come out cheaper than the table projected and two come out dearer, and no shape moves by half. The widest shape falls from 3.90 to 3.47 because most of its bill is the fan-out, and the fan-out reads one document at a time. The retrieval-scoped shape rises from 0.56 to 0.65 because it has a short fan-out and the same four stages that read the whole record, so the understated stages are a larger part of a smaller bill. So the ratio the previous section offered as its claim, the widest shape over the cheapest, falls from 60 to 51 in dollars while the same ratio in calls rises from 156 to 175. The claim that shape moves the bill further than vendor choice survives the counting. It is smaller than we projected it, by about a sixth.

**Table 13.** The four shapes priced twice, once off the assumptions and once off the counts. The shapes are the four of Table 11, named there in full. No column here rests on a token budget nobody measured, the difference between this table and that one.

| Pipeline shape | Calls | | \$ frontier | | \$ self-hosted |
| --- | --- | --- | --- | --- | --- |
|  | assumed | counted | assumed | counted | counted |
| Full stack | 312 | 350 | 3.90 | 3.47 | 0.318 |
| Retrieval-scoped | 39 | 43 | 0.56 | 0.65 | 0.060 |
| Single model | 34 | 38 | 0.45 | 0.40 | 0.036 |
| Lean | 2 | 2 | 0.06 | 0.07 | 0.006 |

Two limits belong on all of this, and neither is a token count. The charts are generated, because both annotated clinical corpora the field uses sit behind a data use agreement, so every input count above is the count for this corpus. A record whose documents are longer moves the input side of every stage that reads one, and the decision stage most of all. The direction of each error does not depend on the corpus, and the direction is the pattern this section reports. The second limit is that the backward verifier was shown no history, because our store holds no adjudicated outcome for this case kind yet, so its prompt carried the sentence that says so, with no sample size and no rate in it. That is the shipped behaviour of an installation with no outcomes in it, and it is a few tokens shorter than the alternative.

## W. The cheap shape and the ranking it gives up

A reader who has read Section S and Section U together is owed an obvious question. The cheapest row of Table 11 costs a fraction of the widest one, so what does buying the widest one get? The two tables above cannot answer it. One of them prices shapes and assumes their token counts, and the other measures accuracy for a single shape. So we ran the cheap shape as well.

The script that produced Section S takes one flag, and three of its settings are compared here. The first sends the audit stage one call for each document of a chart. The second merges every document of that chart into a single block of text and sends one call. The third also sends one call, but it leaves the documents separated inside that prompt and puts each finding back into the document whose line it quotes. We call these three the fan-out arm, the joined arm and the placed arm. The flag has two further settings. They escalate from the cheap audit to the fan-out inside a single run, and the last part of this appendix reports them. Everything else is held fixed across all three: the same charts, the same gold items, the same model, the same prompts, the same number of calls in flight, and the same 3 passes over the corpus. A difference between the arms is therefore attributable to the fan-out and to nothing else the script happened to choose. The build checks this. It refuses to typeset this section if the artefacts disagree on the model, the corpus, the gold count, the chart count, the concurrency or *δ*.

The third arm was added because of what the second one seemed to prove, and the order matters here. The joined arm was run first, it ranked badly, and the reason looked like arithmetic. So we built an arm that removes that reason while paying the same price, to see whether the ranking came back.

The single audit call is shown 57,668 characters of source text at the median, so it reads the whole record. That sentence is here because an earlier comparison of these two shapes was withdrawn for exactly that fault. A character cap sat on the joined arm, so that arm was shown a small part of the corpus, and the same cap decided the labels it was scored against. The gap it reported was the cap talking. The caps are gone from the script, and this is the first comparison of the two shapes in which both arms read the same text.

### Recall came out the same, within what this corpus can resolve

Both stages together named 125/130 gold items in the joined arm, 123/130 in the placed arm and 125/130 in the fan-out. As shares those are 96.2%, 94.6% and 96.2%. The first stage is the same call in all three arms, sent the same prompt, so its own spread is a measure of what a repeat of this corpus can resolve and nothing else. It named 123/130, 121/130 and 122/130. Two items separate the ends of that, from a stage that did not change, so a gap of two items between arms is inside the noise. Across the 3 passes the joined arm’s first stage named 122 to 123 gold items and both stages together named 124 to 125. On this corpus, then, the fan-out is not buying recall.

### The ordering came out different

The area under the curve falls from 0.933 to 0.633, and across the passes the joined arm ran from 0.633 to 0.767. This is the number that tells a reviewer which chart to open first, and it is the one the cheap shape gives up.

### The reason looked arithmetical, so we tested it

The score the library sorts releases by is the fraction of a chart’s documents that raised a finding. Merge every document into one call and that fraction has one denominator, so it can only come out as none of them or all of them. A score that takes two values cannot put twenty charts in an order. The artefacts say so when counted: over the whole corpus the joined arm’s score took 2 distinct values and the fan-out’s took 14. All of that is arithmetic and none of it is in doubt. It does not establish the next step, that the denominator is what the extra calls are being paid for. That step is a claim about cause, and a count of values cannot carry it.

### The denominator came back and the ordering did not

The placed arm sends one call, like the joined arm, and pays 2 calls a chart. But the documents stay separated inside its prompt, and each finding is put back into the document whose line it quotes, so the fraction gets its denominator back. It does: the placed arm’s score took 8 distinct values over the corpus, against 2 for the joined arm. The placement is not achieved by guessing, either, because 0 of its 16 proposals quoted a line that landed in no document. And the ordering did not follow. The area under the curve came out 0.655 against 0.633 for the joined arm and 0.933 for the fan-out, it ran from 0.655 to 0.734 over 3 passes, and its interval 0.381 to 0.899 still covers one half.

We are correcting that explanation here. A two-valued score cannot rank, and giving the score more values did not make it rank. Whatever the extra calls are buying, on this corpus it is not the denominator alone. The plainest guess left is that a call which is shown one document has to reach a verdict about that document, while a call which is shown twenty may spend its attention on a few of them and stay quiet about the rest. Being asked once about twenty documents is not the same task as being asked twenty times about one. The two escalating arms of the same flag do send calls that see one document, so they are worth ruling out as a test of this. They rule themselves out: they send such a call for 13 and 106 documents of 350, and only for documents the cheap audit had already pointed at. A test of the guess has to keep the cheap price and put the question about every document all the same. The fourth arm does exactly that.

### The fourth arm asks the cheap call for a verdict on every document

It sends one audit call and pays 2 calls a chart, the price the other two cheap arms pay. The documents are numbered inside that one prompt, and the reply has to carry one entry per number: a yes or a no, with the item and the line it quotes when the answer is yes. A document the reply says nothing about is counted as flagged, on the reasoning that a document nobody answered for is not a document nobody needs to read. Over 3 passes the reply left out 0 documents, so the question was put and answered for every document in the corpus. The score took 8 distinct values, which is what the placed arm took. Both stages together named 123 of 130 gold items, 94.6% with interval 89.2% to 97.8%. The area under the curve came out 0.649 with interval 0.393 to 0.881 over 6 against 14 charts, and across the passes it ran from 0.649 to 0.780. That interval covers one half, as the intervals of the other two cheap arms do.

### What separates the arms is how often they flag a document at all

Table 15 counts the documents each arm flagged over the whole corpus. The arm that reads one document at a time flags 59 of 350 of them, 16.9%. The arm that reads every document and has to answer for each flags 10 of 350, 2.9%. The arm that reads every document and files findings by quote flags 13 of 350, 3.7%. The cheap call is not waiting to be asked, then, and it is not short of a denominator. It reads the whole record, judges the list of found items to be complete and answers no. Isolation is what produces the flags, and the flags are what carry the ordering.

**Table 14.** Nine times the calls bought no extra recall on this corpus. They bought the ordering, and giving the cheap shape its denominator back did not buy the ordering. All three arms scored the same 20 charts against the same 130 gold items with the same model on the same day. Calls per chart are medians, and every interval is two-sided at *δ* = 0.05. The middle column costs what the left one costs and scores the way the right one does on the row of distinct score values, and it still does not rank.

|  | One call, merged | One call, placed | One call per document |
| --- | --- | --- | --- |
| Provider calls per chart, median | 2 | 2 | 18 |
| Provider calls, whole corpus | 40 | 40 | 370 |
| Gold items the first stage named | 123/130 | 121/130 | 122/130 |
|  | 89.2% to 97.8% | 87.3% to 96.8% | 88.2% to 97.3% |
| Gold items either stage named | 125/130 | 123/130 | 125/130 |
|  | 91.3% to 98.7% | 89.2% to 97.8% | 91.3% to 98.7% |
| Of the items named, matching gold | 124/128 | 122/126 | 123/127 |
| Misses the audit stage recovered | 2/7 | 2/9 | 3/8 |
| Items the audit stage proposed | 21 | 16 | 87 |
| Distinct values the score took | 2 | 8 | 14 |
| Area under the curve, ranking charts | 0.633 | 0.655 | 0.933 |
|  | 0.400 to 0.867 | 0.381 to 0.899 | 0.787 to 1.000 |
| Charts with a miss against without | 5 against 15 | 6 against 14 | 5 against 15 |
| Seconds for one whole chart, median | 15 | 14 | 60 |

**Table 15.** The arm that flags a document six times as often is the arm that ranks, and the two arms that read the record in one call flag rarely whether or not they are made to answer for each document. The three arms scored the same 20 charts against the same 130 gold items with the same model on the same day. Flag counts are over every document of every chart. Calls per chart are medians and every interval is two-sided at *δ* = 0.05.

|  | One call, placed | One call, verdicts | One call per document |
| --- | --- | --- | --- |
| Provider calls per chart, median | 2 | 2 | 18 |
| Provider calls, whole corpus | 40 | 40 | 370 |
| Documents flagged, whole corpus | 13 of 350 | 10 of 350 | 59 of 350 |
| Documents flagged, as a share | 3.7% | 2.9% | 16.9% |
| Distinct values the score took | 8 | 8 | 14 |
| Gold items either stage named | 123/130 | 123 of 130 | 125/130 |
| Area under the curve, ranking charts | 0.655 | 0.649 | 0.933 |
|  | 0.381 to 0.899 | 0.393 to 0.881 | 0.787 to 1.000 |
| Seconds for one whole chart, median | 14 | 22 | 60 |

### What the fourth arm settles and what it leaves open

Why a call shown one document flags so much more often is not measured here, and neither is whether those extra flags are right. The fan-out arm’s recall is no better than the cheap arms’, so the documents it flags and they do not cannot all be carrying a real miss, and some of them are noise which happens to fall on a chart that carries one. The settled part is narrower, and it is the part a person deciding how much to spend needs. Three ways of reshaping the cheap prompt have now been run on this corpus: merging the record into one block of text, keeping the documents apart and putting each finding back where its quote lives, and numbering the documents and demanding an answer for each. Two escalations on top of the cheap flag have been run as well, and Table 16 reports them. None of those five recovers the ordering. A deployment that needs a queue with the worst case at the top has to pay for calls that read one document at a time, and reshaping the prompt of a single cheap call is not a substitute for paying it.

**Table 16.** Escalating on the cheap audit’s own findings cost more than the cheap audit and ordered the charts no better. The two middle columns were run end to end over 3 passes on the same 20 charts, the same 130 gold items and the same model as the two outer ones. Areas are two-sided at *δ* = 0.05. The build refuses to typeset this table if the four artefacts disagree on the model, the corpus, the gold count, the chart count, the concurrency or *δ*.

|  | Cheap only | Escalate on a finding |  | Fan-out only |
| --- | --- | --- | --- | --- |
|  |  | by document | whole chart |  |
| Provider calls, whole corpus | 40 | 53 | 146 | 370 |
| Charts escalated, of 20 | none | 7 | 6 | all |
| Documents given their own audit call | none | 13 | 106 | 350 |
| Gold items either stage named | 125/130 | 123/130 | 123/130 | 125/130 |
| Misses the audit stages recovered | 2/7 | 1/8 | 1/8 | 3/8 |
| Items the audit stages proposed | 21 | 39 | 64 | 87 |
| Distinct values the score took | 2 | 7 | 6 | 14 |
| Area under the curve, ranking charts | 0.633 | 0.580 | 0.627 | 0.933 |
|  | 0.400 to 0.867 | 0.333 to 0.840 | 0.367 to 0.887 | 0.787 to 1.000 |
| Charts with a miss against without | 5 against 15 | 5 against 15 | 5 against 15 | 5 against 15 |
| Seconds for one whole chart, median | 15 | 16 | 16 | 60 |

### Nine times the calls is not nine times the input

A call count is easy to read as a price, and here it must not be. Each document goes to exactly one call in the fan-out arm, so the source text is divided between the calls. Each call repeats the block of instructions and the short list of items the first stage already found. Rendering the prompts of all 20 charts and counting their characters, the fan-out arm sends 65,412 characters for a chart at the median and the one-call arm sends 58,190, a ratio of 1.12 per chart and 1.11 taken over the corpus at once. The instruction block is 412 characters, and repeating it together with the found-items list is the whole of the difference, 7,679 characters a chart. Leaving the found-items list out of the prompt moves the corpus ratio to 1.09, so the figure does not turn on how long an answer happens to be. On input, then, the fan-out costs about a tenth more rather than nine times more, so a deployment that wants the ordering can have the ordering without a bill nine times the size.

A character is not a token, so both arms were run again with the count the provider itself reports kept for every call. Over the same 20 charts, at 370 against 40 calls, the provider counted 1,000,841 input tokens for the fan-out arm and 882,772 for the one-call arm. That is a ratio of 1.13, against 1.11 counted in characters, so counting characters was standing in for the bill to within about two hundredths and the paragraph above holds in the unit that is charged. The build refuses to print the character ratio as a cost if those two ever drift apart by more than a tenth.

The output side is now counted too. The arms were capped at 500 and 2,000 tokens of answer per call, and a fan-out call used 20 at the mean, well inside its cap. Across the corpus the fan-out arm answered in 7,402 tokens against 1,503 on the pass reported here, and pairing each arm’s passes against the other’s puts the ratio anywhere from 3.6 to 5.1. Nine times the calls is not nine times the output either. More to the point, output is the small part of this bill: at the hosted rates this paper prices against, output is 3.6 per cent of what the fan-out audit costs. The figure that decides the price is the input, and on input the two arms are a tenth apart.

Beyond tokens the extra calls cost requests against a rate limit and time on the clock: 60 seconds for a chart against 15.

### What this does not say

It does not say the shapes have equal recall. It says no difference in recall was detected on 20 charts and 130 gold items. Failing to detect a difference is the weaker statement of the two, and the arms would have had to differ by several items before their intervals came apart. It does not say the cheap shapes cannot rank, either. The joined interval 0.400 to 0.867 and the placed interval 0.381 to 0.899 both run from below one half to well above it, and an interval containing one half leaves the ranking undemonstrated without ruling it out. It also does not compare three identical label sets. Which class a chart falls in is decided by whether the first stage missed anything on it, and that stage is a fresh sample in every arm, so the placed arm ranked 6 against 14 charts where the other two ranked 5 against 15. One chart changed sides. Two areas under a curve computed over different label sets are not two measurements of one quantity, so the small gap between the two cheap arms is no evidence that either of them ranks better than the other. Every caution in Section S applies here in full and applies to all three arms equally, including the circularity of the labels and the synthetic corpus.

### What a reader can take from it

The cost of running this library is not a toll fixed by the design. It is a dial, and Table 14 says what the setting changes. If the gate only has to release a case or send it to a person, the cheap setting reached the same recall here for 2 calls a chart instead of 18. That setting saves requests and time and leaves the input tokens where they were, for the reason counted two paragraphs above, and neither precision nor per-call latency moved much with it: of the items the arms named, 124/128 matched gold in the joined arm and 122/126 in the placed one. As rates those are 96.9% and 96.8%, and a call took 3.59 seconds at the median against 2.6 in the fan-out. The first stage is what recall mostly rests on, and it named 94.6% of the gold items in the joined arm and 93.1% in the placed one. If a person has to be handed a queue with the worst case at the top, the fan-out is what produces the order, and that is the reason to pay for it. Stating which of the two a deployment needs is a decision the person deploying makes, so the library takes the shape as an argument and does not choose it.

### A third option is to escalate

The two shapes need not be alternatives. A deployment can run the cheap audit on every chart and pay for the fan-out only on the charts where the cheap audit proposed something. `examples/cascade_cost.py` prices that arrangement by putting the two artefacts already on disk together, taking the ordering score from the fan-out arm for a chart the cheap pass flagged and leaving it at zero for a chart the cheap pass passed. On this corpus the cheap pass flagged 10 of the 20 charts, so the arrangement spends 225 calls over the corpus against 370 for the fan-out everywhere and 40 for the cheap pass alone. The area under the curve comes out 0.853, between 0.933 for the fan-out and 0.633 for the cheap pass. Two fifths of the calls go, and about a tenth of the ordering goes with them.

### Where that tenth went

All of the lost ordering traces to one chart. PatientNo108 carries a miss and the cheap pass proposed nothing on it, so it never escalated, and a chart that never escalates keeps a score of zero and sits at the bottom of the queue. This is the failure mode of every escalating design. The cheap stage decides what the expensive stage is allowed to see, and a case the cheap stage passes is a case the expensive stage never reads. A deployment that cannot accept that has to escalate on something other than the cheap pass having found a candidate, or escalate everything.

### We then ran the escalation as one pipeline

The figure above is arithmetic over two runs, so it was worth running the arrangement instead of adding it up. `examples/accuracy_report.py` takes two further settings that do this, and in both of them a single first stage feeds both audit stages. Under --shape cascade the cheap audit runs on every chart, and every document whose line one of its proposals quoted then gets an audit call of its own. Under --shape cascade_wide a chart with any proposal at all sends every document it has. The composition prices that policy. A proposal whose quote lands in no document widens the referral to the whole chart in both settings, because the cheapest stage may not decide that a finding it could not locate needs no second look. The two runs are in Table 16, beside the two arms they sit between.

### The composition had overstated both halves of the bargain

It priced the chart-wide policy at 225 calls for an area of 0.853. Run as one pipeline that policy spent 146 calls for an area of 0.627. The call count came out lower because the composition escalated 10 charts where the run’s own cheap audit escalated 6. The area came out lower for a reason that is worth separating from the price.

### A composition takes one run’s trigger and another run’s answers

The charts that escalate in it are the charts the cheap arm flagged, and the score each of those charts then receives is the score the fan-out arm gave it. Neither half is wrong where it came from. Put together they describe a pipeline in which the stage that decides what to look at is one sample of the cheap audit and the stage that does the looking answers to a different sample. A run has one cheap audit, and its proposals are both the trigger and the input to everything after it. The cheap audit is the least reliable stage in the arrangement, and an escalation built on it is as reliable as that.

### Three of the five charts carrying a miss escalated nothing at all

In the document-scoped run they were PatientNo107, PatientNo108, PatientNo111, 3/5 of the charts that carried a miss. In the chart-wide run they were PatientNo108, PatientNo117, PatientNo119, the same count from a different sample. PatientNo108 appears on both lists and on the composition’s. A chart nothing escalated on keeps a score of zero and sits at the bottom of the queue, whatever a second audit call would have found in it. Widening the referral from the quoted documents to the whole chart does not address this, because the same proposals still decide whether anything is referred. Escalating on a signal the cheap audit does not produce would address it, and that is a design this appendix has not measured.

### What this leaves for a reader deciding whether to add a loop

The cheap audit on its own ordered these charts at 0.633, 0.400 to 0.867, for 40 calls. The two escalations cost 53 and 146 calls and ordered them at 0.580, 0.333 to 0.840, and 0.627, 0.367 to 0.887. All three of those intervals cover one half and all three overlap each other, so what twenty charts support is that the escalation bought no ordering we could measure, at between a third again and nearly four times the price of the cheap audit alone. Both escalations also ended two gold items behind the two arms outside them, 123/130 against 125/130. That gap is inside the noise of this corpus and is no evidence against them. The arm that did order the charts is the one that audits every document, 0.933, 0.787 to 1.000, at 370 calls. That is the only ranking on this page its own passes support. On its weakest pass the fan-out arm ordered the charts at 0.933 against 0.780, where the second figure is the best pass any other arrangement managed and it came from the verdicts arm. Every other pair of arrangements here overlaps across its passes, and the build refuses to print a ranking between two whose passes cover each other. On this evidence a loop around a cheap first pass is not a cheaper route to that number.

### How much of this moves between passes

Each escalating arm ran 3 passes over the corpus and Table 16 reports the first of them, so the spread across passes tells a reader how much of the ordering figure belongs to the sample and how much to the arrangement. The document-scoped arm ordered the charts at 0.580 on its weakest pass and 0.661 on its strongest. The chart-wide arm ran from 0.627 to 0.733 over its own 3 passes, and the top of that range sits above the cheap audit’s 0.633. We can support only this much. The escalation did not beat the cheap audit by an amount three passes over 20 charts can tell apart from sampling. A better escalation may still exist. Recall moved less than the ordering did, 94.6% (89.2% to 97.8%) for the document-scoped arm against 94.6% (89.2% to 97.8%) for the chart-wide one. The count of blind charts did not move either, 3/5 and 3/5 of the charts carrying a miss, even though the three names differ between the two.

### The middle chart pays the cheap price and a few charts pay for the rest

A reader budgeting for one chart will not see the corpus totals of Table 16. The chart in the middle of the corpus cost 2 calls in the document-scoped arm and 2 in the chart-wide one, the same as the cheap audit alone costs, because most charts escalate nothing. The corpus holds 350 documents and the fan-out arm gives each one a call of its own. The chart-wide escalation gave a call of its own to 106 of them and the document-scoped one to 13, and all of that spending landed on the 6 and 7 charts that escalated. Two consequences follow for anyone sizing a rate limit. The average cost per chart understates what the expensive charts cost, and a corpus in which more charts raise a proposal moves the bill toward the fan-out arm’s without moving the ordering toward it.

### What the composed figure is still good for

Its call counts are exact, and it is the only place here where the two end arms are checked against each other. Everywhere else they are reported side by side. `examples/cascade_cost.py` refuses to compute when the two arms disagree on which of the 20 charts carry a miss, so a later run that disagrees fails outright and returns nothing. Its ordering figure is the part the runs above contradict, and the interval it carried already said as much: 0.580 to 1.000 reaches past 0.933 at one end and past 0.633 at the other. We are keeping the composed row in place with the measured rows beside it, because a projection that was tested and came out wrong is more use to a reader than a projection quietly withdrawn.

### The third arm cannot be put in the cheap slot at all

The placed arm costs what the merged arm costs, so a reader may reasonably ask why the cascade above was not built on it instead. It cannot be. Each arm ran its own first stage, and the placed arm’s first stage missed PatientNo115 while the fan-out arm’s first stage named it. Two runs whose first stages disagree are not auditing the same answer, and an escalation policy composed across them would be answering a question about neither. The script refuses that pair. The same arm also refers less: on the trigger used above it would send 8 of the 20 charts upward against 10, and three of the charts its own first stage got wrong are charts it would never refer. The cheap slot of a cascade goes to the shape that puts forward a candidate on a chart worth a second look. That is not always the shape that costs least, and on this corpus those two properties came apart.

## X. The redaction step against a published threshold

The pipeline has an optional first step that removes direct identifiers from a document before any other stage reads it. Section S measures the stages after it and says nothing about this one. Until the run reported here, the library shipped that step with no recall number attached to it.

De-identification is the one step of this pipeline whose target we did not have to invent. An evaluation of five public de-identification tools, over 2503 annotated radiology reports from a multi-hospital system, quotes the level an earlier comparative study set as acceptable before a clinical release: recall of 95% and precision of 80% for patient names and social security numbers, and recall of 85% and precision of 70% for every other class of protected health information (Steinkamp et al., 2020). The bar belongs to that earlier study, and we use it because it is the number this field quotes. It has two halves, and this section reports both.

The annotated corpora this task is normally measured on are released only under a data use agreement, and we hold no such agreement. The arm under test also sends each note to a hosted model, which our own data rules forbid for a real note, so the corpus had to be one we were free to send. We generated it. The generator writes each note one fragment at a time and records the offset of an identifier at the moment it writes it. That gives a generated corpus a property no annotated corpus has: the answer key cannot disagree with the text. It then asserts that every recorded span still holds its own characters before the note is handed on. A checksum over the text and the spans together pins the corpus, so a silent redraw fails the build and cannot move a rate.

Two corpora came out of that generator, and they are not a matched pair. The first writes every identifier the way a form writes it, with a full name, a spelled month, a bracketed area code and a hyphenated social security number. It holds 396 identifiers over 60 notes, instantiates 16 of the eighteen Safe Harbor classes, and carries checksum c83e935b9618. The second writes identifiers the way a hurried clinician does. A name arrives as a capitalised surname-first pair, in lower case, as three bare initials, and with a letter doubled or an m typed as rn. Two dates in three carry a day and a spelled month and no year at all, and the rest carry a two-digit year. A third of the geographic identifiers are five-digit postcodes and the rest are streets and cities. Social security numbers arrive space-separated, and an age above 89 is spelled out in words. One patient in five has a surname that is also the name of a disease, so the note says both Mr Cushing and Cushing’s syndrome. It holds 300 identifiers over 60 notes, instantiates 13 classes, and carries checksum 98e6071244ec.

The two class lists are not the same list, and that matters for any comparison between the columns below. The second corpus’s 13 classes are a subset of the first’s 16, and the build refuses to typeset this section if that stops being true. Electronic mail addresses, fax numbers and account numbers appear in the first corpus only, and each of those three has a fixed shape a pattern states in one line, so the missing classes account for part of any fall between the two pattern columns. The pattern arm restricted to the 13 classes both corpora hold is reported below beside the unrestricted figure. The three-initial names are the harshest class in either corpus, and deliberately so. Some of the triples the generator produces are also standard clinical abbreviations when read on their own, so this is a class a human annotator could argue about as well.

Both corpora also carry text that must survive. The first holds 138 clinical phrases that are not identifiers, such as a potassium value or a stent placed in a named year, and 12 of its notes carry no identifier at all. The second holds 156 such phrases, drawn from a different list and chosen to look like identifiers on purpose: a bed number, a ward number, a murmur graded three of six, and an age of 89, which is the highest age the rule leaves alone. 12 of its notes carry no identifier. The two kept rows below are therefore measured over different phrases as well as over different counts of them.

Three arms ran against each corpus. The first is a set of regular expressions with no provider call in it at all. A developer reaches for that before reaching for a model. The second is the shipped path and nothing beside it. The shipped agent answers the task the shipped redactor builds, the reply is held to its declared shape by the same parser every other stage uses, and the removals are applied by the shipped rewriter, the function that takes each quoted span and produces the released text. The third feeds the pattern spans and the model spans to that same rewriter together. Every arm that makes a call was run against Claude Sonnet 4.5, one call per note.

Each arm answers four questions, because a redactor fails four different ways. Located asks whether any prediction overlapped the identifier at all. That is the generous convention. The published bar was measured token by token, so a prediction that catches one word of a two-word name counts here and would not count there, and every comparison below should be read with that in our favour. Removed asks whether the released text still carries the identifier anywhere. It is the only question a privacy officer asks. A withheld document releases nothing, so every identifier in one counts as not removed, and the run records the same rate over released documents alone so that both denominators are on the page. Kept asks how many protected clinical phrases survived, counted over released documents, because a document that was never released destroyed nothing in it. Untouched asks how many identifier-free notes the arm left alone. An arm can reach perfect recall by deleting the document, and the last two questions are what stop that from reading as a success. The run records a fifth figure that no release depends on, whether the class the arm attached to a quote was the right class, and it is reported once below.

The scoring does not reuse the library’s own locating code. It re-implements the search for a quote inside a document, because a scorer that shares a bug with the thing it scores reports that bug as a result. The generator is `examples/deid_corpus.py`, the scorer is `examples/deid_report.py`, and every number below is read at build time out of `data/deid_report.json`, `data/deid_report_hard.json` and one third file described further down, and no number here is typed by hand. Table 17 gives every arm on every question.

**Table 17.** Patterns fall from more than half the identifiers to fewer than one in twelve when identifiers are written irregularly, and the shipped redactor does not. Its destruction moves instead. The two corpora do not hold the same set of classes, and the text gives the pattern comparison restricted to the classes they share. Both hold 60 notes and were scored by the same model, named above the table, over 3 passes of the first and 3 of the second. Every count row is the first of those passes, which is what one caller gets, and the last two rows give the weakest and the strongest pass on the located row so that a single pass cannot pass for a stable one. Every interval is two-sided Clopper-Pearson at *δ* = 0.05, and the fraction is printed beside the rate because the denominators differ by class. Located counts an identifier the arm quoted, removed counts one with no occurrence left anywhere in the released text, and kept counts a protected clinical phrase still present, over released documents only, since a withheld document destroys nothing. A clean note holds no identifier at all, and left untouched counts one the arm made no prediction against. A prediction is spurious when the span it quotes overlaps no identifier, and precise when it does overlap one, which is the other half of the published bar. The seconds row is the median time one note spent waiting on the provider. The pattern arm reads none in the last two rows because it makes no provider call to wait on and returns the same spans on every pass.

|  | As a form writes it |  | As a clinician writes it |  |
| --- | --- | --- | --- | --- |
|  | Patterns | Shipped | Patterns | Shipped |
| Identifiers located | 228 of 396<br>57.6%<br>52.5% to 62.5% | 395 of 396<br>99.7%<br>98.6% to 100.0% | 24 of 300<br>8.0%<br>5.2% to 11.7% | 297 of 300<br>99.0%<br>97.1% to 99.8% |
| Identifiers removed | 228 of 396<br>52.5% to 62.5% | 395 of 396<br>98.6% to 100.0% | 24 of 300<br>5.2% to 11.7% | 291 of 300<br>94.4% to 98.6% |
| Protected phrases kept | 138 of 138<br>97.4% to 100.0% | 137 of 138<br>96.0% to 100.0% | 156 of 156<br>97.7% to 100.0% | 131 of 154<br>78.4% to 90.3% |
| Clean notes left untouched | 12 of 12 | 11 of 12 | 12 of 12 | 5 of 12 |
| Spurious predictions | 0 of 228 | 1 of 428 | 0 of 24 | 23 of 326 |
| Precise predictions | 228 of 228<br>98.4% to 100.0% | 427 of 428<br>98.7% to 100.0% | 24 of 24<br>85.8% to 100.0% | 303 of 326<br>89.6% to 95.5% |
| Documents withheld | 0 | 0 | 0 | 1 |
| Provider calls | 0 | 60 | 0 | 60 |
| Seconds per note, median | none | 4.0 | none | 4.0 |
| Located, weakest pass | none | 395 of 396 | none | 297 of 300 |
| Located, strongest pass | none | 396 of 396 | none | 298 of 300 |

### The same command run twice does not give the same numbers

Before any comparison in this section can be read, one fact about all of them has to be on the page. Every model figure here is one pass over the corpus, the run pays for 3 passes so that the size of a pass can be seen, and on the irregular corpus those passes are not close to each other. Identifiers removed ran from 284 of 300 to 297 of 300. Protected clinical phrases kept ran from 128 of 152 to 133 of 156. Diagnoses named after a person that the released text no longer held ran from 9 to 11, and documents the pipeline refused to release ran from 0 to 2. Nothing changed between those passes. The corpus was the same file under the same checksum, the instruction text was the same text, and the model was the same model at the same setting. It answered differently anyway.

That has a consequence we hold ourselves to for the rest of the section. A difference between two wordings that is smaller than the difference between two passes of one wording is not a difference. So every comparison below is checked by the generator against the passes: the weakest pass of the wording we call better has to beat the strongest pass of the wording we call worse, and where it does not, the build stops and the claim goes unprinted. Some of the comparisons we would like to make do not survive that check, and where that happens we say so and leave the claim unmade. We do not quote the two headline numbers and let the reader assume a gap.

The patterns reach what a pattern can state and nothing else. On the regular corpus they locate 57.6% of the identifiers. Write identifiers irregularly and they locate 8.0%. Some of that fall is the three classes the second corpus does not contain, so the same comparison over the 13 shared classes is 180 of 348, a rate of 51.7% with an interval of 46.3% to 57.1%, against 24 of 300, a rate of 8.0% with an interval of 5.2% to 11.7%. The fall survives the correction. A space in place of a hyphen takes the social security number from 12 of 12 to 0 of 12, and writing the date as a day with a spelled month, or as three numbers with full stops between them, takes the date class from 48 of 48 to 0 of 36. The patterns destroy nothing. Every prediction they made overlapped a real identifier, at 228 of 228 and 24 of 24, because a pattern that never fires never over-reaches, and that is the whole of their appeal. Neither arm destroyed a diagnosis named after a person on the regular corpus, at 0 and 0, because that corpus holds no surname that is also a disease name, and the patterns destroyed none on the irregular corpus either, at 0.

The shipped redactor holds across the change, at 99.7% and 99.0% located. Its precision falls and stays above the published bar. 427 of 428 of its predictions overlapped a real identifier on the regular corpus, a rate of 99.8% with an interval of 98.7% to 100.0%, and 303 of 326 on the irregular one, a rate of 92.9% with an interval of 89.6% to 95.5%. Both are above the 80% the threshold asks for on names and social security numbers and the 70% it asks for elsewhere, and both are pooled across classes while the published bar is stated per class. The wording costs the survival of clinical text, and precision against identifiers holds. The redactor keeps 137 of 138 protected phrases on the regular corpus and 131 of 154 on the irregular one, and it leaves 5 of 12 identifier-free notes untouched against 11 of 12. A reader deciding whether to switch this step on is trading one kind of loss for the other, and the rows are printed together for that reason. The class the redactor attached was right for 383 of 396 of the regular corpus, an interval of 94.5% to 98.2%, and 285 of 300 of the irregular one, an interval of 91.9% to 97.2%. The run also records which label was used in place of the right one, and on both corpora the answer is one and the same. Every age above 89 was filed under the date element, 12 of 12 on the regular corpus and 12 of 12 on the irregular one. Beyond those, the number of labels the shipped path got wrong for any other reason was 0 on the regular corpus and 0 on the irregular one.

That single disagreement is one we lose. 45 CFR 164.514(b)(2)(i)(C) files all ages over 89 with the date element, so the label the model chose is the label the rule gives, and the label our corpus calls correct is the one with no rule behind it. Counting the date element as right for such an age, the class was right for 395 of 396 identifiers of the regular corpus, an interval of 98.6% to 100.0%, and 297 of 300 of the irregular one, an interval of 97.1% to 99.8%. We leave the corpus as it stands, because relabelling would move the corpus checksum and would bury a disagreement worth seeing. A wrong label changes nothing about the released text, because a removal does not consult the label. It changes what an auditor reading the log is told, and that is why the run records it.

### Against the published threshold, class by class

An aggregate recall figure for a de-identifier is dominated by whichever class is largest, so Table 18 gives the six largest classes a row and an interval each. Those six cover 252 of 396 identifiers of the regular corpus and 192 of 300 of the irregular one, and they include both classes the threshold names on their own. The classes the table leaves out are the ones with a machine shape, from a telephone number to a device identifier, and the shipped path located every identifier in every one of them in both corpora. The threshold is a lower bound on recall, and the intervals here are two-sided. That makes the comparison conservative by half of *δ* on each side.

**Table 18.** The shipped redactor’s point estimate clears the published bar in each of these six classes in both corpora. The lower end of its interval clears it in four of the six of the regular corpus and three of the six of the irregular one. The bar is 95% recall for names and social security numbers and 85% for the other classes (Steinkamp et al., 2020). Where the lower end falls short the reason differs by row, and the difference matters. A row of twelve identifiers cannot reach either bound even with nothing missed at all, because a clean sweep of twelve supports only the interval printed in the social security row, and the social security and age rows hold twelve each. The name row of the irregular corpus is not like that. With nothing missed it would carry 95.0% to 100.0%, and that clears the bar, so its three misses are what defeat that row. The pattern column is the floor the same class reaches with no provider call. The age row counts ages above 89, which are the only ages the rule names.

|  | Written as a form writes it |  | Written as a clinician does |  |
| --- | --- | --- | --- | --- |
|  | Patterns | Shipped | Patterns | Shipped |
| Names | 0 of 96 | 96 of 96<br>96.2% to 100.0% | 0 of 72 | 69 of 72<br>88.3% to 99.1% |
| Social security numbers | 12 of 12 | 12 of 12<br>73.5% to 100.0% | 0 of 12 | 12 of 12<br>73.5% to 100.0% |
| Dates | 48 of 48 | 48 of 48<br>92.6% to 100.0% | 0 of 36 | 36 of 36<br>90.3% to 100.0% |
| Record numbers | 24 of 24 | 24 of 24<br>85.8% to 100.0% | 0 of 24 | 24 of 24<br>85.8% to 100.0% |
| Geographic subdivisions | 0 of 60 | 59 of 60<br>91.1% to 100.0% | 0 of 36 | 36 of 36<br>90.3% to 100.0% |
| Ages the rule counts | 0 of 12 | 12 of 12<br>73.5% to 100.0% | 0 of 12 | 12 of 12<br>73.5% to 100.0% |

The name row is the one to read first, because it carries the strictest bar and the most identifiers. On the regular corpus the shipped redactor located every name, a rate of 100.0%. On the irregular corpus it located 69 of 72 names, a rate of 95.8%, with an interval of 88.3% to 99.1%. The point estimate is above the published 95%. The lower end of the interval is not, and the reason is the three misses, because a row that size with nothing missed would clear the bar. So we print the row and we do not claim the bar is cleared for names written irregularly. The three misses are named in the artefact and they are of two kinds. Two are sets of three bare initials, which carry no other cue that they are a person. The third is a surname with a doubled letter, and it produced something more interesting than a miss. The initials that do spell a clinical abbreviation were all located, so the redactor was not reading the triples as abbreviations and letting them through.

The geography row covers two things the rule treats differently, so the run reports them apart. 45 CFR 164.514(b)(2)(i)(B) names geographic subdivisions smaller than a state and does not name the hospital a patient was treated in, while the annotation guidelines the field works to do count that hospital. On the regular corpus the shipped path located 24 of 24 of the streets and cities, an interval of 85.8% to 100.0%, and 35 of 36 of the hospital names, an interval of 85.5% to 99.9%, against 0 of 24 and 0 of 36 for the patterns. A reader who reads the rule the narrow way needs the first number without the second folded into it. The irregular corpus names no hospital, so it has no institution row at all. Its geographic identifiers are streets, cities and five-digit postcodes, and its shipped figure of 36 of 36, an interval of 90.3% to 100.0%, is comparable to the regular corpus’s first number rather than to its class total. The pattern arm finds 0 of 36 of them, for two separate reasons. A street, a city and a hospital name have no shape a pattern can state without a list of names to check against. A postcode does have one, and the pattern set we scored carries no expression for it. That is a gap in our pattern set, and it is no limit of patterns themselves. We report the arm as we ran it, and we did not improve it after seeing where it failed.

### The measurement’s first finding was a defect in our own prompt

The first run of the regular corpus located 372 of 396 identifiers, a rate of 93.9% with an interval of 91.1% to 96.1%. The class breakdown put the shortfall in three places. Ages above 89 scored 0 of 12, and the reason was in our own instruction text, which listed the identifier classes without naming that one and told the model not to report a bare year. The rule names both. Names scored 92 of 96, and the four misses were all a bare surname in a note that also gave the full name, because a quote is located at every occurrence of itself but a full name is not an occurrence of the surname alone. Geographic identifiers scored 52 of 60, and all eight misses were a hospital name, or a city name written inside one. Two sentences were added to the prompt, one naming the age and one asking for every surface form of an identifier that appears more than once. Nothing about hospitals was written into it. The three classes then scored 12 of 12, 96 of 96 and 59 of 60, and the run as a whole went to 395 of 396. The geography gain is a side effect of asking for every surface form, and no fix was aimed at that class. The prior artefact is kept in the repository and the build reads it, so the before-number comes from a file, and the build refuses to typeset this paragraph if the two runs disagree on the model, on *δ*, or on the corpus checksum.

One thing about that fix has to be said in the open. The prompt was changed because this corpus showed it was wrong, and then the same corpus scored the new prompt, so the regular corpus’s figures above are in-sample. The irregular corpus was never used to change the prompt or anything else in the redactor, so it is the out-of-sample half of this section, and it is the half that holds the misses. Nothing was adjusted after either of the two runs reported here.

That fix cost precision, and the two artefacts hold both sides of the trade. Under the earlier prompt the shipped path kept 138 of 138 protected phrases, an interval of 97.4% to 100.0%, left 12 of 12 identifier-free notes untouched, an interval of 73.5% to 100.0%, and made 0 of 398 predictions that overlapped no identifier, an interval of 0.0% to 0.9%. Under the current one the same three figures are 137 of 138, 11 of 12 and 1 of 428. Each is a move of a single item and each pair of intervals overlaps, so what the two artefacts establish is the direction of the trade and not its size. Naming the year as something that can carry an age is what makes the redactor flag the year inside a stent placement, and that one flag is the whole of the fall. We report it and do not tune it away, because the corpus we would tune against is our own.

### The quoting rule owns this failure

A quote is removed at every place it occurs, which is what stops a partial match from leaving half a name behind. When a surname is also a disease name, that rule has no good outcome. The corpus writes the case as bluntly as it can, by giving the patient the surname: the note says Mr Cushing was treated for Cushing’s syndrome. Removing Cushing removes it from the diagnosis as well, and scoping the quote to Mr Cushing leaves the surname standing inside the diagnosis. Twelve of the irregular corpus’s 60 notes carry one such diagnosis, and the run shows the redactor going both ways. 11 of the twelve diagnoses were destroyed. In the other two the call quoted the honorific with the surname, so the diagnosis survived and the surname’s own characters survived inside it, which the scorer counts as an identifier not removed. Those two notes are why located exceeds removed on this corpus, at 297 of 300 against 291 of 300, together with a larger cause that has nothing to do with eponyms. One document was withheld, described next, and a document that releases nothing has every identifier in it counted as not removed. Over released documents alone the shipped path removed 291 of 294, an interval of 97.0% to 99.8%, so 3 of 294 identifiers were still in the text it released. That is the figure to hold beside the located row.

The run does not stop at that count. A count of identifiers still present says nothing about why they are there, and the two reasons ask for different repairs, so the run records both. The first is whether any prediction covered the identifier at all. The second is whether its characters survive inside a protected phrase that also survived. That is the eponym case above: the phrase was meant to stay, the surname inside it was meant to go, and one string cannot do both. Of the identifiers the shipped path left standing, 2 were covered by no prediction at all and 1 survive inside a protected phrase that survived, that one being a name. 0 are left over. The three groups cannot overlap, because an identifier nobody predicted is counted as never found whether or not a kept phrase also holds it, and the generator stops the build if they do not add up to the whole.

### Pricing the two ways to give up

A design that removes an identifier by quoting it cannot both delete the surname and keep the diagnosis built from the same characters. Which of the two to give up is a choice, and we ran the second choice to find out. One sentence was added to the instruction text, asking the model not to quote a surname on its own when that surname also sits inside the name of a disease, and to quote the longer form that names the person instead. Nothing else changed. The same 60 notes under the same checksum, the same model, 60 calls again, and 3 passes as before. The artefact records the added sentence beside the numbers it produced, so a reader can see the whole of what was different.

The sentence did what it was written to do, and the check is made pass by pass. Diagnoses named after a person that the released text no longer held ran from 9 to 11 of the twelve under the shipped wording, and from 0 to 1 under the scoped one. Those two ranges do not touch, so this is a real difference and not a lucky pass. The protected clinical phrases the released text no longer held fell from 23 to 16, and the phrases still standing rose from 131 of 154 to 140 of 156, a gap that also holds over every pass of both runs, from 133 of 156 at the shipped wording’s best against 139 of 156 at the scoped wording’s worst. Predictions overlapping no identifier fell from 23 of 326 to 15 of 313, and that gap holds over every pass as well.

Two figures we could have quoted here do not survive the pass check, and we leave them unclaimed. Identifier-free notes left completely untouched were 5 of 12 under the shipped wording and 4 of 12 under the scoped one, but the passes run from 4 of 12 to 5 of 12 against 3 of 12 to 5 of 12. Those ranges overlap, so we do not say the sentence cost anything there. Documents the pipeline refused to release were 0 in the pass reported and ran from 0 to 1 over the three, against 0 to 2 for the shipped wording. Refusal is not a property of the wording. It happens when the model quotes a span it cannot then point at in the document, and whether a given note trips that is one of the things the model decides differently from one pass to the next.

The price is one figure, and it is the figure a privacy officer reads. The scoped wording removed 285 of 300, an interval of 91.9% to 97.2%, over passes running from 278 of 300 to 285 of 300. The shipped wording removed 291 of 300 over passes running from 284 of 300 to 297 of 300. Those two ranges overlap, so this run cannot tell the two wordings apart on removal, and we do not claim the scoped one recalls less. The run can tell apart the released text itself. 15 of 300 identifiers were still present in what the scoped wording released, against 3 of 294 for the shipped wording, and the two counts are read against different totals because a withheld document has no released text to search.

Every one of those survivors is accounted for by the same two facts recorded for the shipped path above. 12 of them survive inside a protected phrase that survived too. This is the eponym case doing exactly what the sentence asked for, and it is no failure of the model. 3 were covered by no prediction at all, a failure of the same kind the shipped path makes. 0 are left over. The surviving group is a name in every case, which is what the sentence is about.

A team choosing between the two wordings is choosing which kind of error it would rather explain. The shipped wording destroys diagnoses and leaves fewer identifier characters behind. The scoped wording keeps the diagnoses and leaves the surname readable inside them. Both are now priced over the same notes. The library ships the first. The second is one sentence, and the command that produced these numbers is the recipe for it.

We also print the name class for each wording, at 69 of 72 located under the scoped wording against 69 of 72 under the shipped one. The run records figures by class for the pass it reports only, so those two numbers have no range behind them and we draw no comparison from the difference between them.

### Reading the wrong predictions instead of only counting them

A rate of wrong predictions tells nobody what to change. So the run records every prediction that overlapped no identifier together with the span it quoted, and those lists are short enough to print. Under the shipped wording the 23 of 326 wrong predictions quoted 10 distinct spans, namely “89 years old” 4 times, “2019” 3 times, “Cushing” 3 times, “bed 12” 3 times, “Crohn” twice, “Graves” twice, “Parkinson” twice, “ward 7” twice, “Bell” once and “Paget” once. Under the scoped wording the 15 of 313 quoted 6 distinct spans, namely “89 years old” 4 times, “ward 7” 4 times, “2019” 3 times, “bed 12” twice, “12” once and “ward 7, bed 12” once. The surnames are gone from the second list, and that is the added sentence working. The leftovers fall into kinds: a bare year, an age written at the boundary of the rule, and a number saying which place inside a hospital the patient occupies. None of those kinds is a near miss on an identifier.

### A third wording, made of what the rule itself leaves alone

All three of those are things 45 CFR 164.514(b)(2)(i) does not ask anyone to remove. Paragraph (C) removes every element of a date except the year, and asks for an age only when that age is above 89, so a bare year and an age of 89 or below are both left in the record by the rule as written. A ward, bay, room or bed number is our own reading, and the rule’s words do not state it: paragraph (R) catches any other number that uniquely identifies the individual, and a bed is reused between patients, so the number names the place only. We mark that third one as a reading because it is one, and a privacy officer who disagrees can drop it and rerun the command.

The third wording states those 3 exclusions and changes nothing else. It is the scoped sentence with the exclusions added after it, and the generator checks that the third wording’s instruction text begins with the second wording’s character for character, so the three wordings differ in what they say and in nothing else. The same 60 notes under the same checksum, the same model, 60 calls, and 3 passes. Table 19 puts the three side by side.

**Table 19.**
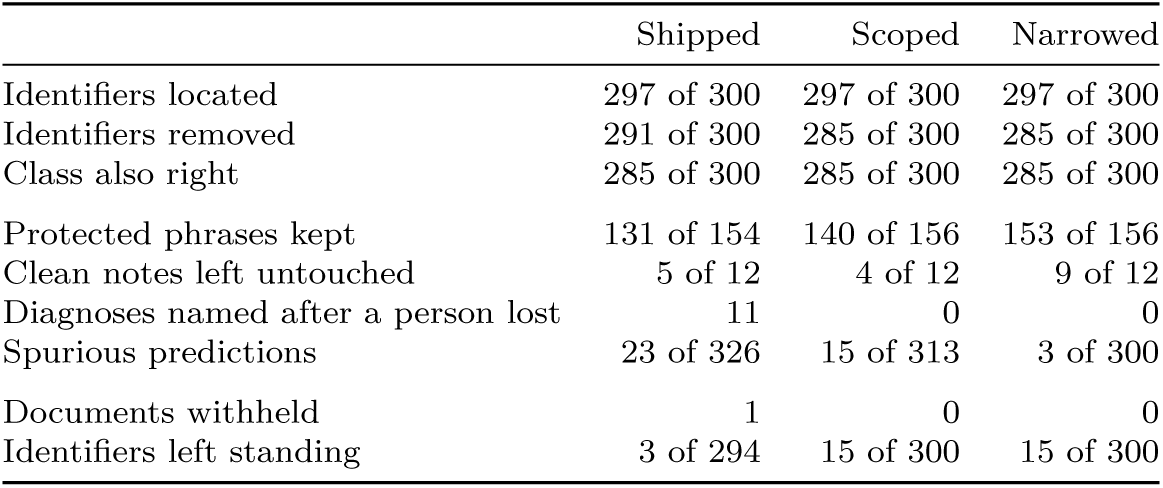
Three wordings of one instruction, run over the same notes under the same checksum, and the last one keeps the most clinical text while quoting the fewest spans that hold no identifier. Shipped is the wording the library installs. Scoped adds one sentence protecting a surname that is also part of a disease name. Narrowed adds to that the 3 exclusions the regulation itself grants, described in the text. Every cell is the first of 3 passes, and that is what one caller gets. The passes disagree with each other by more than some of the columns disagree with each other, so the text gives the range for each comparison and states which comparisons survive it. Identifiers left standing is counted over released documents alone and its total therefore moves with the row above it, since a document that was never released has no released text to search.

Three comparisons against the shipped wording survive the pass check, and a fourth survives against both wordings below. Protected clinical phrases kept rose to 153 of 156, and the weakest pass of the third wording, 149 of 152, still beats the strongest pass of the first, 133 of 156, and of the second, 140 of 156. Identifier-free notes left completely untouched rose to 9 of 12, from a weakest pass of 9 of 12 against strongest passes of 5 of 12 and 5 of 12. Wrong predictions fell to 3 of 300, from a worst pass of 3 of 300 against best passes of 22 of 324 and 15 of 313. Diagnoses named after a person survived, at 0 lost against 11 for the shipped wording, and that comparison is made against the shipped wording only, because the sentence that saves those names is already in the second wording and between the second and the third there is nothing left to win.

The exclusions did not cost recall, and the honest form of that sentence is that this run cannot show a difference either way. Identifiers located ran from 296 of 300 to 299 of 300 against 297 of 300 to 298 of 300 for the shipped wording, and identifiers removed from 275 of 300 to 287 of 300 against 284 of 300 to 297 of 300. Both pairs of ranges overlap. The generator holds that too, in the other direction: if a wording ever did pull clear of another on recall the build would stop, because the sentence saying the two cannot be told apart would have gone stale.

Underneath that equal total, the misses moved. Splitting the same identifiers by class shows the third wording finding fewer device serial numbers, from 12 of 12 to 10 of 12 and more names, from 69 of 72 to 71 of 72 than the first, with the total identical on both sides, so a reader would be wrong to take the tie above as nothing having changed. The direction of the first of those is the one to worry about. A serial number on an infusion pump is a device identifier, the rule asks for it to go, and our exclusions say nothing about equipment, so the most likely reading is that a sentence telling the model to leave the number of a bed also taught it to leave the number of a pump. We cannot prove that from one run. The class figures are recorded for the pass the artefact reports only, so this shift has no range behind it and none of the checks above applies to it. We report it as an observation and a warning, and we do not count it as a cost. It does establish that a sentence written to protect clinical prose can reach a class it never mentions, and that anyone adopting an exclusion of ours should score the classes as well as the total.

The identifiers the third wording leaves standing in what it released, 15 of 300, split the same way the second wording’s do and with nothing left over. 12 survive inside a protected phrase that survived too, every one of them a name, the eponym case behaving as the second sentence asks. 3 were covered by no prediction at all, and 0 are left over. The pump numbers of the paragraph above sit in that middle group. That is where the shift between classes and this row meet: the same two identifiers counted once as a class the wording found less of and once as characters still readable in a released document.

We report the over-quoting that is left. The third wording still made 3 of 300 wrong predictions, and every one of them quoted the same thing, “89 years old” 3 times. The rule asks for an age only above 89, the exclusion we wrote says an age of 89 or below stays, and 89 itself is the boundary of both. The model removes it anyway. That is caution at a boundary, and it is no misreading of the rule. It is the sort of thing a real deployment would settle with its own privacy officer, and we leave it in the number. Tuning the wording until one corpus of 60 notes shows nothing would produce a wording fitted to that corpus and would tell a reader less than this does.

### Failing closed, once

One document of the irregular corpus was withheld outright. The rewriter refuses when a quoted span cannot be found in the document character for character, and that is the only condition under which it refuses. It keeps the quote out of the error it raises, so that an identifier the model may have invented does not travel into a log. The artefact therefore records that this happened and leaves out which quote did it. It records one thing about that note. Its doubled-letter surname was covered by no prediction at all. The instruction text warns the model that an unfindable quote costs the whole document, and the model was held to it. The pipeline released nothing for that note. An offset-based redactor given the same reply would have removed whatever sat at the offsets and returned a document that looked clean. This is the behaviour the rest of the paper argues for, observed on a live call, and it costs one document of 60.

The other way this step can fail is that the provider does not answer at all, and that path is covered by tests alone. When the redaction call fails, the task is escalated with the reason recorded, the stored trace holds a placeholder in place of the prompt so that an un-redacted note is never written down, and no later stage runs against the text. Five tests in tests/test_stage_availability.py hold that behaviour in place, including one that asserts the prompt the redactor could not de-identify is not what gets stored.

### Adding the patterns to the model buys nothing

The third arm applies both sets of spans together. On both corpora it returns the shipped path’s own figures, digit for digit, on every question in Table 17 and on the withheld notes, the destroyed diagnoses and the call count as well. We state that and do not print it twice, because a reader asked to compare sixteen pairs of identical numbers learns the same one fact more slowly. The generator holds us to it: it compares the two arms field by field and fails the build if any of them parts company, so the sentence cannot quietly go stale. The denominator the spurious count is measured over does move: 1 of 656 against 1 of 428 on the regular corpus and 23 of 350 against 23 of 326 on the irregular one, the same number of wrong predictions inside a larger set of predictions. The patterns are precise and, once a call is being made, redundant. They earn their place only where a call cannot be afforded, and Table 17 says what that saving costs.

### What these numbers are not

Every recall and precision rate here is a ceiling. The corpus is generated, so its identifiers are more regular than a real note’s even in the irregular variant, and the annotated corpora that would settle the question are the ones we cannot use. The counts of destroyed clinical text do not run the same way, and we cannot sign their direction. Our corpus plants a disease-surname in one note in five, far denser than a real chart, and it carries far less clinical prose per note than a real one. That leaves fewer chances to remove something that should have stayed. Those counts show that the failure happens and how it happens. They are not an estimate of how often it would. The redactor is also zero-shot, meaning its instruction text carries no worked examples. On real notes, a prompted general-purpose model evaluated zero-shot on oncology notes reached recall of 0.8318 (Altalla’ et al., 2025), and a local task-specific model reached 97.9 F1 on the same radiology corpus that carries the threshold (Chambon et al., 2023). The honest reading of Table 17 is that our corpus is easier than theirs, not that our redactor is better than theirs. The run establishes something narrower. The shipped path has a number attached to it where it had none. The free alternative has one beside it, measured the same way and over the same notes. Each arm’s destruction is printed next to what it removes. And the refusal this design was built around was seen to fire on a live call.

Three things follow for the next run. Adding worked examples to the instruction text costs no extra provider call, so that is the cheapest untried change to this step. The citations above point at a second direction that is not cheaper but is different in kind, a small task-specific model run on the machine that holds the note, which removes the hosted call this arm depends on and adds a model to host in its place. And the three arms and the four questions are written against a corpus object, so the corpus is the only piece that has to change. The day a data use agreement for an annotated corpus is in place, the same arms and the same questions score it, and what we report stops being a ceiling.

## Y. Provenance of every generated quantity

Table 20 lists every quantity this paper states about our own system next to the command that produced it. A row is the whole account of where one number came from: the first column is the name the quantity carries in the generated file the manuscript reads, which is how a reader finds it in the source archive, the third column is the command or the recipe that emitted it, run from the library root, and the second column is what that emitted. Most rows are a command. The rest name the thing counted, because a count of files or of public symbols has no command shorter than its own description. Every row ends with what the quantity is in words, so the name in the first column never has to be decoded. The rows are in the order the generator produces them. That puts every quantity from the reviewer simulation first and the quantities describing the library and its test suite last. A quantity with no row here cannot reach the page at all, and that is why the table is printed.

**Table 20.**
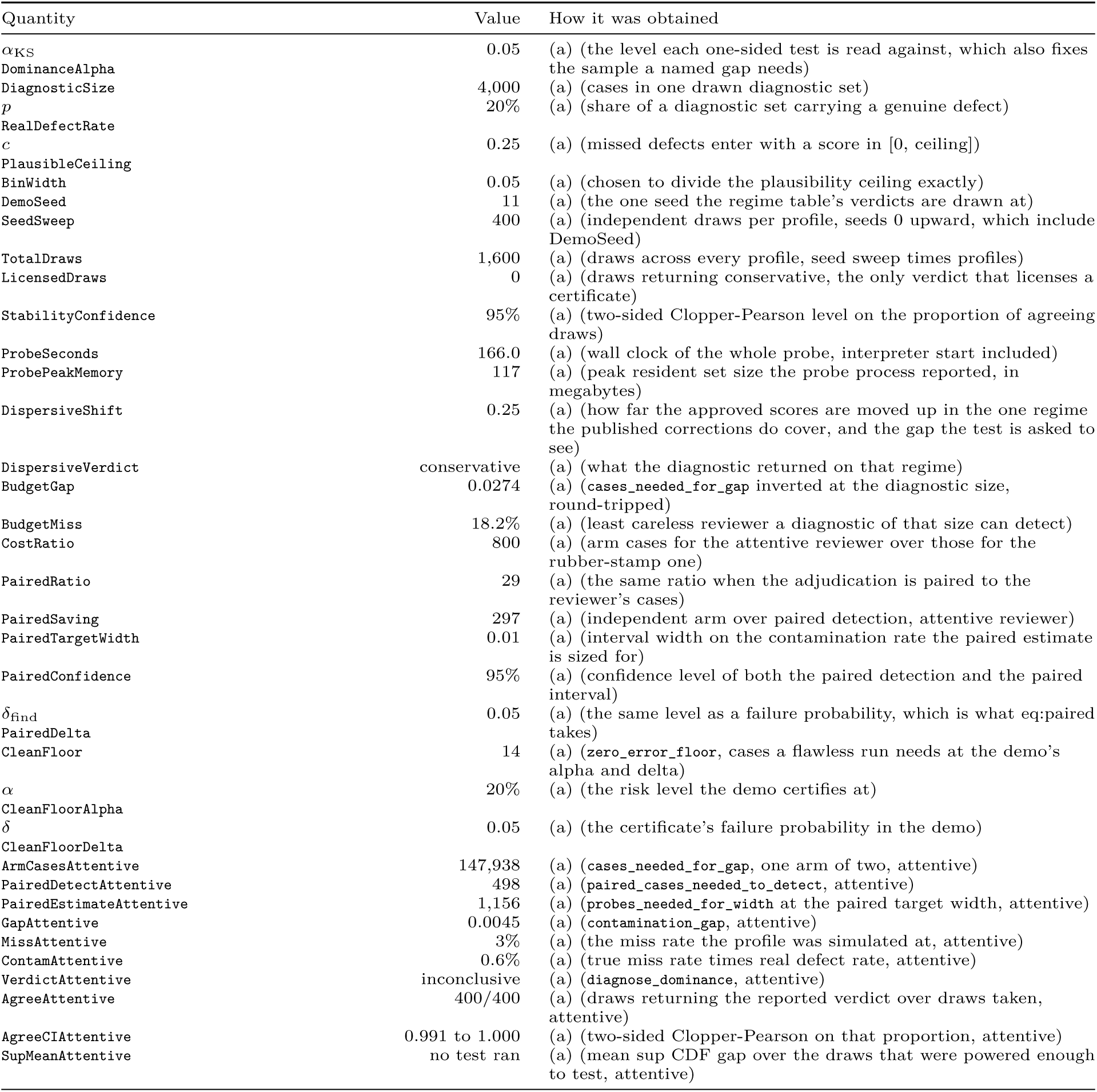
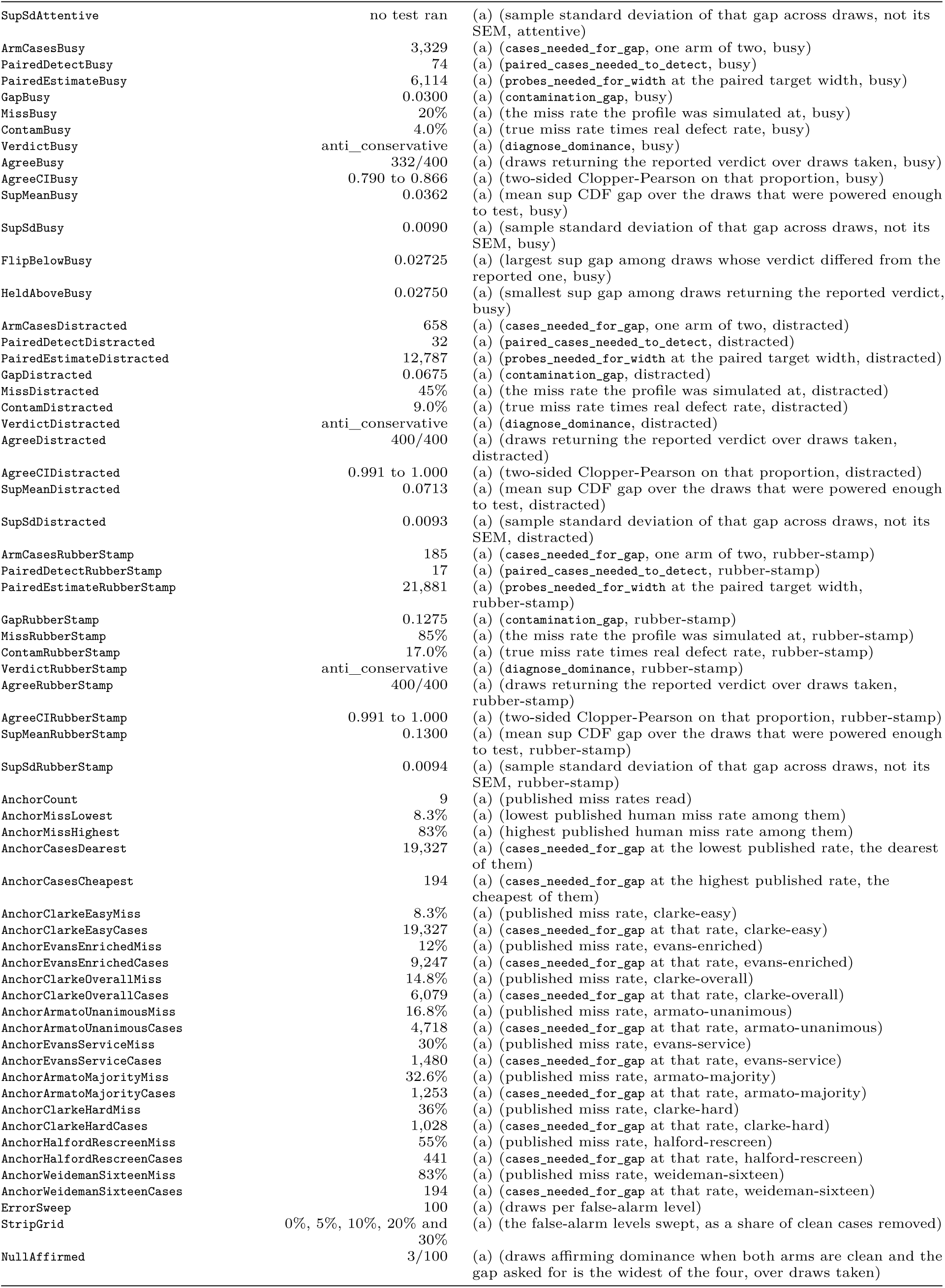
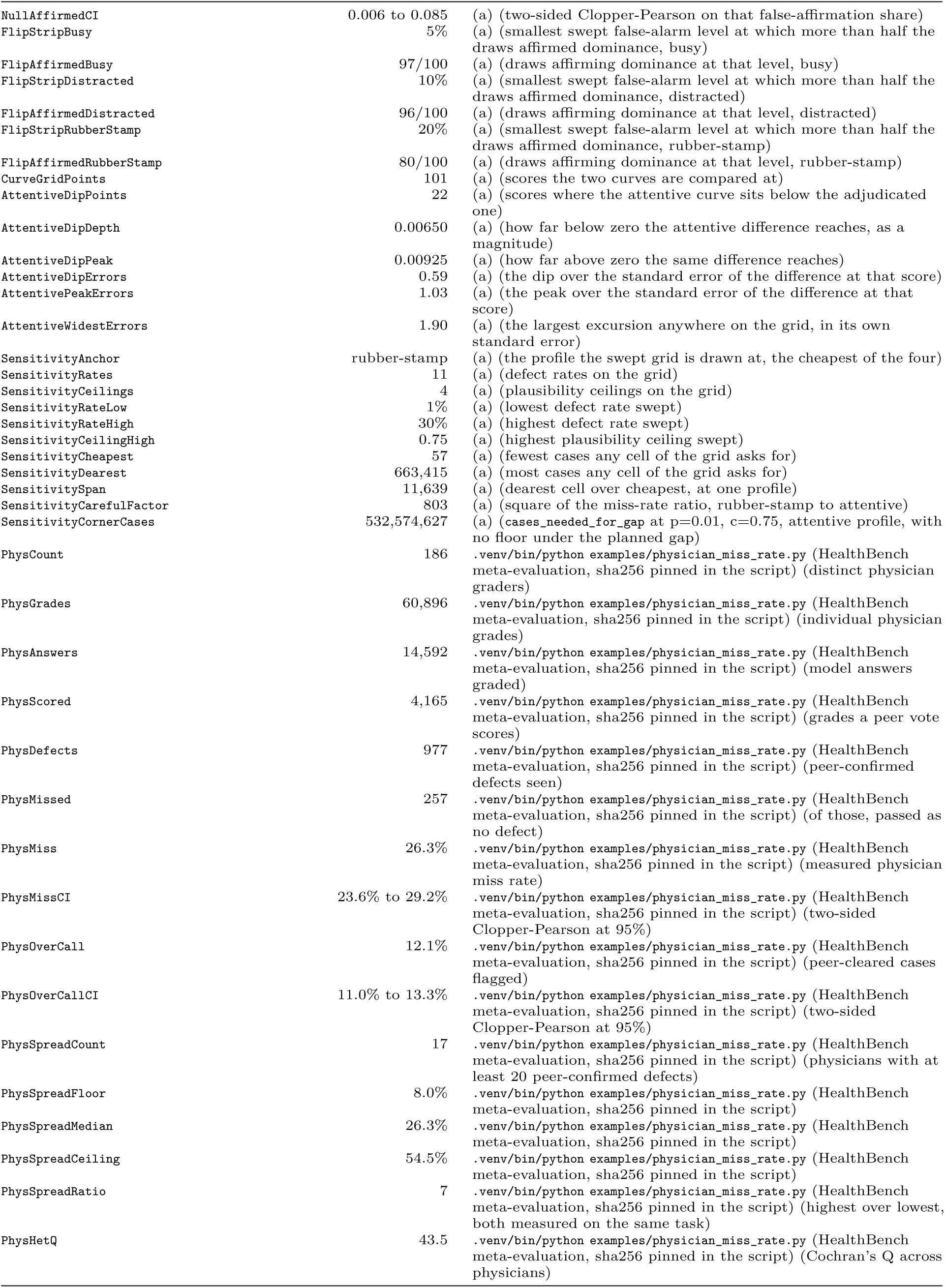
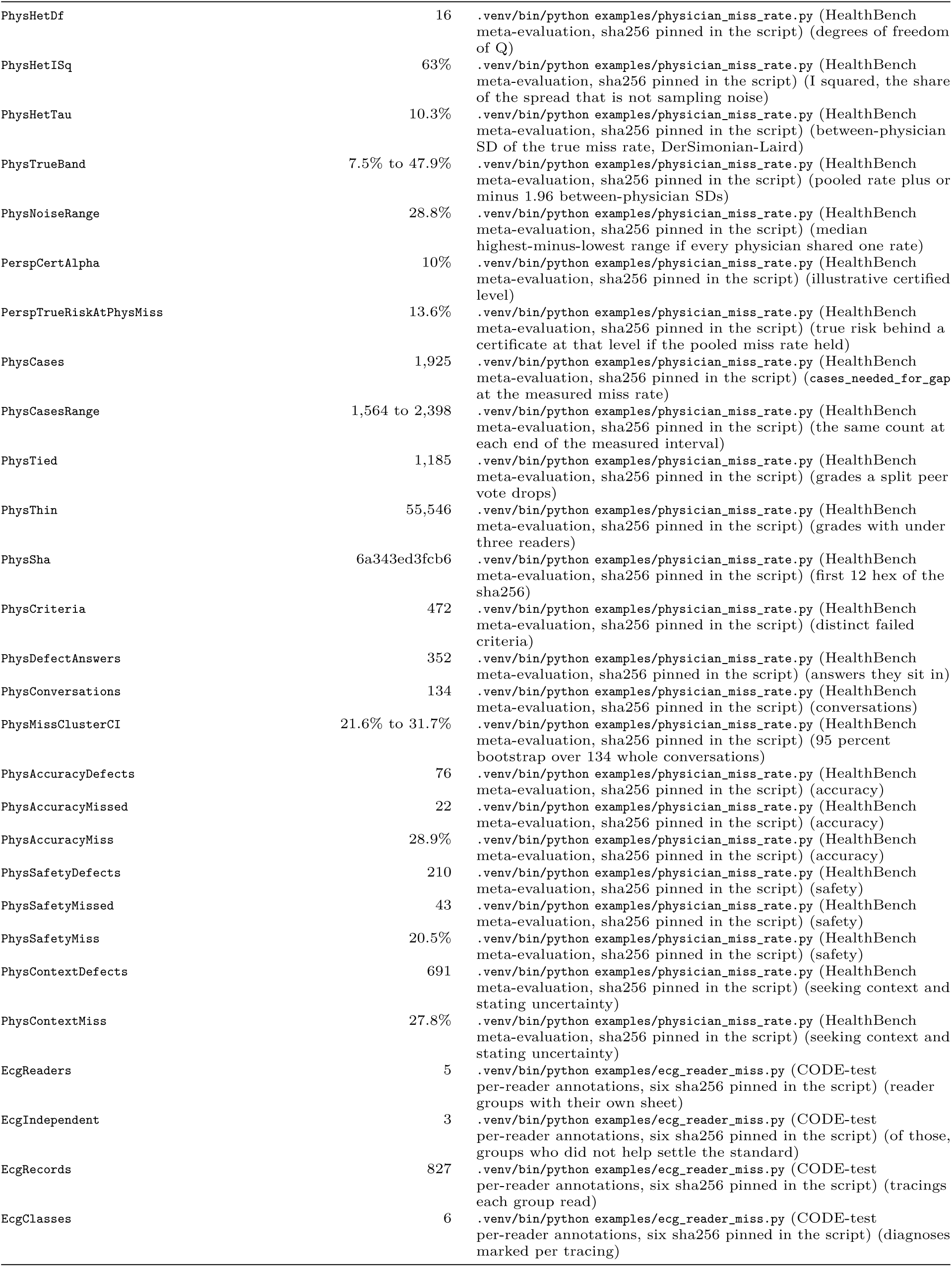
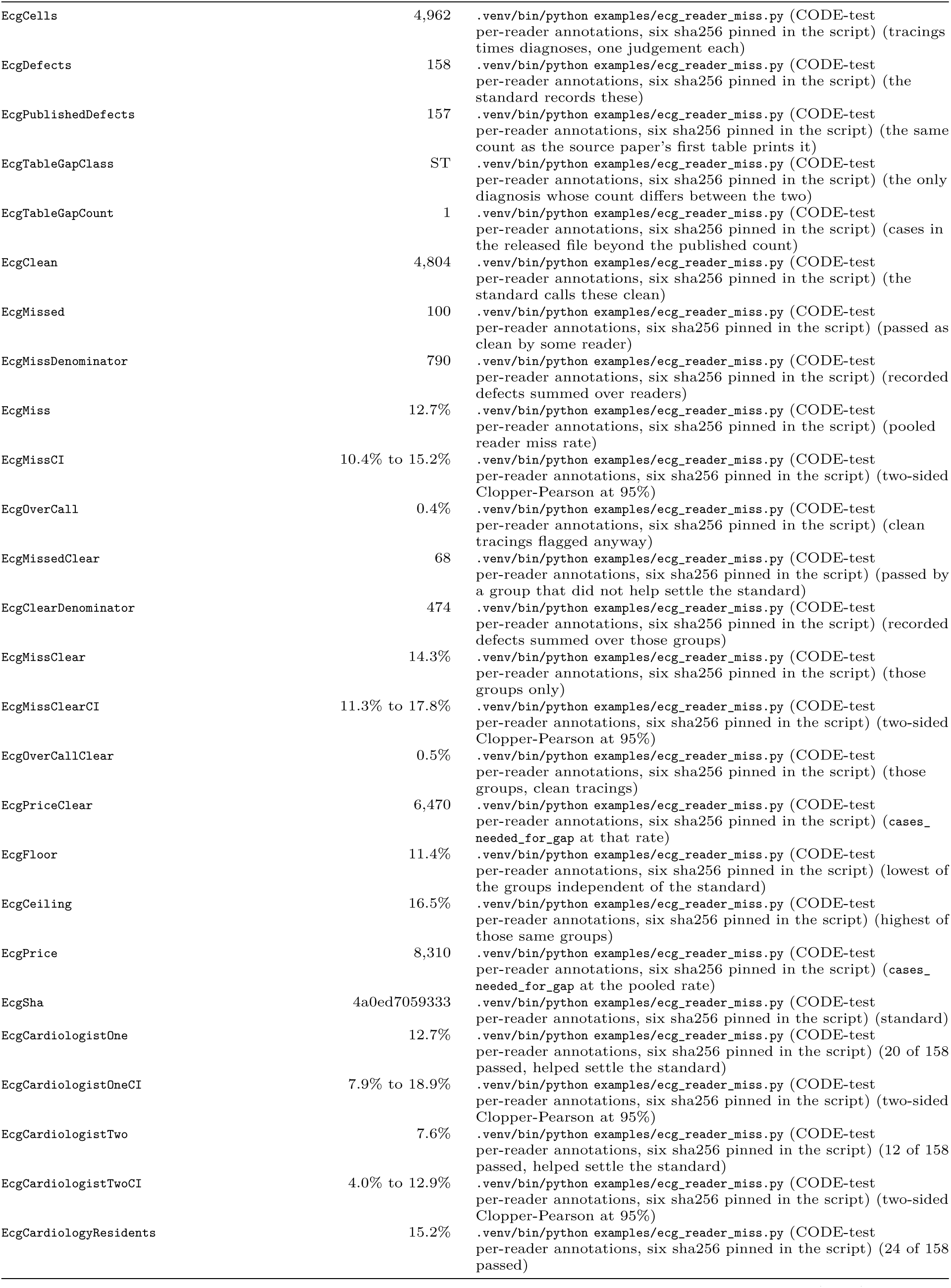
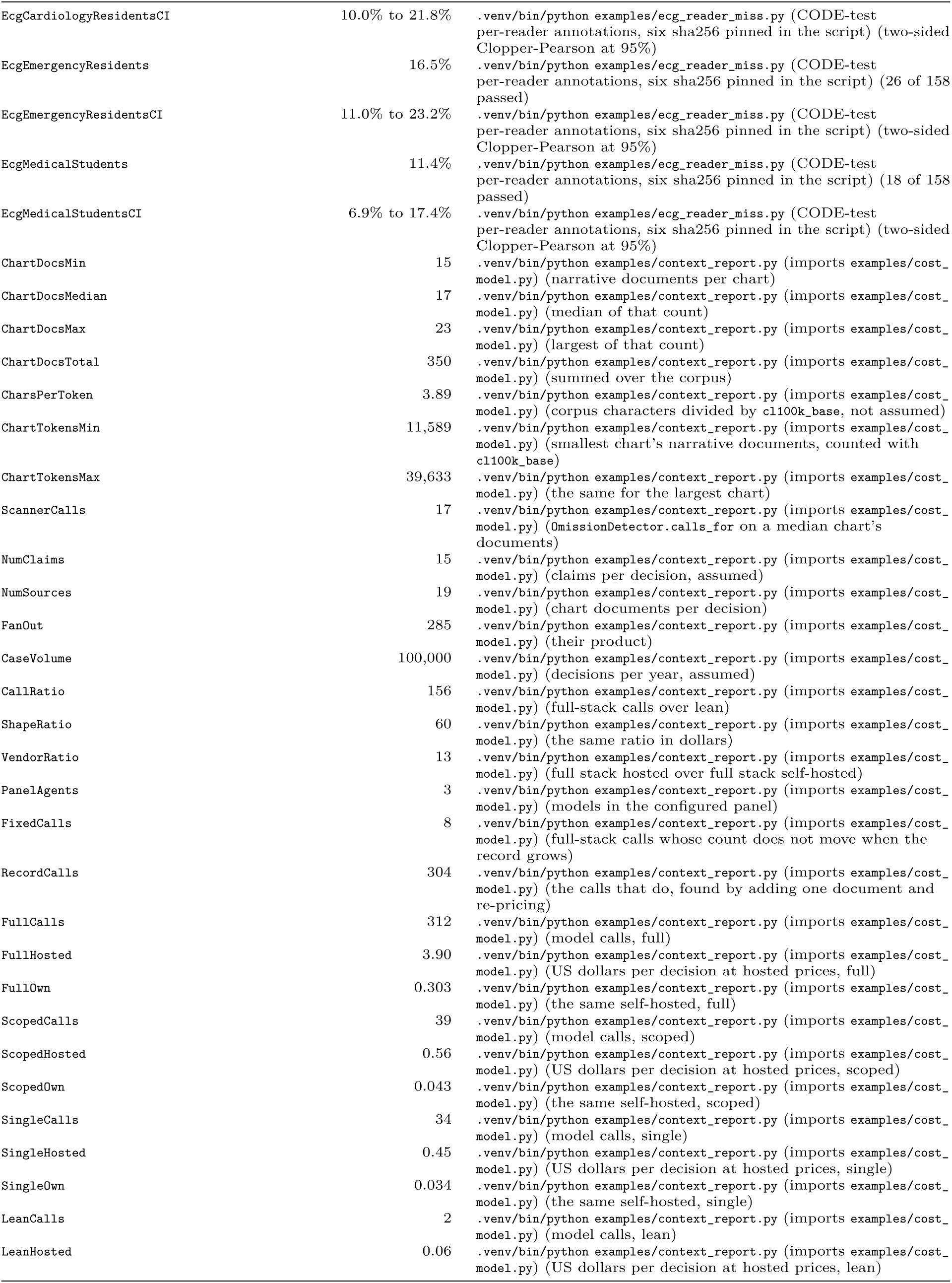
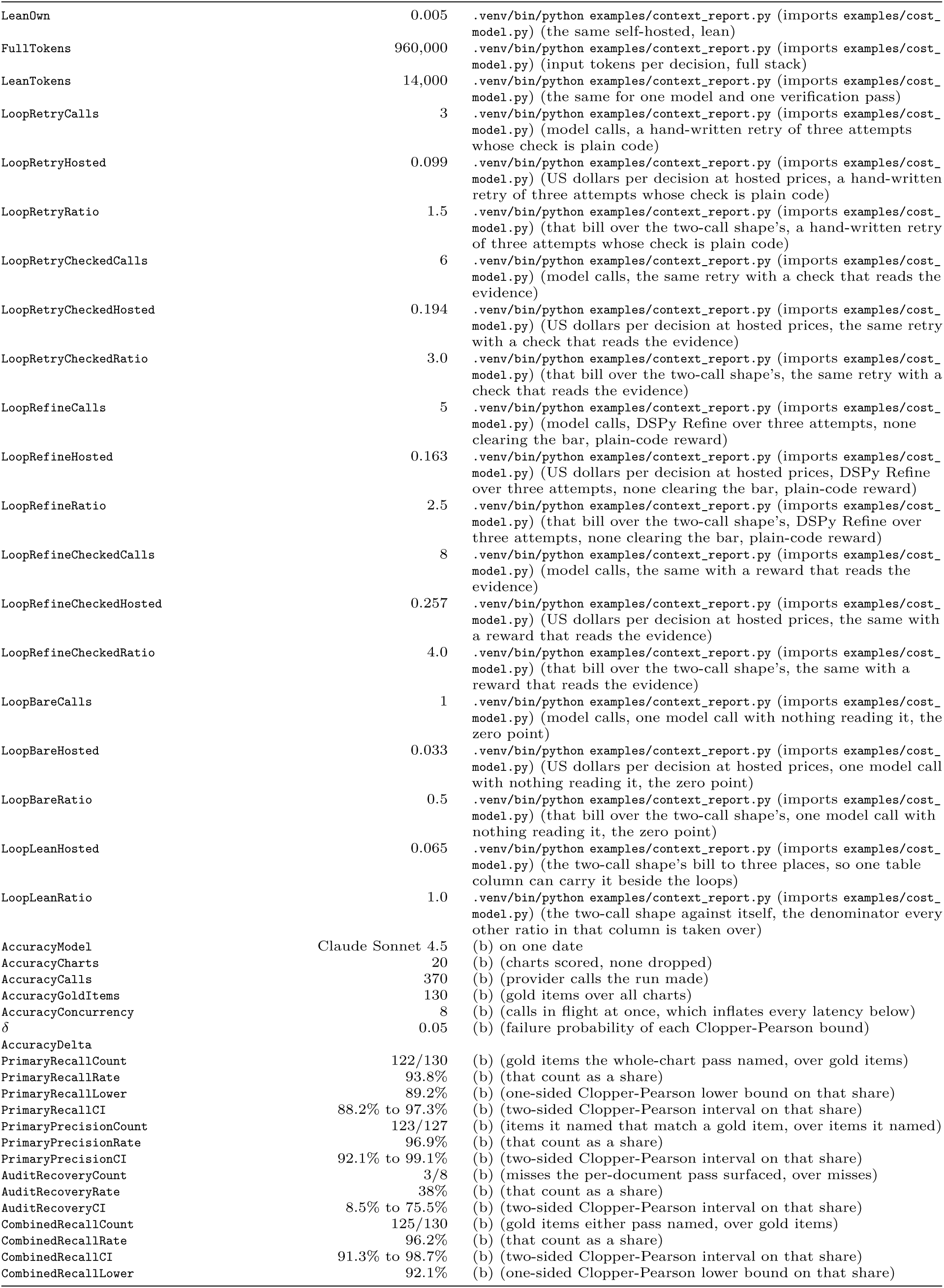
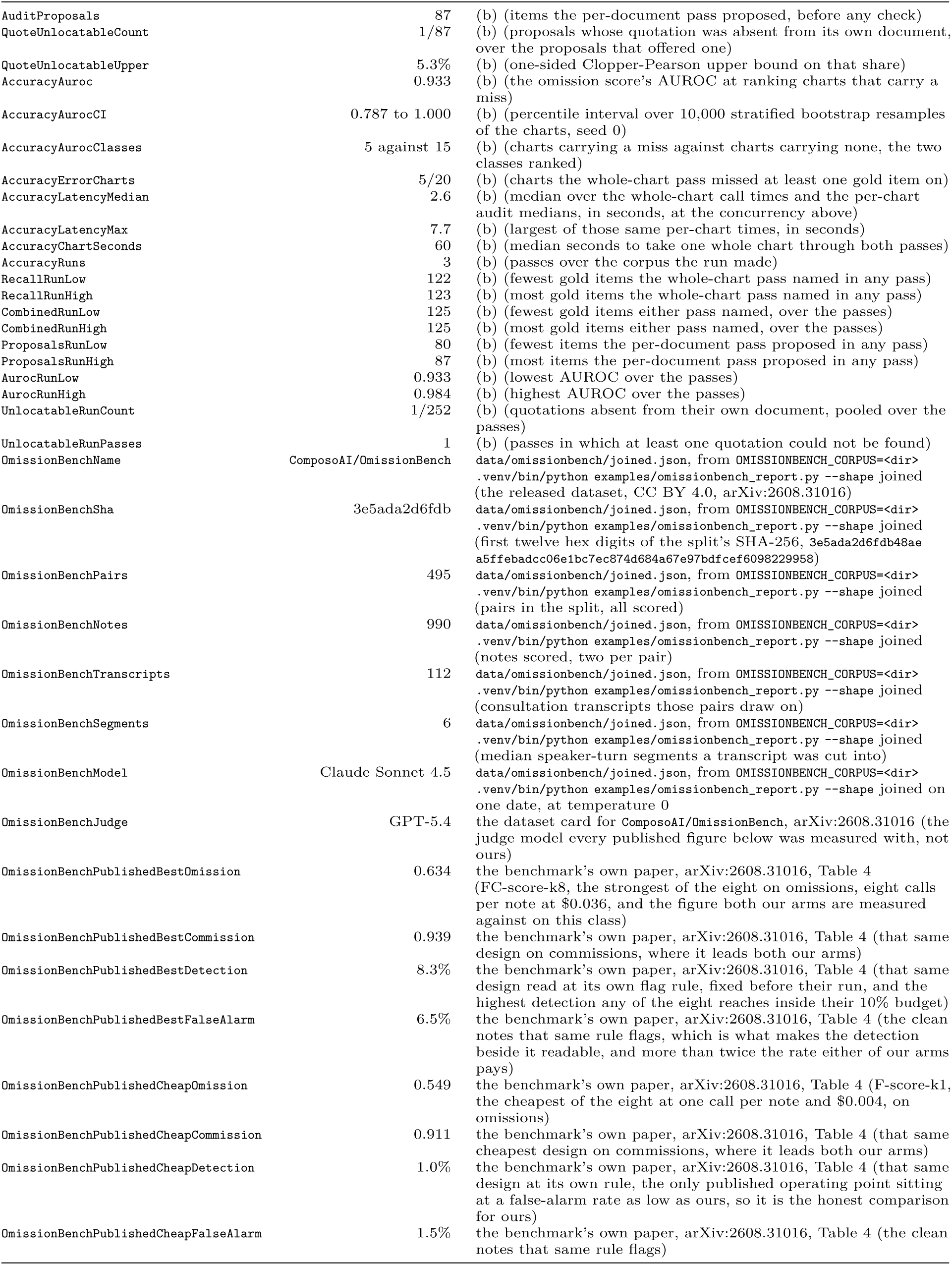
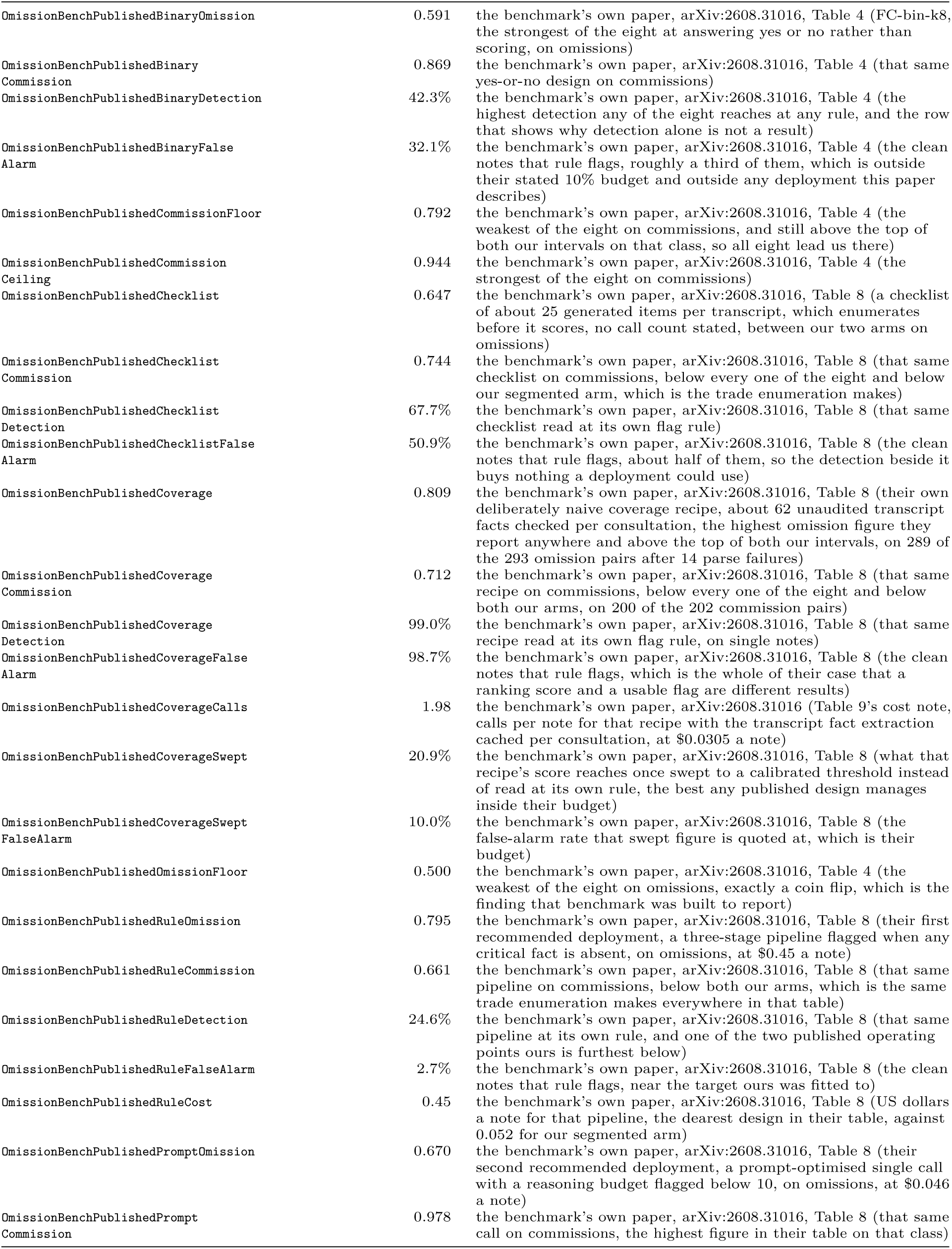
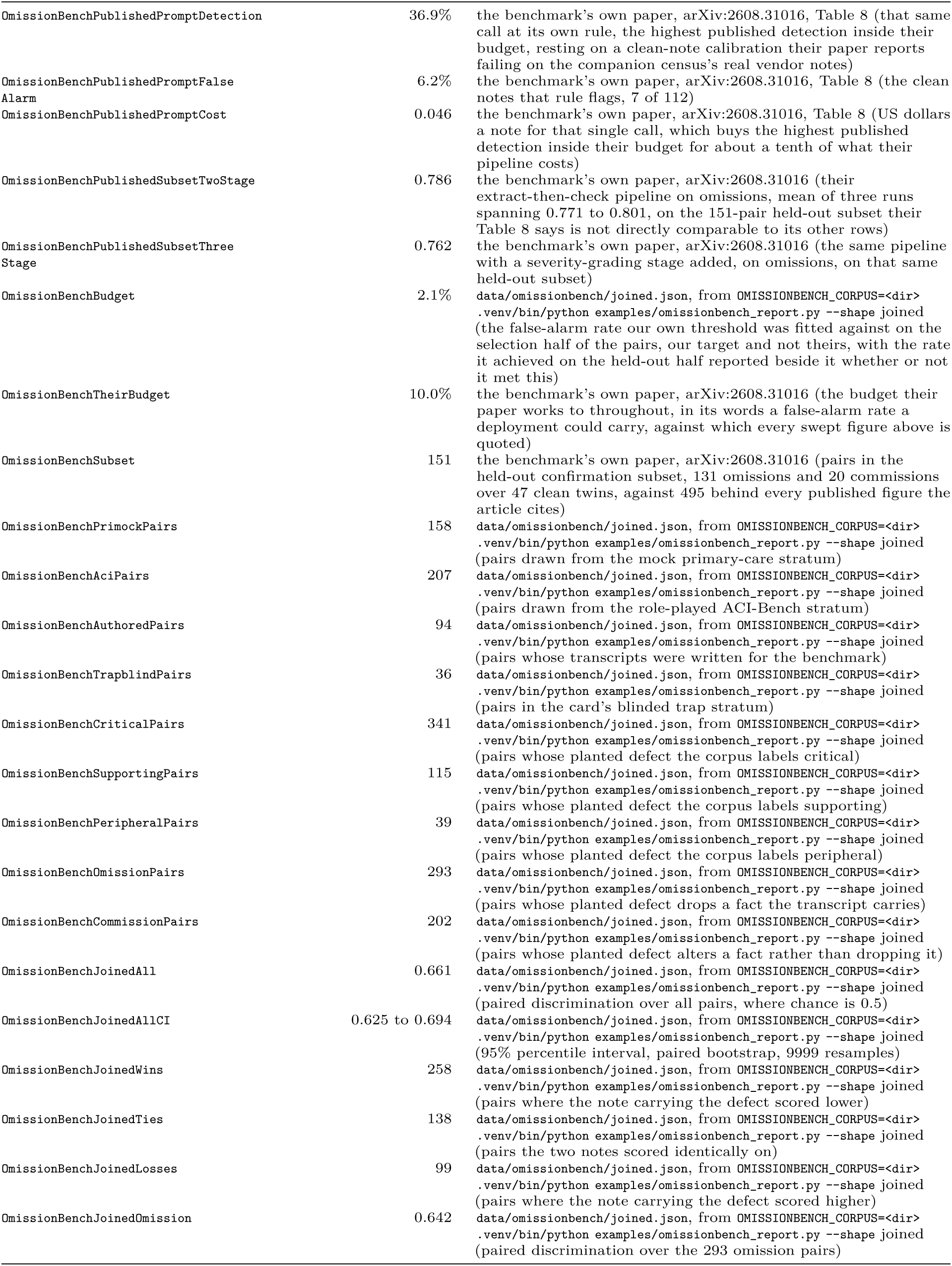
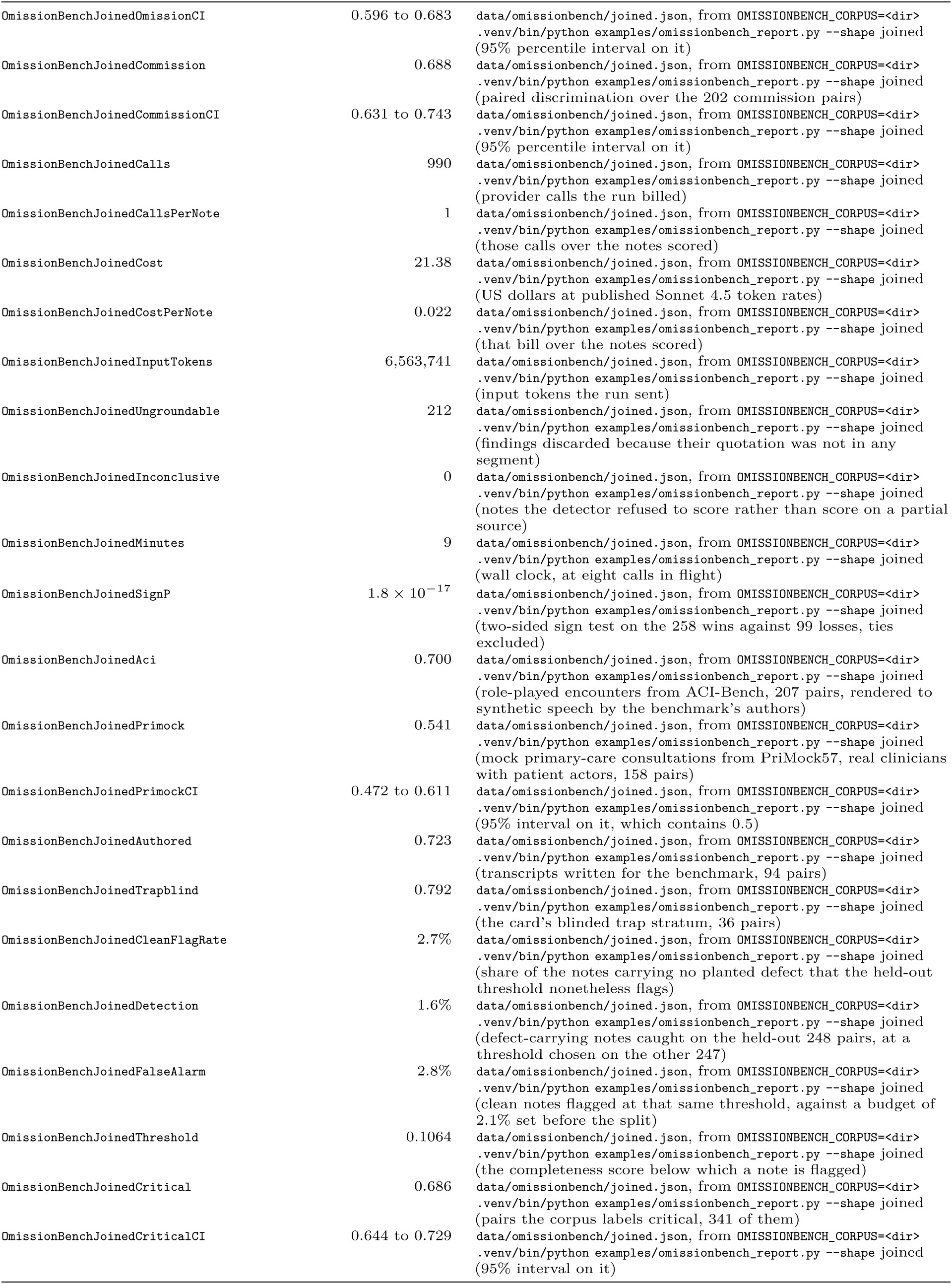
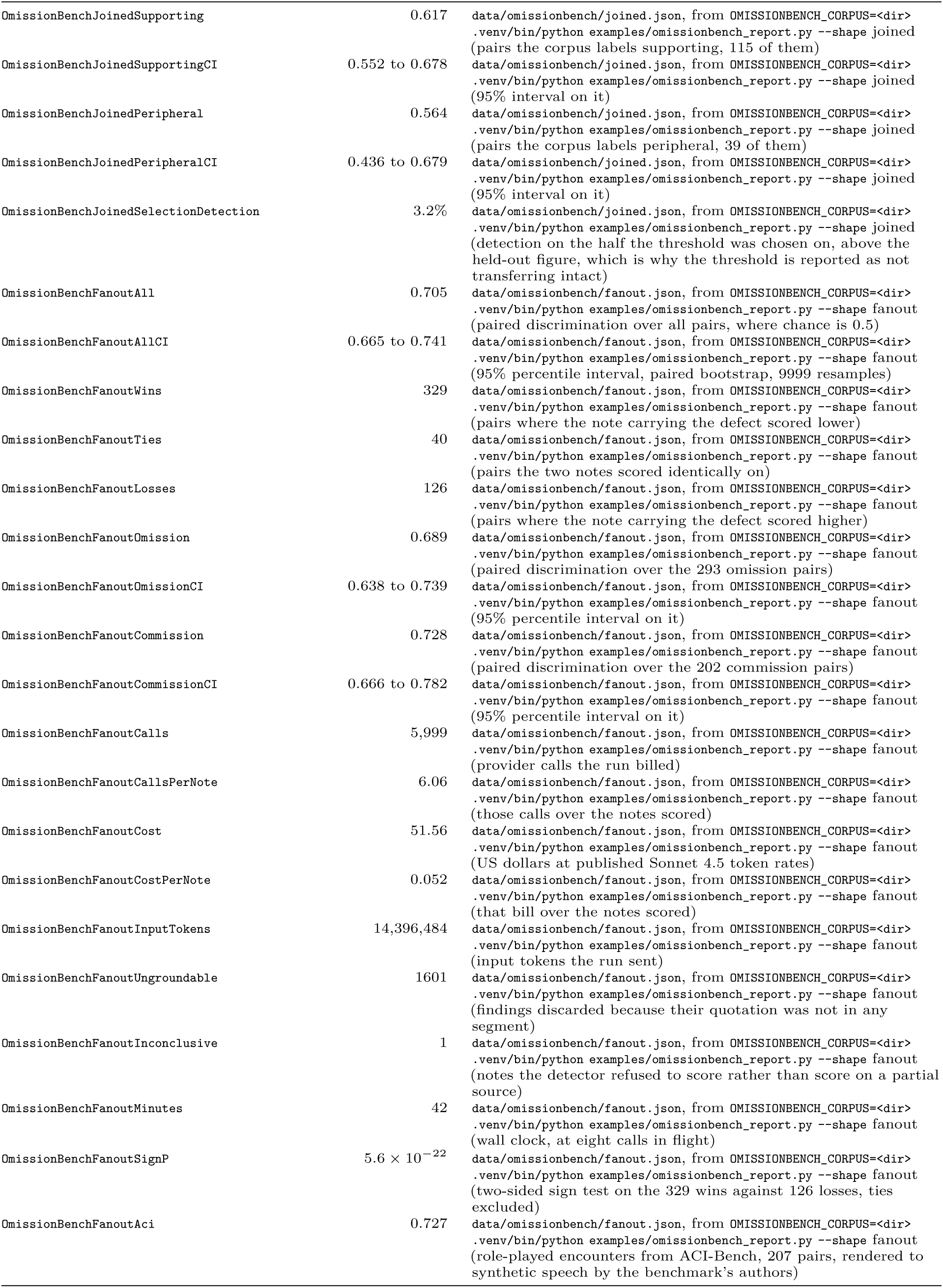
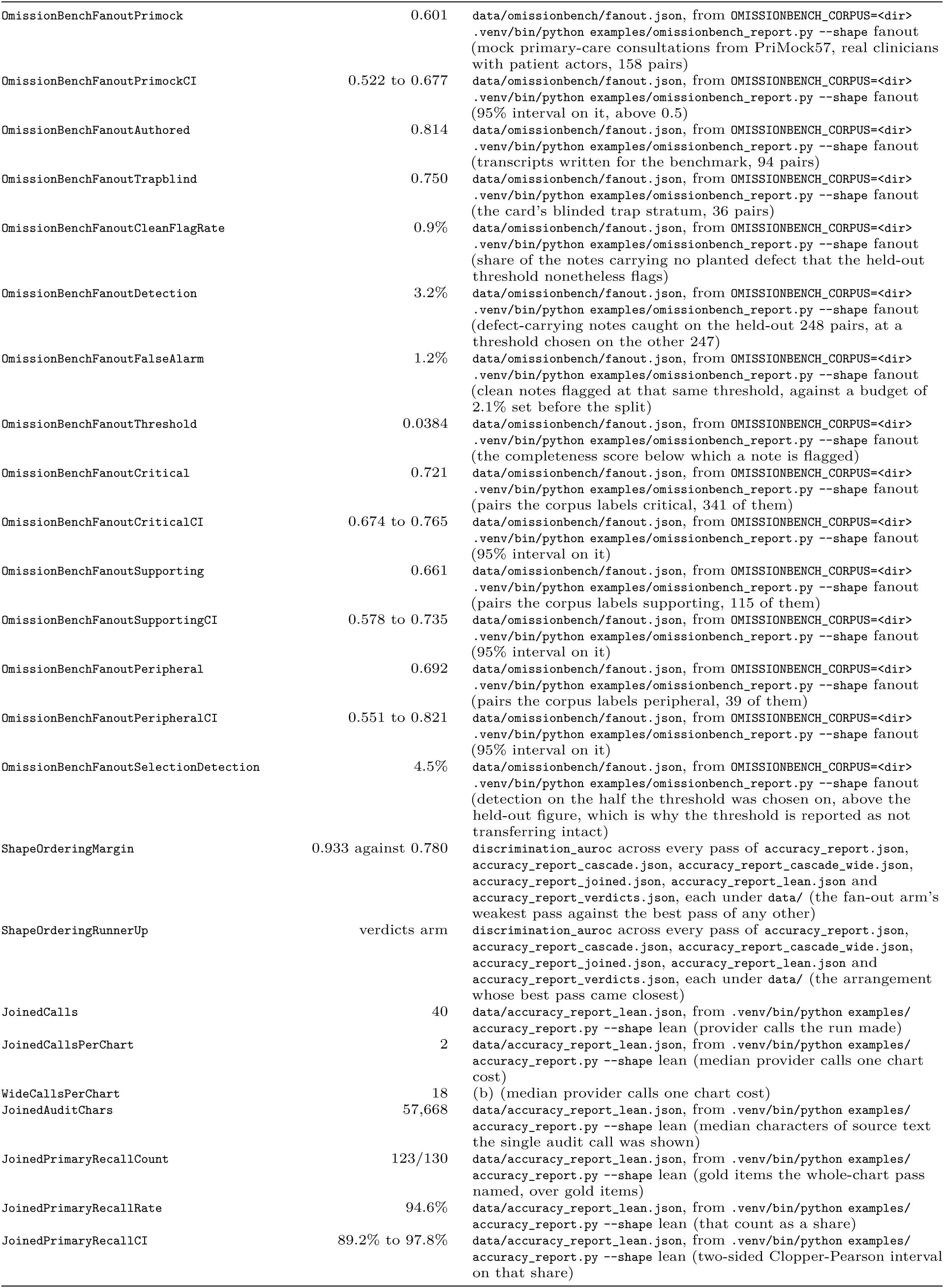
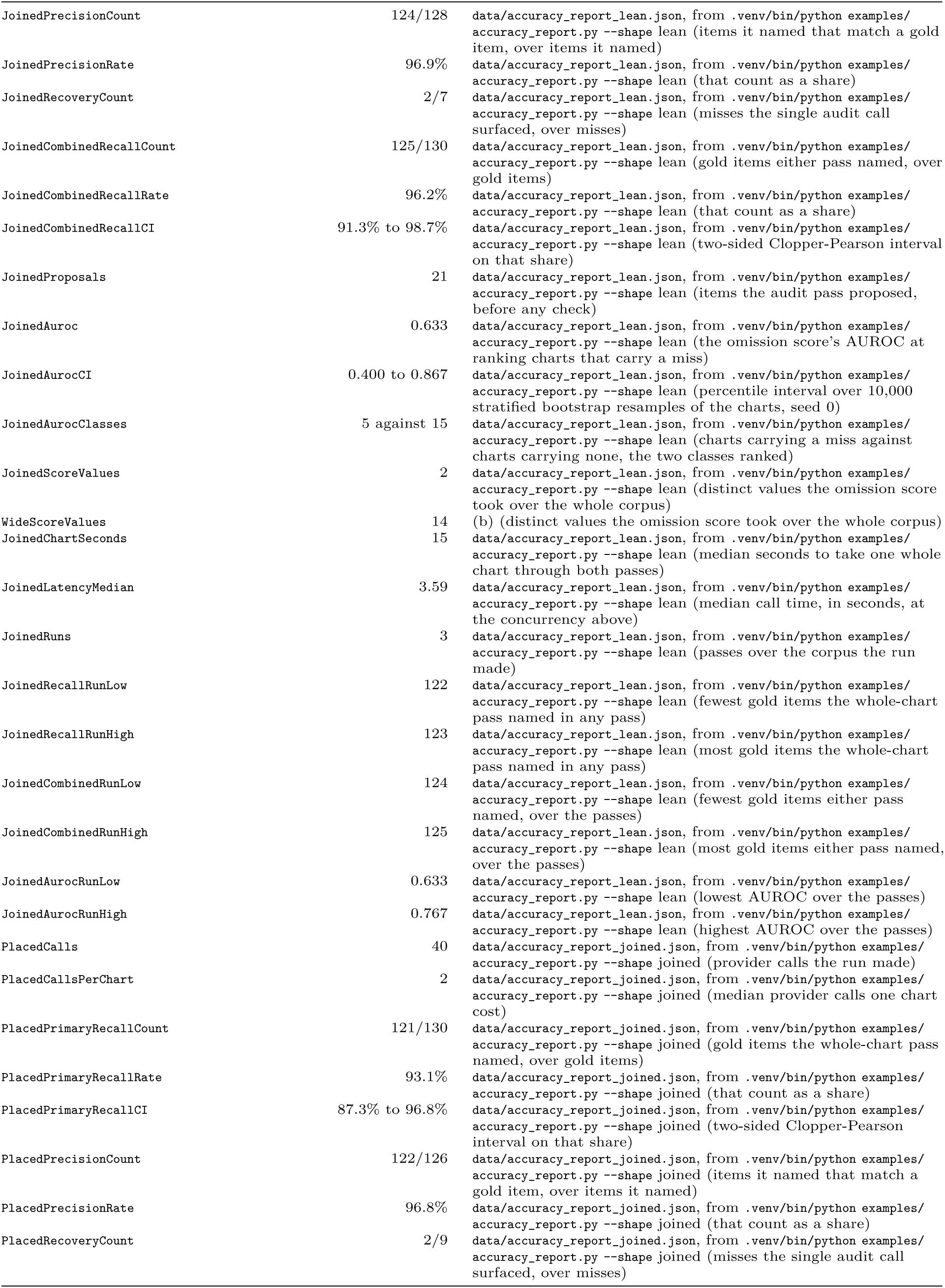
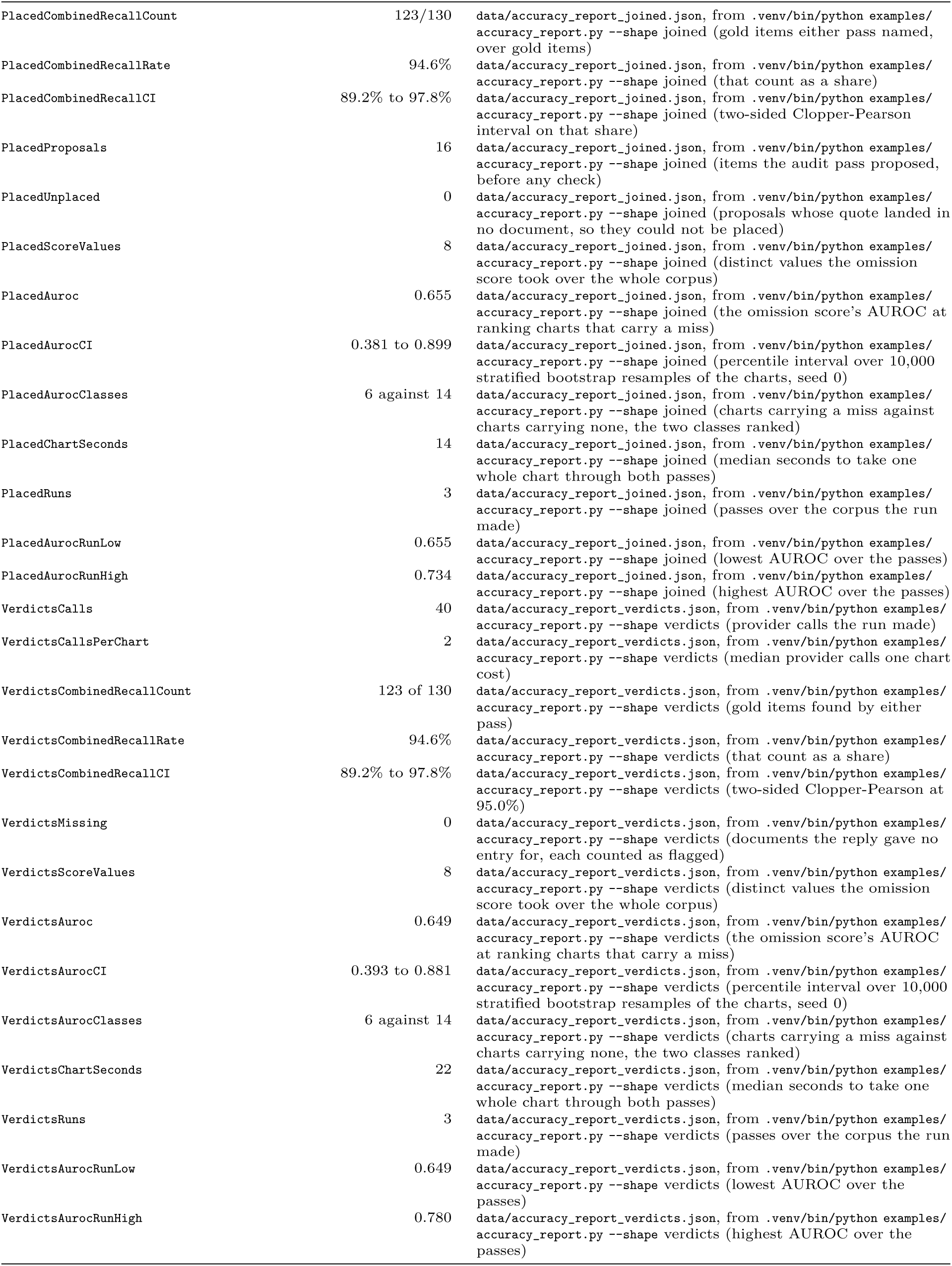
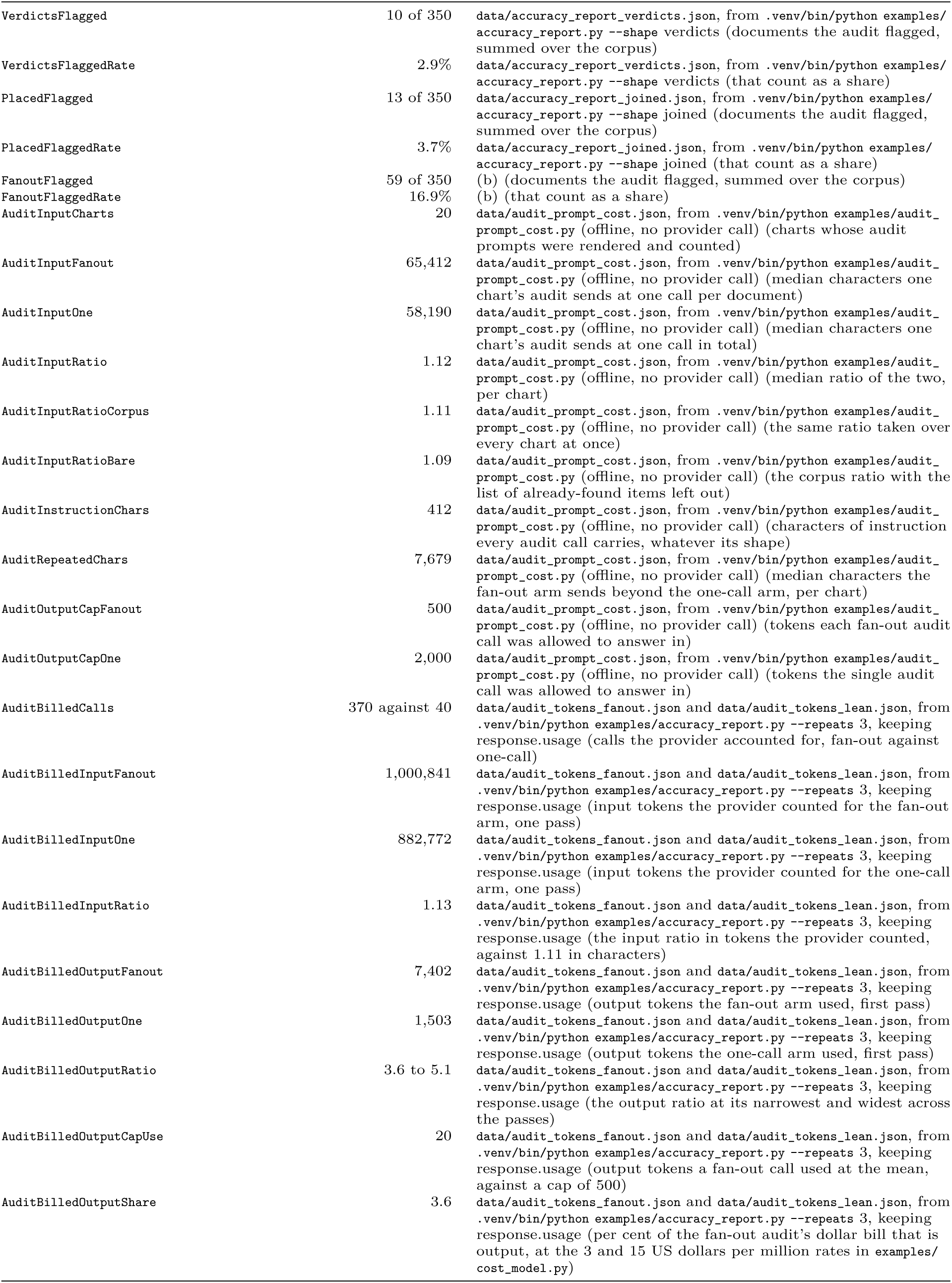
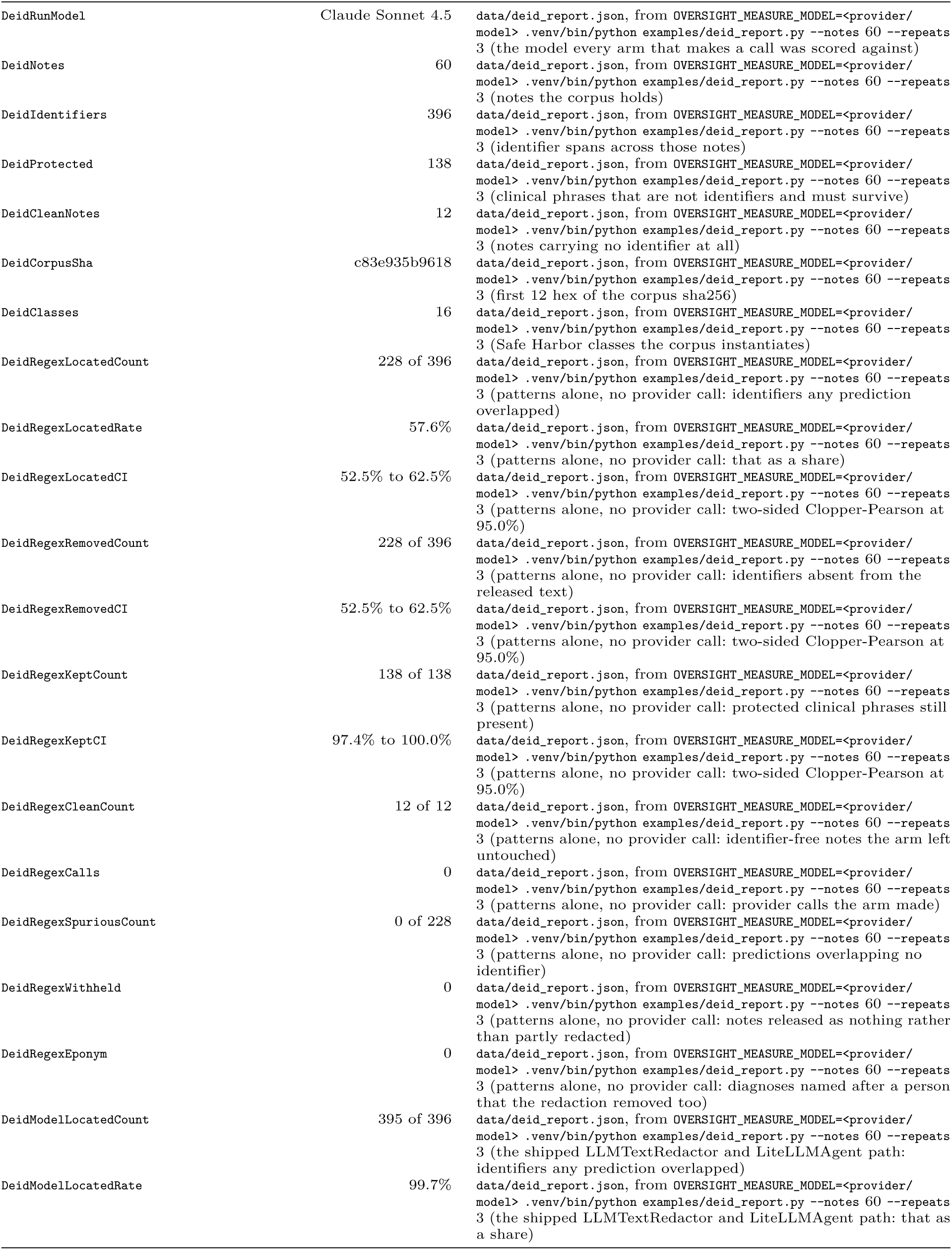
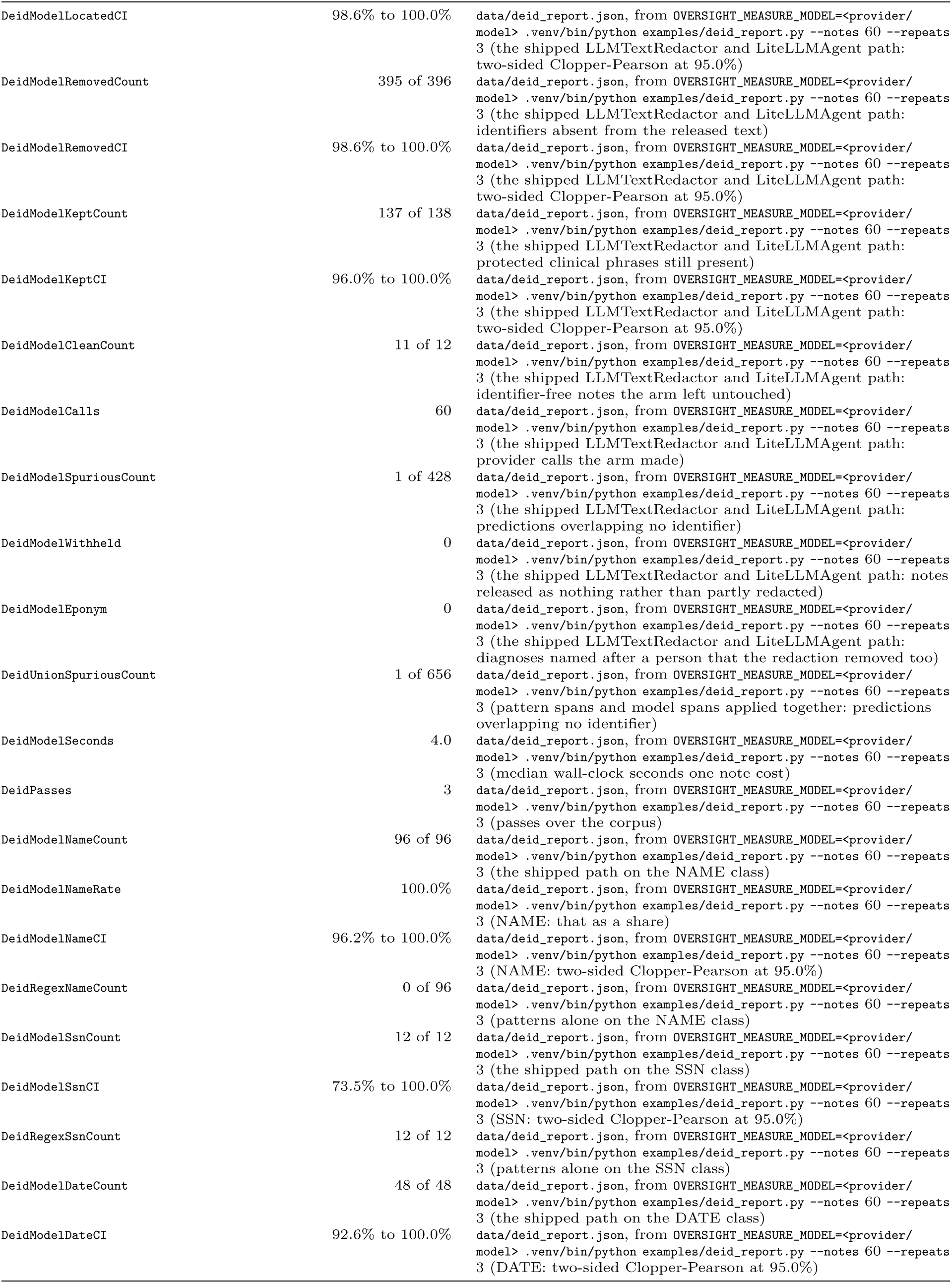
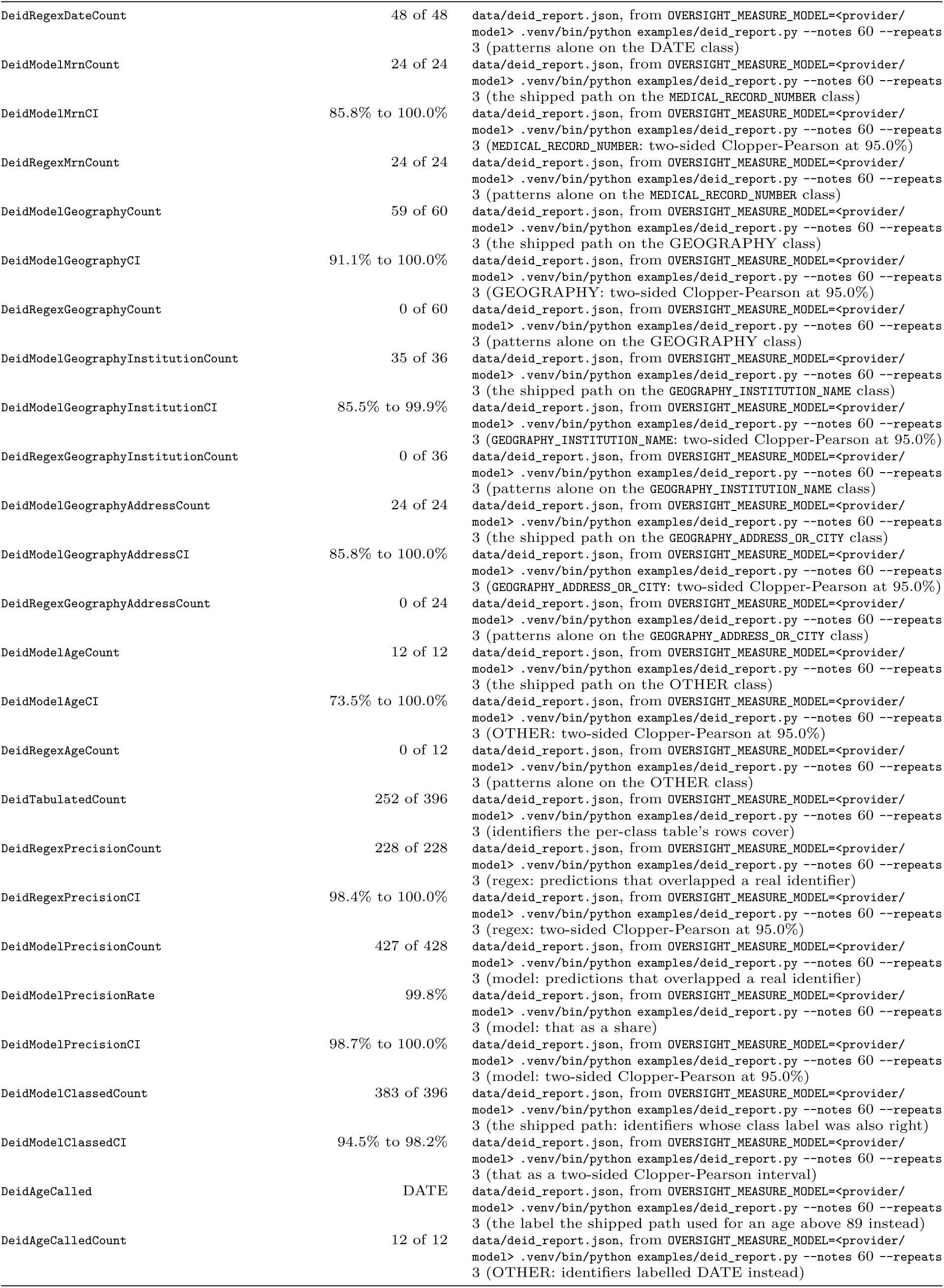
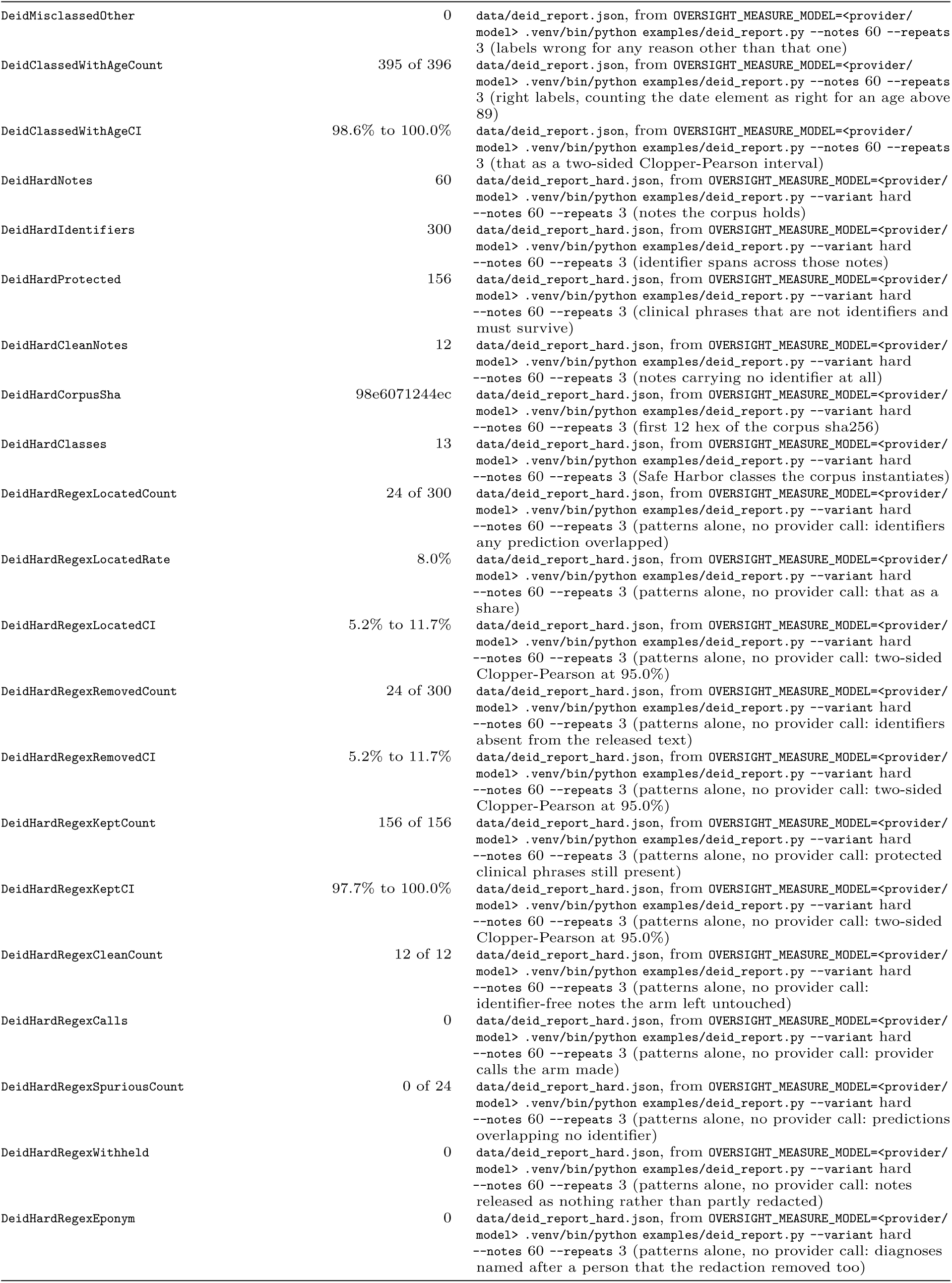
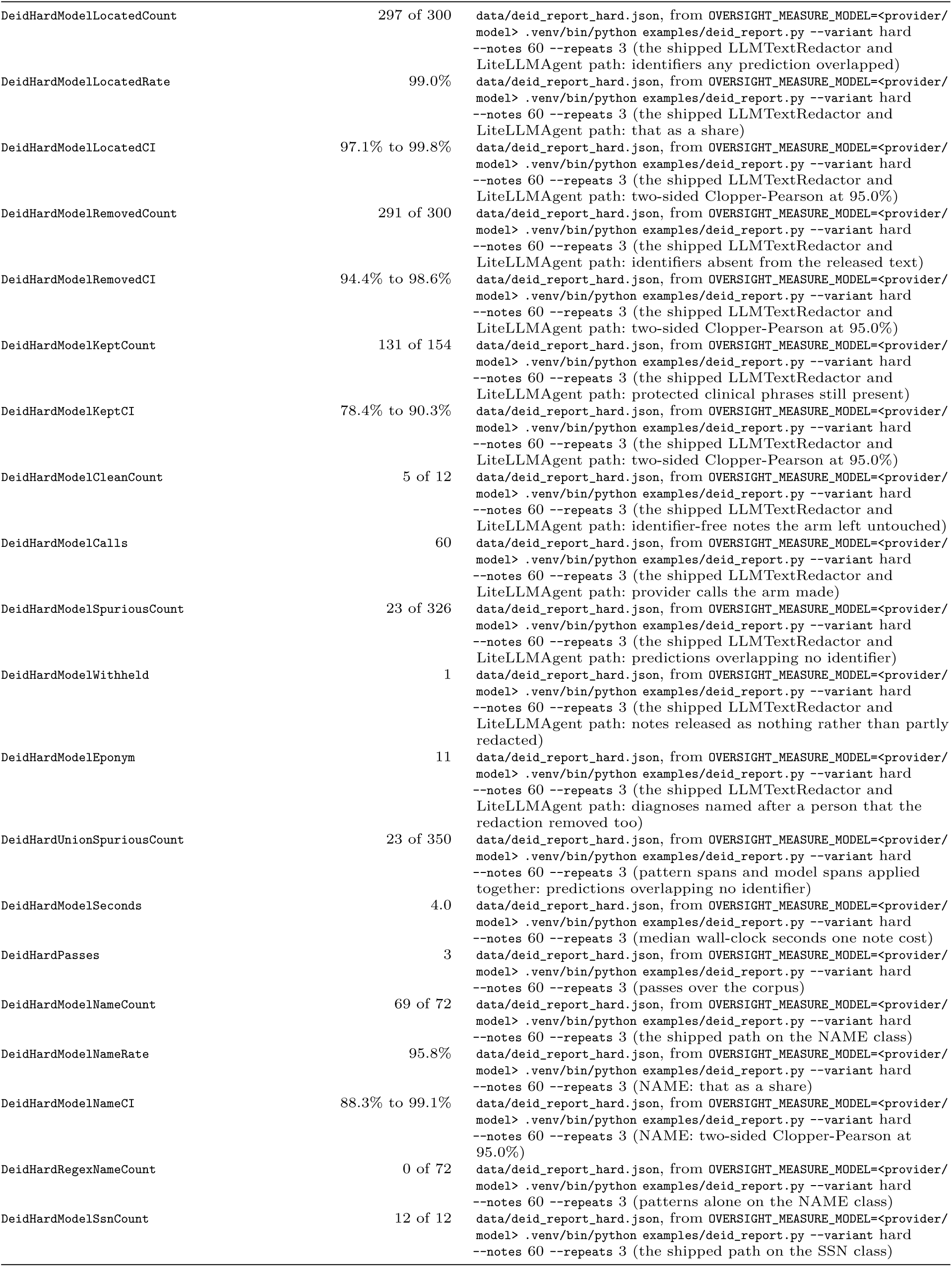
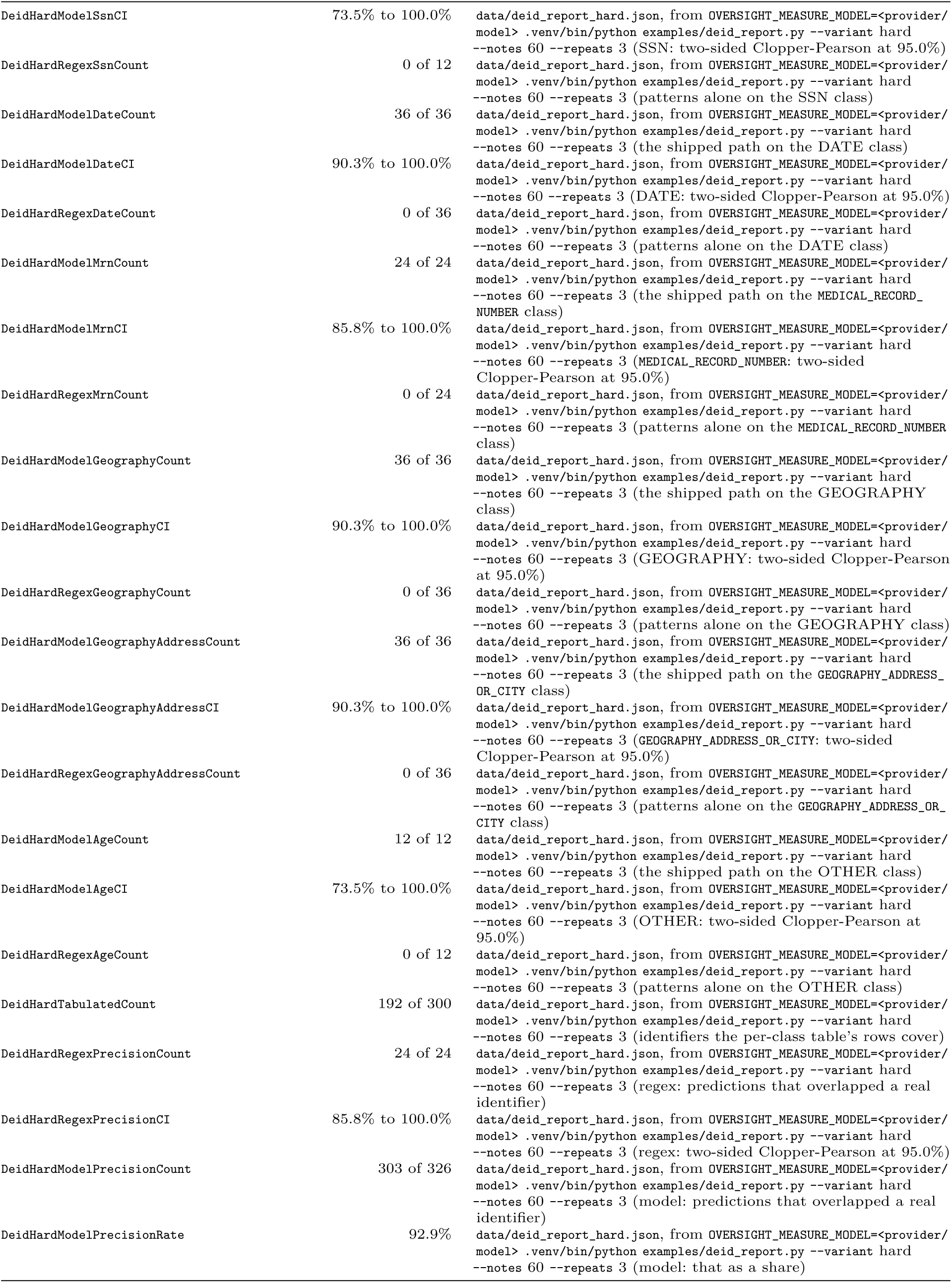
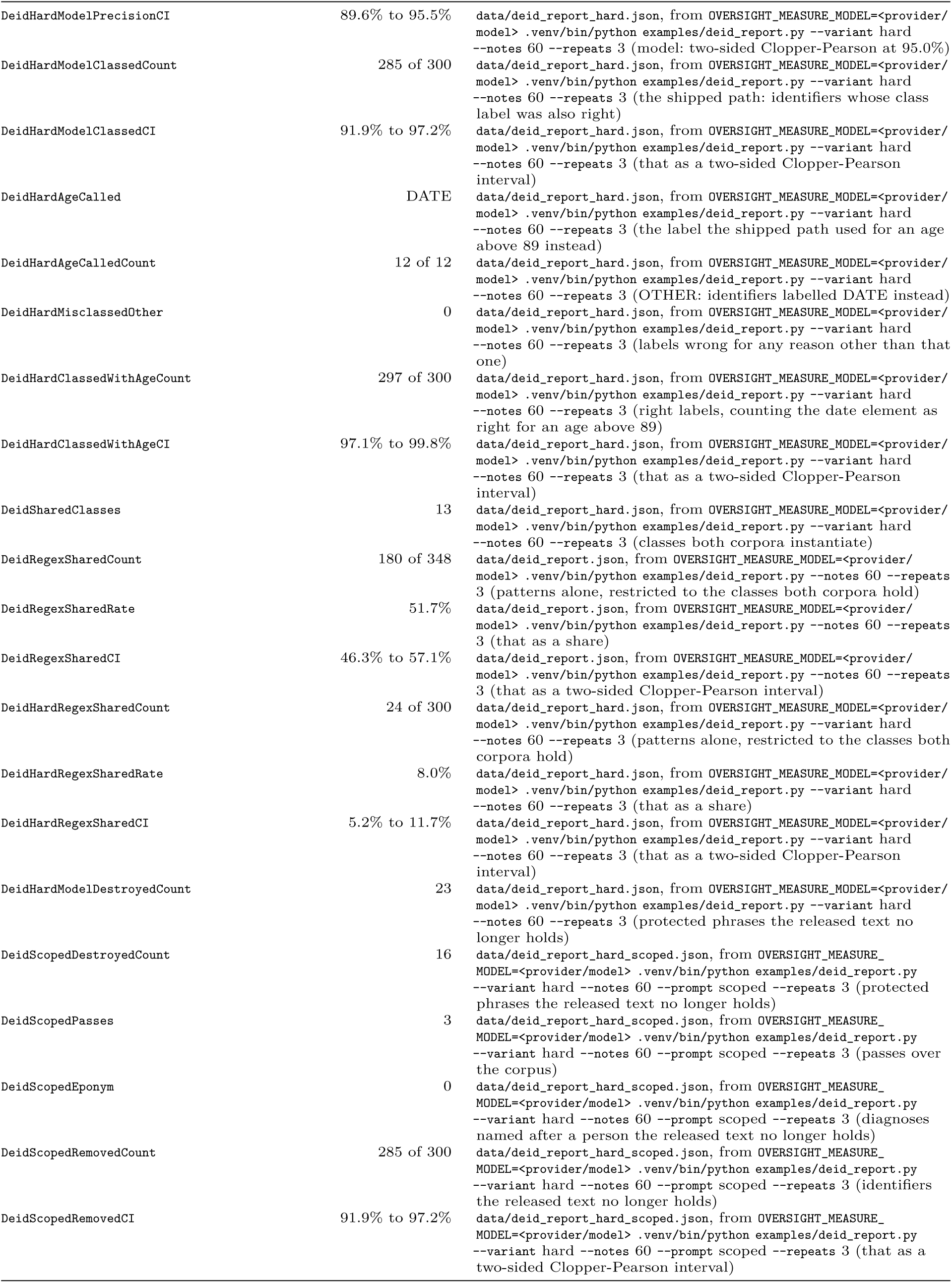
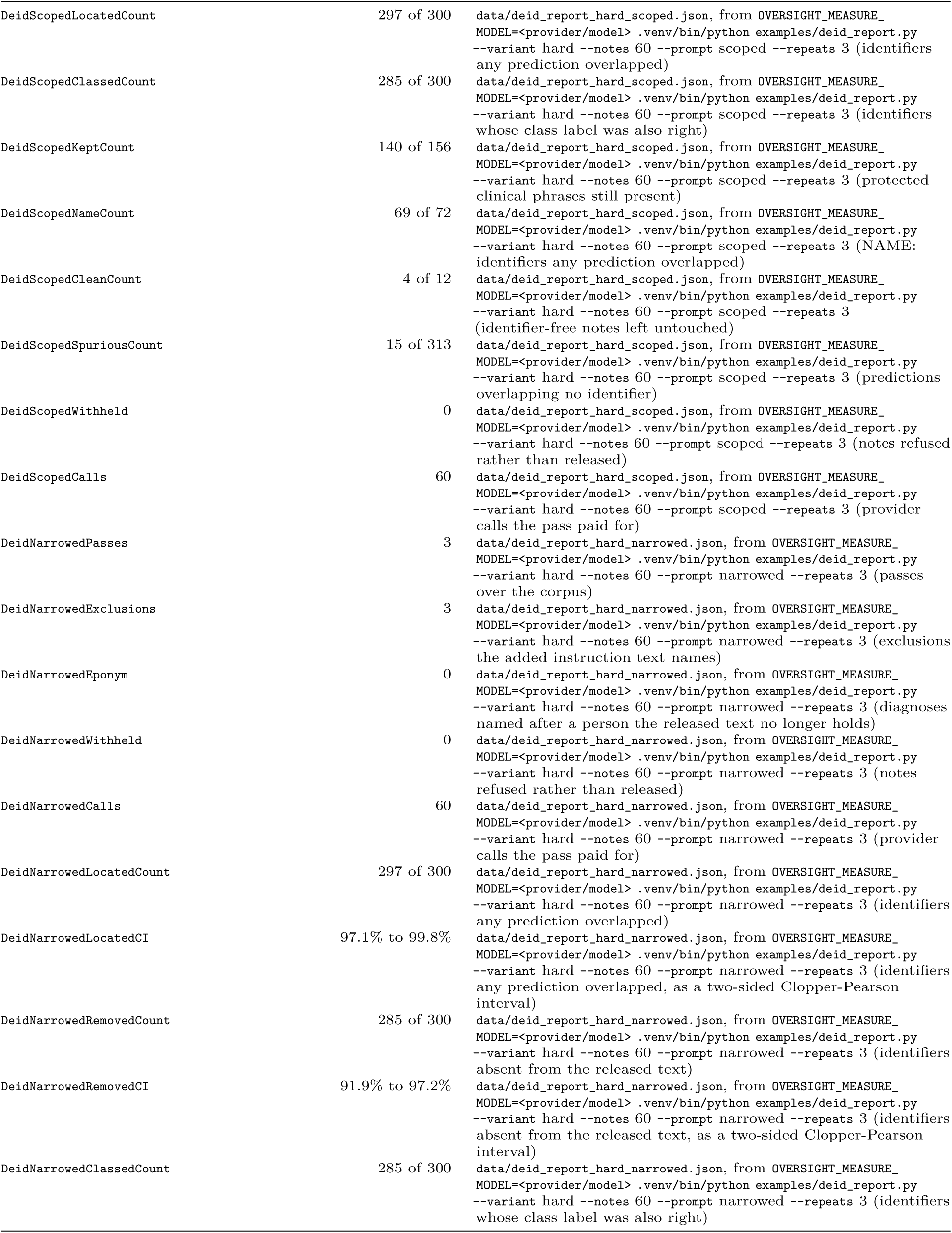
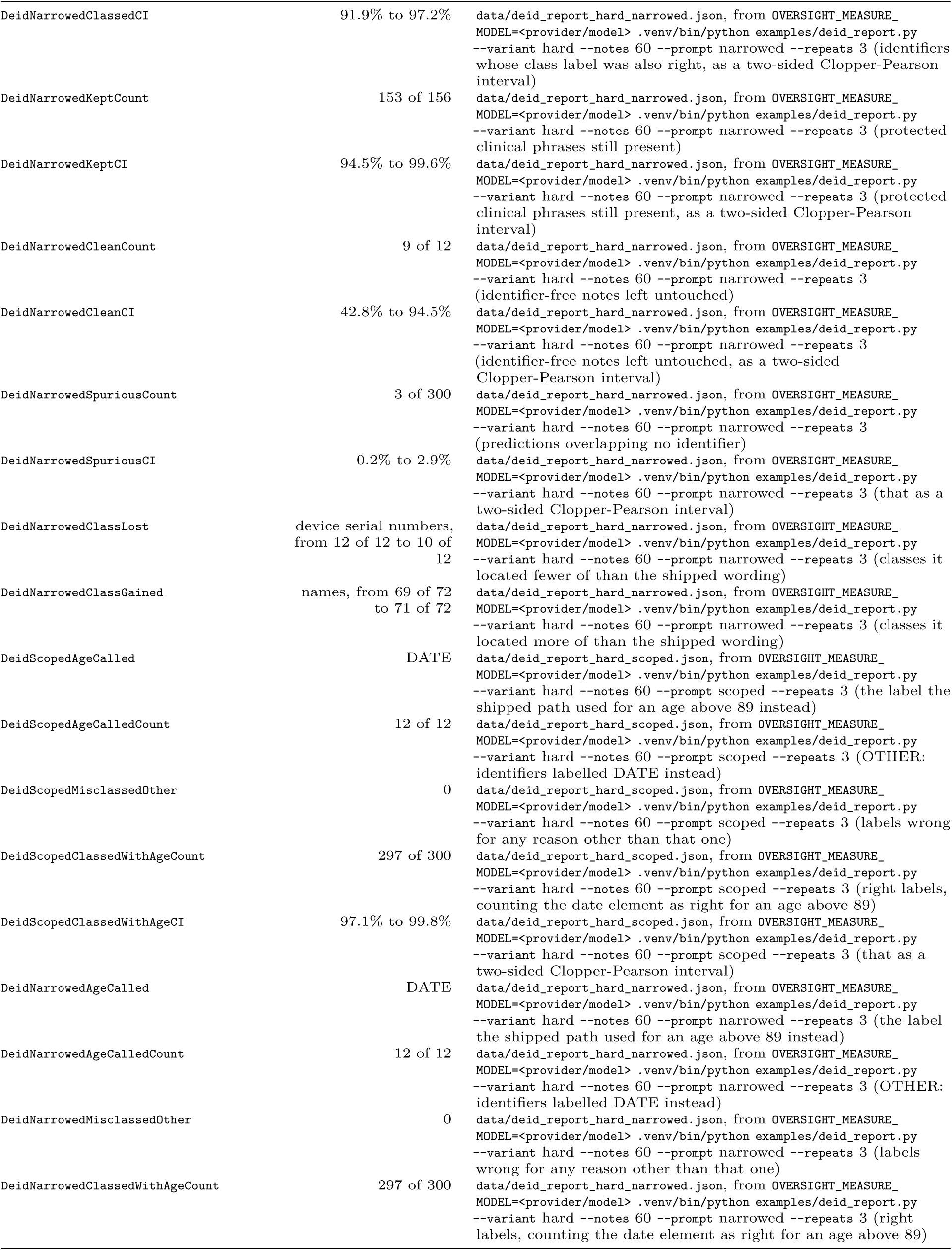
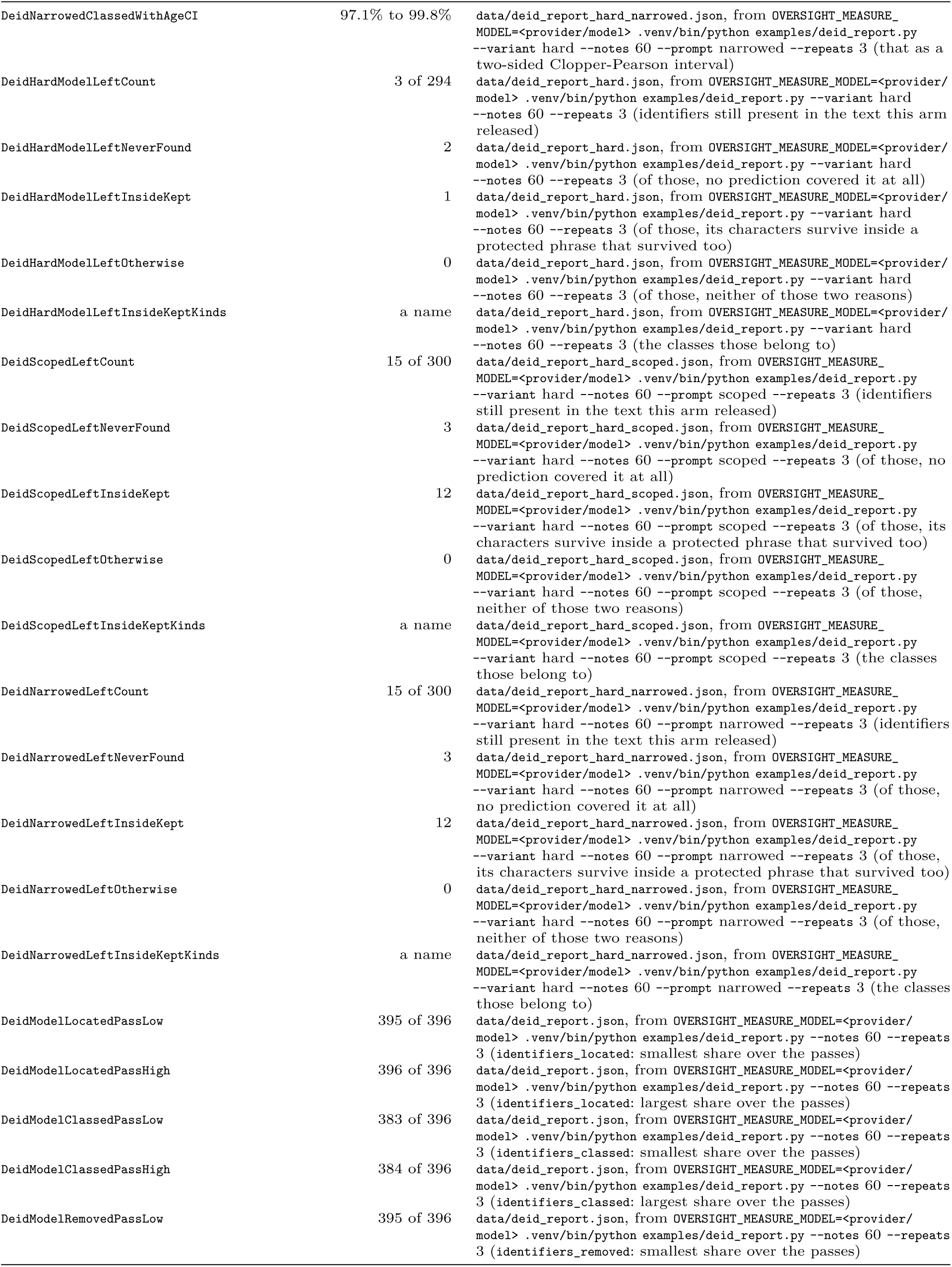
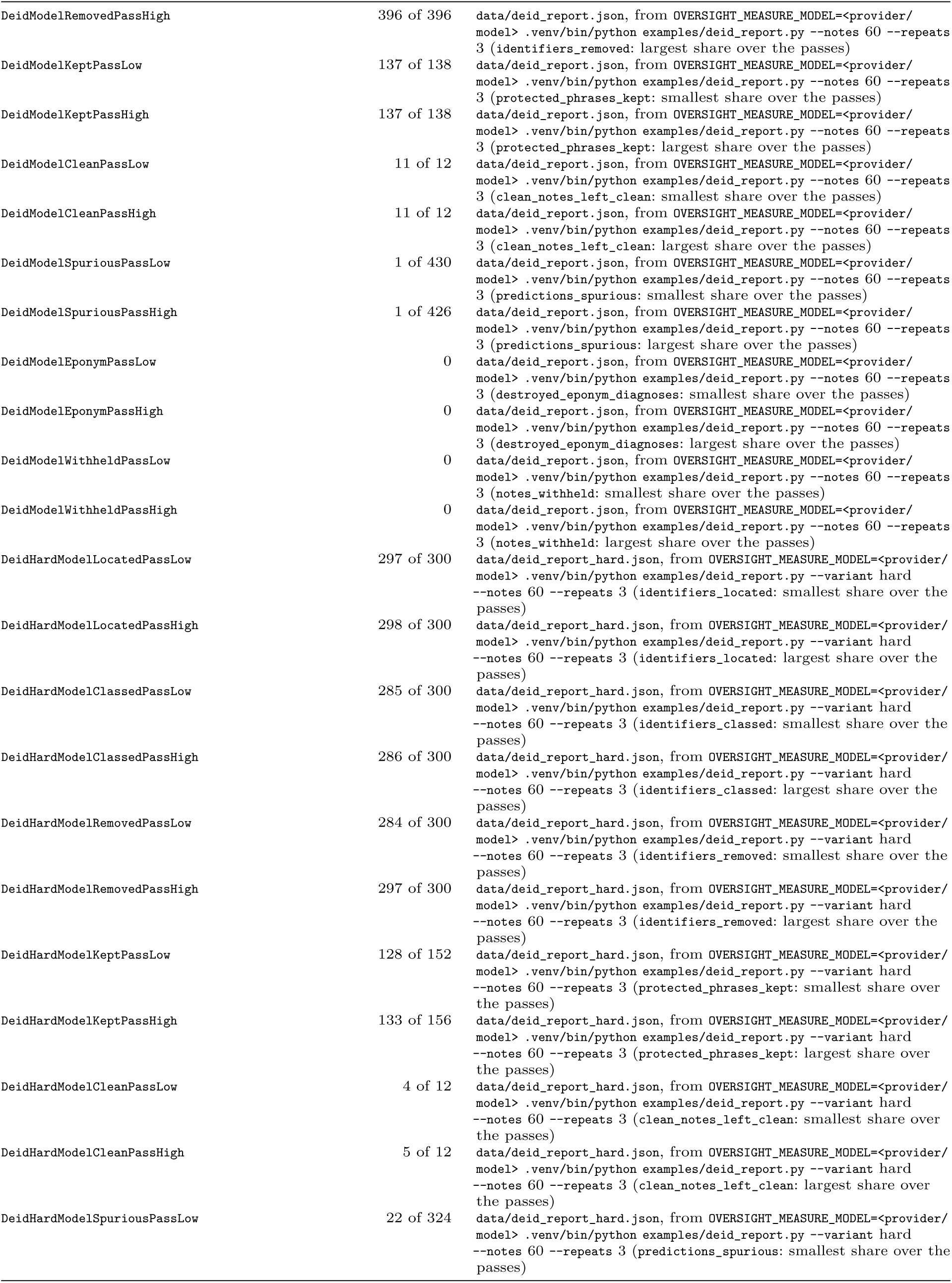
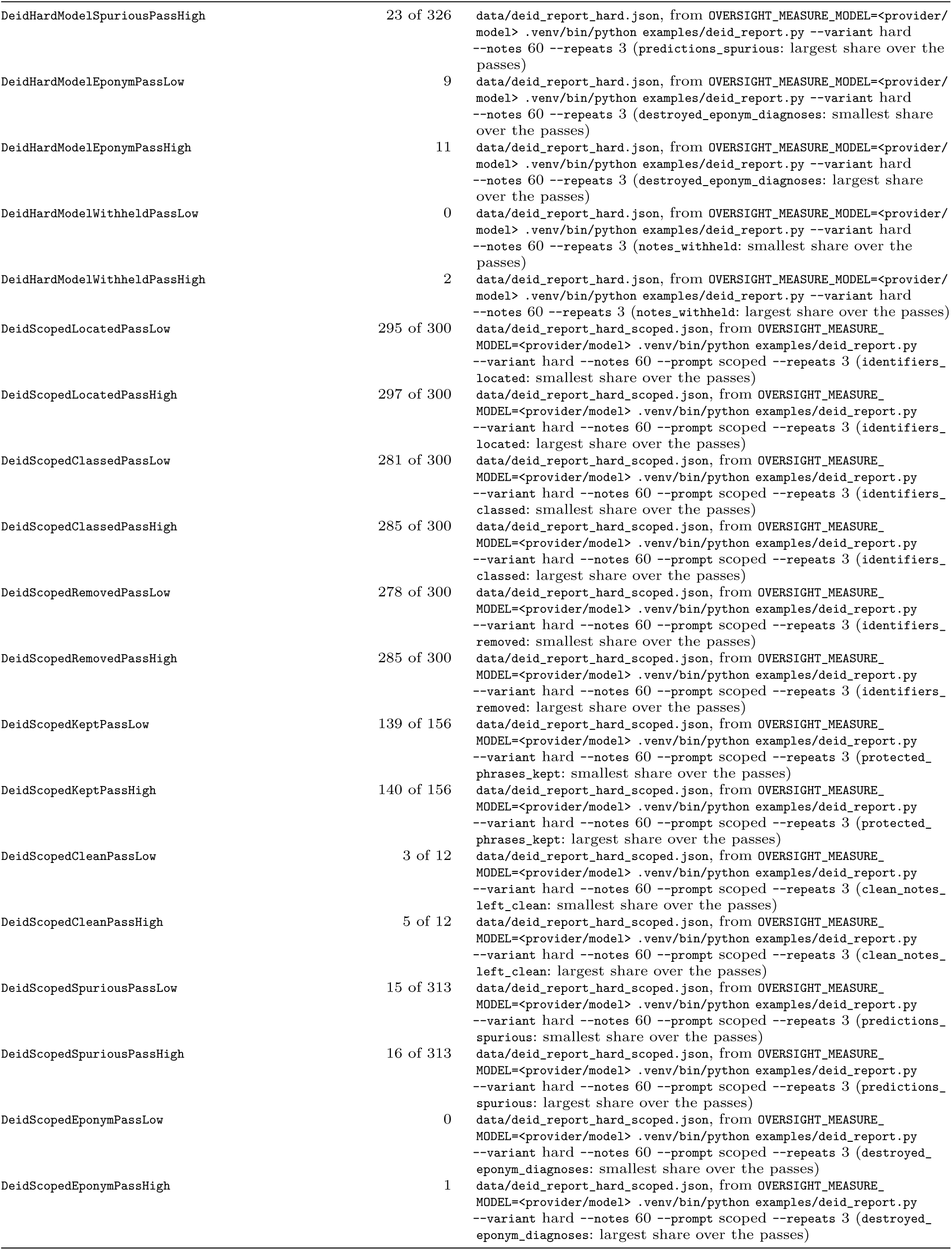
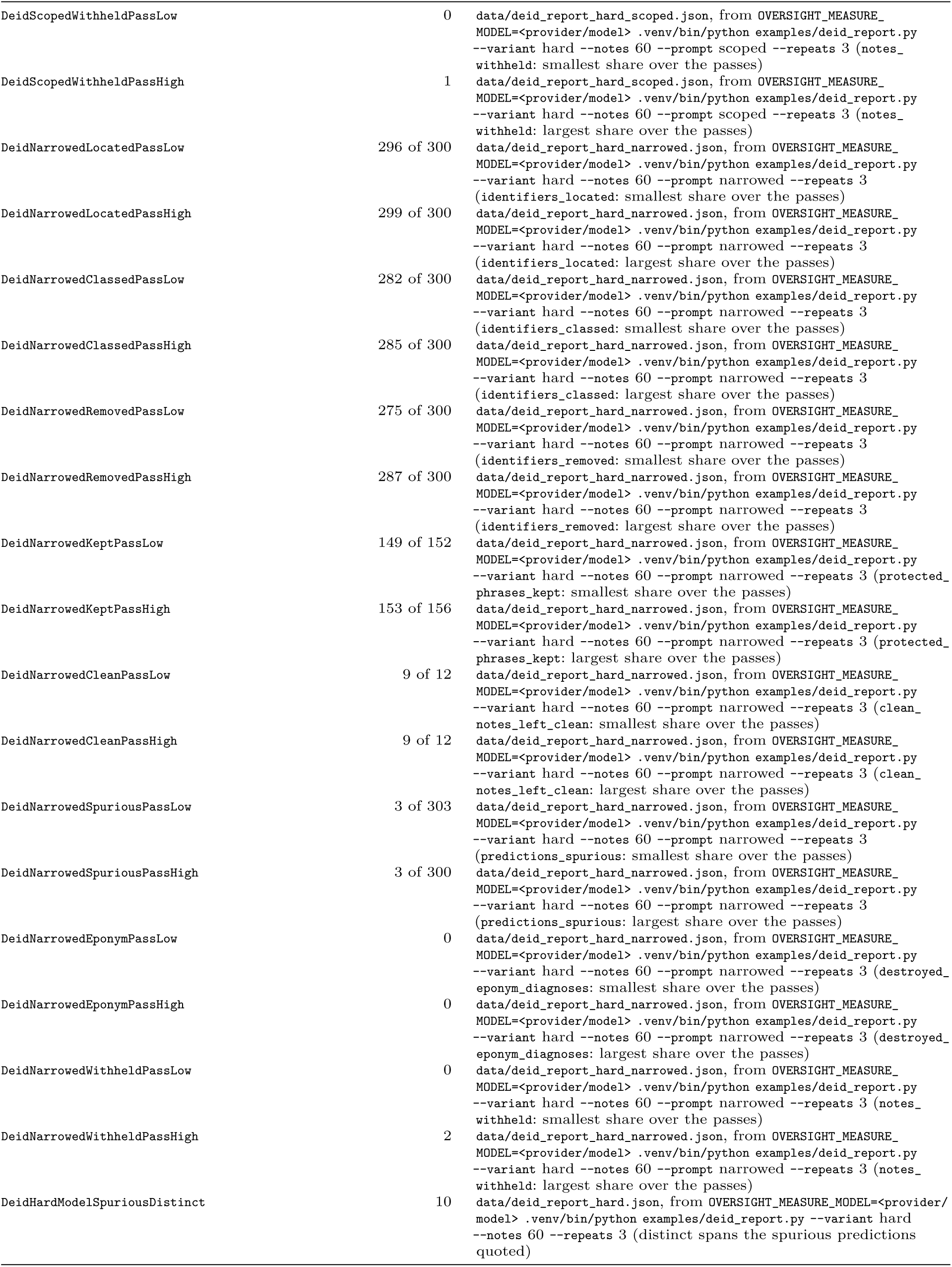
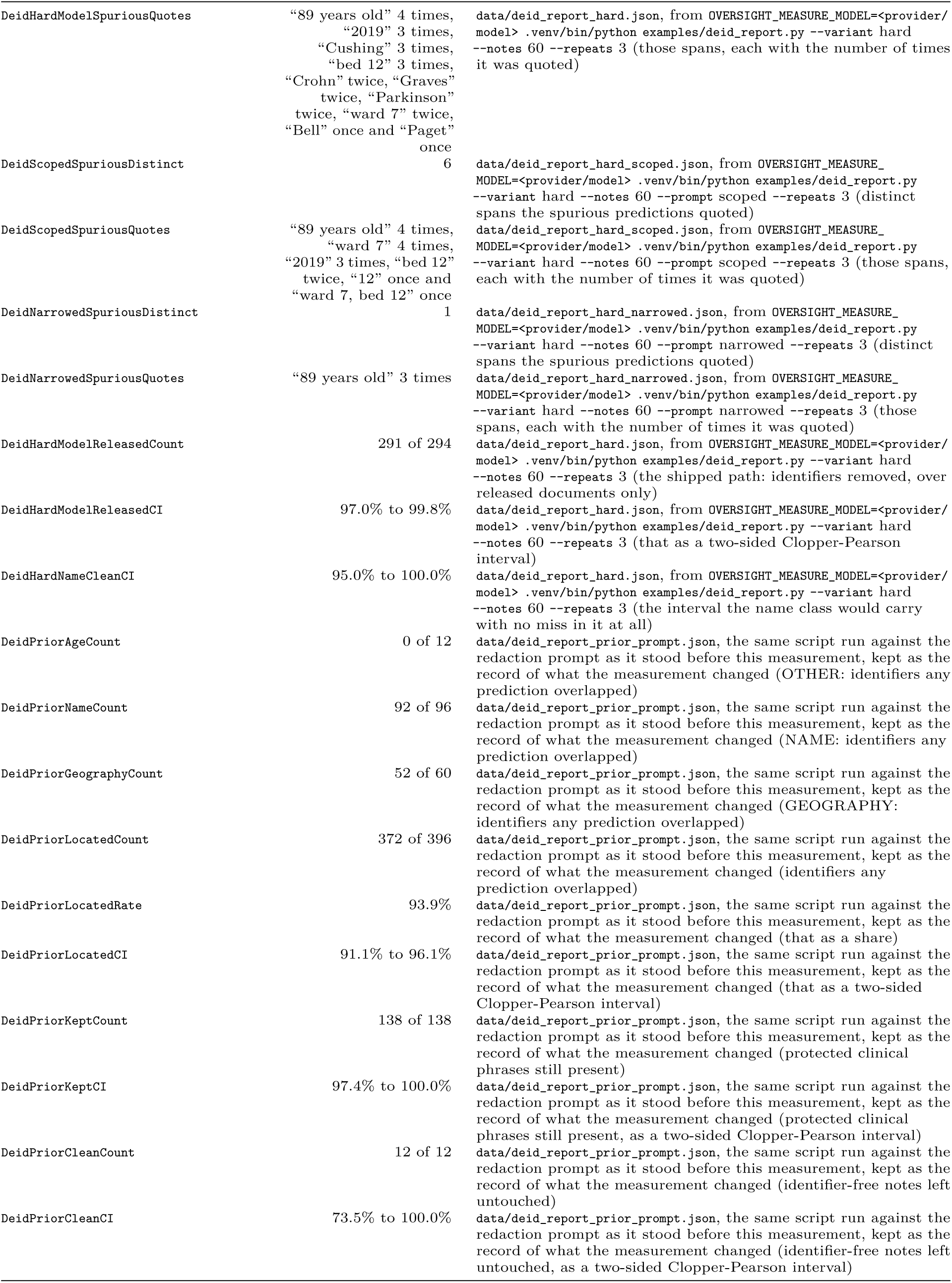
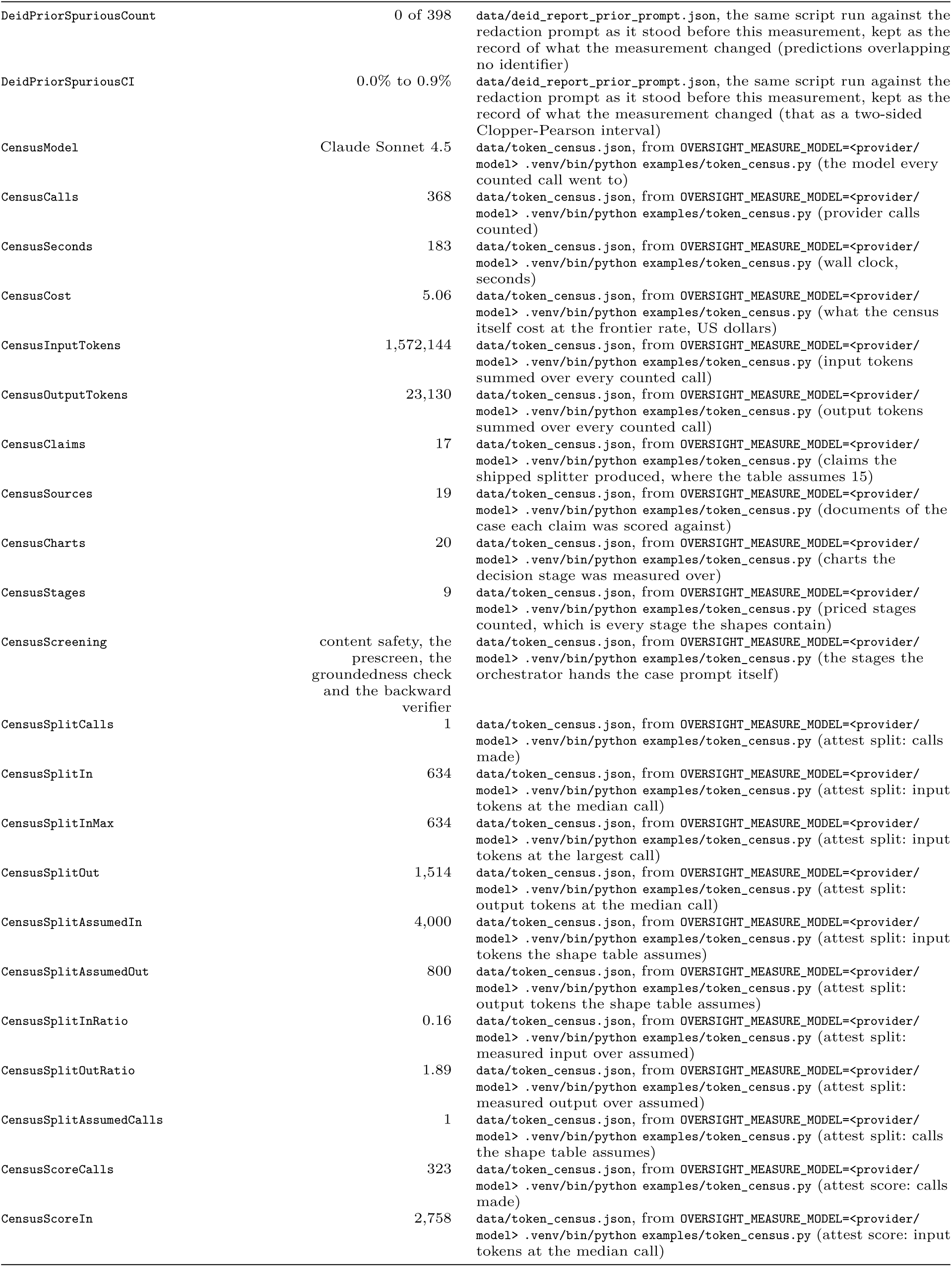
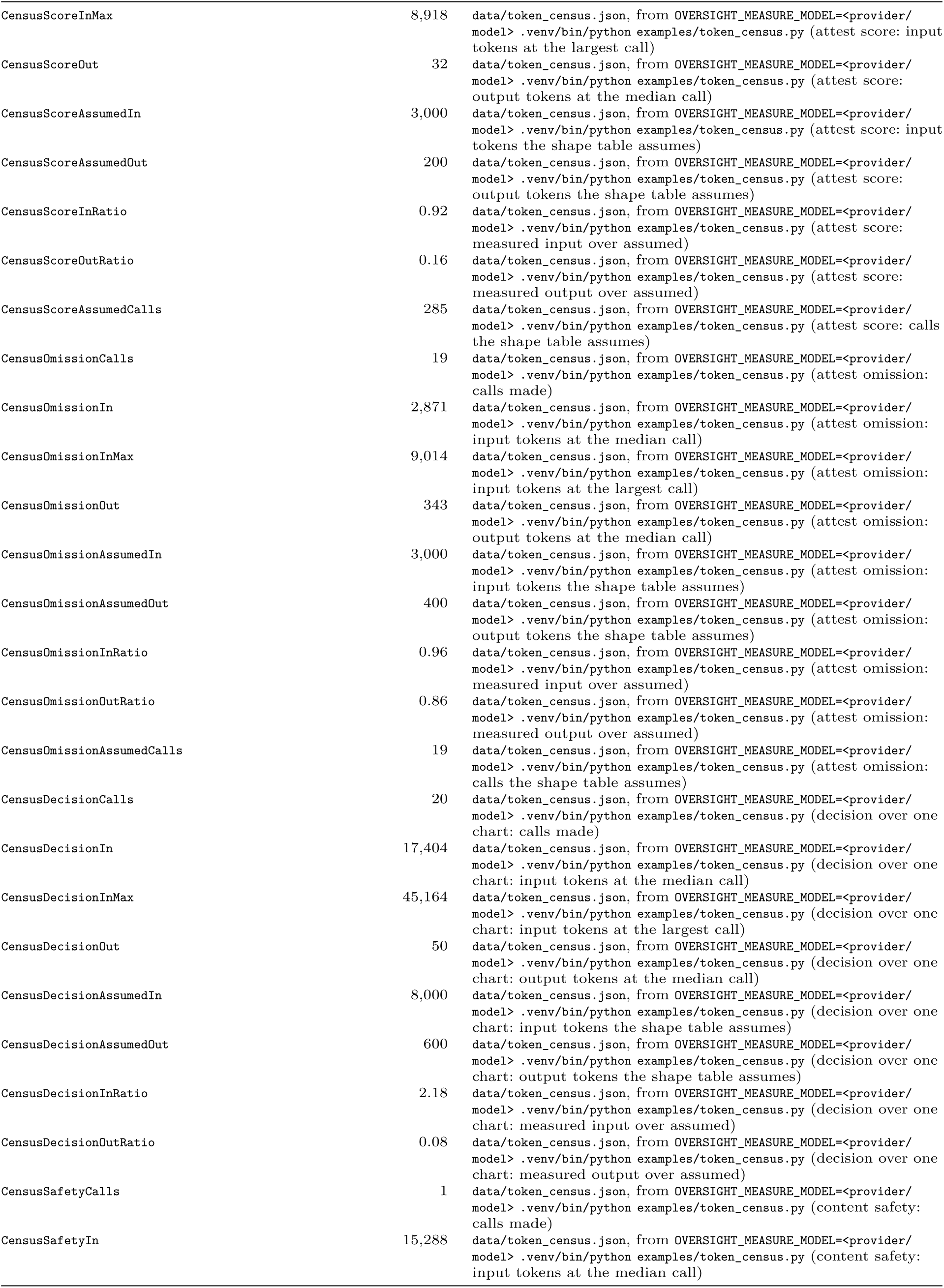
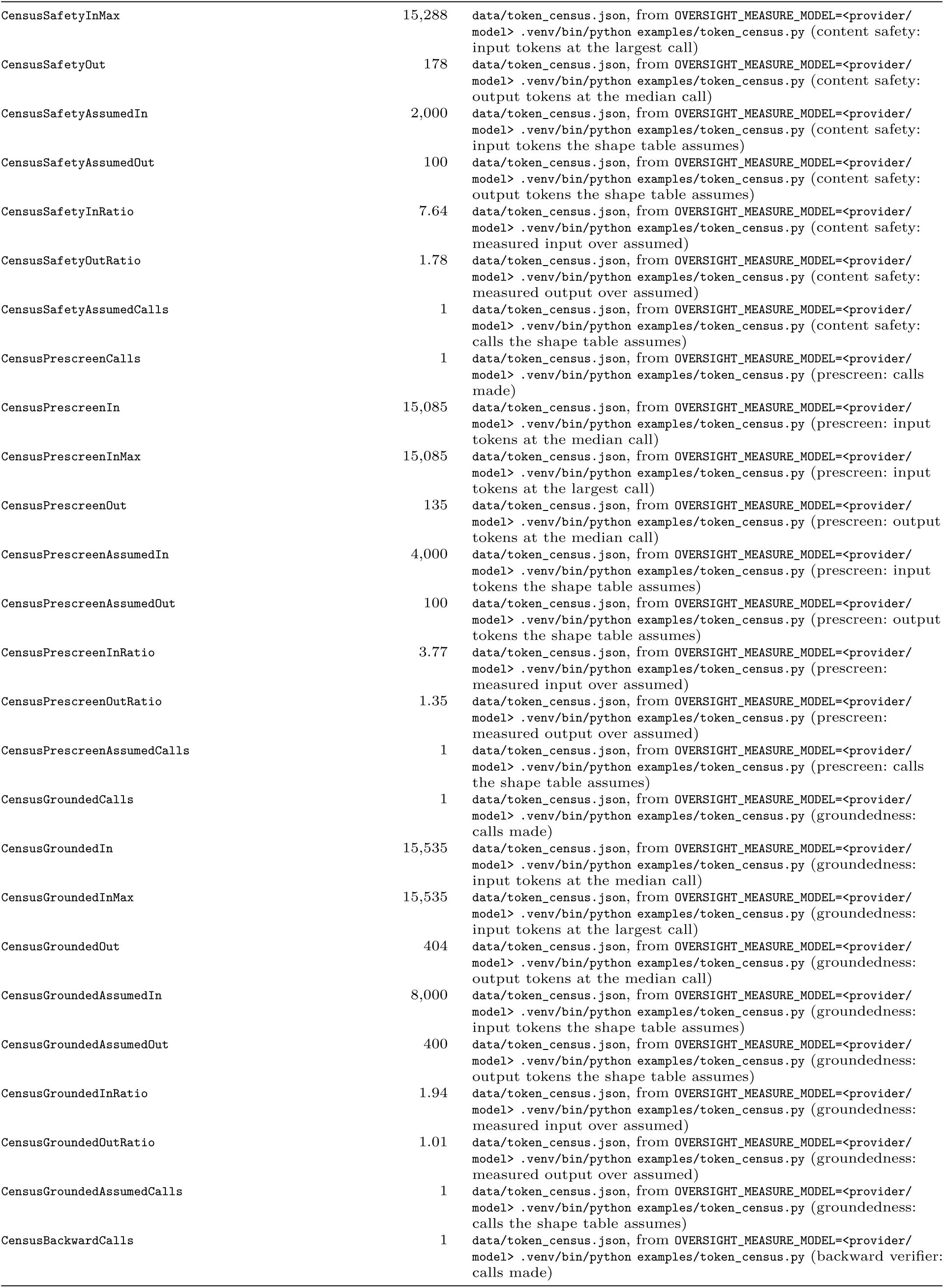
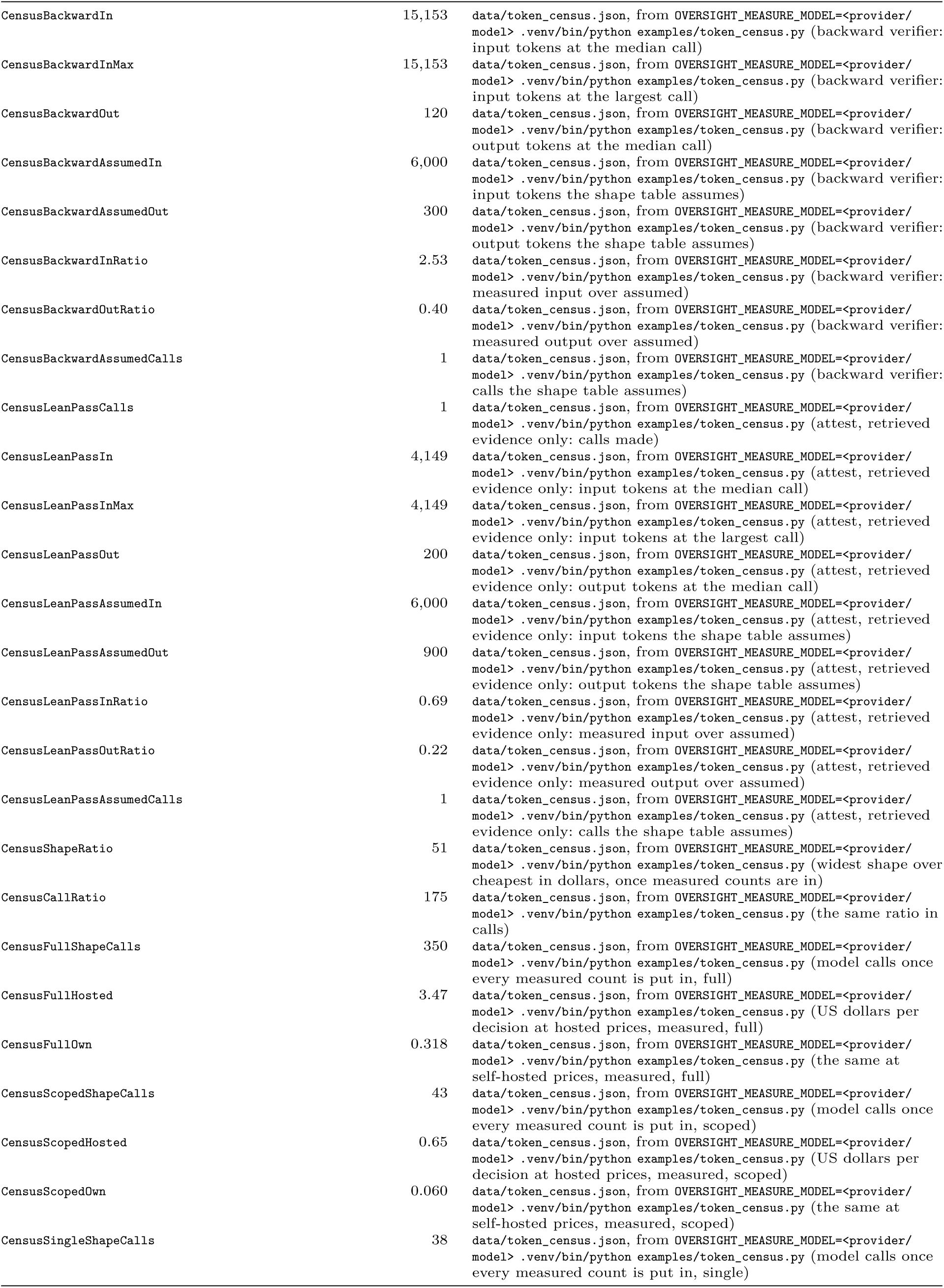
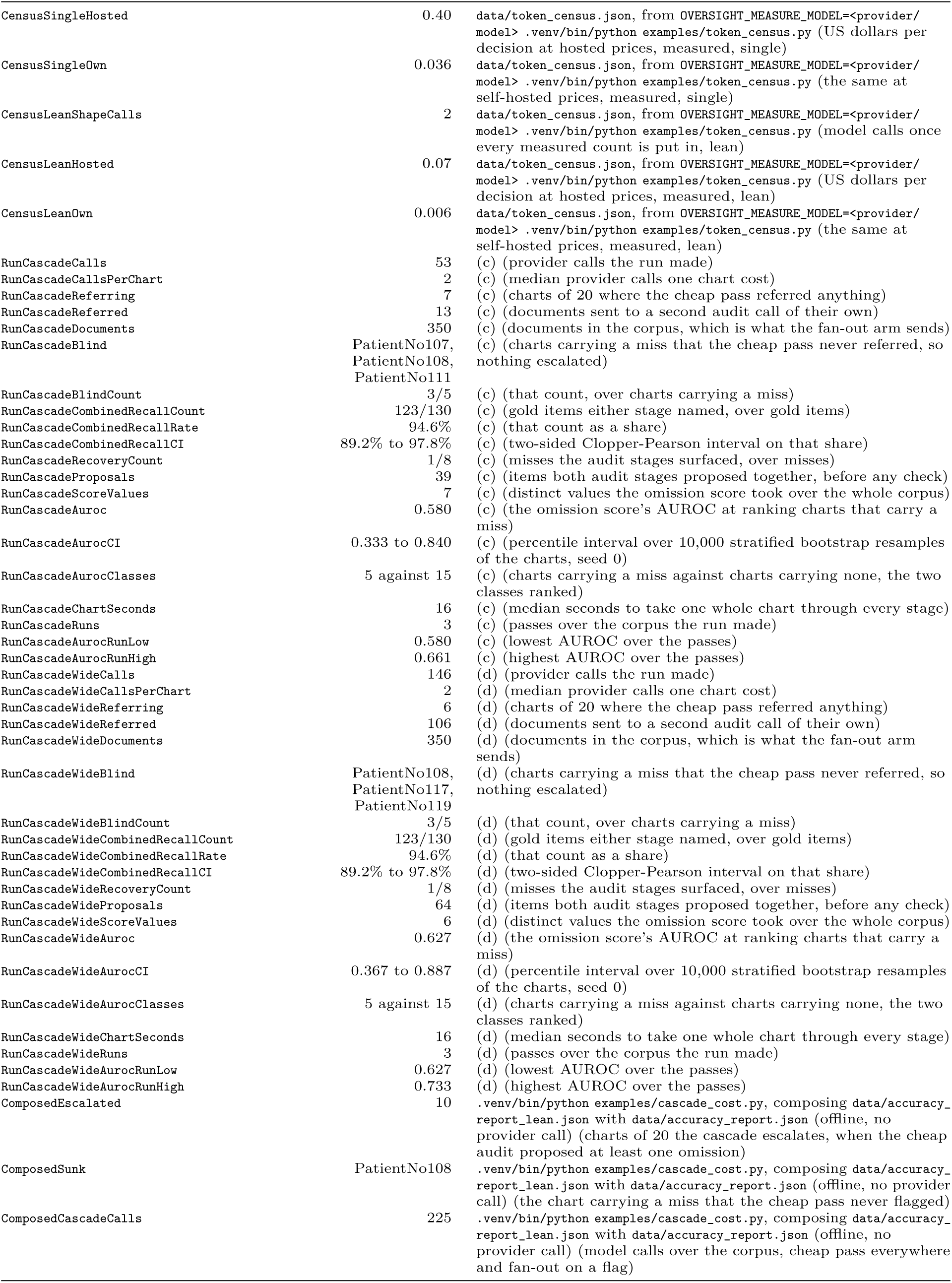
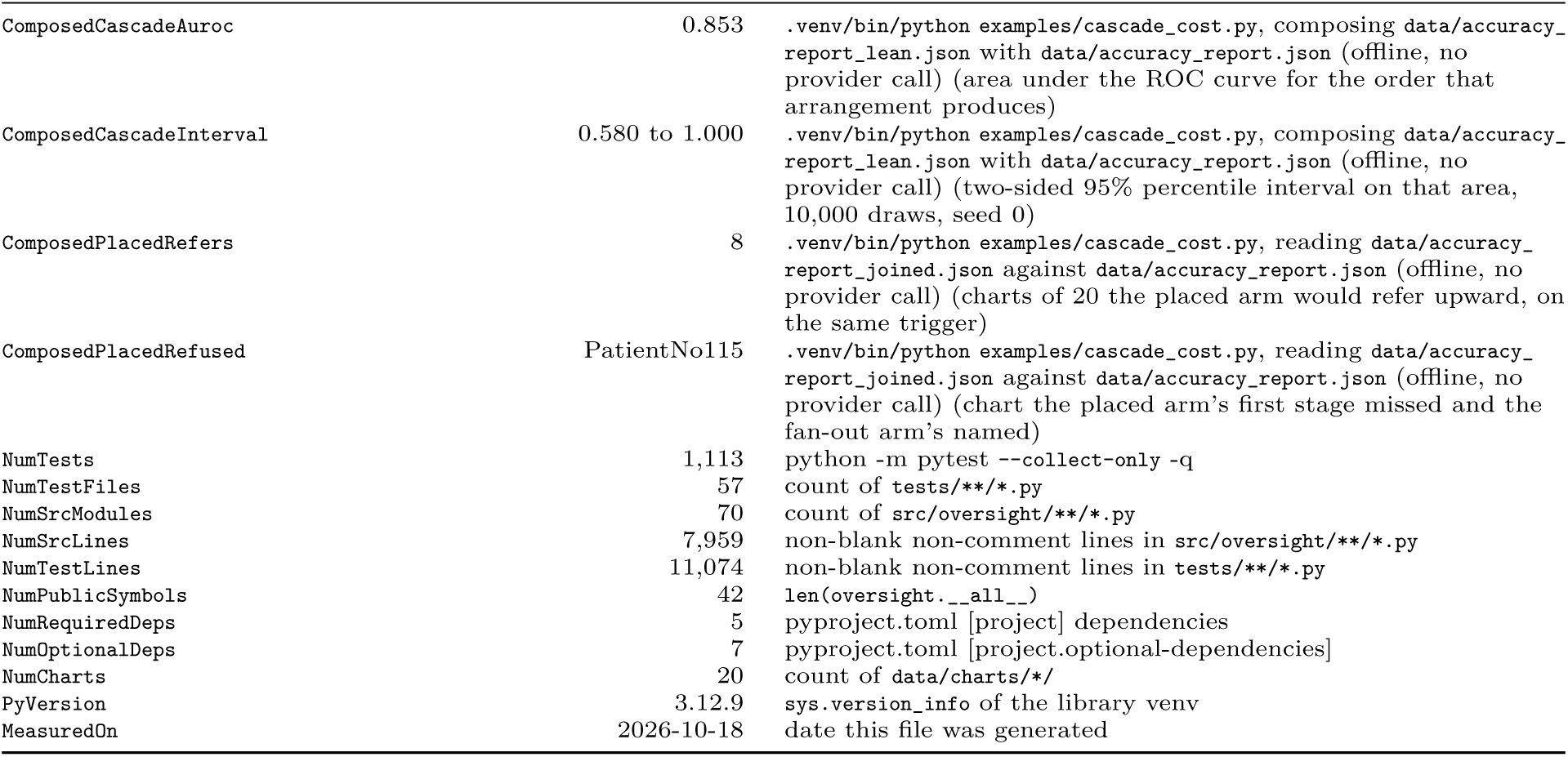
Every quantity the paper states about our own system, with the command that produced it. Where the text writes a quantity as a symbol, the symbol is printed above its name. Entries reading (a) were obtained by `.venv/bin/python examples/vigilance_report.py`, run from the library root, which imports `examples/reviewer_certificate_demo.py`. The parenthesis after that marker says what the quantity is, and where a function returned it or an argument was varied to obtain it, names that too. Entries reading (b) were read from data/accuracy_report.json, which `.venv/bin/python examples/accuracy_report.py` wrote on one date against one hosted model. That is the only report here that reaches a provider, so it is the only one whose digits a reader’s own run will not reproduce, and Section S says what it does and does not measure. Entries reading (c) and (d) were read from `data/accuracy_report_cascade.json` and `data/accuracy_report_cascade_wide.json`, which the same script wrote under --shape cascade and --shape cascade_wide. Those two reach a provider as well, and Table 16 is where they are reported.

## Notes

### Competing Interest Statement

All authors are employees of ConcertAI, which owns the oversight system described in this work. No author or institution received any payment or service from a third party in the past 36 months in connection with this work. No external funding was received.

### Author Declarations

This work used only simulated data and data that were openly available to the public before the study began. No ethics committee or institutional review board decision applies, no dataset was obtained by request, application or screening, and no new human data were collected. The simulated reviewer data are generated by code written for this paper and are reproducible from the seeds recorded in the replication deposit. Two openly available third-party corpora supply the measured physician miss rates. Both were public before this study started and neither is redistributed by us. 1. HealthBench meta-evaluation, released publicly by OpenAI, downloaded from https://openaipublic.blob.core.windows.net/simple-evals/healthbench/2025-05-07-06-14-12_oss_meta_eval.jsonl and verified against the SHA-256 checksum 6a343ed3fcb6f28353c2c6c157ae30ddf2db874c947b27ba474ad54da9c5dbac. It contains grades that physicians gave to model-generated answers under anonymised identifiers. 2. CODE-test reader annotations, released publicly at https://github.com/antonior92/automatic-ecg-diagnosis under data/annotations/. It contains independent readings of electrocardiograms, used in the appendix. Our loaders refuse to report a rate from bytes whose checksum does not match the pinned value, so a reader cannot reproduce us against a different release.

### Summary of Updates

Revised the reporting of the physician miss rate measured from the HealthBench meta-evaluation. The interval now resamples whole conversations, because grades from the same conversation are not independent, and runs from 21.6 to 31.7 percent around the same 26.3 percent estimate. The text now states that every scored defect comes from the context-seeking and hedging parts of HealthBench, the only answers graded by three or more physicians, and reports the rate separately for accuracy and for helpful-and-safe criteria. We no longer claim that most of the spread in miss rates between physicians is real, because the physicians graded different answers, and we cite Borgohain and Mariathas (2026), who find that physician identity explains little of the disagreement in this corpus. Correction to the version 2 note: the electrocardiogram reader analysis was not removed. It remains in the appendix, and the data statements now list both corpora.

